# Trends and patterns of sedative prescribing in primary care in Ireland between 2014 and 2022 - a repeated cross-sectional study

**DOI:** 10.1101/2024.11.26.24317964

**Authors:** Molly Mattsson, Ahmed Hassan Ali, Fiona Boland, Michelle Flood, Ciara Kirke, Emma Wallace, Derek Corrigan, Mary E Walsh, Tom Fahey, Frank Moriarty

## Abstract

**Background:** The trends in sedative use have varied in recent years. Benzodiazepines and z-drugs are indicated for anxiety and sleep disorders, but should be limited to short term use. The aim of this study is to examine trends and patterns in sedative prescribing in Ireland between 2014 and 2022, as well as comparing trends between Ireland and England within the same period.

**Methods:** Monthly data on medicines prescribed and dispensed in primary care on the means-tested General Medical Services (GMS) scheme in Ireland was used. Volumes of prescribed benzodiazepine and z-drug use and patterns of prescribing, including initiations, discontinuations, chronic use, and high-risk prescribing were summarised per year. Other sedating agents (sedating antihistamines, antidepressants, and antipsychotics) were also analysed. Volume of use outcomes were compared with NHS data from England for the same period.

**Results:** The rate of benzodiazepine and z-drug dispensings per 1,000 GMS population decreased by 4%, from 1,531 in 2014 to 1,474 in 2022. T By comparison in England, there was a steeper decrease of 27% in the dispensing rate and the level of use was substantially lower, falling from 288 dispensings per 1,000 population in 2014 to 210 in 2022. In Ireland, dispensing rates were highest amongst women and older age groups. High-risk dispensings of benzodiazepines and z-drugs decreased over the study period

**Discussion:** Despite decreases in benzodiazepine and z-drug dispensings, rates remain high in Ireland and may suggest a need for enhanced availability of non-pharmacological interventions, and improved education and deprescribing support for healthcare professionals.

**Statement of Significance:** The use of benzodiazepines and z-drugs has decreased in many populations in recent years, while prescribing of other sedatives has increased. In Ireland, comparatively higher levels of prescribing have been previously identified. Describing and quantifying medication use is important to monitor medication safety at the population level, however this can be complex and therefore a range of indicators are needed to capture differences.

The findings of this study suggest that benzodiazepine and z-drug dispensing is decreasing in Ireland, including high-risk dispensings and dispensings to older age groups. However, in comparison to England, dispensing rates remain high and suggest a need for enhanced availability of non-pharmacological services and interventions, as well as improved education and deprescribing support for healthcare professionals.

## Background

Benzodiazepines and z-drugs are indicated for acute anxiety and sleep disorders, as well as for a number of primary health conditions, including the management of epilepsy and muscle spasm. The use of benzodiazepines and z-drugs is associated with significant side effects and a high risk of dependency, with cessation often accompanied by withdrawal symptoms, in particular for chronic users of short-acting variants^1^. Benzodiazepines and z-drugs may also increase the risk of accidents, with chronic and regular use found to be associated with a high risk of road collisions^2^. For these reasons, many guidelines indicate that prescriptions should be limited to short term use, with the British National Formulary (BNF) recommending that uninterrupted usage does not exceed four weeks^3^. In Ireland, controlled drugs are subject to additional legislation and regulations including restrictions on possession and supply, additional requirements for prescriptions, and specific law enforcement powers. Controlled drugs are divided into schedules, which restrict their use according to their relative scientific and/or medicinal value^4^. In Ireland, most benzodiazepines and z-drugs were historically not classified as controlled drugs, however a new schedule, Schedule 4, Part 1, was created for these drugs in 2017^5^, thus restricting their supply.

A recent analysis of global trends in benzodiazepine and z-drug use found distinct differences in consumption and trends across countries in region, with the consumption in high-income countries much higher than in middle-income countries^6^. In Ireland, previous research has shown that there was a reduced trend in benzodiazepine prescribing between 2005 and 2015, while prescribing of Z-drugs increased^7^, however, a more recent analysis found that both benzodiazepine and Z-drug prescribing increased between 2017 and 2019^8^. In the UK, prescribing has reduced in recent years, with the number of people prescribed a benzodiazepine reducing from 1.4 million in 2017 to 1.1 million in 2021^9^. Outside of benzodiazepines, drugs from other drug classes may also be used off-licence (i.e. prescribed for an unlicensed indication) for sedative purposes, particularly at lower doses This includes antidepressants with sedative properties, including doxepin, trazodone, and mirtazapine, all of which have seen increased dispensings in recent years in the UK^10^. Similarly, dispensings of atypical antipsychotics (risperidone, quetiapine, and olanzapine), often prescribed at low dosages for sleep difficulties, have increased^11^. A recent Canadian study found that while prescribing of benzodiazepines and z-drugs have decreased in recent years, there has been an increase in low dose trazodone and quetiapine^12^.

Describing and quantifying patterns and quality of medication use is important to monitor medication safety at the population level, however this can be complex and therefore a range of indicators are needed to capture differences. The OECD has outlined 11 quality indicators for prescribing in primary care, including indicators of long-term use of benzodiazepines amongst older adults^13^. Ireland has had the second highest rate of chronic benzodiazepine use among older adults among OECD countries since 2011, and unlike other countries, saw little decline in this indicator between 2011 and 2021^14^. While this is an important indicator of appropriate prescribing of these medications across different national contexts^15^, it provides a limited assessment of utilisation, not accounting for rates of initiation or discontinuation, or other forms of high-risk prescribing such as combination with other sedative drugs or utilisation of high doses.

Therefore, the aim of this study was to examine trends in volume and patterns (including volume of use stratified by age group and sex, prevalence of use, and rates of initiation, discontinuation, and chronic use, as well as high-risk dispensing indicators) of sedative dispensings, particularly benzodiazepines and z-drugs, in Ireland between 2014 and 2022. Additionally, it aimed to assess trends in volume of sedatives in Ireland versus England in a cross-country comparison.

## Methods

This is a repeated cross-sectional study of medicines dispensing in primary care in Ireland between 2014 to 2022. This study forms part of a larger project focused on analgesic and sedative prescribing, the protocol for which has been previously published^16^. The study was approved by the RCSI University of Medicine and Health Sciences Research Ethics Committee (ref: REC202201015) and the Health Service Executive (HSE) Reference Research Ethics Committee B (ref: RRECB1022FM).

## Population and data sources

This study includes individuals eligible for the General Medical Services (GMS) scheme in Ireland. Eligibility to this scheme is based on age and income and covers approximately 32% of the population, and therefore eligible persons tend to be more socioeconomically deprived than the general population^17^. However, the scheme does cover the vast majority of adults aged 70 years and over, and the data provides complete information on prescribed medications that are dispensed to eligible people.

Data were obtained from the Health Service Executive (HSE) Primary Care Reimbursement Service (PCRS), which administers community drug schemes in Ireland. Pharmacies transmit claims for medications dispensed under these schemes to the PCRS at the end of each month for reimbursement. The pharmacy claims database from the PCRS contains records of prescribed medications which were dispensed to individuals eligible for community drug schemes. This is a national database and is used extensively for pharmaceutical policy and pharmacoepidemiology research in Ireland^17^. The data are hierarchical, with dispensed medications nested within prescriptions, prescriptions nested within individuals, and individuals nested within prescribers^17^. Data were requested from the PCRS for this study through an information request, and data were provided to the research team under a data transfer agreement. Supplementary Table 1 provides a list of the medications for which data were requested and that are included in the analysis. Data were at the level of individual medications dispensed to a patient. For each item, information included anonymised identifiers for individuals, prescribers and pharmacies, Local Health Office (LHO) where the drug was dispensed, date of dispensing, sex and age group of the individual, drug information (World Health Organisation (WHO) Anatomical Therapeutic Chemical (ATC) code, product name and strength, and defined daily dosage (DDD)), quantity dispensed, and cost.

## National Health Service data (England)

For the cross-country comparison of volume of sedative use, aggregate data relating to prescribing in all GP practices in England were used as an international comparator. These data are available from the English Prescribing Dataset on National Health Service (NHS) Business Services Authority website, and provides monthly statistics of prescribing of different medications aggregated at the level of GP practices for all practices in England^18^. The data relate to NHS prescriptions issued by general practices in England (by any practice prescribing staff) and dispensed in any community pharmacy in the UK. Prescribed products are coded based on their British National Formulary (BNF) classification. In this study we used the ‘items’ variable within the English Prescribing Dataset, which corresponds to the number of items of each prescribed product that was dispensed in the specified month, and equivalent information to the data from Ireland on product strength, quantity dispensed and cost. A prescription item refers to a single supply of a medicine, dressing or appliance prescribed on a prescription form. Item figures do not provide any indication of the length of treatment or quantity of medicine prescribed, however usually these will be issued for one month.

## Outcomes

Outcomes were derived to capture volume of prescribed sedative utilisation for Ireland and England. Sedatives were identified using WHO ATC codes in data for Ireland, and included benzodiazepines and z-drugs, as well as other sedating agents, including certain antidepressants, antihistamines, and atypical antipsychotics dispensed as low-dose products. For data for England, relevant ATC codes were mapped to the equivalent BNF codes to ensure comparability of analysis (See Supplementary Table 1 for included drugs with ATC and BNF codes). Outcomes included number of dispensings, costs, and standard doses, expressed as rates per 1,000 patients. For Ireland, denominators were obtained from PCRS annual reports which report number of eligible individuals at the end of each calendar year^19^. For England, denominators were obtained from the NHS Digital portal^20^. Standard doses provide a fixed unit of measurement dependent on prescription quantity, pack size and strength to capture medications consumption on a common scale across different medications, drugs, or drug classes. The WHO DDD, defined as “the assumed average maintenance dose per day for a drug used for its main indication in adults”^21^ was used for all medications. In addition, for benzodiazepines and z-drugs, diazepam dose equivalents (DDE) were used^22^ (see Supplementary Table 2).

For Ireland, in addition to the volume indicators outlined above, pattern of use indicators were derived, including volume of use stratified by age group and sex, prevalence of use, and rates of initiation, discontinuation, and chronic use, as well as high-risk dispensing indicators as defined by the Scottish Polypharmacy Guidance^23^. An overview of the included outcomes is provided in Supplementary Table 3.

## Analysis

The outcomes above were calculated in each data source for each year from 2014 to 2022. Outcomes were derived for all drug classes and individual drugs, as well as well as for long-acting benzodiazepines^24^ (See Supplementary Table 1). Rates for each outcome were summarised per year overall and for each drug in both countries. The absolute and relative change from 2014 to 2022 was calculated for each, as well as the rate ratio (rate in Ireland divided by the rate in England). The Jonckheere–Terpstra test for trend across years was run for each drug separately for Ireland and England. Considering data from Ireland relates to the GMS scheme, which overrepresents those with lower SES, a subgroup analysis was conducted using data for England from higher deprivation clinical commissioning groups (CCGs) only (those CCGs in the top 33 centiles of deprivation, based on the Index of Multiple of Deprivation 2019^25^

For Ireland, all volume indicators were further calculated by age group and sex. Prevalence of use was calculated as the proportion of individuals dispensed a relevant medicine within each year, as well as by age group and sex. To capture patterns of use, initiation was defined as a dispensing to an individual with no dispensing in the previous 90 days, and discontinuation as an individual with no dispensing in the 90 days following a dispensing. Both of these analyses were repeated using a longer, more conservative 180-day window. Long-term use was analysed for >30 days and for >90 days. To capture high-risk dispensings, the rate of dispensings of benzodiazepines/z-drugs with two or more other sedating or anticholinergic drugs (see Supplementary Table 4) in people aged ≥65 years, as well as the rate of dispensing at a dose equivalent to >40 mg diazepam per day were calculated. All analyses were conducted in Stata version 18 (College Station, TX: StataCorp LLC) and statistical significance for trend tests was defined as p<0.05.

## Results

The number of GMS eligible patients during the study period decreased from 1,768,700 in 2014 to 1,568,379 in 2022. Conversely, the NHS population in England increased year-on-year from 56,545,892 in 2014 to 61,768,942 in 2022. See Supplementary Table 5 for populations for both countries between 2014 and 2022.

## Volume of utilisation

The rate of benzodiazepine and z-drug dispensings per 1,000 GMS population increased in the initial study period from 1,521per 1,000 population in 2014, peaking at 1,569 per 1,000 in 2019, and then decreasing to 1,445 in 2022: a 5% decrease from 2014-2022. The rate of long-acting benzodiazepine dispensings decreased from 437 to 399, a 9% decrease. The trends for individual benzodiazepines and z-drugs varied, with moderate increases seen for clonazepam (17%), lorazepam (12%), zolpidem (7%), and zopiclone (6%). The largest relative decreases were seen lormetazepam (99%), nitrazepam (97%), and prazepam (58%). Zopiclone and zolpidem had the highest rates of dispensing, followed by diazepam and alprazolam (see Figure 1). The rate of dispensings of sedating antidepressants (including low dose doxepin, trazodone, and mirtazapine) increased by 112%, while sedating antihistamines (including promethazine, cyclizine, and ketotifen) increased by 570% and sedating antipsychotics (including low dose olanzapine, quetiapine, and risperidone) by 95%. Combining these with benzodiazepine and z-drugs, the overall dispensing rate of sedative drugs rose from 1,741 per 1,000 in 2014 to 1,940 in 2022, corresponding to an 11% relative increase. See Table 1 for rates of dispensings per 1,000 GMS population for all drugs included in the study. In England, the rate of benzodiazepine and z-drug dispensings decreased by 27% from 288 in 2014 to 210 in 2022. Multiple drugs dispensed in Ireland were not dispensed in England during the study period, including potassium clorazepate, bromazepam, prazepam, alprazolam, flurazepam, and triazolam. The rate of all sedative dispensings per 1,000 NHS population remained steady throughout the study period. See Supplementary Table 6. Figure 2 reports rate ratios for dispensings in Ireland versus England for 2014 and 2022.

**Figure 1.**
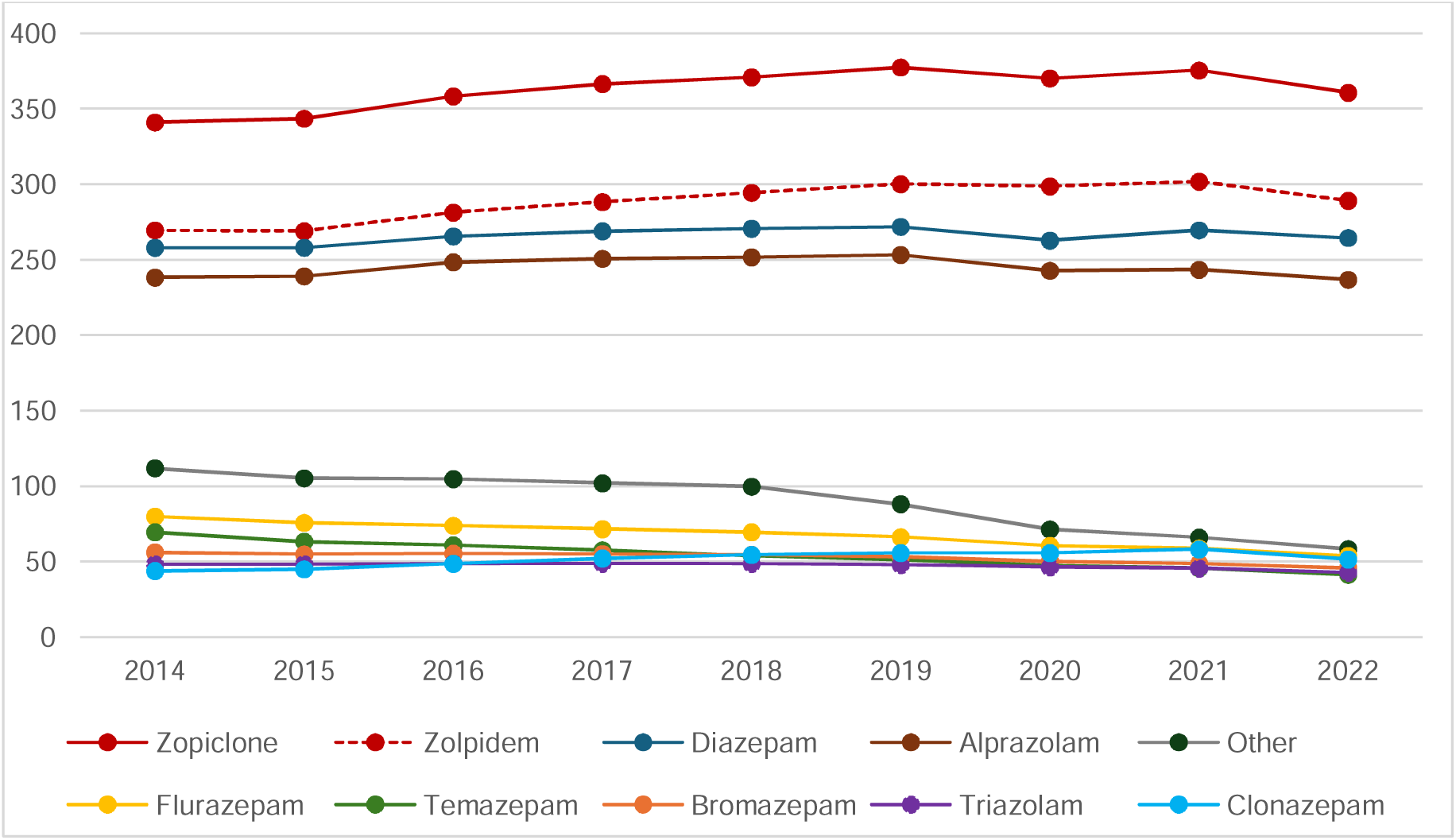
Rate of dispensing per 1,000 GMS population for individual benzodiazepines and z-drugs.

**Figure 2.**
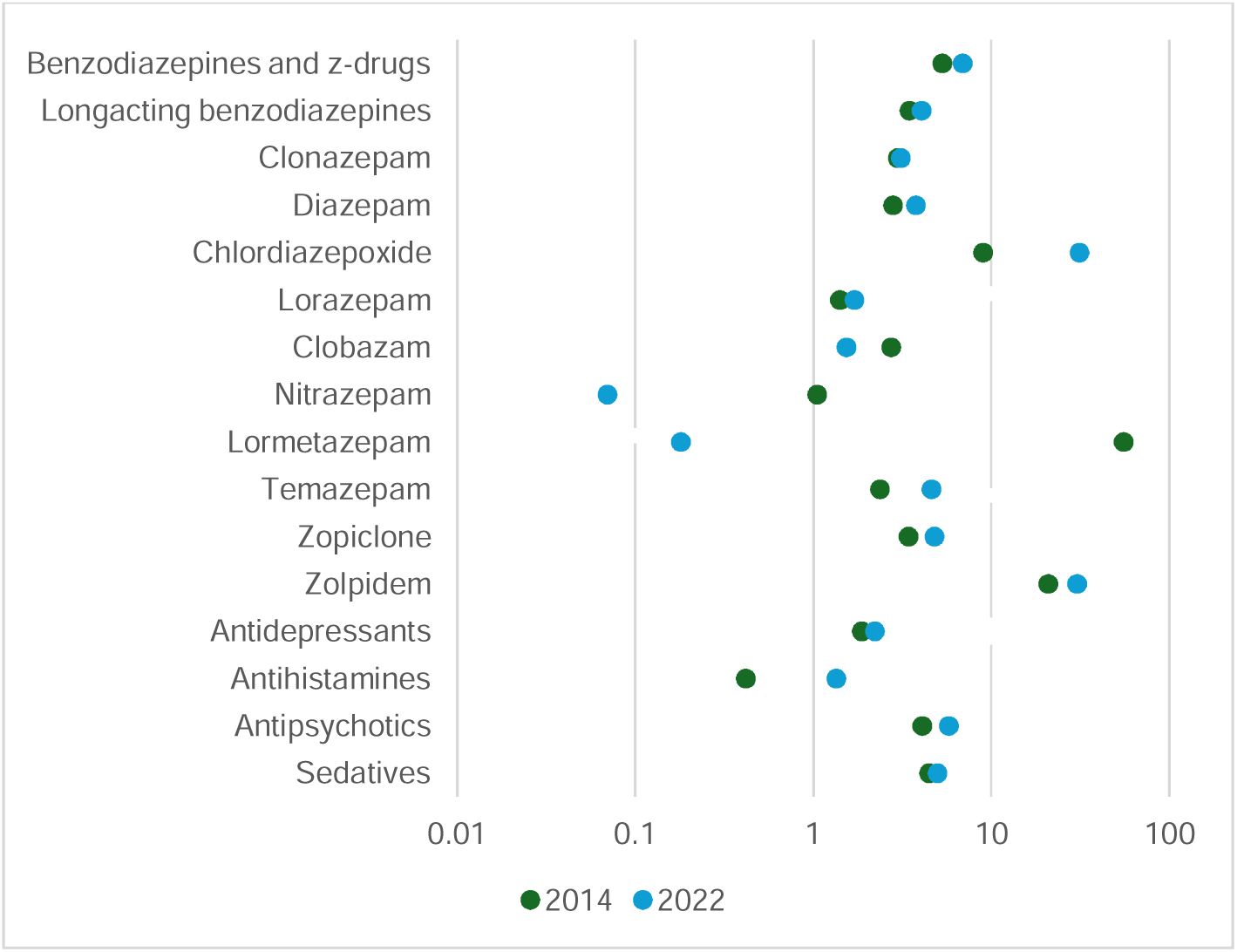
Rate ratio for dispensings in Ireland versus England for 2014 and 2022*. *Rates above 1.0 indicate higher dispensings in Ireland

**Table 1.** Rate of dispensings per 1,000 GMS population in Ireland.

|  | 2014 | 2015 | 2016 | 2017 | 2018 | 2019 | 2020 | 2021 | 2022 | Absolute change | Relative change |
| --- | --- | --- | --- | --- | --- | --- | --- | --- | --- | --- | --- |
| Benzodiazepines and z-drugs | 1521.0 | 1507.2 | 1549.0 | 1562.0 | 1568.9 | 1565.6 | 1506.4 | 1513.4 | 1444.9 | -76.1 | -0.05 |
| Long-acting benzodiazepines | 436.7 | 429.7 | 438.3 | 441.4 | 441.6 | 440.1 | 421.1 | 423.2 | 399.1 | -37.6 | -0.09 |
| *Clonazepam* | 43.8 | 44.9 | 48.5 | 52.1 | 54.5 | 55.7 | 55.7 | 58.1 | 51.4 | 7.6* | 0.17 |
| *Diazepam* | 258.1 | 258.1 | 265.6 | 269.0 | 270.8 | 272.0 | 262.9 | 269.7 | 264.6 | 6.4 | 0.02 |
| *Chloridiazepoxide* | 18.5 | 17.8 | 18.2 | 18.0 | 17.5 | 18.0 | 16.5 | 16.8 | 15.1 | -3.3* | -0.18 |
| *Potassium Clorazepate* | 0.1 | 0.1 | 0.1 | 0.1 | 0.1 | 0.1 | 0.1 | 0.1 | 0.1 | 0.0 | -0.13 |
| *Lorazepam* | 26.0 | 25.4 | 26.0 | 26.3 | 27.4 | 28.1 | 28.2 | 29.2 | 29.0 | 3.0* | 0.12 |
| *Bromazepam* | 56.1 | 54.8 | 55.3 | 55.1 | 54.5 | 53.1 | 50.2 | 48.7 | 45.8 | -10.3* | -0.18 |
| *Clobazam* | 12.0 | 10.6 | 10.2 | 9.9 | 9.6 | 9.1 | 8.7 | 8.9 | 9.2 | -2.8* | -0.23 |
| *Prazepam* | 10.8 | 10.3 | 10.0 | 9.7 | 9.6 | 9.2 | 7.8 | 7.6 | 4.5 | -6.2* | -0.58 |
| *Alprazolam* | 238.4 | 239.2 | 248.5 | 250.8 | 251.8 | 253.3 | 242.8 | 243.4 | 236.8 | -1.5 | -0.01 |
| *Flurazepam* | 79.9 | 75.7 | 73.8 | 71.6 | 69.5 | 66.3 | 60.5 | 58.7 | 53.8 | -26.1* | -0.33 |
| *Nitrazepam* | 13.6 | 12.2 | 11.7 | 10.9 | 10.1 | 9.7 | 8.9 | 3.3 | 0.4 | -13.2* | -0.97 |
| *Triazolam* | 48.2 | 48.4 | 48.8 | 48.7 | 48.5 | 47.9 | 46.2 | 45.5 | 42.8 | -5.4* | -0.11 |
| *Lormetazepam* | 31.0 | 28.8 | 28.4 | 27.0 | 25.5 | 14.0 | 1.1 | 0.1 | 0.0 | -31.0* | -1.00 |
| *Temazepam* | 69.5 | 63.3 | 60.9 | 57.5 | 53.9 | 51.2 | 47.7 | 45.8 | 41.3 | -28.2* | -0.41 |
| *Zopiclone* | 341.1 | 343.7 | 358.5 | 366.6 | 371.1 | 377.6 | 370.3 | 375.6 | 360.8 | 19.7* | 0.06 |
| *Zolpidem* | 269.7 | 269.1 | 281.4 | 288.4 | 294.5 | 300.2 | 298.8 | 301.9 | 289.2 | 19.5* | 0.07 |
| Antidepressants<sup>1</sup> | 91.7 | 104.6 | 118.4 | 131.7 | 146.3 | 159.0 | 172.4 | 192.9 | 194.6 | 102.9* | 1.12 |
| Antihistamines<sup>2</sup> | 10.5 | 13.2 | 17.8 | 24.1 | 29.5 | 39.3 | 50.6 | 64.8 | 70.5 | 60.0* | 5.70 |
| Antipsychotics<sup>3</sup> | 118.1 | 131.4 | 146.2 | 162.2 | 179.1 | 198.2 | 211.6 | 230.3 | 229.9 | 111.8* | 0.95 |
| All sedatives | 1741.4 | 1756.4 | 1831.5 | 1880.0 | 1923.8 | 1962.1 | 1941.1 | 2001.3 | 1940.0 | 198.7* | 0.11 |

<sup>1</sup>doxepin 10mg, trazodone 50 and 100mg, mirtazapine 15mg
<sup>2</sup>promethazine, cyclizine, ketotifen
<sup>3</sup>olanzapine 2.5mg, quetiapine 25mg, risperidone 0.5mg
\*test for trend significant at 0.05

There were similar changes in the cost rates per 1,000 GMS/NHS population, as shown in Supplementary Table 7.

The rate of diazepam dose equivalent (DDEs) dispensed per 1,000 GMS population per day for all benzodiazepines decreased from 834 in 2014 to 764 in 2022, a 9% decrease, with most individual drugs decreasing. The largest relative decreases were similarly to rate of dispensings seen for flunitrazepam, lormetazepam, nitrazepam, and prazepam. Full results for rates of DDEs and DDDs per 1,000 GMS/NHS population are available in Supplementary Table 7. Rate ratio plots for DDEs and DDDs in Ireland vs England for 2014 and 2022 are available in Supplementary Figure 1.

Results of the subgroup analysis of the CCGs in England in the top 33 centiles for deprivation showed dispensing rates of benzodiazepines lower than those of the general population in 2014 (238 vs 288), and slightly higher in 2022 (216 vs 210). For all sedatives the rates of dispensing was stable throughout the study period, while there was an increase of 31% in the most deprived CCGs. See Supplementary Table 8 for rates of dispensings, costs, DDDs, and DDEs for this subgroup.

## Pattern of utilisation

Based on analysis of individual-level data in Ireland, the rate of benzodiazepine and z-drug dispensings varied substantially between age groups. Throughout the study period the 75+ years age group had the highest rate of dispensings, however it also saw the largest relative decrease between 2014 and 2022, decreasing from 3,736to 2,826 per 1,000 populations. All other age groups also saw lower rates in decreased for all age groups between 2014 and 2022, however for the youngest age groups (<5, 5-11, 12-15, 16-24) the differences were small, and for younger adults (25-34, 35-44) there was no significant linear change in trend. See Figure 3A and Supplementary Table 9.

**Figure 3A.**
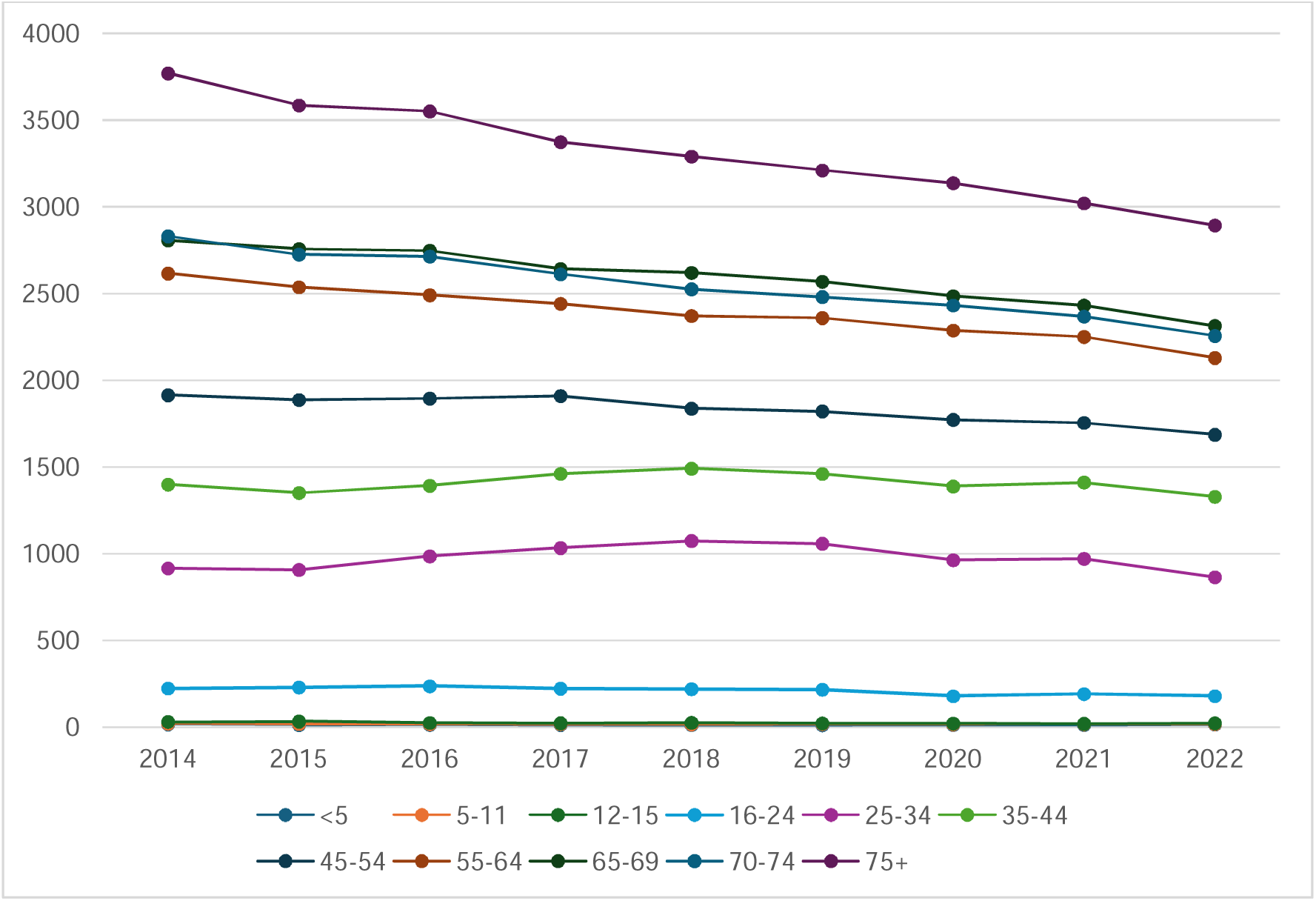
Rate of benzodiazepine and z-drug dispensings per 1,000 GMS population by age group.

The rate of dispensings for benzodiazepines and z-drugs was higher for females compared to men throughout the study period, decreasing slightly from 1,865 in 2014 to 1,730 in 2022 for females and 1,133in 2014 to 1,100in 2022 for males. See Figure 3B and Supplementary Table 9.

**Figure 3B.**
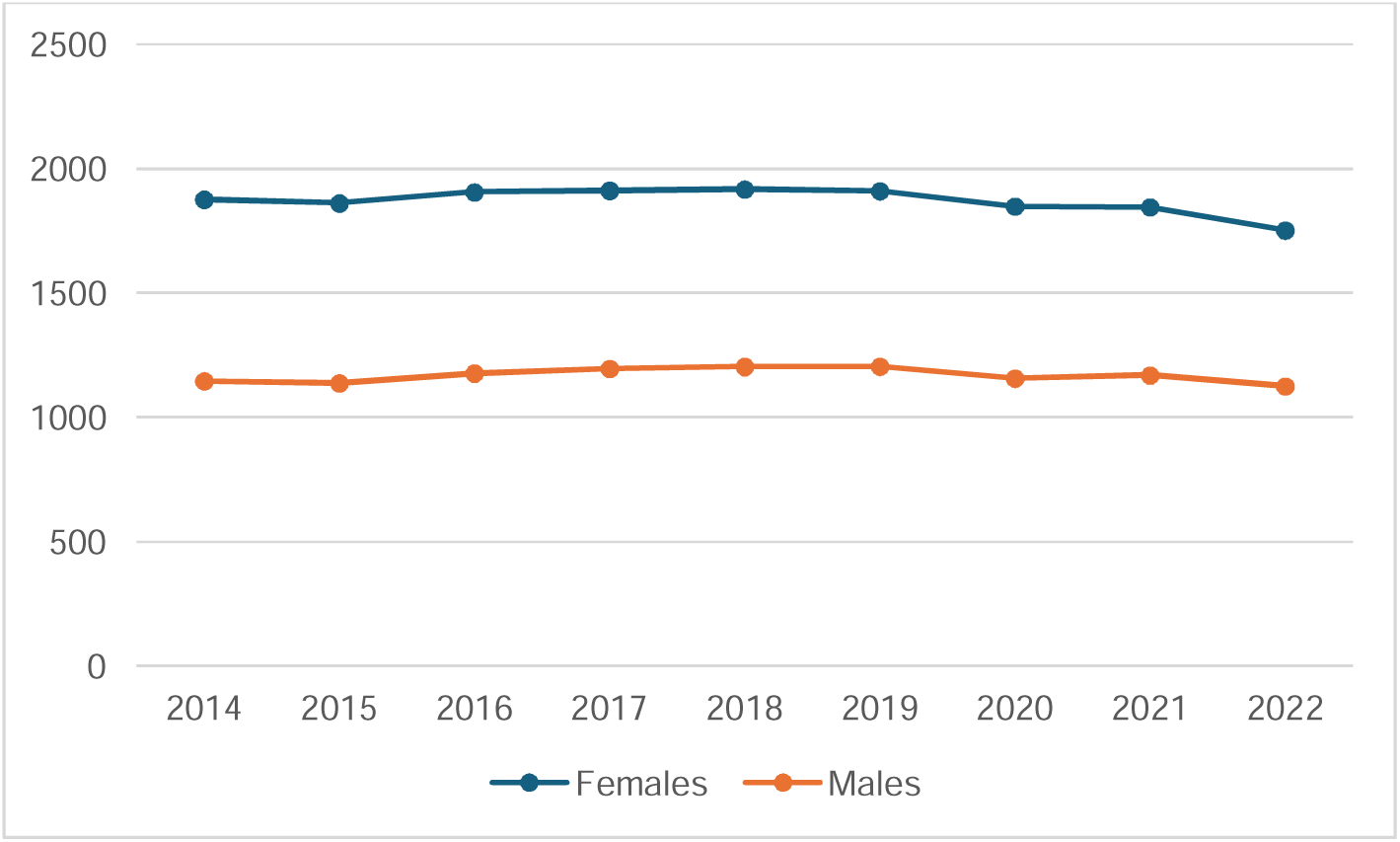
Rate of benzodiazepine and z-drug dispensings per 1,000 GMS population by sex.

Similar to the rate dispensings, the rate of DDEs per 1,000 GMS population was highest in the 75+ age group, followed by the 55-64, 65-69, and 70-74 age groups, and higher for women compared to men. See Supplementary Table 9 for rate of DDE, DDD, and cost per 1,000 GMS population by age group and sex.

The prevalence of benzodiazepines and z-drugs use in the GMS population decreased slightly during the study period, albeit not significantly, with 18.2% of eligible individuals receiving at least one dispensing in 2014, compared to 16.4% in 2022. The prevalence of benzodiazepine and z-drug initiations, based on a medication-free interval of 90 or 180 days, was stable at 7.7-9.0% and 6.3- 7.2% respectively, while discontinuations, based on a medication-free interval of 90 and 180 days, ranged between 4.9-5.6% and 3.2-3.6% respectively. Similarly, chronic use of benzodiazepines and z-drugs for more than 30 days was stable at 10.6-11.7%, while 8.0-9.6 of the GMS population received dispensings for more than 90 days. Prevalence of initiations, discontinuations, and chronic use was not calculated for 2014, as results would be skewed due to the lack of access to 2013 data. All results were non-significant for trend between 2015 and 2022. See Table 2 and Supplementary Table 10.

**Table 2.** Prevalence (%) of any dispensings, initiations (90/180 days), discontinuations (90/180 days), and chronic use (30/90 days) of benzodiazepines and z-drugs in the GMS population.

|  | 2014 | 2015 | 2016 | 2017 | 2018 | 2019 | 2020 | 2021 | 2022 |
| --- | --- | --- | --- | --- | --- | --- | --- | --- | --- |
| Prevalence of any dispensings | 18.2 | 17.8 | 18.3 | 18.4 | 18.3 | 18.1 | 16.5 | 16.9 | 16.4 |
| Prevalence of initiations after 90 days | - | 8.2 | 8.9 | 9.0 | 8.9 | 8.8 | 7.7 | 7.8 | 7.7 |
| Prevalence of initiations after 180 days | - | 6.3 | 7.1 | 7.2 | 7.1 | 7.1 | 6.3 | 6.4 | 6.4 |
| Prevalence of discontinuations after 90 days | - | 5.2 | 5.5 | 5.6 | 5.5 | 5.6 | 4.9 | 5.0 | 5.0 |
| Prevalence of discontinuations after 180days | - | 3.2 | 3.5 | 3.5 | 3.5 | 3.6 | 3.3 | 3.3 | 3.3 |
| Prevalence of chronic use >30 days | - | 11.2 | 11.5 | 11.6 | 11.7 | 11.6 | 11.1 | 11.2 | 10.6 |
| Prevalence of chronic use >90 days | - | 9.1 | 9.3 | 9.5 | 9.6 | 9.6 | 9.2 | 9.3 | 8.9 |

Results by age group and sex generally followed age/sex figures presented above. See Supplementary Table 11 and Supplementary Figure 2 for prevalence of initiations, discontinuations, chronic use, and high risk dispensings by age group and sex.

The prevalence of high-risk dispensings decreased during the study period. The proportion of GMS eligible individuals aged 65 years or over receiving a benzodiazepine or z-drug dispensing with two or more other sedating or anticholinergic drugs (see Supplementary Table 4 for a full list of anticholinergics) decreased from 18.5% in 2014 to 13.1% in 2022. Out of all benzodiazepine and z-drug dispensings to people aged 65 years or over, this constituted 53.8% of dispensings in 2014, decreasing slightly to 49.0% in 2022. The proportion of GMS eligible individuals receiving benzodiazepines or z-drugs dispensed at a dose equivalent to >40 mg diazepam per day remained steady at 0.9-1.0% during the study period, constituting 5.7% of dispensings in 2014 and 5.4% in 2022 (see Table 3).

**Table 3.** Prevalence (%) of high risk dispensings in the GMS population.

|  | 2014 | 2015 | 2016 | 2017 | 2018 | 2019 | 2020 | 2021 | 2022 |
| --- | --- | --- | --- | --- | --- | --- | --- | --- | --- |
| **Prevalence of benzodiazepine or z-drug dispensings with two or more other sedating or anticholinergic drugs in people aged ≥65 years** |  |  |  |  |  |  |  |  |  |
| Benzodiazepines and z-drugs | 18.5 | 17.5 | 17.1 | 16.1 | 15.6 | 15.0 | 14.1 | 13.6 | 13.1 |
| Long-acting benzodiazepines | 5.4 | 5.1 | 5.0 | 4.7 | 4.5 | 4.3 | 4.1 | 3.9 | 3.7 |
| **Proportion of individuals receiving benzodiazepine or z-drug dispensings with two or more other sedating or anticholinergic drugs out of all benzodiazepine or z-drug dispensings in people aged ≥65 years** |  |  |  |  |  |  |  |  |  |
| Benzodiazepines and z-drugs | 53.8 | 54.1 | 53.1 | 52.2 | 51.4 | 50.8 | 50.7 | 49.7 | 49.0 |
| Long-acting benzodiazepines | 54.3 | 54.0 | 53.1 | 52.1 | 50.9 | 50.7 | 51.5 | 50.1 | 49.2 |
| **Benzodiazepines or z-drugs dispensed at a dose equivalent to >40 mg diazepam per day** |  |  |  |  |  |  |  |  |  |
| Benzodiazepines and z-drugs | 1.04 | 1.02 | 1.03 | 1.04 | 1.02 | 1.01 | 0.94 | 0.92 | 0.88 |
| Long-acting benzodiazepines | 0.66 | 0.65 | 0.66 | 0.66 | 0.66 | 0.65 | 0.60 | 0.59 | 0.57 |
| **Proportion of individuals receiving benzodiazepines or z-drugs dispensed at a dose equivalent to >40 mg diazepam per day out of all benzodiazepine or z-drug dispensings** |  |  |  |  |  |  |  |  |  |
| Benzodiazepines and z-drugs | 5.7 | 5.7 | 5.6 | 5.6 | 5.6 | 5.6 | 5.7 | 5.4 | 5.4 |
| Long-acting benzodiazepines | 9.2 | 9.2 | 9.1 | 9.2 | 9.1 | 9.1 | 9.3 | 8.9 | 9.0 |

## Discussion

Between 2014 and 2022, there was an overall rise in sedative dispensings, but a decrease in benzodiazepine and z-drug dispensings. Previous research has found that benzodiazepine dispensings in this population decreased significantly from 2005 to 2015, while z-drugs increased^7^. Our research indicates that benzodiazepine use increased up to 2019, but has fallen below 2015 levels in recent years, while z-drugs have continued to increase with the exception of a small drop in 2022. This is in line with literature from the UK, where the trend in benzodiazepine and z-drug use similarly has decreased in recent years, consistent with our England analysis^9^. Recent research from the US has shown that the onset of the COVID-19 pandemic was associated with increased rates in z-drug prescribing in both men and women, along with an increase in benzodiazepine prescriptions in women, with higher rates in 2020 and 2021 compared to 2018 and 2019^26^. Our research does not show any evidence of a similar trend in Ireland.

In Ireland, the creation of a new controlled schedule for benzodiazepines and z-drugs in 2017 was followed in 2018 by the Medicines Management Programme (MMP) issuing the first formal guidance on appropriate prescribing of these medications, aligning with long-standing guidance in other jurisdictions, such as the UK^27^ and accompanied by educational resources targeting GPs and community pharmacists. Among their recommendations is the restriction of benzodiazepine prescribing to the shortest possible duration and for a maximum period of 2–4 weeks for the treatment of anxiety and the avoidance of long-acting benzodiazepines for insomnia. The MMP recommends selective serotonin reuptake Inhibitors (SSRIs) as first-line pharmacological treatment for anxiety, however no specific guidance or national guidelines exist in Ireland on the prescription of the antidepressants included in this study (trazodone, doxepin, and mirtazapine), systemic antihistamines, or atypical antipsychotics in the general public, except for that they should be avoided when discontinuing long-term benzodiazepine treatment. National clinical guidelines on appropriate prescribing of psychotropic medication for non-cognitive symptoms in people with dementia recommend that prior to considering any psychotropic medication, a comprehensive assessment should be performed and that non-pharmacological interventions should be used initially to treat non-cognitive symptoms, unless there is severe distress, or an identifiable risk of harm to the person and/or others^28^. Despite the decrease in benzodiazepine and z-drug dispensings, overall sedative dispensings increased, driven by an increase in antidepressants, first-generation antihistamines, and atypical antipsychotics with sedating properties. Previous research from the UK shows an increase in prescribing of these medications^10, 11^, and our findings are in line with that.

Without information on the clinical characteristics of patients it is unclear what the implications for clinical practice are, and if this is a positive change. There is no clear evidence or guidance on the comparative effectiveness and safety of benzodiazepines and z-drugs versus these alternative sedatives, and more research is needed to examine this.

Our analysis of benzodiazepine and z-drug dispensing rates by age group and sex show higher rates for women and older people, which is a well-established pattern in large parts of the world^29, 30^.

Rates of dispensings decreased in all older age groups, with the largest decrease over the study period in the over 75 years age group. Two OECD health indicators for prescribing in primary care relate to the use of benzodiazepines: individuals aged 65 years and over receiving long-acting benzodiazepines and over 65s receiving benzodiazepines long-term, both indicators which have been found to have decreased in most OECD countries between 2009 and 2019^31^. The prevalence of benzodiazepine and z-drug initiations, discontinuations, and chronic use remained steady throughout the study period, while there was a slight decrease in the percentage of individuals with any dispensings. In the context of the decreasing rate of dispensings, this may indicate that a reduction in the frequency of dispensings, rather than a decrease in the amount of people receiving dispensings, is the main driver. The larger relative reduction in DDEs compared to dispensings suggest an additional trend towards prescribing lower quantity, strength, and/or potency of benzodiazepines and z-drugs with the implications that patients may be less at risk of dependence/tolerance developing

During the study period, England saw a substantially lower rate of dispensings for all drugs. One explanation may be the difference between the two populations, as the English data included all individuals registered with a GP practice, compared to the Irish data which was restricted to a means-tested population. Benzodiazepine and z-drug prescribing has been found to be associated with deprivation, with higher prescribing rates in GP practices in deprived areas^32^, which may contribute to the higher rates of dispensings in the GMS population. To account for differences in deprivation (albeit not in age) between Ireland and England, volume outcomes were repeated in the one third of NHS CCGs that are most deprived according to IMD 2019, with results showing that dispensing rates were not higher for most drugs in this population throughout the study period.

Although recommendations regarding benzodiazepine and z-drug prescribing, as well as the scheduling of controlled drugs, are similar in the two countries, Ireland has for a long time had higher rates of prescribing. As highlighted above, Ireland has one of the highest rates of chronic benzodiazepine and z-drug use in people aged 65 and older among OECD countries^14^, and previous studies have found higher rates of potentially inappropriate prescribing in the GMS population compared to the NHS population of Northern Ireland^33^. Hospitalisation and unintended long-term continuation of hospital initiated benzodiazepines and z-drugs also likely contribute in Ireland, potentially driven by a lack of shared care record to support transitions of care^34^. This more restrictive stance on prescribing of these drugs in the UK is also mirrored by a much more restricted set of drugs available to be prescribed in the NHS, with clorazepate, prazepam, alprazolam, flurazepam, flunitrazepam, and triazolam not dispensed at all during the study period. There are also differences in access to non-pharmacological therapies and in waiting time for mental health specialists between Ireland and UK, both which may impact on prescribing.

The main strength of our study is the data used. As GMS dispensing data are primarily used for pharmacies to be reimbursed, the data cover all prescriptions dispensed to the GMS population within the time period, and the accuracy and completeness are high. Another strength is the availability of dispensing-level data, where each dispensed medication is associated to a specific individual, allowing more detailed analysis than aggregate data. Our study does however have limitations. Firstly, the study only includes data dispensed to the means-tested GMS eligible population, and so it is not directly comparable to the NHS population in England. This was addressed through the subgroup analysis of the most deprived CCGs in England, however this may not fully account for potential differences in populations. Secondly, dispensings do not include indications for the prescription, nor do we know the medical history or diagnoses of the individuals. Our analysis of other sedative agents should therefore also be interpreted with caution as a proportion of these dispensings will be for indications unrelated to sedation. Similarly, we have no information on whether prescribing was specialist or GP initiated, as all GMS patients will have their secondary care prescriptions transcribed by their GP, and interventions aiming to reduce sedative prescribing need to take account for this. Finally, as we do not have access to eligibility start or end dates, or data on mortality, we are not eligible to account for these factors when analysing patterns of use (e.g. initiations, discontinuations, and chronic use).

The findings of this study suggest that benzodiazepine and z-drug dispensing is decreasing in Ireland, including high-risk dispensings and dispensings to older age groups. However, in comparison to England, dispensing rates remain high and suggest a need for enhanced availability of non-pharmacological services and interventions to address the two main indications of benzodiazepine and z-drug prescribing: anxiety and insomnia, as well as improved education and deprescribing support for healthcare professionals. Qualitative work may be beneficial to identify and contextualise sedative use in primary and secondary specialist care, and the impact on individuals of issues such as access to specialised mental health care and the availability of non-pharmacological intervention. An important aspect of drug utilisation research is the access to data sources.

Restrictions to data sources, including charges and lengthy application processes, may be a challenge to timely analysis and should be addressed by data governance policies, with the availability of open data when appropriate being crucial.

## Data availability

The datasets analysed during the current study are not publicly available as this was not covered by the Data Exchange Agreement between RCSI and HSE-PCRS.

