## Supplementary Material for "Trends and patterns of sedative prescribing in primary care in Ireland between 2014 and 2022 - a repeated cross-sectional study"

**Supplementary figure 1. Rate ratios for DDDs and DDEs in Ireland versus England for 2014 and 2022**

|  | 2014 | 2022 |
| --- | --- | --- |
| Sedatives | 5.96123 | 6.80072 |
| Antipsychotics3 | 4.36356 | 6.48963 |
| Antihistamines2 | 0.591735 | 1.85362 |
| Antidepressants1 | 1.92413 | 2.30591 |
| Zolpidem | 25.2106 | 42.2775 |
| Zopiclone | 5.53811 | 9.06041 |
| Temazepam | 3.04617 | 6.28097 |
| Lormetazepam | 55.7321 | 0.156935 |
| Nitrazepam | 1.29723 | 0.0797615 |
| Clobazam | 2.78192 | 1.6334 |
| Lorazepam | 1.80803 | 2.56702 |
| Chloridiazepoxide | 8.91547 | 30.3132 |
| Diazepam | 4.76015 | 7.53367 |
| Clonazepam | 3.0182 | 3.29068 |
| Long-acting benzodiazepines | 5.01632 | 6.8969 |
| Benzodiazepines and z-drugs | 7.4147 | 11.4844 |

|  | 2014 |
| --- | --- |
| 5.96123 | 0.5 |
| 4.36356 | 1.5 |
| 0.591735 | 2.5 |
| 1.92413 | 3.5 |
| 25.2106 | 4.5 |
| 5.53811 | 5.5 |
| 3.04617 | 6.5 |
| 55.7321 | 7.5 |
| 1.29723 | 8.5 |
| 2.78192 | 9.5 |
| 1.80803 | 10.5 |
| 8.91547 | 11.5 |
| 4.76015 | 12.5 |
| 3.0182 | 13.5 |
| 5.01632 | 14.5 |
| 7.4147 | 15.5 |

|  | 2022 |
| --- | --- |
| 6.80072 | 0.5 |
| 6.48963 | 1.5 |
| 1.85362 | 2.5 |
| 2.30591 | 3.5 |
| 42.2775 | 4.5 |
| 9.06041 | 5.5 |
| 6.28097 | 6.5 |
| 0.156935 | 7.5 |
| 0.0797615 | 8.5 |
| 1.6334 | 9.5 |
| 2.56702 | 10.5 |
| 30.3132 | 11.5 |
| 7.53367 | 12.5 |
| 3.29068 | 13.5 |
| 6.8969 | 14.5 |
| 11.4844 | 15.5 |

**Rate ratios for DDDs**

|  | 2014 | 2022 |
| --- | --- | --- |
| Zolpidem | 25.1969 | 42.3465 |
| Zopiclone | 5.53808 | 9.06032 |
| Temazepam | 3.0455 | 6.27449 |
| Lormetazepam | 56.2352 | 0.160695 |
| Nitrazepam | 1.29774 | 0.0797144 |
| Clobazam | 2.78219 | 1.63183 |
| Lorazepam | 1.8076 | 2.56616 |
| Chloridiazepoxide | 8.97339 | 31.2879 |
| Diazepam | 4.75959 | 7.53477 |
| Clonazepam | 3.01772 | 3.29057 |
| Long-acting benzodiazepines | 4.81769 | 5.65752 |
| Benzodiazepines and z-drugs | 6.96387 | 9.31543 |

|  | 2014 |
| --- | --- |
| 25.1969 | 0.5 |
| 5.53808 | 1.5 |
| 3.0455 | 2.5 |
| 56.2352 | 3.5 |
| 1.29774 | 4.5 |
| 2.78219 | 5.5 |
| 1.8076 | 6.5 |
| 8.97339 | 7.5 |
| 4.75959 | 8.5 |
| 3.01772 | 9.5 |
| 4.81769 | 10.5 |
| 6.96387 | 11.5 |

|  | 2022 |
| --- | --- |
| 42.3465 | 0.5 |
| 9.06032 | 1.5 |
| 6.27449 | 2.5 |
| 0.160695 | 3.5 |
| 0.0797144 | 4.5 |
| 1.63183 | 5.5 |
| 2.56616 | 6.5 |
| 31.2879 | 7.5 |
| 7.53477 | 8.5 |
| 3.29057 | 9.5 |
| 5.65752 | 10.5 |
| 9.31543 | 11.5 |

**Rate ratios for DDEs**

**Supplemental figure 2. Prevalence of initiations (90/180 days), discontinuations (90/180 days), chronic use (30/90 days), and dispensings at a dose equivalent to >40 mg diazepam per day of benzodiazepines and z-drugs, by age group and sex**

|  | <5 | 5-11 | 12-15 | 16-24 | 25-34 | 35-44 | 45-54 | 55-64 | 65-69 | 70-74 | 75+ |
| --- | --- | --- | --- | --- | --- | --- | --- | --- | --- | --- | --- |
| 2015 | 0.00100844 | 0.00100705 | 0.00312049 | 0.0385728 | 0.0952036 | 0.108454 | 0.121063 | 0.121697 | 0.116105 | 0.118154 | 0.128064 |
| 2016 | 0.00108258 | 0.00129027 | 0.00315228 | 0.0423245 | 0.112637 | 0.120753 | 0.127855 | 0.127529 | 0.118732 | 0.119271 | 0.129964 |
| 2017 | 0.000929202 | 0.00148484 | 0.00316316 | 0.0411269 | 0.117738 | 0.128756 | 0.130051 | 0.12658 | 0.114717 | 0.113352 | 0.124431 |
| 2018 | 0.000916425 | 0.00136471 | 0.00342326 | 0.0410598 | 0.121035 | 0.128903 | 0.124985 | 0.121805 | 0.113471 | 0.111584 | 0.120586 |
| 2019 | 0.000709163 | 0.00127816 | 0.0028413 | 0.0415871 | 0.119753 | 0.125854 | 0.124218 | 0.120126 | 0.11459 | 0.108044 | 0.116535 |
| 2020 | 0.000597873 | 0.00111985 | 0.00259798 | 0.0309441 | 0.102308 | 0.110527 | 0.110926 | 0.104564 | 0.100345 | 0.0961355 | 0.106676 |
| 2021 | 0.000511354 | 0.00119552 | 0.00289396 | 0.0341768 | 0.105426 | 0.114819 | 0.112024 | 0.104635 | 0.0982399 | 0.0915456 | 0.10257 |
| 2022 | 0.000430567 | 0.00123297 | 0.00243924 | 0.0294964 | 0.0967478 | 0.108796 | 0.110264 | 0.105286 | 0.0988984 | 0.0953786 | 0.103101 |

**Prevalence of initiations after >90 days by age group**

|  | <5 | 5-11 | 12-15 | 16-24 | 25-34 | 35-44 | 45-54 | 55-64 | 65-69 | 70-74 | 75+ |
| --- | --- | --- | --- | --- | --- | --- | --- | --- | --- | --- | --- |
| 2015 | 0.000692069 | 0.000698008 | 0.0024434 | 0.030991 | 0.0737646 | 0.0835702 | 0.0925983 | 0.0920351 | 0.087144 | 0.0886338 | 0.0960932 |
| 2016 | 0.000874814 | 0.00105668 | 0.00272469 | 0.0362571 | 0.0936844 | 0.0994479 | 0.10229 | 0.0998293 | 0.0910513 | 0.0905847 | 0.0992064 |
| 2017 | 0.000690945 | 0.00125505 | 0.00267975 | 0.0358885 | 0.0994302 | 0.106556 | 0.104788 | 0.099548 | 0.0886794 | 0.0867022 | 0.0952184 |
| 2018 | 0.000800258 | 0.00119258 | 0.0029496 | 0.0359466 | 0.102622 | 0.106847 | 0.101564 | 0.0967625 | 0.0882937 | 0.0861015 | 0.0930045 |
| 2019 | 0.000561978 | 0.00109735 | 0.0025529 | 0.0363623 | 0.101085 | 0.103981 | 0.101297 | 0.0957471 | 0.0893353 | 0.0843564 | 0.0899116 |
| 2020 | 0.000483993 | 0.000921864 | 0.0021819 | 0.0274183 | 0.0878751 | 0.0918858 | 0.0912965 | 0.0836966 | 0.07825 | 0.0744895 | 0.0841526 |
| 2021 | 0.000335576 | 0.00103958 | 0.00253096 | 0.030065 | 0.0905496 | 0.0976433 | 0.0934413 | 0.0857104 | 0.0779608 | 0.0723617 | 0.0817428 |
| 2022 | 0.000287045 | 0.0011025 | 0.00221302 | 0.0256808 | 0.0837527 | 0.0917702 | 0.0928904 | 0.0866425 | 0.0789169 | 0.0761089 | 0.083 |

**Prevalence of initiations after >180 days by age group**

|  | 90 days - Female | 90 days - Male | 180 days - Female | 180 days - Male |
| --- | --- | --- | --- | --- |
| 2015 | 0.0988812 | 0.0635192 | 0.0751112 | 0.048719 |
| 2016 | 0.10621 | 0.0690279 | 0.0839006 | 0.0554491 |
| 2017 | 0.106579 | 0.069924 | 0.0844589 | 0.0567853 |
| 2018 | 0.106019 | 0.0688347 | 0.084684 | 0.056052 |
| 2019 | 0.104641 | 0.0680746 | 0.0833736 | 0.0557398 |
| 2020 | 0.0922256 | 0.0598766 | 0.0740565 | 0.0495224 |
| 2021 | 0.0933372 | 0.0603 | 0.0761659 | 0.0505003 |
| 2022 | 0.0918413 | 0.0596186 | 0.0753878 | 0.0499353 |

**Prevalence of initiations after >90/180 days by sex**

|  | <5 | 5-11 | 12-15 | 16-24 | 25-34 | 35-44 | 45-54 | 55-64 | 65-69 | 70-74 | 75+ |
| --- | --- | --- | --- | --- | --- | --- | --- | --- | --- | --- | --- |
| 2015 | 0.000563542 | 0.000479547 | 0.00158968 | 0.0192893 | 0.0517736 | 0.0637771 | 0.0763074 | 0.082016 | 0.0784701 | 0.0798456 | 0.0874005 |
| 2016 | 0.00061237 | 0.000583956 | 0.00136234 | 0.020411 | 0.0591763 | 0.0675763 | 0.0783671 | 0.0841725 | 0.0813721 | 0.0823834 | 0.0891276 |
| 2017 | 0.000536078 | 0.000571546 | 0.00128208 | 0.0191623 | 0.0601987 | 0.0709556 | 0.0791626 | 0.0836123 | 0.078879 | 0.0782597 | 0.0863804 |
| 2018 | 0.000400129 | 0.000430314 | 0.00164704 | 0.0195389 | 0.0624134 | 0.0705767 | 0.0755235 | 0.08004 | 0.0758994 | 0.0762167 | 0.0835186 |
| 2019 | 0.000347891 | 0.000554908 | 0.00113225 | 0.0203791 | 0.0649747 | 0.0715881 | 0.0774321 | 0.0804114 | 0.0793341 | 0.0746107 | 0.0815582 |
| 2020 | 0.000156586 | 0.000433091 | 0.00106558 | 0.0160756 | 0.0578238 | 0.0637907 | 0.0686141 | 0.0700106 | 0.0674114 | 0.0653937 | 0.0718589 |
| 2021 | 0.000175778 | 0.000396341 | 0.00116969 | 0.0183548 | 0.0591401 | 0.0666603 | 0.0704846 | 0.0703256 | 0.0676803 | 0.0626408 | 0.0719962 |
| 2022 | 0.000334885 | 0.000437086 | 0.00103274 | 0.0159261 | 0.0542036 | 0.0627064 | 0.0688452 | 0.0710256 | 0.069071 | 0.067564 | 0.0730485 |

**Prevalence of discontinuations after >90 days by age group**

|  | <5 | 5-11 | 12-15 | 16-24 | 25-34 | 35-44 | 45-54 | 55-64 | 65-69 | 70-74 | 75+ |
| --- | --- | --- | --- | --- | --- | --- | --- | --- | --- | --- | --- |
| 2015 | 0.000306488 | 0.000261087 | 0.00104998 | 0.0120132 | 0.0323815 | 0.0399894 | 0.0479277 | 0.0497118 | 0.0480024 | 0.0478572 | 0.0523597 |
| 2016 | 0.00029525 | 0.000328128 | 0.000865138 | 0.0129567 | 0.0382747 | 0.0438959 | 0.050719 | 0.0526167 | 0.0494844 | 0.0499214 | 0.0540837 |
| 2017 | 0.000238257 | 0.000353534 | 0.000798672 | 0.0126821 | 0.0402469 | 0.0463813 | 0.0512393 | 0.0532218 | 0.0488052 | 0.0483781 | 0.0535414 |
| 2018 | 0.000219426 | 0.000282778 | 0.00100114 | 0.0129523 | 0.0420357 | 0.0462321 | 0.049631 | 0.0513428 | 0.0479387 | 0.0466055 | 0.0524803 |
| 2019 | 0.000200706 | 0.000374095 | 0.000790438 | 0.0138055 | 0.044297 | 0.0474387 | 0.050782 | 0.0518895 | 0.0489768 | 0.046391 | 0.0504609 |
| 2020 | 9.96455e-05 | 0.000296976 | 0.000700237 | 0.0116077 | 0.0418298 | 0.0435518 | 0.0469369 | 0.0469974 | 0.0439117 | 0.0417761 | 0.0465511 |
| 2021 | 6.39192e-05 | 0.00029888 | 0.000897431 | 0.0130095 | 0.042367 | 0.045911 | 0.0476861 | 0.0467198 | 0.0438116 | 0.0387254 | 0.0462962 |
| 2022 | 0.000191363 | 0.000300089 | 0.000708167 | 0.0114718 | 0.0385888 | 0.0434057 | 0.0470347 | 0.0464715 | 0.0440216 | 0.0426004 | 0.0476152 |

**Prevalence of discontinuations after >180 days by age group**

|  | 90 days - Female | 90 days - Male | 180 days - Female | 180 days - Male |
| --- | --- | --- | --- | --- |
| 2015 | 0.0634045 | 0.0389769 | 0.0386483 | 0.0242209 |
| 2016 | 0.0672337 | 0.0413694 | 0.0419219 | 0.026273 |
| 2017 | 0.0674495 | 0.0417226 | 0.0426411 | 0.0269972 |
| 2018 | 0.0668262 | 0.041038 | 0.042547 | 0.0268199 |
| 2019 | 0.067889 | 0.0416179 | 0.0433707 | 0.0270726 |
| 2020 | 0.0594902 | 0.0364352 | 0.0395239 | 0.0248937 |
| 2021 | 0.0612282 | 0.037222 | 0.0405476 | 0.0249413 |
| 2022 | 0.0608036 | 0.0371229 | 0.0403004 | 0.0249515 |

**Prevalence of discontinuations after >90/180 days by sex**

|  | <5 | 5-11 | 12-15 | 16-24 | 25-34 | 35-44 | 45-54 | 55-64 | 65-69 | 70-74 | 75+ |
| --- | --- | --- | --- | --- | --- | --- | --- | --- | --- | --- | --- |
| 2015 | 0.000711842 | 0.000996393 | 0.00181538 | 0.0209585 | 0.0678918 | 0.0943064 | 0.130952 | 0.179762 | 0.200933 | 0.206333 | 0.277558 |
| 2016 | 0.000677981 | 0.00100107 | 0.00171039 | 0.0208091 | 0.0728768 | 0.0971068 | 0.131254 | 0.176265 | 0.200871 | 0.204717 | 0.274978 |
| 2017 | 0.000655207 | 0.00100757 | 0.00171294 | 0.0196305 | 0.0758621 | 0.102528 | 0.133358 | 0.17399 | 0.193337 | 0.196621 | 0.261784 |
| 2018 | 0.000619555 | 0.000934396 | 0.00180851 | 0.0198999 | 0.0784774 | 0.103573 | 0.12592 | 0.167973 | 0.192112 | 0.190276 | 0.256011 |
| 2019 | 0.000495076 | 0.000904063 | 0.00152747 | 0.0193797 | 0.0775694 | 0.102027 | 0.124431 | 0.16366 | 0.1888 | 0.186741 | 0.247078 |
| 2020 | 0.000669049 | 0.000810498 | 0.00149181 | 0.015921 | 0.0717309 | 0.095994 | 0.120305 | 0.157067 | 0.180879 | 0.18127 | 0.239548 |
| 2021 | 0.000543313 | 0.000903137 | 0.00137135 | 0.0172611 | 0.0726241 | 0.0970917 | 0.1202 | 0.154453 | 0.179866 | 0.177062 | 0.231884 |
| 2022 | 0.00062193 | 0.000835029 | 0.00132781 | 0.0152293 | 0.0638896 | 0.0910219 | 0.113844 | 0.145292 | 0.168034 | 0.167833 | 0.222405 |

**Prevalence of chronic use >30 days by age group**

|  | <5 | 5-11 | 12-15 | 16-24 | 25-34 | 35-44 | 45-54 | 55-64 | 65-69 | 70-74 | 75+ |
| --- | --- | --- | --- | --- | --- | --- | --- | --- | --- | --- | --- |
| 2015 | 0.000395468 | 0.000682023 | 0.000961661 | 0.0108554 | 0.0437089 | 0.0672736 | 0.10056 | 0.148063 | 0.17141 | 0.175282 | 0.244856 |
| 2016 | 0.000360861 | 0.000689624 | 0.000825361 | 0.0111911 | 0.0474125 | 0.0692646 | 0.100444 | 0.144237 | 0.170268 | 0.174287 | 0.242228 |
| 2017 | 0.000381211 | 0.000642253 | 0.000830198 | 0.0108025 | 0.0507452 | 0.0738323 | 0.103159 | 0.14229 | 0.164174 | 0.16885 | 0.231211 |
| 2018 | 0.000361407 | 0.000534819 | 0.00111956 | 0.0110146 | 0.0528646 | 0.0752474 | 0.0978678 | 0.138107 | 0.163741 | 0.162849 | 0.226125 |
| 2019 | 0.000280989 | 0.000586082 | 0.000790438 | 0.0106852 | 0.0528459 | 0.0741805 | 0.0962491 | 0.13571 | 0.161659 | 0.160455 | 0.219141 |
| 2020 | 0.000412817 | 0.000556831 | 0.000730683 | 0.00913454 | 0.0490171 | 0.070386 | 0.0942 | 0.131281 | 0.154881 | 0.156534 | 0.212438 |
| 2021 | 0.000383515 | 0.000636744 | 0.000675594 | 0.00995864 | 0.0498848 | 0.0714264 | 0.0942704 | 0.128795 | 0.154393 | 0.153072 | 0.206958 |
| 2022 | 0.000382726 | 0.000469704 | 0.000737673 | 0.00862662 | 0.0438368 | 0.0673385 | 0.0896161 | 0.121046 | 0.14438 | 0.14456 | 0.197568 |

**Prevalence of chronic use >90 days by age group**

|  | 30 days - Female | 30 days - Male | 90 days - Female | 90 days - Male |
| --- | --- | --- | --- | --- |
| 2015 | 0.135298 | 0.0855037 | 0.111115 | 0.068094 |
| 2016 | 0.138488 | 0.0879859 | 0.113548 | 0.0702113 |
| 2017 | 0.139562 | 0.0895865 | 0.115191 | 0.0721637 |
| 2018 | 0.139866 | 0.0895547 | 0.115805 | 0.0725982 |
| 2019 | 0.138507 | 0.0889925 | 0.115199 | 0.0725982 |
| 2020 | 0.133277 | 0.0846701 | 0.111363 | 0.0695843 |
| 2021 | 0.133984 | 0.0855034 | 0.112199 | 0.070716 |
| 2022 | 0.126587 | 0.082182 | 0.106257 | 0.0677113 |

**Prevalence of chronic use >30/90 days by sex**

|  | <5 | 5-11 | 12-15 | 16-24 | 25-34 | 35-44 | 45-54 | 55-64 | 65-69 | 70-74 | 75+ |
| --- | --- | --- | --- | --- | --- | --- | --- | --- | --- | --- | --- |
| 2014 | 0.000381572 | 0.000276827 | 0.000285711 | 0.00218757 | 0.0108286 | 0.015322 | 0.0178036 | 0.0203505 | 0.0186405 | 0.014336 | 0.0118033 |
| 2015 | 0.000237281 | 0.000223789 | 0.000304199 | 0.00218048 | 0.0109593 | 0.0149444 | 0.0171826 | 0.0192711 | 0.0169323 | 0.0138551 | 0.0110187 |
| 2016 | 0.000207768 | 0.000216898 | 0.000258547 | 0.00243982 | 0.0115946 | 0.0152536 | 0.017215 | 0.0182082 | 0.0165086 | 0.0133949 | 0.0109714 |
| 2017 | 0.00016678 | 0.000182659 | 0.000241703 | 0.002341 | 0.0123977 | 0.0165449 | 0.0178202 | 0.0183054 | 0.0153474 | 0.0127163 | 0.00983097 |
| 2018 | 0.000167796 | 0.000165978 | 0.000258359 | 0.0022766 | 0.0133976 | 0.0166141 | 0.0169701 | 0.0176756 | 0.0148627 | 0.0117777 | 0.00934977 |
| 2019 | 0.000120424 | 0.000205752 | 0.000245677 | 0.00214517 | 0.0132589 | 0.0166591 | 0.0163913 | 0.0170288 | 0.0144712 | 0.0116293 | 0.00924313 |
| 2020 | 0.000156586 | 0.000179423 | 0.000345045 | 0.00184016 | 0.0111774 | 0.0150499 | 0.0158381 | 0.0164186 | 0.0141084 | 0.0108603 | 0.00861968 |
| 2021 | 0.000143818 | 0.000253398 | 0.000211753 | 0.00186673 | 0.0108422 | 0.0146222 | 0.0156099 | 0.0155261 | 0.013664 | 0.0102447 | 0.00821339 |
| 2022 | 0.000111628 | 0.000208757 | 0.000216384 | 0.00188293 | 0.00961704 | 0.0139143 | 0.0151816 | 0.0148863 | 0.0131635 | 0.0100368 | 0.00805995 |

**Prevalence of dispensings at a dose equivalent to >40 mg diazepam per day by age group**

|  | Female | Male |
| --- | --- | --- |
| 2014 | 0.0110295 | 0.00970801 |
| 2015 | 0.0108053 | 0.00943293 |
| 2016 | 0.0108678 | 0.00964359 |
| 2017 | 0.0107501 | 0.00992893 |
| 2018 | 0.0105853 | 0.00980116 |
| 2019 | 0.0104751 | 0.00961386 |
| 2020 | 0.00977704 | 0.00894992 |
| 2021 | 0.00942604 | 0.00884889 |
| 2022 | 0.00895294 | 0.00868159 |

**Prevalence of dispensings at a dose equivalent to >40 mg diazepam per day by sex**

**Supplementary table 1. Drugs included in the analysis**

|  | **ATC-code** | **BNF-code** |
| --- | --- | --- |
| **Benzodiazepines and Z-drug hypnotics** |  |  |
| ***Antiepileptics*** |  |  |
| Clonazepam | N03AE01* | 0408010F0 |
| ***Anxiolytics*** |  |  |
| Diazepam | N05BA01* | 0401020K0 |
| Chloridiazepoxide | N05BA02* | 0401020E0 |
| Potassium Clorazepate | N05BA05* | 0401020V0 |
| Lorazepam | N05BA06 | 0401020P0 |
| Bromazepam | N05BA08 | 0401020G0 |
| Clobazam | N05BA09* | 040801060 |
| Prazepam | N05BA11* | 0401020U0 |
| Alprazolam | N05BA12 | 0401020A0 |
| ***Hypnotics and sedatives*** |  |  |
| Flurazepam | N05CD01* | 0401010L0 |
| Nitrazepam | N05CD02* | 0401010R0 |
| Flunitrazepam | N05CD03* | 0401010I0 |
| Triazolam | N05CD05 | 0401010V0 |
| Lormetazepam | N05CD06 | 0401010P0 |
| Temazepam | N05CD07 | 0401010T0 |
| Midazolam | N05CD08 | 0401010Q0 |
| Zopiclone | N05CF01 | 0401010Z0 |
| Zolpidem | N05CF02 | 0401010Y0 |
| **Other sedating agents** |  |  |
| ***Antidepressants*** |  |  |
| Amitriptyline | N06AA09 | 0403010B0 |
| Doxepin | N06AA12 | 0403010L0 |
| Trazodone | N06AX05 | 0403010X0 |
| Mirtazapine | N06AX11 | 0403040X0 |
| ***Antihistamines for systemic use*** |  |  |
| Promethazine | R06AD02 | 0304010W0 |
| Cyclizine | R06AE03 | 0406000F0,  0406000G0 |
| Ketotifen | R06AX17 | 0304010AG |
| ***Atypical antipsychotics*** |  |  |
| Olanzapine | N05AH03 | 040201060 |
| Quetiapine | N05AH04 | 0402010AB |
| Risperidone | N05AX08 | 040201030 |

*Long-acting benzodiazepines

**Supplementary table 2. Diazepam Dose Equivalent (DDE)^1^**

| **Drug** | **DDE equivalent** |
| --- | --- |
| Clonazepam | 20 |
| Diazepam | 1 |
| Chloridiazepoxide | 0.4 |
| Potassium Clorazepate | 0.7 |
| Lorazepam | 10 |
| Bromazepam | 1.8 |
| Clobazam | 0.5 |
| Prazepam | 0.7 |
| Alprazolam | 20 |
| Flurazepam | 0.4 |
| Nitrazepam | 1 |
| Flunitrazepam | 1 |
| Triazolam | 20 |
| Lormetazepam | 6.7 |
| Temazepam | 0.5 |
| Zopiclone | 0.7 |
| Zolpidem | 0.5 |

^1^Benzo.org.uk. Benzodiazepine Equivalency Table. https://www.benzo.org.uk/bzequiv.htm

**Supplementary Table 3. Overview of outcomes**

| **Outcomes** | **Definition** |
| --- | --- |
| **(i) Volume of dispensing outcomes** | **Derived for Ireland and England** |
| Dispensings | Rate of prescription dispensings per 1,000 population |
| Standard daily doses | Rate of dosage units, multiplied by the strength in standard units (i.e. Diazepam Dose Equivalents and WHO Defined Daily Doses) per 1,000 population |
| Cost | Rate of costs to the Health Service Executive/National Health Services per 1,000 population |
| **(ii) Pattern of dispensing outcomes** | **Derived for Ireland only** |
| Volume of dispensing outcomes by age and sex sub-groups. | As above. |
| Prevalence of use | Proportion of individuals dispensed a relevant medicine |
| Prevalence of initiations | Proportion of individuals dispensed a relevant medicine with no use in the previous 90 or 180 days |
| Prevalence of discontinuations | Proportion of individuals dispensed a relevant medicine with no further dispensings in the following 90 or 180 days |
| Chronic use | Proportion of individuals dispensed a relevant medicine for >30 days and for >90 days, calculated as a rolling sum over the last 30/90 days, where two or more dispensings in 30 days and four or more dispensings in 90 days equate chronic use |
| **(iii) High-risk dispensings^a^** | **Derived for Ireland only** |
| Benzodiazepines/z-drugs + medication increasing the risk of falls, fractures, and delirium | Proportion of individuals aged ≥65 years dispensed a benzodiazepine/z-drug within 30 days of being dispensed two or more other sedating or anticholinergic^b^ drugs |
| Benzodiazepines/z-drugs at a dose increasing the risk of dependency | Proportion of individuals dispensed the dose equivalent to >40 mg diazepam per day^d^, defined as the 30-day average DDE exceeding 40 |

^a^based on Scottish Polypharmacy Guidance^1^

^b^anticholinergic drugs identified using the Anticholinergic Cognitive Burden (ACB) scale^2^

1.	Polypharmacy Guidance, Realistic Prescribing (Scottish Government) (2018).

2.	Boustani M, Campbell N, Munger S, Maidment I, Fox C. Impact of anticholinergics on the aging brain: a review and practical application. Aging Health. 2008/06/01 2008;4(3):311-320. doi:10.2217/1745509X.4.3.311

**Supplementary Table 4. Anticholinergic drugs^1^**

| **Score 1** | | **Score 2** | | **Score 3** | |
| --- | --- | --- | --- | --- | --- |
| **Drug** | **ATC** | **Drug** | **ATC** | **Drug** | **ATC** |
| Alimemazine | R06AD01 | Amantadine | N04BB01 | Amitriptyline | N06AA09 |
| Alverine | A03AX08, A03AX58 | Belladonna alkaloids | A03BA01, A03BA04,  A03BB01, A03BB02,  A03BB03, A03BB04,  A03BB05, A03BB06,  A06AB30 | Amoxapine | N06AA17 |
| Alprazolam | N05BA12 | Carbamazepine | N03AF01 | Atropine | A03BA01, A03CB03 |
| Atenolol | C07AB03, C07FB03,  C07CB03, C07CB53,  C07BB03, C07DB01 | Cyclobenzaprine | M03BX08 | Benzatropine | N04AC01 |
| Brompheniramine maleate | R06AB01, R06AB51 | Cyproheptadine | R06AX02 | Brompheniramine | R06AB01, R06AB51 |
| Bupropion hydrochloride | N06AX12, A08AA62 | Levomepromazine | N05AA02 | Carbinoxamine | R06AA08 |
| Captopri | C09AA01, C09BA01 | Loxapine | N05AH01 | Chlorphenamine | R06AB04, R06AB54 |
| Chlortalidone | C03BA04, C03BB04,  C03EA06 | Molindone | N05AE02 | Chlorpromazine | N05AA01 |
| Cimetidine hydrochloride | A02BA01, A02BA51 | Oxcarbazepine | N03AF02 | Clemastine | R06AA04, R06AA54 |
| Clorazepate | N05BA05 | Pethidine hydrochloride | N02AB02, N02AG03,  N02AB52, N02AB72 | Clomipramine | N06AA04 |
| Codeine | R05DA04, N02AJ06,  N02AJ07, N02AJ08,  N02AJ09, N02AA59,  N02AA79, | Pimozide | N05AG02 | Clozapine | N05AH02 |
| Colchicine | M04AC01 |  |  | Darifenacin | G04BD10 |
| Diazepam | N05BA01 |  |  | Desipramine | N06AA01 |
| Digoxin | C01AA05 |  |  | Dicycloverine | A03AA07 |
| Dipyridamole | B01AC07 |  |  | Dimenhydrinate | R06AA11 |
| Disopyramide phosphate | C01BA03 |  |  | Diphenhydramine | R06AA02, R06AA52 |
| Fentanyl | N02AB03 |  |  | Doxepin | N06AA12 |
| Furosemide | C03CA01, C03CB01,  C03EB01 |  |  | Flavoxate | G04BD02 |
| Fluvoxamine | N06AB08 |  |  | Hydroxyzine | N05BB01,N05BB51 |
| Haloperidol | N05AD01 |  |  | Hyoscyamine | A03BA03, A03CB31 |
| Hydralazine | C02DB02, C02LG02 |  |  | Imipramine | N06AA02, N06AA03 |
| Hydrocortisone | H02AB09 |  |  | Mepyramine | R06AC01, R03DA12 |
| Isosorbide | C01DA08, C01DA58,  C05AE02, C01DA14 |  |  | Meclozine | R06AE05, R06AE55 |
| Loperamide | A07DA03, A07DA05,  A07DA53 |  |  | Nortriptyline | N06AA10 |
| Metoprolol | C07AB02, C07FX03,  C07FB13, C07FB02,  C07FX05, C07CB02,  C07BB02, C07BB52 |  |  | Olanzapine | N05AH03 |
| Morphine | N02AA01, N02AA51,  N02AG01, A07DA52,  R05DA05 |  |  | Orphenadrine | N04AB02, M03BC01,  M03BC51 |
| Nifedipine | C08CA05, C07FB03,  C08GA01, C08CA55 |  |  | Oxybutynin | G04BD04 |
| Prednisone | H02AB07, A07EA03 |  |  | Paroxetine | N06AB05 |
| Quinidine | C01BA01, C01BA51,  C01BA71 |  |  | Perphenazine | N05AB03 |
| Ranitidine | A02BA02, A02BA07 |  |  | Procyclidine | N04AA04 |
| Risperidone | N05AX08 |  |  | Promazine | N05AA03 |
| Theophylline | R03DA04, R03DB04,  R03DA54, R03DA74 |  |  | Promethazine | R06AD02, R06AD52, |
| Trazodone | N06AX05 |  |  | Propantheline | A03AB05, A03CA34 |
| Triamterene | C03DB02 |  |  | Quetiapine | N05AH04 |
| Warfarin | B01AA03 |  |  | Scopolamine | A04AD01, A04AD51,  N05CM05 |
|  |  |  |  | Thioridazine | N05AC02 |
|  |  |  |  | Tolterodine | G04BD07 |
|  |  |  |  | Trifluoperazine | N05AB06 |
|  |  |  |  | Trihexyphenidy | N04AA01 |
|  |  |  |  | Trimipramine | N06AA06 |

^1^Boustani M, Campbell N, Munger S, Maidment I, Fox C. Impact of anticholinergics on the aging brain: a review and practical application. *Aging Health.* 2008/06/01 2008;4(3):311-320. doi:10.2217/1745509X.4.3.311

**Supplementary Table 5. GMS (Ireland) and NHS (England) populations**

|  | **GMS** | **NHS** |
| --- | --- | --- |
| **2014** | 1,768,700 | 56,545,892 |
| **2015** | 1,734,853 | 57,111,235 |
| **2016** | 1,683,792 | 57,744,814 |
| **2017** | 1,609,820 | 58,492,541 |
| **2018** | 1,565,049 | 59,178,163 |
| **2019** | 1,544,374 | 59,901,236 |
| **2020** | 1,584,790 | 60,413,787 |
| **2021** | 1,545,222 | 60,970,002 |
| **2022** | 1,568,379 | 61,768,942 |

**Supplementary Table 5. Rate of dispensings per 1,000 NHS population**

|  | **2014** | **2015** | **2016** | **2017** | **2018** | **2019** | **2020** | **2021** | **2022** | **Absolute change** | **Relative change** |
| --- | --- | --- | --- | --- | --- | --- | --- | --- | --- | --- | --- |
| Benzodiazepines and z-drugs | 288.0 | 278.8 | 270.9 | 260.1 | 248.5 | 238.1 | 230.3 | 220.9 | 210.1 | -77.83 | -0.27 |
| Long-acting benzodiazepines | 126.6 | 124.2 | 122.2 | 118.1 | 113.9 | 110.1 | 106.0 | 101.4 | 98.7 | -27.90 | -0.22 |
| Clonazepam | 14.8 | 15.4 | 15.9 | 16.0 | 16.2 | 16.5 | 16.8 | 16.7 | 16.7 | 1.89 | 0.13 |
| Diazepam | 92.4 | 91.2 | 89.8 | 87.0 | 83.5 | 80.3 | 76.6 | 72.8 | 70.5 | -21.94 | -0.24 |
| Chloridiazepoxide | 2.1 | 1.5 | 1.3 | 1.1 | 0.9 | 0.8 | 0.7 | 0.6 | 0.5 | -1.58 | -0.76 |
| Lorazepam | 18.5 | 18.6 | 18.7 | 18.3 | 17.9 | 17.7 | 18.0 | 17.5 | 17.1 | -1.39 | -0.08 |
| Clobazam | 4.4 | 4.7 | 4.9 | 5.0 | 5.2 | 5.4 | 5.6 | 5.8 | 6.0 | 1.62 | 0.37 |
| Nitrazepam | 13.0 | 11.5 | 10.2 | 9.0 | 8.0 | 7.1 | 6.3 | 5.5 | 5.1 | -7.89 | -0.61 |
| Lormetazepam | 0.6 | 0.5 | 0.4 | 0.4 | 0.3 | 0.3 | 0.2 | 0.2 | 0.2 | -0.37 | -0.65 |
| Temazepam | 29.5 | 24.7 | 21.8 | 19.1 | 16.8 | 14.8 | 13.3 | 11.8 | 9.0 | -20.46 | -0.69 |
| Zopiclone | 99.9 | 98.0 | 95.4 | 92.5 | 88.6 | 84.6 | 82.5 | 80.0 | 75.6 | -24.32 | -0.24 |
| Zolpidem | 13.0 | 12.7 | 12.4 | 11.7 | 11.1 | 10.6 | 10.2 | 9.9 | 9.6 | -3.40 | -0.26 |
| Antidepressants^1^ | 49.3 | 54.1 | 59.0 | 63.2 | 68.5 | 73.6 | 79.3 | 84.8 | 88.1 | 38.83 | 0.79 |
| Antihistamines^2^ | 25.3 | 28.4 | 31.3 | 33.8 | 35.7 | 37.6 | 44.0 | 49.2 | 52.6 | 27.25 | 1.08 |
| Antipsychotics^3^ | 29.0 | 31.0 | 33.3 | 35.1 | 36.1 | 37.5 | 39.5 | 39.9 | 40.0 | 11.05 | 0.38 |
| Sedatives | 391.5 | 392.4 | 394.5 | 392.2 | 388.9 | 386.8 | 393.0 | 394.8 | 390.8 | -0.71 | 0.00 |

^1^doxepin, trazodone, mirtazapine

^2^promethazine, cyclizine, ketotifen

^3^olanzapine, quetiapine, risperidone

**Supplementary Table 7. Rate of cost, DDEs, and DDDs per 1,000 GMS (Ireland) and NHS (England) populations**

**Rate of cost per 1,000 GMS population**

|  | **2014** | **2015** | **2016** | **2017** | **2018** | **2019** | **2020** | **2021** | **2022** | **Absolute change** | **Relative change** |
| --- | --- | --- | --- | --- | --- | --- | --- | --- | --- | --- | --- |
| Benzodiazepines and z-drugs | 10242.28 | 10047.37 | 10324.70 | 10480.79 | 10676.62 | 10703.11 | 10293.09 | 10945.72 | 10351.45 | 109.17 | 0.01 |
| Long-acting benzodiazepines | 2960.31 | 3105.57 | 3195.31 | 3223.15 | 3263.81 | 3257.90 | 3155.87 | 3438.58 | 3227.92 | 267.61 | 0.09 |
| Clonazepam | 389.47 | 396.67 | 422.76 | 452.56 | 464.45 | 466.73 | 450.75 | 506.12 | 484.84 | 95.36 | 0.24 |
| Diazepam | 1369.10 | 1515.26 | 1577.01 | 1610.61 | 1656.19 | 1676.00 | 1667.03 | 1890.76 | 1802.98 | 433.88 | 0.32 |
| Chloridiazepoxide | 114.96 | 111.33 | 114.57 | 110.62 | 110.58 | 112.41 | 103.48 | 113.12 | 97.84 | -17.12 | -0.15 |
| Potassium Clorazepate | 1.54 | 1.60 | 1.36 | 1.68 | 1.88 | 2.00 | 1.82 | 1.64 | 1.62 | 0.08 | 0.05 |
| Lorazepam | 142.27 | 143.56 | 147.24 | 150.32 | 156.05 | 161.53 | 161.86 | 176.00 | 173.70 | 31.43 | 0.22 |
| Bromazepam | 390.99 | 386.46 | 383.00 | 382.32 | 388.94 | 384.90 | 363.95 | 361.26 | 337.40 | -53.60 | -0.14 |
| Clobazam | 138.75 | 121.01 | 124.88 | 112.08 | 113.18 | 122.44 | 138.29 | 163.02 | 178.67 | 39.92 | 0.29 |
| Prazepam | 64.57 | 62.47 | 61.38 | 60.39 | 60.45 | 58.24 | 48.80 | 49.78 | 29.33 | -35.24 | -0.55 |
| Alprazolam | 1147.40 | 1168.56 | 1199.05 | 1230.21 | 1278.55 | 1290.72 | 1241.87 | 1312.39 | 1252.87 | 105.48 | 0.09 |
| Flurazepam | 813.37 | 833.19 | 831.78 | 816.36 | 802.87 | 766.10 | 695.63 | 694.82 | 630.58 | -182.79 | -0.22 |
| Nitrazepam | 68.54 | 64.04 | 61.59 | 58.86 | 54.20 | 53.98 | 50.07 | 19.32 | 2.05 | -66.49 | -0.97 |
| Triazolam | 298.52 | 308.54 | 314.53 | 321.64 | 319.16 | 319.17 | 309.45 | 318.94 | 300.07 | 1.55 | 0.01 |
| Lormetazepam | 217.73 | 205.75 | 202.49 | 196.20 | 187.90 | 104.65 | 8.08 | 0.77 | 0.25 | -217.48 | -1.00 |
| Temazepam | 449.23 | 426.42 | 414.94 | 402.94 | 381.37 | 367.06 | 337.08 | 336.28 | 299.32 | -149.91 | -0.33 |
| Zopiclone | 1654.17 | 1682.15 | 1760.71 | 1817.91 | 1888.31 | 1942.83 | 1924.37 | 2031.84 | 1934.66 | 280.48 | 0.17 |
| Zolpidem | 1187.06 | 1292.03 | 1298.30 | 1337.31 | 1462.89 | 1563.63 | 1668.58 | 1944.33 | 1991.94 | 804.88 | 0.68 |
| Antidepressants^1^ | 62.93 | 81.70 | 110.02 | 142.73 | 179.08 | 235.01 | 298.14 | 410.09 | 430.40 | 367.47 | 5.84 |
| Antihistamines^2^ | 1984.62 | 1975.44 | 2012.66 | 1638.97 | 1799.19 | 1978.68 | 2117.54 | 2372.19 | 2339.54 | 354.92 | 0.18 |
| Antipsychotics^3^ | 10242.28 | 10047.37 | 10324.70 | 10480.79 | 10676.62 | 10703.11 | 10293.09 | 10945.72 | 10351.45 | 109.17 | 0.01 |
| Sedatives | 13476.89 | 13396.54 | 13745.67 | 13599.79 | 14117.77 | 14480.43 | 14377.34 | 15672.34 | 15113.33 | 1636.44 | 0.12 |

**Rate of DDE per 1,000 GMS population per day**

|  | **2014** | **2015** | **2016** | **2017** | **2018** | **2019** | **2020** | **2021** | **2022** | **Absolute change** | **Relative change** |
| --- | --- | --- | --- | --- | --- | --- | --- | --- | --- | --- | --- |
| Benzodiazepines and z-drugs | 836.22 | 827.50 | 847.28 | 859.57 | 861.57 | 851.83 | 813.10 | 811.03 | 761.72 | -74.499 | -0.089 |
| Long-acting benzodiazepines | 297.35 | 294.27 | 300.96 | 307.13 | 309.07 | 305.14 | 291.76 | 292.44 | 268.73 | -28.616 | -0.096 |
| Clonazepam | 73.93 | 74.95 | 79.94 | 86.16 | 89.68 | 87.97 | 86.33 | 89.89 | 78.05 | 4.118 | 0.056 |
| Diazepam | 136.08 | 137.01 | 140.52 | 143.06 | 144.35 | 145.47 | 140.63 | 142.17 | 138.19 | 2.111 | 0.016 |
| Chloridiazepoxide | 6.64 | 6.29 | 6.41 | 6.22 | 5.89 | 5.86 | 5.22 | 5.20 | 4.69 | -1.947 | -0.293 |
| Potassium Clorazepate | 0.08 | 0.08 | 0.07 | 0.08 | 0.09 | 0.10 | 0.09 | 0.08 | 0.08 | 0.000 | -0.003 |
| Lorazepam | 31.54 | 30.83 | 31.53 | 31.92 | 32.59 | 33.39 | 33.51 | 34.26 | 33.62 | 2.074 | 0.066 |
| Bromazepam | 27.10 | 26.34 | 26.20 | 25.75 | 25.33 | 24.49 | 22.97 | 22.21 | 20.78 | -6.318 | -0.233 |
| Clobazam | 7.60 | 6.69 | 6.34 | 6.11 | 5.85 | 5.63 | 5.42 | 5.51 | 5.63 | -1.966 | -0.259 |
| Prazepam | 8.03 | 7.78 | 7.63 | 7.33 | 7.13 | 6.75 | 5.68 | 5.46 | 3.22 | -4.811 | -0.599 |
| Alprazolam | 186.30 | 184.86 | 190.17 | 192.89 | 193.43 | 192.13 | 182.31 | 179.40 | 170.10 | -16.199 | -0.087 |
| Flurazepam | 58.29 | 55.46 | 54.28 | 52.75 | 51.09 | 48.57 | 44.00 | 42.53 | 38.72 | -19.571 | -0.336 |
| Nitrazepam | 6.70 | 6.01 | 5.77 | 5.43 | 5.00 | 4.79 | 4.39 | 1.59 | 0.15 | -6.550 | -0.978 |
| Triazolam | 19.71 | 19.77 | 19.83 | 19.79 | 19.66 | 19.50 | 18.66 | 18.31 | 17.15 | -2.557 | -0.130 |
| Lormetazepam | 16.31 | 15.18 | 14.90 | 14.31 | 13.41 | 7.35 | 0.59 | 0.05 | 0.02 | -16.292 | -0.999 |
| Temazepam | 40.26 | 36.93 | 35.32 | 33.53 | 31.42 | 29.87 | 27.55 | 26.26 | 23.59 | -16.669 | -0.414 |
| Zopiclone | 133.25 | 134.69 | 139.69 | 143.13 | 143.90 | 145.83 | 142.53 | 143.91 | 137.54 | 4.289 | 0.032 |
| Zolpidem | 84.41 | 84.63 | 88.68 | 91.13 | 92.75 | 94.12 | 93.24 | 94.20 | 90.20 | 5.788 | 0.069 |

**Rate of DDD per 1,000 GMS population per day**

|  | **2014** | **2015** | **2016** | **2017** | **2018** | **2019** | **2020** | **2021** | **2022** | **Absolute change** | **Relative change** |
| --- | --- | --- | --- | --- | --- | --- | --- | --- | --- | --- | --- |
| Benzodiazepines and z-drugs | 86.71 | 85.99 | 88.06 | 89.10 | 89.09 | 88.37 | 84.50 | 84.01 | 79.71 | -7.005 | -0.081 |
| Long-acting benzodiazepines | 21.97 | 21.56 | 21.77 | 21.81 | 21.67 | 21.48 | 20.40 | 19.89 | 18.68 | -3.292 | -0.150 |
| Clonazepam | 0.46 | 0.47 | 0.50 | 0.54 | 0.56 | 0.55 | 0.54 | 0.56 | 0.49 | 0.026 | 0.056 |
| Diazepam | 13.61 | 13.70 | 14.05 | 14.31 | 14.43 | 14.55 | 14.06 | 14.22 | 13.82 | 0.211 | 0.016 |
| Chloridiazepoxide | 0.55 | 0.52 | 0.53 | 0.52 | 0.49 | 0.49 | 0.44 | 0.43 | 0.39 | -0.162 | -0.293 |
| Potassium Clorazepate | 0.01 | 0.01 | 0.01 | 0.01 | 0.01 | 0.01 | 0.01 | 0.01 | 0.01 | 0.000 | -0.003 |
| Lorazepam | 1.26 | 1.23 | 1.26 | 1.28 | 1.30 | 1.34 | 1.34 | 1.37 | 1.34 | 0.083 | 0.066 |
| Bromazepam | 1.51 | 1.46 | 1.46 | 1.43 | 1.41 | 1.36 | 1.28 | 1.23 | 1.15 | -0.351 | -0.233 |
| Clobazam | 0.76 | 0.67 | 0.63 | 0.61 | 0.59 | 0.56 | 0.54 | 0.55 | 0.56 | -0.197 | -0.259 |
| Prazepam | 0.38 | 0.37 | 0.36 | 0.35 | 0.34 | 0.32 | 0.27 | 0.26 | 0.15 | -0.229 | -0.599 |
| Alprazolam | 9.31 | 9.24 | 9.51 | 9.64 | 9.67 | 9.61 | 9.12 | 8.97 | 8.50 | -0.810 | -0.087 |
| Flurazepam | 4.86 | 4.62 | 4.52 | 4.40 | 4.26 | 4.05 | 3.67 | 3.54 | 3.23 | -1.631 | -0.336 |
| Nitrazepam | 1.34 | 1.20 | 1.15 | 1.09 | 1.00 | 0.96 | 0.88 | 0.32 | 0.03 | -1.310 | -0.978 |
| Triazolam | 3.94 | 3.95 | 3.97 | 3.96 | 3.93 | 3.90 | 3.73 | 3.66 | 3.43 | -0.511 | -0.130 |
| Lormetazepam | 2.43 | 2.27 | 2.22 | 2.14 | 2.00 | 1.10 | 0.09 | 0.01 | 0.00 | -2.432 | -0.999 |
| Temazepam | 4.03 | 3.69 | 3.53 | 3.35 | 3.14 | 2.99 | 2.75 | 2.63 | 2.36 | -1.667 | -0.414 |
| Zopiclone | 16.88 | 16.93 | 17.74 | 18.23 | 18.55 | 18.82 | 18.65 | 18.84 | 18.04 | 1.158 | 0.069 |
| Zolpidem | 3.13 | 3.58 | 4.05 | 4.44 | 4.94 | 5.37 | 5.85 | 6.65 | 6.91 | 3.776 | 1.206 |
| Antidepressants^1^ | 1.09 | 1.35 | 1.79 | 2.41 | 2.94 | 3.87 | 5.04 | 6.51 | 7.09 | 5.995 | 5.484 |
| Antihistamines^2^ | 1.39 | 1.53 | 1.70 | 1.89 | 2.08 | 2.27 | 2.45 | 2.66 | 2.63 | 1.244 | 0.896 |
| Antipsychotics^3^ | 86.71 | 85.99 | 88.06 | 89.10 | 89.09 | 88.37 | 84.50 | 84.01 | 79.71 | -7.005 | -0.081 |
| Sedatives | 92.33 | 92.45 | 95.59 | 97.84 | 99.05 | 99.88 | 97.84 | 99.84 | 96.34 | 4.010 | 0.043 |

**Rate of cost per 1,000 NHS population**

|  | **2014** | **2015** | **2016** | **2017** | **2018** | **2019** | **2020** | **2021** | **2022** | **Absolute change** | **Relative change** |
| --- | --- | --- | --- | --- | --- | --- | --- | --- | --- | --- | --- |
| Benzodiazepines and z-drugs | 1069.57 | 882.08 | 780.06 | 740.54 | 672.87 | 720.52 | 836.10 | 874.72 | 949.73 | -119.84 | -0.11 |
| Long-acting benzodiazepines | 334.69 | 393.88 | 514.75 | 510.67 | 494.34 | 542.80 | 625.92 | 642.88 | 655.44 | 320.75 | 0.96 |
| Clonazepam | 56.58 | 83.44 | 197.87 | 211.79 | 207.29 | 218.01 | 241.87 | 244.26 | 247.56 | 190.98 | 3.38 |
| Diazepam | 132.64 | 156.08 | 141.40 | 129.05 | 112.11 | 137.40 | 166.39 | 171.36 | 168.71 | 36.06 | 0.27 |
| Chloridiazepoxide | 9.16 | 10.68 | 9.51 | 7.26 | 5.73 | 4.96 | 4.62 | 3.93 | 3.75 | -5.41 | -0.59 |
| Potassium Clorazepate | 0 | 0 | 0 | 0 | 0 | 0 | 0 | 0 | 0 | 0.00 | 0.00 |
| Lorazepam | 58.49 | 56.93 | 67.97 | 99.34 | 71.02 | 65.39 | 86.89 | 118.95 | 136.37 | 77.87 | 1.33 |
| Bromazepam | 0 | 0 | 0 | 0 | 0 | 0 | 0 | 0 | 0 | 0.00 | 0.00 |
| Clobazam | 108.59 | 115.90 | 108.42 | 120.09 | 131.98 | 145.14 | 161.50 | 178.38 | 199.86 | 91.27 | 0.84 |
| Prazepam | 0 | 0 | 0 | 0 | 0 | 0 | 0 | 0 | 0 | 0.00 | 0.00 |
| Alprazolam | 0 | 0 | 0 | 0 | 0 | 0 | 0 | 0 | 0 | 0.00 | 0.00 |
| Flurazepam | 0 | 0 | 0 | 0 | 0 | 0 | 0 | 0 | 0 | 0.00 | 0.00 |
| Nitrazepam | 27.72 | 27.77 | 57.55 | 42.49 | 37.24 | 37.29 | 51.54 | 44.95 | 35.57 | 7.84 | 0.28 |
| Triazolam | 0 | 0 | 0 | 0 | 0 | 0 | 0 | 0 | 0 | 0.00 | 0.00 |
| Lormetazepam | 19.08 | 11.81 | 5.81 | 3.32 | 2.07 | 1.80 | 4.71 | 3.97 | 3.70 | -15.38 | -0.81 |
| Temazepam | 538.13 | 283.17 | 89.82 | 46.72 | 42.82 | 40.32 | 34.49 | 28.93 | 89.25 | -448.88 | -0.83 |
| Zopiclone | 102.12 | 119.83 | 89.05 | 70.48 | 54.84 | 60.83 | 69.62 | 68.24 | 54.79 | -47.32 | -0.46 |
| Zolpidem | 17.05 | 16.46 | 12.65 | 10.02 | 7.78 | 9.39 | 14.46 | 11.74 | 10.19 | -6.86 | -0.40 |
| Antidepressants^1^ | 230.86 | 245.01 | 308.24 | 287.59 | 148.18 | 133.61 | 195.05 | 177.88 | 124.86 | -105.99 | -0.46 |
| Antihistamines^2^ | 121.57 | 143.32 | 141.41 | 140.13 | 148.50 | 155.24 | 193.14 | 181.95 | 442.39 | 320.82 | 2.64 |
| Antipsychotics^3^ | 51.91 | 54.04 | 48.75 | 296.03 | 191.48 | 110.69 | 88.13 | 72.85 | 64.74 | 12.83 | 0.25 |
| Sedatives | 1473.91 | 1324.45 | 1278.46 | 1464.29 | 1161.03 | 1120.05 | 1312.42 | 1307.40 | 1581.73 | 107.82 | 0.07 |

**Rate of DDE per 1,000 NHS population per day**

|  | **2014** | **2015** | **2016** | **2017** | **2018** | **2019** | **2020** | **2021** | **2022** | **Absolute change** | **Relative change** |
| --- | --- | --- | --- | --- | --- | --- | --- | --- | --- | --- | --- |
| Benzodiazepines and z-drugs | 120.08 | 114.70 | 109.73 | 104.55 | 98.93 | 93.76 | 91.52 | 86.07 | 81.77 | -38.31 | -0.32 |
| Long-acting benzodiazepines | 61.72 | 60.28 | 58.55 | 56.68 | 54.45 | 52.04 | 51.64 | 48.89 | 47.50 | -14.22 | -0.23 |
| Clonazepam | 24.50 | 24.84 | 24.88 | 25.08 | 24.78 | 24.23 | 24.88 | 24.03 | 23.72 | -0.78 | -0.03 |
| Diazepam | 28.59 | 27.51 | 26.29 | 24.77 | 23.25 | 21.74 | 20.92 | 19.30 | 18.34 | -10.24 | -0.36 |
| Chloridiazepoxide | 0.74 | 0.55 | 0.46 | 0.38 | 0.31 | 0.27 | 0.22 | 0.19 | 0.15 | -0.59 | -0.79 |
| Potassium Clorazepate | 0.00 | 0.00 | 0.00 | 0.00 | 0.00 | 0.00 | 0.00 | 0.00 | 0.00 | 0.00 | 0.00 |
| Lorazepam | 17.45 | 17.01 | 16.59 | 15.89 | 15.10 | 14.66 | 14.46 | 13.63 | 13.10 | -4.35 | -0.25 |
| Bromazepam | 0.00 | 0.00 | 0.00 | 0.00 | 0.00 | 0.00 | 0.00 | 0.00 | 0.00 | 0.00 | 0.00 |
| Clobazam | 2.73 | 2.86 | 2.94 | 3.01 | 3.09 | 3.17 | 3.29 | 3.36 | 3.45 | 0.72 | 0.26 |
| Prazepam | 0.00 | 0.00 | 0.00 | 0.00 | 0.00 | 0.00 | 0.00 | 0.00 | 0.00 | 0.00 | 0.00 |
| Alprazolam | 0.00 | 0.00 | 0.00 | 0.00 | 0.00 | 0.00 | 0.00 | 0.00 | 0.00 | 0.00 | 0.00 |
| Flurazepam | 0.00 | 0.00 | 0.00 | 0.00 | 0.00 | 0.00 | 0.00 | 0.00 | 0.00 | 0.00 | 0.00 |
| Nitrazepam | 5.16 | 4.52 | 3.97 | 3.44 | 3.01 | 2.64 | 2.33 | 2.01 | 1.84 | -3.32 | -0.64 |
| Triazolam | 0.00 | 0.00 | 0.00 | 0.00 | 0.00 | 0.00 | 0.00 | 0.00 | 0.00 | 0.00 | 0.00 |
| Lormetazepam | 0.29 | 0.26 | 0.23 | 0.20 | 0.18 | 0.16 | 0.12 | 0.11 | 0.10 | -0.19 | -0.65 |
| Temazepam | 13.22 | 11.02 | 9.59 | 8.31 | 7.24 | 6.37 | 5.72 | 4.99 | 3.76 | -9.46 | -0.72 |
| Zopiclone | 24.06 | 22.93 | 21.71 | 20.60 | 19.28 | 18.02 | 17.20 | 16.22 | 15.18 | -8.88 | -0.37 |
| Zolpidem | 3.35 | 3.20 | 3.06 | 2.87 | 2.68 | 2.51 | 2.37 | 2.24 | 2.13 | -1.21 | -0.36 |

**Rate of DDD per 1,000 NHS population per day**

|  | **2014** | **2015** | **2016** | **2017** | **2018** | **2019** | **2020** | **2021** | **2022** | **Absolute change** | **Relative change** |
| --- | --- | --- | --- | --- | --- | --- | --- | --- | --- | --- | --- |
| Benzodiazepines and z-drugs | 11.69 | 10.97 | 10.32 | 9.65 | 8.98 | 8.37 | 7.98 | 7.43 | 6.94 | -4.75 | -0.41 |
| Long-acting benzodiazepines | 4.38 | 4.14 | 3.91 | 3.66 | 3.42 | 3.19 | 3.06 | 2.83 | 2.71 | -1.67 | -0.38 |
| Clonazepam | 0.15 | 0.16 | 0.16 | 0.16 | 0.15 | 0.15 | 0.16 | 0.15 | 0.15 | 0.00 | -0.03 |
| Diazepam | 2.86 | 2.75 | 2.63 | 2.48 | 2.33 | 2.17 | 2.09 | 1.93 | 1.83 | -1.02 | -0.36 |
| Chloridiazepoxide | 0.06 | 0.05 | 0.04 | 0.03 | 0.03 | 0.02 | 0.02 | 0.02 | 0.01 | -0.05 | -0.79 |
| Potassium Clorazepate | 0 | 0 | 0 | 0 | 0 | 0 | 0 | 0 | 0 | 0.00 | 0.00 |
| Lorazepam | 0.70 | 0.68 | 0.66 | 0.64 | 0.60 | 0.59 | 0.58 | 0.55 | 0.52 | -0.17 | -0.25 |
| Bromazepam | 0 | 0 | 0 | 0 | 0 | 0 | 0 | 0 | 0 | 0.00 | 0.00 |
| Clobazam | 0.27 | 0.29 | 0.29 | 0.30 | 0.31 | 0.32 | 0.33 | 0.34 | 0.34 | 0.07 | 0.26 |
| Prazepam | 0 | 0 | 0 | 0 | 0 | 0 | 0 | 0 | 0 | 0.00 | 0.00 |
| Alprazolam | 0 | 0 | 0 | 0 | 0 | 0 | 0 | 0 | 0 | 0.00 | 0.00 |
| Flurazepam | 0 | 0 | 0 | 0 | 0 | 0 | 0 | 0 | 0 | 0.00 | 0.00 |
| Nitrazepam | 1.03 | 0.90 | 0.79 | 0.69 | 0.60 | 0.53 | 0.47 | 0.40 | 0.37 | -0.66 | -0.64 |
| Triazolam | 0 | 0 | 0 | 0 | 0 | 0 | 0 | 0 | 0 | 0.00 | 0.00 |
| Lormetazepam | 0.04 | 0.04 | 0.03 | 0.03 | 0.03 | 0.02 | 0.02 | 0.02 | 0.02 | -0.03 | -0.65 |
| Temazepam | 1.32 | 1.10 | 0.96 | 0.83 | 0.72 | 0.64 | 0.57 | 0.50 | 0.38 | -0.95 | -0.72 |
| Zopiclone | 4.58 | 4.37 | 4.14 | 3.92 | 3.67 | 3.43 | 3.28 | 3.09 | 2.89 | -1.69 | -0.37 |
| Zolpidem | 0.67 | 0.64 | 0.61 | 0.57 | 0.54 | 0.50 | 0.47 | 0.45 | 0.43 | -0.24 | -0.36 |
| Antidepressants^1^ | 1.63 | 1.79 | 1.95 | 2.09 | 2.27 | 2.45 | 2.67 | 2.86 | 3.00 | 1.37 | 0.84 |
| Antihistamines^2^ | 1.85 | 2.07 | 2.27 | 2.45 | 2.56 | 2.70 | 3.18 | 3.56 | 3.82 | 1.98 | 1.07 |
| Antipsychotics^3^ | 0.32 | 0.34 | 0.36 | 0.37 | 0.38 | 0.39 | 0.41 | 0.41 | 0.41 | 0.09 | 0.27 |
| Sedatives | 15.49 | 15.17 | 14.90 | 14.56 | 14.19 | 13.91 | 14.23 | 14.25 | 14.17 | -1.32 | -0.09 |

^1^doxepin, trazodone, mirtazapine

^2^promethazine, cyclizine, ketotifen

^3^olanzapine, quetiapine, risperidone

**Supplementary Table 8. Rate of claims, cost, DDEs, and DDDs per 1,000 NHS (England) population (top 33% of CCGs for deprivation)**

**Rate of claims**

|  | **2014** | **2015** | **2016** | **2017** | **2018** | **2019** | **2020** | **2021** | **2022** | **Absolute change** | **Relative change** |
| --- | --- | --- | --- | --- | --- | --- | --- | --- | --- | --- | --- |
| Benzodiazepines and z-drugs | 238.43 | 271.83 | 262.44 | 248.63 | 237.15 | 231.17 | 220.96 | 212.65 | 216.30 | -22.13 | -0.09 |
| Long-acting benzodiazepines | 106.66 | 123.29 | 120.76 | 115.23 | 110.84 | 108.82 | 103.60 | 98.66 | 103.80 | -2.86 | -0.03 |
| Clonazepam | 12.45 | 14.53 | 15.07 | 15.09 | 15.12 | 15.71 | 14.69 | 14.84 | 15.25 | 2.80 | 0.23 |
| Diazepam | 75.44 | 88.63 | 87.03 | 83.21 | 79.90 | 77.94 | 74.27 | 69.94 | 73.77 | -1.68 | -0.02 |
| Chloridiazepoxide | 1.83 | 1.56 | 1.30 | 1.02 | 0.87 | 0.72 | 0.62 | 0.52 | 0.50 | -1.34 | -0.73 |
| Potassium Clorazepate | 0 | 0 | 0 | 0 | 0 | 0 | 0 | 0 | 0 | 0.00 | 0.00 |
| Lorazepam | 14.98 | 17.72 | 17.85 | 17.20 | 16.75 | 17.10 | 17.23 | 16.83 | 17.63 | 2.64 | 0.18 |
| Bromazepam | 0 | 0 | 0 | 0 | 0 | 0 | 0 | 0 | 0 | 0.00 | 0.00 |
| Clobazam | 4.20 | 5.23 | 5.44 | 5.58 | 5.82 | 6.15 | 6.48 | 6.61 | 7.31 | 3.10 | 0.74 |
| Prazepam | 0 | 0 | 0 | 0 | 0 | 0 | 0 | 0 | 0 | 0.00 | 0.00 |
| Alprazolam | 0 | 0 | 0 | 0 | 0 | 0 | 0 | 0 | 0 | 0.00 | 0.00 |
| Flurazepam | 0 | 0 | 0 | 0 | 0 | 0 | 0 | 0 | 0 | 0.00 | 0.00 |
| Nitrazepam | 12.74 | 13.35 | 11.92 | 10.32 | 9.14 | 8.30 | 7.53 | 6.75 | 6.98 | -5.76 | -0.45 |
| Flunitrazepam | 0 | 0 | 0 | 0 | 0 | 0 | 0 | 0 | 0 | 0.00 | 0.00 |
| Triazolam | 0 | 0 | 0 | 0 | 0 | 0 | 0 | 0 | 0 | 0.00 | 0.00 |
| Lormetazepam | 0.49 | 0.52 | 0.46 | 0.39 | 0.33 | 0.31 | 0.27 | 0.24 | 0.24 | -0.26 | -0.52 |
| Temazepam | 23.51 | 24.91 | 21.92 | 18.83 | 16.64 | 15.09 | 14.21 | 12.67 | 10.69 | -12.82 | -0.55 |
| Zopiclone | 81.59 | 93.00 | 89.63 | 86.10 | 82.33 | 80.06 | 76.53 | 75.33 | 75.20 | -6.39 | -0.08 |
| Zolpidem | 11.19 | 12.38 | 11.83 | 10.89 | 10.26 | 9.79 | 9.11 | 8.92 | 8.75 | -2.44 | -0.22 |
| Antidepressants^1^ | 45.98 | 60.97 | 66.65 | 70.71 | 76.73 | 84.91 | 95.46 | 103.20 | 117.61 | 71.62 | 1.56 |
| Antihistamines^2^ | 25.33 | 33.13 | 36.45 | 38.95 | 41.16 | 43.71 | 50.28 | 57.93 | 62.56 | 37.23 | 1.47 |
| Antipsychotics^3^ | 22.93 | 29.90 | 32.21 | 33.71 | 34.51 | 36.35 | 37.38 | 37.66 | 39.30 | 16.36 | 0.71 |
| Sedatives | 332.67 | 395.83 | 397.74 | 392.00 | 389.55 | 396.14 | 404.08 | 411.45 | 435.76 | 103.09 | 0.31 |

**Rate of DDE**

|  | **2014** | **2015** | **2016** | **2017** | **2018** | **2019** | **2020** | **2021** | **2022** | **Absolute change** | **Relative change** |
| --- | --- | --- | --- | --- | --- | --- | --- | --- | --- | --- | --- |
| Benzodiazepines and z-drugs | 36293.20 | 40515.82 | 38540.41 | 36127.29 | 33992.14 | 32964.36 | 30985.40 | 29396.39 | 29727.98 | -6565.22 | -0.18 |
| Long-acting benzodiazepines | 19074.78 | 21631.88 | 20850.60 | 19808.56 | 18936.49 | 18547.24 | 17435.60 | 16615.71 | 17273.18 | -1801.59 | -0.09 |
| Clonazepam | 7396.11 | 8557.20 | 8499.28 | 8340.79 | 8209.52 | 8338.50 | 7755.99 | 7660.18 | 7917.21 | 521.10 | 0.07 |
| Diazepam | 8677.67 | 9845.67 | 9342.62 | 8710.63 | 8128.79 | 7700.67 | 7257.25 | 6627.67 | 6873.23 | -1804.45 | -0.21 |
| Chloridiazepoxide | 235.64 | 201.68 | 165.58 | 131.90 | 110.61 | 94.44 | 81.64 | 69.09 | 65.74 | -169.89 | -0.72 |
| Potassium Clorazepate | 0 | 0 | 0 | 0 | 0 | 0 | 0 | 0 | 0 | 0.00 | 0.00 |
| Lorazepam | 5198.46 | 5930.00 | 5855.52 | 5567.16 | 5206.55 | 5225.36 | 5062.77 | 4829.84 | 4954.89 | -243.57 | -0.05 |
| Bromazepam | 0 | 0 | 0 | 0 | 0 | 0 | 0 | 0 | 0 | 0.00 | 0.00 |
| Clobazam | 951.55 | 1150.55 | 1191.38 | 1214.78 | 1253.23 | 1313.92 | 1366.89 | 1382.75 | 1530.56 | 579.01 | 0.61 |
| Prazepam | 0 | 0 | 0 | 0 | 0 | 0 | 0 | 0 | 0 | 0.00 | 0.00 |
| Alprazolam | 0 | 0 | 0 | 0 | 0 | 0 | 0 | 0 | 0 | 0.00 | 0.00 |
| Flurazepam | 0 | 0 | 0 | 0 | 0 | 0 | 0 | 0 | 0 | 0.00 | 0.00 |
| Nitrazepam | 1813.80 | 1876.77 | 1651.75 | 1410.46 | 1234.34 | 1099.71 | 973.83 | 876.02 | 886.45 | -927.36 | -0.51 |
| Flunitrazepam | 0 | 0 | 0 | 0 | 0 | 0 | 0 | 0 | 0 | 0.00 | 0.00 |
| Triazolam | 0 | 0 | 0 | 0 | 0 | 0 | 0 | 0 | 0 | 0.00 | 0.00 |
| Lormetazepam | 90.50 | 96.74 | 83.55 | 72.70 | 63.11 | 57.91 | 47.69 | 43.35 | 44.23 | -46.27 | -0.51 |
| Temazepam | 3947.76 | 4077.33 | 3543.73 | 3018.36 | 2645.04 | 2382.63 | 2207.64 | 1948.28 | 1610.89 | -2336.87 | -0.59 |
| Zopiclone | 6940.88 | 7657.42 | 7158.56 | 6702.00 | 6253.73 | 5922.12 | 5471.04 | 5239.19 | 5155.04 | -1785.84 | -0.26 |
| Zolpidem | 1040.83 | 1122.46 | 1048.45 | 958.50 | 887.22 | 829.09 | 760.66 | 720.02 | 689.74 | -351.09 | -0.34 |

**Rate of DDD**

|  | **2014** | **2015** | **2016** | **2017** | **2018** | **2019** | **2020** | **2021** | **2022** | **Absolute change** | **Relative change** |
| --- | --- | --- | --- | --- | --- | --- | --- | --- | --- | --- | --- |
| Benzodiazepines and z-drugs | 3538.01 | 3887.69 | 3624.96 | 3341.40 | 3096.41 | 2931.15 | 2737.08 | 2566.32 | 2558.38 | -979.63 | -0.28 |
| Long-acting benzodiazepines | 1391.55 | 1545.27 | 1450.67 | 1337.75 | 1245.60 | 1181.39 | 1112.46 | 1029.88 | 1072.63 | -318.92 | -0.23 |
| Clonazepam | 46.23 | 53.48 | 53.12 | 52.13 | 51.31 | 52.12 | 48.47 | 47.88 | 49.48 | 3.26 | 0.07 |
| Diazepam | 867.77 | 984.57 | 934.26 | 871.06 | 812.88 | 770.07 | 725.72 | 662.77 | 687.32 | -180.44 | -0.21 |
| Chloridiazepoxide | 19.64 | 16.81 | 13.80 | 10.99 | 9.22 | 7.87 | 6.80 | 5.76 | 5.48 | -14.16 | -0.72 |
| Potassium Clorazepate | 0 | 0 | 0 | 0 | 0 | 0 | 0 | 0 | 0 | 0.00 | 0.00 |
| Lorazepam | 207.94 | 237.20 | 234.22 | 222.69 | 208.26 | 209.01 | 202.51 | 193.19 | 198.20 | -9.74 | -0.05 |
| Bromazepam | 0 | 0 | 0 | 0 | 0 | 0 | 0 | 0 | 0 | 0.00 | 0.00 |
| Clobazam | 95.16 | 115.06 | 119.14 | 121.48 | 125.32 | 131.39 | 136.69 | 138.28 | 153.06 | 57.90 | 0.61 |
| Prazepam | 0 | 0 | 0 | 0 | 0 | 0 | 0 | 0 | 0 | 0.00 | 0.00 |
| Alprazolam | 0 | 0 | 0 | 0 | 0 | 0 | 0 | 0 | 0 | 0.00 | 0.00 |
| Flurazepam | 0 | 0 | 0 | 0 | 0 | 0 | 0 | 0 | 0 | 0.00 | 0.00 |
| Nitrazepam | 362.76 | 375.35 | 330.35 | 282.09 | 246.87 | 219.94 | 194.77 | 175.20 | 177.29 | -185.47 | -0.51 |
| Flunitrazepam | 0 | 0 | 0 | 0 | 0 | 0 | 0 | 0 | 0 | 0.00 | 0.00 |
| Triazolam | 0 | 0 | 0 | 0 | 0 | 0 | 0 | 0 | 0 | 0.00 | 0.00 |
| Lormetazepam | 13.51 | 14.44 | 12.47 | 10.85 | 9.42 | 8.64 | 7.12 | 6.47 | 6.60 | -6.91 | -0.51 |
| Temazepam | 394.78 | 407.73 | 354.37 | 301.84 | 264.50 | 238.26 | 220.76 | 194.83 | 161.09 | -233.69 | -0.59 |
| Zopiclone | 1322.07 | 1458.56 | 1363.53 | 1276.57 | 1191.19 | 1128.02 | 1042.10 | 997.94 | 981.91 | -340.16 | -0.26 |
| Zolpidem | 208.17 | 224.49 | 209.69 | 191.70 | 177.44 | 165.82 | 152.13 | 144.00 | 137.95 | -70.22 | -0.34 |
| Antidepressants^1^ | 540.00 | 706.25 | 769.27 | 814.54 | 882.10 | 979.78 | 1091.93 | 1170.41 | 1348.10 | 808.09 | 1.50 |
| Antihistamines^2^ | 681.74 | 877.28 | 958.02 | 1019.36 | 1066.17 | 1127.05 | 1278.18 | 1479.18 | 1590.92 | 909.18 | 1.33 |
| Antipsychotics^3^ | 90.59 | 114.91 | 121.82 | 125.89 | 127.31 | 132.38 | 135.19 | 133.36 | 139.46 | 48.87 | 0.54 |
| Sedatives | 4850.34 | 5586.12 | 5474.06 | 5301.19 | 5171.98 | 5170.36 | 5242.39 | 5349.27 | 5636.84 | 786.51 | 0.16 |

**Rate of cost**

|  | **2014** | **2015** | **2016** | **2017** | **2018** | **2019** | **2020** | **2021** | **2022** | **Absolute change** | **Relative change** |
| --- | --- | --- | --- | --- | --- | --- | --- | --- | --- | --- | --- |
| Benzodiazepines and z-drugs | 868.35 | 874.20 | 794.86 | 738.69 | 682.25 | 746.57 | 869.05 | 932.78 | 1094.07 | 225.72 | 0.26 |
| Long-acting benzodiazepines | 291.87 | 408.49 | 542.36 | 522.13 | 511.03 | 573.56 | 666.68 | 702.24 | 770.02 | 478.15 | 1.64 |
| Clonazepam | 50.17 | 85.50 | 188.98 | 199.37 | 193.19 | 207.50 | 214.27 | 220.03 | 236.55 | 186.38 | 3.72 |
| Diazepam | 111.28 | 160.70 | 150.69 | 136.33 | 126.00 | 156.11 | 197.38 | 206.89 | 229.56 | 118.28 | 1.06 |
| Chloridiazepoxide | 7.93 | 10.61 | 9.12 | 7.02 | 5.51 | 4.77 | 4.60 | 3.97 | 4.39 | -3.54 | -0.45 |
| Potassium Clorazepate | 0 | 0 | 0 | 0 | 0 | 0 | 0 | 0 | 0 | 0.00 | 0.00 |
| Lorazepam | 48.53 | 54.85 | 66.04 | 94.54 | 67.76 | 65.19 | 86.52 | 123.04 | 157.56 | 109.03 | 2.25 |
| Bromazepam | 0 | 0 | 0 | 0 | 0 | 0 | 0 | 0 | 0 | 0.00 | 0.00 |
| Clobazam | 95.35 | 118.49 | 112.10 | 122.74 | 137.96 | 158.73 | 184.82 | 208.21 | 246.24 | 150.89 | 1.58 |
| Prazepam | 0 | 0 | 0 | 0 | 0 | 0 | 0 | 0 | 0 | 0.00 | 0.00 |
| Alprazolam | 0 | 0 | 0 | 0 | 0 | 0 | 0 | 0 | 0 | 0.00 | 0.00 |
| Flurazepam | 0 | 0 | 0 | 0 | 0 | 0 | 0 | 0 | 0 | 0.00 | 0.00 |
| Nitrazepam | 27.15 | 33.18 | 81.49 | 56.67 | 48.38 | 46.46 | 65.61 | 63.14 | 53.29 | 26.14 | 0.96 |
| Flunitrazepam | 0 | 0 | 0 | 0 | 0 | 0 | 0 | 0 | 0 | 0.00 | 0.00 |
| Triazolam | 0 | 0 | 0 | 0 | 0 | 0 | 0 | 0 | 0 | 0.00 | 0.00 |
| Lormetazepam | 15.97 | 12.04 | 5.83 | 3.20 | 1.95 | 1.71 | 4.71 | 4.27 | 4.41 | -11.56 | -0.72 |
| Temazepam | 418.58 | 276.05 | 90.18 | 47.59 | 45.64 | 43.16 | 38.24 | 32.97 | 102.54 | -316.04 | -0.76 |
| Zopiclone | 79.15 | 107.28 | 78.79 | 62.20 | 48.89 | 54.52 | 60.61 | 60.17 | 50.79 | -28.36 | -0.36 |
| Zolpidem | 14.24 | 15.51 | 11.66 | 9.02 | 6.97 | 8.43 | 12.29 | 10.09 | 8.75 | -5.50 | -0.39 |
| Antidepressants^1^ | 223.34 | 286.61 | 364.85 | 321.63 | 162.99 | 150.56 | 220.51 | 202.00 | 161.69 | -61.65 | -0.28 |
| Antihistamines^2^ | 117.64 | 157.70 | 152.75 | 145.22 | 154.91 | 163.95 | 200.75 | 200.50 | 484.54 | 366.90 | 3.12 |
| Antipsychotics^3^ | 39.63 | 49.51 | 45.63 | 280.72 | 179.78 | 104.02 | 81.53 | 67.28 | 63.46 | 23.83 | 0.60 |
| Sedatives | 1248.96 | 1368.03 | 1358.10 | 1486.26 | 1179.92 | 1165.10 | 1371.83 | 1402.55 | 1803.76 | 554.80 | 0.44 |

^1^doxepin, trazodone, mirtazapine

^2^promethazine, cyclizine, ketotifen

^3^olanzapine, quetiapine, risperidone

**Supplementary Table 9. Rate of claims, cost, DDEs, and DDDs per 1,000 GMS population by age group and sex**

**Rate of claims per 1,000 GMS population by age group**

|  | **<5** | **5-11** | **12-15** | **16-24** | **25-34** | **35-44** | **45-54** | **55-64** | **65-69** | **70-74** | **75+** |
| --- | --- | --- | --- | --- | --- | --- | --- | --- | --- | --- | --- |
| **2014** |  |  |  |  |  |  |  |  |  |  |  |
| Benzodiazepines and z-drugs | 10.02 | 12.21 | 22.75 | 214.66 | 907.22 | 1390.87 | 1903.49 | 2603.56 | 2794.78 | 2815.63 | 3735.69 |
| Long-acting benzodiazepines | 9.03 | 9.96 | 16.82 | 91.21 | 401.50 | 587.00 | 681.63 | 794.82 | 737.96 | 660.44 | 685.24 |
| Clonazepam | 1.80 | 1.73 | 2.24 | 13.79 | 41.37 | 56.09 | 75.40 | 79.69 | 68.67 | 60.45 | 64.51 |
| Diazepam | 2.61 | 3.32 | 5.88 | 60.14 | 281.42 | 389.11 | 397.70 | 443.41 | 410.35 | 365.59 | 373.21 |
| Chloridiazepoxide | 0.01 | 0.03 | 0.66 | 2.50 | 15.55 | 24.14 | 39.56 | 40.31 | 26.00 | 23.71 | 22.99 |
| Potassium Clorazepate | 0.00 | 0.00 | 0.00 | 0.07 | 0.03 | 0.09 | 0.38 | 0.27 | 0.14 | 0.15 | 0.17 |
| Lorazepam | 0.02 | 0.21 | 0.66 | 4.39 | 11.84 | 15.32 | 24.95 | 39.62 | 52.27 | 50.74 | 81.02 |
| Bromazepam | 0.00 | 0.01 | 0.27 | 3.32 | 12.12 | 21.62 | 50.45 | 107.15 | 148.58 | 145.09 | 158.78 |
| Clobazam | 4.27 | 4.62 | 5.17 | 7.52 | 12.12 | 14.53 | 18.02 | 19.07 | 17.41 | 15.28 | 12.41 |
| Prazepam | 0.01 | 0.01 | 0.01 | 0.63 | 1.85 | 3.34 | 9.91 | 24.58 | 29.24 | 29.80 | 27.13 |
| Alprazolam | 0.28 | 0.58 | 2.98 | 45.86 | 168.81 | 230.97 | 310.28 | 413.78 | 432.84 | 423.14 | 538.54 |
| Flurazepam | 0.02 | 0.04 | 2.48 | 5.85 | 46.98 | 95.99 | 132.33 | 172.17 | 163.01 | 134.80 | 126.24 |
| Nitrazepam | 0.32 | 0.21 | 0.37 | 0.71 | 2.18 | 3.72 | 8.33 | 15.33 | 23.13 | 30.65 | 58.56 |
| Triazolam | 0.01 | 0.06 | 0.01 | 3.67 | 21.62 | 26.45 | 45.89 | 72.92 | 93.51 | 100.81 | 156.16 |
| Lormetazepam | 0.01 | 0.05 | 0.18 | 0.80 | 2.46 | 6.24 | 17.54 | 35.70 | 56.56 | 80.34 | 134.44 |
| Temazepam | 0.13 | 0.01 | 0.08 | 2.12 | 10.05 | 21.31 | 47.90 | 90.42 | 134.33 | 141.49 | 289.91 |
| Zopiclone | 0.19 | 0.71 | 0.81 | 36.07 | 175.08 | 295.36 | 412.11 | 577.90 | 600.73 | 638.28 | 917.40 |
| Zolpidem | 0.25 | 0.52 | 0.71 | 25.26 | 101.20 | 182.53 | 307.79 | 464.72 | 529.93 | 569.17 | 763.86 |
| Antidepressants^1^ | 0.07 | 0.11 | 0.17 | 17.00 | 51.69 | 75.11 | 107.37 | 124.89 | 128.48 | 145.54 | 292.47 |
| Antihistamines^2^ | 0.00 | 0.18 | 0.65 | 5.38 | 11.74 | 15.14 | 20.37 | 19.77 | 12.50 | 10.32 | 12.42 |
| Antipsychotics^3^ | 0.05 | 0.48 | 3.43 | 32.64 | 76.61 | 95.49 | 123.23 | 131.54 | 136.34 | 165.33 | 414.82 |
| Sedatives | 10.14 | 12.98 | 26.99 | 269.68 | 1047.26 | 1576.61 | 2154.46 | 2879.75 | 3072.10 | 3136.82 | 4455.41 |
| **2015** |  |  |  |  |  |  |  |  |  |  |  |
| Benzodiazepines and z-drugs | 6.89 | 10.59 | 25.20 | 219.50 | 899.11 | 1342.79 | 1875.60 | 2523.11 | 2742.96 | 2708.33 | 3543.13 |
| Long-acting benzodiazepines | 6.13 | 8.55 | 17.74 | 93.43 | 393.55 | 568.54 | 671.22 | 761.71 | 724.18 | 633.44 | 639.76 |
| Clonazepam | 0.93 | 1.35 | 1.80 | 15.19 | 45.81 | 56.33 | 74.90 | 80.29 | 70.07 | 58.65 | 62.99 |
| Diazepam | 2.20 | 3.00 | 7.60 | 61.75 | 279.96 | 381.73 | 402.31 | 429.96 | 412.61 | 351.74 | 356.02 |
| Chloridiazepoxide | 0.01 | 0.03 | 0.99 | 1.93 | 14.37 | 25.72 | 35.16 | 37.46 | 26.06 | 22.31 | 20.43 |
| Potassium Clorazepate | 0.00 | 0.00 | 0.00 | 0.05 | 0.11 | 0.08 | 0.40 | 0.41 | 0.12 | 0.12 | 0.11 |
| Lorazepam | 0.01 | 0.11 | 1.03 | 5.34 | 13.36 | 15.39 | 23.91 | 37.29 | 49.28 | 48.94 | 73.96 |
| Bromazepam | 0.01 | 0.01 | 0.23 | 4.21 | 11.62 | 20.59 | 46.53 | 97.84 | 143.04 | 137.47 | 152.06 |
| Clobazam | 2.58 | 3.82 | 3.39 | 6.78 | 9.27 | 11.69 | 17.02 | 16.73 | 16.90 | 14.07 | 12.18 |
| Prazepam | 0.00 | 0.00 | 0.01 | 0.46 | 1.77 | 3.59 | 8.57 | 22.38 | 27.81 | 28.46 | 25.02 |
| Alprazolam | 0.22 | 0.74 | 3.17 | 45.29 | 174.58 | 224.60 | 305.62 | 403.20 | 424.81 | 417.98 | 521.39 |
| Flurazepam | 0.01 | 0.05 | 3.56 | 6.38 | 40.39 | 86.53 | 125.87 | 161.23 | 149.59 | 129.36 | 113.25 |
| Nitrazepam | 0.40 | 0.30 | 0.39 | 0.89 | 1.87 | 2.88 | 6.99 | 13.25 | 21.03 | 28.72 | 49.76 |
| Triazolam | 0.01 | 0.01 | 0.14 | 3.07 | 22.63 | 27.43 | 43.25 | 72.36 | 97.85 | 93.73 | 149.80 |
| Lormetazepam | 0.01 | 0.01 | 0.18 | 0.69 | 2.76 | 5.80 | 15.71 | 30.37 | 49.02 | 72.48 | 122.41 |
| Temazepam | 0.00 | 0.25 | 0.08 | 2.60 | 9.06 | 19.17 | 41.87 | 80.54 | 118.66 | 125.75 | 254.12 |
| Zopiclone | 0.13 | 0.47 | 1.30 | 37.43 | 170.27 | 283.21 | 414.23 | 572.19 | 605.25 | 623.54 | 889.81 |
| Zolpidem | 0.27 | 0.22 | 1.09 | 25.40 | 98.28 | 172.42 | 307.42 | 460.21 | 522.67 | 548.56 | 729.08 |
| Antidepressants^1^ | 0.10 | 0.06 | 0.70 | 21.29 | 59.69 | 85.24 | 116.83 | 137.93 | 147.69 | 162.99 | 321.15 |
| Antihistamines^2^ | 0.00 | 0.18 | 0.88 | 7.20 | 15.90 | 19.98 | 25.11 | 23.56 | 15.38 | 11.02 | 14.07 |
| Antipsychotics^3^ | 0.01 | 0.70 | 3.83 | 43.12 | 90.08 | 108.40 | 134.75 | 144.92 | 152.57 | 165.70 | 439.08 |
| Sedatives | 7.00 | 11.53 | 30.61 | 291.11 | 1064.78 | 1556.40 | 2152.29 | 2829.51 | 3058.61 | 3048.04 | 4317.43 |
| **2016** |  |  |  |  |  |  |  |  |  |  |  |
| Benzodiazepines and z-drugs | 7.52 | 10.44 | 16.49 | 225.38 | 975.80 | 1381.89 | 1882.97 | 2474.58 | 2731.29 | 2698.39 | 3501.71 |
| Long-acting benzodiazepines | 6.10 | 8.37 | 10.38 | 96.49 | 427.39 | 581.95 | 679.16 | 739.09 | 714.28 | 627.48 | 620.47 |
| Clonazepam | 0.79 | 1.21 | 1.44 | 14.94 | 52.81 | 61.60 | 75.96 | 83.12 | 71.54 | 67.14 | 67.00 |
| Diazepam | 2.93 | 2.69 | 5.11 | 62.83 | 305.41 | 396.21 | 413.11 | 422.40 | 412.23 | 348.48 | 346.24 |
| Chloridiazepoxide | 0.02 | 0.05 | 0.08 | 2.84 | 16.68 | 27.77 | 34.73 | 36.90 | 25.61 | 21.51 | 18.58 |
| Potassium Clorazepate | 0.00 | 0.00 | 0.00 | 0.00 | 0.13 | 0.02 | 0.28 | 0.23 | 0.39 | 0.03 | 0.17 |
| Lorazepam | 0.08 | 0.18 | 1.44 | 6.73 | 15.48 | 16.60 | 24.30 | 37.20 | 44.67 | 46.08 | 72.54 |
| Bromazepam | 0.02 | 0.04 | 0.16 | 4.19 | 13.74 | 22.01 | 43.87 | 90.57 | 137.16 | 139.16 | 147.63 |
| Clobazam | 1.95 | 3.82 | 3.15 | 6.81 | 9.50 | 10.61 | 15.94 | 15.37 | 15.98 | 12.65 | 12.02 |
| Prazepam | 0.00 | 0.00 | 0.01 | 0.44 | 1.60 | 3.32 | 7.89 | 20.09 | 25.05 | 26.81 | 24.57 |
| Alprazolam | 0.21 | 0.62 | 1.99 | 46.52 | 189.67 | 234.84 | 310.81 | 399.67 | 426.12 | 417.58 | 524.05 |
| Flurazepam | 0.15 | 0.11 | 0.04 | 7.85 | 39.27 | 79.79 | 124.26 | 149.87 | 142.98 | 126.05 | 104.88 |
| Nitrazepam | 0.26 | 0.49 | 0.55 | 0.78 | 2.00 | 2.63 | 6.99 | 11.10 | 20.50 | 24.82 | 47.01 |
| Triazolam | 0.08 | 0.04 | 0.17 | 3.33 | 22.68 | 28.62 | 41.91 | 69.66 | 93.09 | 91.50 | 146.91 |
| Lormetazepam | 0.00 | 0.01 | 0.14 | 0.62 | 2.89 | 5.05 | 13.96 | 29.23 | 47.60 | 67.61 | 117.45 |
| Temazepam | 0.00 | 0.09 | 0.03 | 2.07 | 8.77 | 19.38 | 38.66 | 74.12 | 110.37 | 120.92 | 233.55 |
| Zopiclone | 0.55 | 0.58 | 1.11 | 39.63 | 187.95 | 294.36 | 421.65 | 570.46 | 614.39 | 625.57 | 892.04 |
| Zolpidem | 0.44 | 0.41 | 0.92 | 24.60 | 105.00 | 176.26 | 304.75 | 459.71 | 539.18 | 557.47 | 740.35 |
| Antidepressants^1^ | 0.15 | 0.06 | 0.24 | 23.24 | 73.80 | 96.81 | 129.58 | 153.20 | 152.55 | 178.01 | 350.24 |
| Antihistamines^2^ | 0.01 | 0.17 | 1.24 | 10.55 | 23.20 | 27.66 | 31.17 | 31.69 | 18.66 | 15.91 | 17.63 |
| Antipsychotics^3^ | 0.04 | 0.96 | 3.99 | 53.82 | 115.61 | 122.85 | 151.81 | 158.67 | 158.91 | 175.90 | 456.00 |
| Sedatives | 7.73 | 11.63 | 21.96 | 312.98 | 1188.40 | 1629.20 | 2195.53 | 2818.14 | 3061.41 | 3068.22 | 4325.59 |
| **2017** |  |  |  |  |  |  |  |  |  |  |  |
| Benzodiazepines and z-drugs | 6.96 | 9.48 | 15.67 | 211.79 | 1022.20 | 1450.58 | 1897.01 | 2425.48 | 2625.03 | 2593.59 | 3320.02 |
| Long-acting benzodiazepines | 5.47 | 7.44 | 10.68 | 93.71 | 449.80 | 615.22 | 692.24 | 729.55 | 689.92 | 596.86 | 585.01 |
| Clonazepam | 0.68 | 1.05 | 1.74 | 15.04 | 58.14 | 66.50 | 80.96 | 91.06 | 71.42 | 72.78 | 68.16 |
| Diazepam | 2.19 | 2.49 | 5.21 | 60.76 | 326.34 | 424.29 | 428.27 | 418.39 | 398.73 | 330.00 | 326.89 |
| Chloridiazepoxide | 0.00 | 0.04 | 0.05 | 2.66 | 16.50 | 28.62 | 35.51 | 35.13 | 24.65 | 18.76 | 18.03 |
| Potassium Clorazepate | 0.00 | 0.00 | 0.00 | 0.02 | 0.01 | 0.15 | 0.11 | 0.50 | 0.26 | 0.17 | 0.21 |
| Lorazepam | 0.05 | 0.22 | 1.53 | 6.42 | 17.37 | 18.30 | 24.12 | 38.49 | 42.85 | 45.28 | 65.73 |
| Bromazepam | 0.00 | 0.01 | 0.23 | 3.66 | 13.38 | 23.13 | 41.58 | 83.20 | 127.44 | 135.93 | 140.53 |
| Clobazam | 1.91 | 3.14 | 2.87 | 6.51 | 10.40 | 10.16 | 14.73 | 15.46 | 14.86 | 11.85 | 11.16 |
| Prazepam | 0.00 | 0.01 | 0.02 | 0.31 | 1.91 | 2.95 | 6.85 | 17.31 | 23.44 | 24.65 | 23.72 |
| Alprazolam | 0.31 | 0.47 | 1.35 | 44.89 | 198.79 | 249.96 | 310.92 | 392.90 | 406.51 | 402.69 | 499.20 |
| Flurazepam | 0.25 | 0.01 | 0.12 | 7.48 | 34.40 | 79.89 | 119.54 | 141.53 | 139.33 | 117.36 | 95.69 |
| Nitrazepam | 0.44 | 0.71 | 0.66 | 0.93 | 2.10 | 2.65 | 6.27 | 10.17 | 17.23 | 21.29 | 41.16 |
| Triazolam | 0.00 | 0.01 | 0.07 | 2.59 | 21.35 | 30.67 | 41.20 | 68.01 | 88.01 | 86.54 | 138.37 |
| Lormetazepam | 0.01 | 0.10 | 0.01 | 0.92 | 2.93 | 3.99 | 13.42 | 28.09 | 42.38 | 58.10 | 104.90 |
| Temazepam | 0.07 | 0.01 | 0.06 | 1.71 | 9.72 | 17.44 | 35.20 | 68.11 | 98.96 | 109.93 | 206.47 |
| Zopiclone | 0.80 | 0.81 | 1.09 | 36.00 | 204.54 | 309.65 | 429.01 | 565.76 | 601.47 | 608.09 | 857.36 |
| Zolpidem | 0.25 | 0.40 | 0.63 | 21.76 | 104.14 | 181.87 | 309.06 | 450.78 | 526.83 | 549.61 | 721.89 |
| Antidepressants^1^ | 0.20 | 0.03 | 0.32 | 26.54 | 87.63 | 113.00 | 143.12 | 166.61 | 156.70 | 186.19 | 365.66 |
| Antihistamines^2^ | 0.00 | 0.11 | 1.37 | 14.98 | 33.17 | 37.22 | 42.17 | 42.81 | 26.50 | 21.47 | 22.48 |
| Antipsychotics^3^ | 0.13 | 0.59 | 3.92 | 60.13 | 139.19 | 144.25 | 172.40 | 177.32 | 165.92 | 183.22 | 466.13 |
| Sedatives | 7.29 | 10.21 | 21.27 | 313.43 | 1282.19 | 1745.04 | 2254.70 | 2812.22 | 2974.16 | 2984.47 | 4174.29 |
| **2018** |  |  |  |  |  |  |  |  |  |  |  |
| Benzodiazepines and z-drugs | 5.38 | 8.29 | 17.91 | 207.91 | 1062.50 | 1479.82 | 1823.62 | 2354.69 | 2600.02 | 2509.07 | 3236.88 |
| Long-acting benzodiazepines | 4.78 | 6.73 | 12.71 | 88.91 | 468.43 | 628.91 | 669.64 | 708.95 | 680.37 | 577.19 | 572.07 |
| Clonazepam | 0.93 | 0.93 | 2.12 | 16.13 | 61.92 | 69.56 | 80.72 | 92.61 | 76.89 | 70.65 | 71.66 |
| Diazepam | 1.72 | 2.33 | 6.50 | 55.78 | 343.25 | 439.16 | 420.46 | 408.64 | 388.11 | 327.90 | 319.56 |
| Chloridiazepoxide | 0.03 | 0.04 | 0.00 | 2.83 | 16.77 | 27.16 | 31.33 | 37.02 | 26.47 | 16.48 | 16.19 |
| Potassium Clorazepate | 0.00 | 0.00 | 0.00 | 0.00 | 0.04 | 0.03 | 0.14 | 0.35 | 0.46 | 0.14 | 0.23 |
| Lorazepam | 0.06 | 0.25 | 1.39 | 8.22 | 20.82 | 21.00 | 25.17 | 37.33 | 45.18 | 43.78 | 63.71 |
| Bromazepam | 0.06 | 0.01 | 0.33 | 3.34 | 12.44 | 22.38 | 36.99 | 77.70 | 121.05 | 131.62 | 137.93 |
| Clobazam | 1.61 | 2.82 | 3.15 | 6.42 | 9.47 | 10.49 | 13.72 | 14.68 | 15.38 | 11.44 | 10.44 |
| Prazepam | 0.00 | 0.00 | 0.01 | 0.27 | 1.73 | 3.38 | 6.59 | 14.49 | 22.41 | 23.55 | 23.63 |
| Alprazolam | 0.10 | 0.37 | 1.58 | 45.80 | 211.07 | 259.83 | 302.86 | 382.19 | 402.95 | 385.92 | 479.16 |
| Flurazepam | 0.01 | 0.01 | 0.01 | 6.62 | 33.52 | 76.79 | 111.07 | 132.21 | 134.35 | 110.35 | 93.24 |
| Nitrazepam | 0.48 | 0.60 | 0.92 | 0.85 | 1.75 | 2.32 | 5.61 | 8.95 | 16.30 | 16.69 | 37.12 |
| Triazolam | 0.01 | 0.01 | 0.04 | 2.23 | 22.09 | 32.39 | 41.11 | 64.95 | 82.04 | 83.43 | 132.59 |
| Lormetazepam | 0.00 | 0.09 | 0.03 | 1.11 | 2.70 | 3.75 | 10.54 | 25.45 | 39.70 | 48.31 | 98.16 |
| Temazepam | 0.00 | 0.01 | 0.17 | 1.66 | 8.86 | 17.75 | 29.24 | 62.44 | 88.15 | 101.05 | 185.84 |
| Zopiclone | 0.27 | 0.59 | 1.22 | 34.09 | 209.82 | 310.73 | 414.42 | 556.86 | 601.55 | 597.37 | 844.00 |
| Zolpidem | 0.09 | 0.22 | 0.43 | 22.57 | 106.15 | 182.96 | 293.62 | 438.70 | 538.81 | 540.28 | 723.21 |
| Antidepressants^1^ | 0.12 | 0.12 | 0.38 | 29.16 | 106.79 | 124.75 | 153.56 | 179.39 | 175.94 | 195.00 | 391.46 |
| Antihistamines^2^ | 0.19 | 0.20 | 1.38 | 18.18 | 41.40 | 45.57 | 51.16 | 53.30 | 32.92 | 25.91 | 25.50 |
| Antipsychotics^3^ | 0.15 | 0.77 | 5.78 | 70.01 | 172.97 | 166.35 | 186.43 | 196.51 | 176.59 | 193.36 | 480.91 |
| Sedatives | 5.85 | 9.39 | 25.45 | 325.27 | 1383.66 | 1816.49 | 2214.77 | 2783.88 | 2985.46 | 2923.35 | 4134.75 |
| **2019** |  |  |  |  |  |  |  |  |  |  |  |
| Benzodiazepines and z-drugs | 4.34 | 8.79 | 14.64 | 204.26 | 1046.17 | 1449.07 | 1808.65 | 2340.72 | 2548.26 | 2460.76 | 3153.53 |
| Long-acting benzodiazepines | 3.84 | 7.51 | 10.33 | 87.64 | 456.26 | 622.07 | 670.40 | 708.21 | 663.42 | 556.68 | 564.21 |
| Clonazepam | 0.56 | 1.15 | 1.75 | 16.02 | 61.05 | 69.31 | 79.22 | 93.54 | 76.43 | 72.08 | 77.03 |
| Diazepam | 1.65 | 2.23 | 4.86 | 53.69 | 336.70 | 437.96 | 431.01 | 414.61 | 383.98 | 317.27 | 315.18 |
| Chloridiazepoxide | 0.00 | 0.04 | 0.06 | 3.36 | 17.58 | 31.54 | 31.98 | 38.38 | 24.68 | 14.10 | 14.71 |
| Potassium Clorazepate | 0.00 | 0.00 | 0.00 | 0.00 | 0.04 | 0.01 | 0.19 | 0.40 | 0.63 | 0.13 | 0.15 |
| Lorazepam | 0.00 | 0.19 | 0.68 | 9.75 | 21.33 | 23.04 | 25.10 | 36.62 | 45.39 | 43.93 | 63.77 |
| Bromazepam | 0.04 | 0.01 | 0.16 | 3.58 | 12.67 | 20.36 | 37.37 | 70.97 | 114.17 | 127.22 | 131.91 |
| Clobazam | 1.24 | 2.99 | 2.72 | 6.61 | 9.07 | 10.57 | 12.40 | 13.26 | 14.73 | 11.15 | 9.90 |
| Prazepam | 0.00 | 0.00 | 0.01 | 0.33 | 1.36 | 2.96 | 5.63 | 13.16 | 22.95 | 22.60 | 22.05 |
| Alprazolam | 0.31 | 0.41 | 1.77 | 45.56 | 218.31 | 257.78 | 303.72 | 378.97 | 404.79 | 380.36 | 468.25 |
| Flurazepam | 0.00 | 0.02 | 0.15 | 6.46 | 29.12 | 67.89 | 104.63 | 126.26 | 126.16 | 103.31 | 90.31 |
| Nitrazepam | 0.39 | 1.09 | 0.77 | 1.17 | 1.34 | 1.82 | 5.35 | 8.59 | 13.87 | 16.04 | 34.88 |
| Triazolam | 0.03 | 0.01 | 0.04 | 1.68 | 19.71 | 28.33 | 40.13 | 68.74 | 76.44 | 82.52 | 126.76 |
| Lormetazepam | 0.00 | 0.09 | 0.00 | 0.32 | 1.98 | 1.52 | 5.06 | 12.92 | 19.49 | 26.99 | 53.57 |
| Temazepam | 0.01 | 0.00 | 0.22 | 1.84 | 9.37 | 15.78 | 26.88 | 58.39 | 79.30 | 94.18 | 172.65 |
| Zopiclone | 0.08 | 0.33 | 0.81 | 32.89 | 201.13 | 304.68 | 417.52 | 561.24 | 611.24 | 595.20 | 844.88 |
| Zolpidem | 0.03 | 0.24 | 0.62 | 21.00 | 105.41 | 175.50 | 282.46 | 444.66 | 534.03 | 553.67 | 727.53 |
| Antidepressants^1^ | 0.04 | 0.13 | 0.25 | 30.67 | 118.32 | 131.09 | 165.36 | 194.09 | 199.08 | 206.28 | 412.07 |
| Antihistamines^2^ | 0.15 | 0.34 | 1.77 | 26.23 | 61.15 | 62.52 | 66.52 | 69.24 | 42.13 | 33.02 | 31.75 |
| Antipsychotics^3^ | 0.05 | 0.79 | 6.41 | 86.73 | 204.96 | 192.44 | 205.41 | 219.54 | 194.01 | 201.09 | 503.25 |
| Sedatives | 4.58 | 10.04 | 23.07 | 347.89 | 1430.60 | 1835.12 | 2245.93 | 2823.60 | 2983.48 | 2901.15 | 4100.59 |
| **2020** |  |  |  |  |  |  |  |  |  |  |  |
| Benzodiazepines and z-drugs | 5.31 | 7.65 | 13.68 | 166.25 | 949.46 | 1377.32 | 1757.65 | 2264.59 | 2460.45 | 2405.67 | 3063.81 |
| Long-acting benzodiazepines | 4.85 | 6.60 | 9.78 | 74.38 | 410.47 | 585.67 | 654.00 | 691.29 | 625.11 | 547.74 | 546.06 |
| Clonazepam | 0.58 | 1.10 | 2.07 | 13.25 | 54.47 | 67.28 | 76.65 | 94.03 | 83.88 | 74.85 | 80.64 |
| Diazepam | 1.47 | 1.56 | 4.34 | 46.84 | 304.19 | 421.43 | 429.23 | 412.20 | 357.36 | 314.70 | 305.25 |
| Chloridiazepoxide | 0.04 | 0.06 | 0.05 | 2.38 | 15.75 | 28.69 | 31.46 | 35.07 | 21.45 | 13.57 | 13.22 |
| Potassium Clorazepate | 0.00 | 0.00 | 0.00 | 0.00 | 0.06 | 0.02 | 0.04 | 0.38 | 0.60 | 0.12 | 0.15 |
| Lorazepam | 0.01 | 0.40 | 0.92 | 9.94 | 21.23 | 22.22 | 26.65 | 35.72 | 43.39 | 44.54 | 64.81 |
| Bromazepam | 0.03 | 0.01 | 0.11 | 2.60 | 11.65 | 18.53 | 33.67 | 66.84 | 108.22 | 120.56 | 127.97 |
| Clobazam | 2.42 | 3.05 | 2.15 | 5.74 | 8.53 | 10.18 | 11.67 | 12.52 | 13.44 | 10.30 | 10.14 |
| Prazepam | 0.00 | 0.01 | 0.01 | 0.23 | 0.98 | 2.19 | 5.44 | 11.11 | 17.91 | 20.40 | 18.73 |
| Alprazolam | 0.19 | 0.30 | 1.33 | 34.32 | 203.06 | 244.92 | 292.41 | 358.57 | 396.22 | 373.53 | 455.86 |
| Flurazepam | 0.00 | 0.01 | 0.13 | 4.86 | 24.80 | 54.26 | 94.91 | 117.74 | 118.32 | 98.31 | 86.04 |
| Nitrazepam | 0.34 | 0.81 | 1.02 | 1.09 | 1.69 | 1.61 | 4.60 | 8.25 | 12.14 | 15.49 | 31.89 |
| Triazolam | 0.00 | 0.00 | 0.06 | 1.49 | 16.59 | 27.33 | 38.71 | 66.48 | 71.59 | 84.65 | 122.51 |
| Lormetazepam | 0.00 | 0.00 | 0.05 | 0.01 | 0.08 | 0.07 | 0.35 | 1.08 | 1.98 | 2.00 | 4.26 |
| Temazepam | 0.00 | 0.02 | 0.27 | 1.12 | 9.34 | 14.86 | 25.67 | 53.94 | 72.14 | 87.40 | 161.13 |
| Zopiclone | 0.10 | 0.23 | 0.80 | 26.51 | 181.78 | 292.85 | 407.35 | 548.06 | 608.70 | 585.91 | 845.97 |
| Zolpidem | 0.13 | 0.10 | 0.35 | 15.88 | 95.27 | 170.87 | 278.84 | 442.61 | 533.09 | 559.35 | 735.23 |
| Antidepressants^1^ | 0.04 | 0.13 | 0.26 | 30.75 | 127.29 | 143.92 | 178.34 | 213.76 | 218.00 | 226.25 | 442.87 |
| Antihistamines^2^ | 0.01 | 0.39 | 1.56 | 28.07 | 77.21 | 82.65 | 88.74 | 87.10 | 56.28 | 41.37 | 42.16 |
| Antipsychotics^3^ | 0.01 | 0.96 | 6.03 | 84.57 | 227.46 | 214.59 | 228.91 | 236.62 | 206.19 | 210.83 | 521.47 |
| Sedatives | 5.38 | 9.13 | 21.53 | 309.64 | 1381.41 | 1818.48 | 2253.63 | 2802.07 | 2940.93 | 2884.12 | 4070.32 |
| **2021** |  |  |  |  |  |  |  |  |  |  |  |
| Benzodiazepines and z-drugs | 5.48 | 9.04 | 12.21 | 179.47 | 957.83 | 1402.18 | 1742.54 | 2229.48 | 2407.23 | 2341.88 | 2950.66 |
| Long-acting benzodiazepines | 5.23 | 8.09 | 8.64 | 85.16 | 425.74 | 600.55 | 657.15 | 686.90 | 618.31 | 522.99 | 512.53 |
| Clonazepam | 0.91 | 1.53 | 1.63 | 14.42 | 57.66 | 71.11 | 78.16 | 92.77 | 88.01 | 73.04 | 84.27 |
| Diazepam | 1.15 | 2.11 | 4.04 | 54.09 | 317.89 | 433.55 | 439.00 | 422.89 | 360.30 | 310.14 | 294.66 |
| Chloridiazepoxide | 0.02 | 0.03 | 0.03 | 4.05 | 15.77 | 30.79 | 31.38 | 36.12 | 20.09 | 12.63 | 11.88 |
| Potassium Clorazepate | 0.00 | 0.00 | 0.00 | 0.00 | 0.19 | 0.00 | 0.03 | 0.24 | 0.22 | 0.32 | 0.17 |
| Lorazepam | 0.00 | 0.45 | 1.11 | 11.31 | 23.76 | 23.61 | 27.55 | 36.72 | 45.28 | 42.62 | 62.74 |
| Bromazepam | 0.00 | 0.01 | 0.18 | 2.43 | 12.54 | 17.30 | 29.80 | 60.51 | 101.27 | 110.49 | 124.49 |
| Clobazam | 2.72 | 3.85 | 2.62 | 6.13 | 9.11 | 10.37 | 11.84 | 13.50 | 11.94 | 10.33 | 9.32 |
| Prazepam | 0.00 | 0.01 | 0.00 | 0.49 | 1.07 | 2.57 | 5.07 | 9.12 | 16.77 | 19.41 | 18.20 |
| Alprazolam | 0.06 | 0.23 | 1.17 | 34.19 | 197.01 | 253.31 | 290.38 | 354.55 | 382.87 | 365.92 | 438.38 |
| Flurazepam | 0.00 | 0.04 | 0.00 | 5.50 | 23.44 | 51.56 | 90.10 | 109.53 | 117.23 | 91.67 | 82.56 |
| Nitrazepam | 0.43 | 0.51 | 0.31 | 0.47 | 0.60 | 0.60 | 1.56 | 2.74 | 3.75 | 5.44 | 11.48 |
| Triazolam | 0.00 | 0.00 | 0.01 | 1.54 | 14.61 | 26.60 | 37.25 | 61.26 | 70.51 | 81.03 | 117.68 |
| Lormetazepam | 0.00 | 0.00 | 0.00 | 0.00 | 0.00 | 0.01 | 0.03 | 0.04 | 0.23 | 0.28 | 0.43 |
| Temazepam | 0.00 | 0.00 | 0.16 | 1.17 | 7.90 | 13.62 | 24.88 | 49.09 | 67.93 | 82.41 | 148.74 |
| Zopiclone | 0.13 | 0.06 | 0.61 | 27.19 | 179.32 | 299.35 | 401.58 | 547.77 | 593.66 | 585.30 | 826.16 |
| Zolpidem | 0.06 | 0.19 | 0.33 | 16.49 | 96.96 | 167.83 | 273.92 | 432.62 | 527.18 | 550.85 | 719.52 |
| Antidepressants^1^ | 0.02 | 0.18 | 0.07 | 34.43 | 144.38 | 160.83 | 194.29 | 235.29 | 238.66 | 245.24 | 476.99 |
| Antihistamines^2^ | 0.05 | 0.36 | 2.39 | 36.64 | 100.61 | 105.40 | 111.65 | 113.71 | 72.84 | 50.92 | 50.79 |
| Antipsychotics^3^ | 0.00 | 0.86 | 5.20 | 96.49 | 257.15 | 247.06 | 249.20 | 256.51 | 231.07 | 217.33 | 526.94 |
| Sedatives | 5.54 | 10.45 | 19.87 | 347.03 | 1459.96 | 1915.47 | 2297.68 | 2834.99 | 2949.81 | 2855.38 | 4005.37 |
| **2022** |  |  |  |  |  |  |  |  |  |  |  |
| Benzodiazepines and z-drugs | 5.61 | 7.80 | 12.83 | 164.79 | 849.15 | 1318.20 | 1672.42 | 2108.42 | 2291.51 | 2233.35 | 2826.32 |
| Long-acting benzodiazepines | 5.31 | 6.77 | 9.14 | 78.59 | 371.55 | 561.48 | 627.89 | 650.66 | 582.42 | 499.01 | 480.11 |
| Clonazepam | 0.75 | 1.27 | 1.63 | 10.73 | 45.99 | 61.35 | 68.67 | 79.83 | 82.14 | 66.25 | 77.19 |
| Diazepam | 1.45 | 1.92 | 4.39 | 50.82 | 280.47 | 418.76 | 435.24 | 418.59 | 355.85 | 310.06 | 292.18 |
| Chloridiazepoxide | 0.00 | 0.02 | 0.02 | 4.35 | 14.66 | 26.55 | 28.34 | 31.11 | 18.17 | 13.45 | 10.55 |
| Potassium Clorazepate | 0.00 | 0.00 | 0.00 | 0.00 | 0.03 | 0.07 | 0.00 | 0.30 | 0.24 | 0.21 | 0.21 |
| Lorazepam | 0.08 | 0.63 | 1.64 | 11.84 | 23.28 | 25.54 | 29.13 | 34.79 | 42.33 | 41.68 | 60.94 |
| Bromazepam | 0.03 | 0.00 | 0.19 | 2.12 | 11.24 | 17.53 | 27.51 | 54.67 | 92.05 | 102.36 | 118.93 |
| Clobazam | 3.01 | 3.32 | 3.06 | 6.74 | 10.63 | 10.02 | 11.82 | 14.75 | 12.44 | 9.53 | 9.42 |
| Prazepam | 0.00 | 0.00 | 0.00 | 0.21 | 0.53 | 1.10 | 2.95 | 4.64 | 9.77 | 11.18 | 12.04 |
| Alprazolam | 0.08 | 0.16 | 0.94 | 31.08 | 185.62 | 247.68 | 286.80 | 344.04 | 371.61 | 351.71 | 422.18 |
| Flurazepam | 0.00 | 0.01 | 0.03 | 5.66 | 19.13 | 43.52 | 80.61 | 101.18 | 103.61 | 88.05 | 77.31 |
| Nitrazepam | 0.10 | 0.23 | 0.01 | 0.08 | 0.10 | 0.12 | 0.27 | 0.26 | 0.20 | 0.27 | 1.20 |
| Triazolam | 0.00 | 0.00 | 0.10 | 1.03 | 12.61 | 24.51 | 35.80 | 56.40 | 63.91 | 75.73 | 111.45 |
| Lormetazepam | 0.00 | 0.00 | 0.00 | 0.00 | 0.01 | 0.00 | 0.01 | 0.02 | 0.00 | 0.12 | 0.13 |
| Temazepam | 0.00 | 0.01 | 0.02 | 1.19 | 6.13 | 11.13 | 21.01 | 42.37 | 60.40 | 73.92 | 136.22 |
| Zopiclone | 0.05 | 0.16 | 0.43 | 24.02 | 160.51 | 280.47 | 382.23 | 519.93 | 571.45 | 562.62 | 797.65 |
| Zolpidem | 0.06 | 0.08 | 0.36 | 14.92 | 78.21 | 149.85 | 262.03 | 405.55 | 507.35 | 526.20 | 698.72 |
| Antidepressants^1^ | 0.00 | 0.17 | 0.23 | 28.02 | 130.35 | 153.95 | 194.49 | 240.09 | 236.55 | 248.51 | 489.19 |
| Antihistamines^2^ | 0.02 | 0.46 | 2.87 | 31.45 | 100.51 | 110.94 | 125.63 | 124.03 | 82.56 | 58.21 | 58.46 |
| Antipsychotics^3^ | 0.00 | 0.53 | 4.01 | 82.73 | 252.11 | 250.48 | 257.92 | 262.42 | 230.64 | 213.86 | 514.89 |
| Sedatives | 5.63 | 8.97 | 19.94 | 307.00 | 1332.12 | 1833.57 | 2250.46 | 2734.96 | 2841.25 | 2753.92 | 3888.85 |

**Rate of claims per 1,000 GMS population by sex**

|  | **Females** | **Males** |
| --- | --- | --- |
| **2014** |  |  |
| Benzodiazepines and z-drugs | 1864.50 | 1133.12 |
| Long-acting benzodiazepines | 483.20 | 384.22 |
| Clonazepam | 46.26 | 41.05 |
| Diazepam | 291.82 | 220.04 |
| Chloridiazepoxide | 14.40 | 23.06 |
| Potassium Clorazepate | 0.18 | 0.07 |
| Lorazepam | 29.75 | 21.73 |
| Bromazepam | 79.80 | 29.42 |
| Clobazam | 12.31 | 11.62 |
| Prazepam | 14.68 | 6.31 |
| Alprazolam | 309.94 | 157.56 |
| Flurazepam | 85.03 | 74.08 |
| Nitrazepam | 18.52 | 7.99 |
| Triazolam | 60.19 | 34.68 |
| Lormetazepam | 43.30 | 17.22 |
| Temazepam | 89.57 | 46.86 |
| Zopiclone | 409.56 | 263.83 |
| Zolpidem | 354.12 | 174.33 |
| Antidepressants^1^ | 110.73 | 70.30 |
| Antihistamines^2^ | 12.08 | 8.79 |
| Antipsychotics^3^ | 134.97 | 99.07 |
| Sedatives | 2122.28 | 1311.28 |
| **2015** |  |  |
| Benzodiazepines and z-drugs | 1847.76 | 1122.96 |
| Long-acting benzodiazepines | 473.18 | 380.72 |
| Clonazepam | 47.33 | 42.18 |
| Diazepam | 290.69 | 221.38 |
| Chloridiazepoxide | 13.43 | 22.63 |
| Potassium Clorazepate | 0.20 | 0.07 |
| Lorazepam | 29.16 | 21.18 |
| Bromazepam | 78.19 | 28.45 |
| Clobazam | 10.80 | 10.48 |
| Prazepam | 13.98 | 6.17 |
| Alprazolam | 312.27 | 156.76 |
| Flurazepam | 80.13 | 70.69 |
| Nitrazepam | 16.63 | 7.12 |
| Triazolam | 60.28 | 34.90 |
| Lormetazepam | 40.30 | 15.90 |
| Temazepam | 81.47 | 42.75 |
| Zopiclone | 412.29 | 266.21 |
| Zolpidem | 354.68 | 172.51 |
| Antidepressants^1^ | 125.77 | 80.82 |
| Antihistamines^2^ | 15.06 | 11.09 |
| Antipsychotics^3^ | 149.77 | 110.63 |
| Sedatives | 2138.35 | 1325.49 |
| **2016** |  |  |
| Benzodiazepines and z-drugs | 1890.12 | 1160.16 |
| Long-acting benzodiazepines | 479.42 | 391.34 |
| Clonazepam | 50.94 | 45.83 |
| Diazepam | 298.18 | 228.52 |
| Chloridiazepoxide | 13.33 | 23.80 |
| Potassium Clorazepate | 0.17 | 0.05 |
| Lorazepam | 29.39 | 22.20 |
| Bromazepam | 78.54 | 28.76 |
| Clobazam | 10.14 | 10.32 |
| Prazepam | 13.54 | 5.98 |
| Alprazolam | 323.02 | 163.49 |
| Flurazepam | 77.23 | 69.89 |
| Nitrazepam | 15.88 | 6.95 |
| Triazolam | 60.58 | 35.31 |
| Lormetazepam | 39.56 | 15.73 |
| Temazepam | 77.96 | 41.37 |
| Zopiclone | 427.02 | 280.27 |
| Zolpidem | 370.80 | 179.40 |
| Antidepressants^1^ | 141.71 | 91.91 |
| Antihistamines^2^ | 20.30 | 14.96 |
| Antipsychotics^3^ | 165.72 | 123.94 |
| Sedatives | 2217.85 | 1390.97 |
| **2017** |  |  |
| Benzodiazepines and z-drugs | 1895.30 | 1177.53 |
| Long-acting benzodiazepines | 479.26 | 397.62 |
| Clonazepam | 54.45 | 49.41 |
| Diazepam | 299.79 | 233.54 |
| Chloridiazepoxide | 13.05 | 23.76 |
| Potassium Clorazepate | 0.18 | 0.10 |
| Lorazepam | 28.91 | 23.22 |
| Bromazepam | 77.69 | 28.95 |
| Clobazam | 9.80 | 9.94 |
| Prazepam | 13.13 | 5.71 |
| Alprazolam | 324.08 | 166.25 |
| Flurazepam | 74.25 | 68.45 |
| Nitrazepam | 14.61 | 6.71 |
| Triazolam | 60.10 | 35.49 |
| Lormetazepam | 37.81 | 14.57 |
| Temazepam | 73.51 | 39.13 |
| Zopiclone | 434.59 | 288.18 |
| Zolpidem | 378.90 | 183.93 |
| Antidepressants^1^ | 156.83 | 102.65 |
| Antihistamines^2^ | 27.57 | 20.17 |
| Antipsychotics^3^ | 183.11 | 138.06 |
| Sedatives | 2262.80 | 1438.41 |
| **2018** |  |  |
| Benzodiazepines and z-drugs | 1899.43 | 1184.63 |
| Long-acting benzodiazepines | 477.82 | 399.51 |
| Clonazepam | 56.69 | 51.87 |
| Diazepam | 300.36 | 236.35 |
| Chloridiazepoxide | 12.35 | 23.57 |
| Potassium Clorazepate | 0.16 | 0.10 |
| Lorazepam | 30.15 | 24.17 |
| Bromazepam | 76.99 | 28.31 |
| Clobazam | 9.55 | 9.61 |
| Prazepam | 12.81 | 5.78 |
| Alprazolam | 325.67 | 165.87 |
| Flurazepam | 72.52 | 66.02 |
| Nitrazepam | 13.38 | 6.20 |
| Triazolam | 59.72 | 35.54 |
| Lormetazepam | 35.79 | 13.58 |
| Temazepam | 68.90 | 36.41 |
| Zopiclone | 438.56 | 292.65 |
| Zolpidem | 385.70 | 188.55 |
| Antidepressants^1^ | 174.92 | 113.08 |
| Antihistamines^2^ | 33.84 | 24.51 |
| Antipsychotics^3^ | 201.95 | 152.54 |
| Sedatives | 2310.14 | 1474.76 |
| **2019** |  |  |
| Benzodiazepines and z-drugs | 1891.54 | 1183.62 |
| Long-acting benzodiazepines | 474.39 | 399.83 |
| Clonazepam | 57.53 | 53.52 |
| Diazepam | 301.15 | 237.84 |
| Chloridiazepoxide | 12.29 | 24.65 |
| Potassium Clorazepate | 0.17 | 0.11 |
| Lorazepam | 30.56 | 25.17 |
| Bromazepam | 75.35 | 27.00 |
| Clobazam | 9.03 | 9.26 |
| Prazepam | 12.19 | 5.59 |
| Alprazolam | 327.65 | 166.26 |
| Flurazepam | 69.35 | 62.70 |
| Nitrazepam | 12.66 | 6.17 |
| Triazolam | 59.02 | 34.81 |
| Lormetazepam | 19.81 | 7.18 |
| Temazepam | 66.03 | 33.91 |
| Zopiclone | 445.39 | 298.28 |
| Zolpidem | 393.33 | 191.18 |
| Antidepressants^1^ | 189.24 | 123.65 |
| Antihistamines^2^ | 44.99 | 32.64 |
| Antipsychotics^3^ | 221.89 | 170.45 |
| Sedatives | 2347.66 | 1510.36 |
| **2020** |  |  |
| Benzodiazepines and z-drugs | 1825.39 | 1131.53 |
| Long-acting benzodiazepines | 455.37 | 380.75 |
| Clonazepam | 58.34 | 52.51 |
| Diazepam | 290.48 | 230.39 |
| Chloridiazepoxide | 11.30 | 22.66 |
| Potassium Clorazepate | 0.16 | 0.08 |
| Lorazepam | 31.02 | 24.92 |
| Bromazepam | 71.21 | 25.47 |
| Clobazam | 8.73 | 8.74 |
| Prazepam | 10.34 | 4.73 |
| Alprazolam | 316.93 | 155.76 |
| Flurazepam | 64.66 | 55.60 |
| Nitrazepam | 11.37 | 6.03 |
| Triazolam | 56.40 | 34.27 |
| Lormetazepam | 1.57 | 0.58 |
| Temazepam | 61.23 | 31.71 |
| Zopiclone | 439.26 | 289.29 |
| Zolpidem | 392.40 | 188.78 |
| Antidepressants^1^ | 206.06 | 132.75 |
| Antihistamines^2^ | 58.10 | 41.86 |
| Antipsychotics^3^ | 238.53 | 180.03 |
| Sedatives | 2328.09 | 1486.18 |
| **2021** |  |  |
| Benzodiazepines and z-drugs | 1823.41 | 1144.63 |
| Long-acting benzodiazepines | 453.20 | 387.48 |
| Clonazepam | 60.64 | 55.05 |
| Diazepam | 295.84 | 238.53 |
| Chloridiazepoxide | 11.23 | 23.44 |
| Potassium Clorazepate | 0.15 | 0.07 |
| Lorazepam | 32.02 | 25.76 |
| Bromazepam | 69.22 | 24.27 |
| Clobazam | 8.75 | 9.16 |
| Prazepam | 9.95 | 4.73 |
| Alprazolam | 316.11 | 156.94 |
| Flurazepam | 62.51 | 54.24 |
| Nitrazepam | 4.12 | 2.28 |
| Triazolam | 55.35 | 33.73 |
| Lormetazepam | 0.16 | 0.06 |
| Temazepam | 58.34 | 30.88 |
| Zopiclone | 443.74 | 294.59 |
| Zolpidem | 395.27 | 190.91 |
| Antidepressants^1^ | 229.95 | 148.76 |
| Antihistamines^2^ | 74.54 | 53.15 |
| Antipsychotics^3^ | 261.29 | 193.38 |
| Sedatives | 2389.20 | 1539.92 |
| **2022** |  |  |
| Benzodiazepines and z-drugs | 1729.91 | 1100.11 |
| Long-acting benzodiazepines | 424.04 | 368.90 |
| Clonazepam | 53.40 | 49.07 |
| Diazepam | 288.65 | 235.40 |
| Chloridiazepoxide | 10.02 | 21.33 |
| Potassium Clorazepate | 0.14 | 0.07 |
| Lorazepam | 31.32 | 26.19 |
| Bromazepam | 64.45 | 23.27 |
| Clobazam | 9.10 | 9.32 |
| Prazepam | 5.97 | 2.78 |
| Alprazolam | 305.46 | 153.85 |
| Flurazepam | 56.35 | 50.64 |
| Nitrazepam | 0.41 | 0.29 |
| Triazolam | 51.81 | 31.87 |
| Lormetazepam | 0.06 | 0.01 |
| Temazepam | 52.36 | 27.92 |
| Zopiclone | 423.79 | 284.66 |
| Zolpidem | 376.64 | 183.45 |
| Antidepressants^1^ | 230.49 | 151.29 |
| Antihistamines^2^ | 80.58 | 58.39 |
| Antipsychotics^3^ | 260.62 | 192.84 |
| Sedatives | 2301.60 | 1502.63 |

**Rate of cost per 1,000 GMS population by age group**

|  | **<5** | **5-11** | **12-15** | **16-24** | **25-34** | **35-44** | **45-54** | **55-64** | **65-69** | **70-74** | **75+** |
| --- | --- | --- | --- | --- | --- | --- | --- | --- | --- | --- | --- |
| **2014** |  |  |  |  |  |  |  |  |  |  |  |
| Benzodiazepines and z-drugs | 552.46 | 400.71 | 272.21 | 1237.23 | 6472.87 | 9979.03 | 12775.42 | 17068.62 | 17857.97 | 17974.89 | 25119.03 |
| Long-acting benzodiazepines | 548.09 | 388.40 | 246.95 | 571.45 | 2927.49 | 4295.49 | 4647.04 | 5250.34 | 4602.92 | 3928.57 | 4137.80 |
| Clonazepam | 166.12 | 156.91 | 104.28 | 107.37 | 370.57 | 463.50 | 662.22 | 668.48 | 554.72 | 468.46 | 475.94 |
| Diazepam | 27.93 | 32.18 | 34.22 | 306.67 | 1853.03 | 2447.47 | 2086.97 | 2142.19 | 1820.23 | 1596.71 | 1779.84 |
| Chloridiazepoxide | 0.03 | 0.16 | 4.10 | 13.70 | 80.21 | 127.49 | 223.21 | 270.25 | 184.27 | 165.67 | 167.40 |
| Potassium Clorazepate | 0.00 | 0.00 | 0.00 | 0.56 | 0.18 | 0.60 | 5.57 | 3.88 | 2.69 | 1.89 | 1.55 |
| Lorazepam | 0.05 | 0.88 | 2.87 | 23.67 | 68.15 | 90.13 | 156.87 | 218.85 | 302.82 | 270.17 | 414.02 |
| Bromazepam | 0.00 | 0.04 | 1.07 | 13.89 | 74.50 | 135.34 | 352.58 | 768.53 | 1033.50 | 1038.46 | 1107.22 |
| Clobazam | 338.51 | 195.21 | 92.75 | 82.30 | 93.90 | 136.91 | 144.14 | 177.63 | 123.45 | 103.16 | 91.12 |
| Prazepam | 0.03 | 0.03 | 0.03 | 2.49 | 8.28 | 18.24 | 58.31 | 144.45 | 186.41 | 192.45 | 160.58 |
| Alprazolam | 1.16 | 2.05 | 11.19 | 192.69 | 849.46 | 1123.72 | 1486.06 | 1962.59 | 2036.67 | 1994.36 | 2642.70 |
| Flurazepam | 0.10 | 0.33 | 9.53 | 50.92 | 506.30 | 1077.95 | 1424.78 | 1767.36 | 1628.05 | 1255.76 | 1173.84 |
| Nitrazepam | 15.38 | 3.59 | 2.05 | 7.44 | 15.01 | 23.33 | 41.84 | 76.08 | 103.10 | 144.47 | 287.54 |
| Triazolam | 0.04 | 0.35 | 0.03 | 17.90 | 147.23 | 180.84 | 282.57 | 429.60 | 548.96 | 602.02 | 984.23 |
| Lormetazepam | 0.14 | 0.29 | 1.23 | 5.82 | 13.56 | 38.08 | 117.86 | 233.63 | 378.22 | 547.11 | 983.35 |
| Temazepam | 0.55 | 0.06 | 0.35 | 12.74 | 58.73 | 142.37 | 312.60 | 572.11 | 854.88 | 897.08 | 1896.19 |
| Zopiclone | 0.91 | 4.63 | 4.26 | 255.69 | 1703.18 | 2827.44 | 3534.22 | 4771.09 | 4897.91 | 5221.85 | 8066.54 |
| Zolpidem | 1.02 | 3.22 | 3.29 | 129.38 | 620.53 | 1125.28 | 1849.86 | 2821.84 | 3148.51 | 3438.20 | 4823.63 |
| Antidepressants^1^ | 0.83 | 0.87 | 1.41 | 171.64 | 588.29 | 884.85 | 1329.60 | 1594.32 | 1647.20 | 1926.69 | 4029.71 |
| Antihistamines^2^ | 0.00 | 0.65 | 2.93 | 33.66 | 71.44 | 90.51 | 121.26 | 130.15 | 72.29 | 52.54 | 69.26 |
| Antipsychotics^3^ | 0.32 | 3.59 | 41.37 | 466.27 | 1326.66 | 1726.38 | 2310.44 | 2526.18 | 2545.55 | 2976.20 | 6270.37 |
| Sedatives | 553.61 | 405.81 | 317.92 | 1908.81 | 8459.26 | 12680.76 | 16536.72 | 21319.28 | 22123.01 | 22930.33 | 35488.36 |
| **2015** |  |  |  |  |  |  |  |  |  |  |  |
| Benzodiazepines and z-drugs | 371.72 | 333.22 | 276.06 | 1231.09 | 6591.47 | 9530.51 | 12883.70 | 16489.07 | 17169.18 | 16973.51 | 23285.87 |
| Long-acting benzodiazepines | 368.75 | 322.98 | 239.34 | 606.91 | 3152.66 | 4366.02 | 5037.44 | 5464.14 | 4723.41 | 4044.48 | 4123.73 |
| Clonazepam | 69.61 | 122.87 | 87.28 | 119.09 | 405.70 | 482.05 | 690.53 | 714.71 | 560.94 | 451.41 | 457.74 |
| Diazepam | 19.07 | 42.84 | 47.39 | 334.19 | 2096.34 | 2553.59 | 2406.60 | 2343.60 | 2002.04 | 1705.66 | 1881.24 |
| Chloridiazepoxide | 0.03 | 0.08 | 4.51 | 8.81 | 77.21 | 140.20 | 204.24 | 239.65 | 185.40 | 153.15 | 155.82 |
| Potassium Clorazepate | 0.00 | 0.00 | 0.00 | 0.22 | 1.01 | 0.52 | 6.15 | 4.63 | 2.43 | 1.02 | 0.79 |
| Lorazepam | 0.02 | 0.41 | 4.56 | 24.35 | 82.10 | 96.46 | 156.41 | 216.80 | 278.59 | 276.58 | 386.44 |
| Bromazepam | 0.03 | 0.06 | 0.80 | 18.07 | 65.92 | 137.96 | 323.05 | 716.49 | 1002.99 | 966.67 | 1088.07 |
| Clobazam | 271.42 | 155.41 | 83.19 | 74.82 | 75.32 | 106.96 | 148.89 | 144.00 | 123.56 | 98.22 | 95.98 |
| Prazepam | 0.00 | 0.00 | 0.05 | 1.67 | 7.78 | 18.09 | 48.17 | 136.07 | 174.83 | 173.55 | 157.83 |
| Alprazolam | 0.77 | 2.61 | 12.80 | 173.18 | 890.14 | 1099.07 | 1505.44 | 1928.74 | 2022.27 | 2045.93 | 2595.06 |
| Flurazepam | 0.03 | 0.58 | 14.97 | 59.21 | 477.70 | 1045.24 | 1491.81 | 1814.45 | 1572.21 | 1320.10 | 1116.47 |
| Nitrazepam | 8.60 | 1.20 | 1.94 | 8.91 | 11.60 | 19.37 | 41.05 | 67.02 | 102.00 | 141.37 | 257.87 |
| Triazolam | 0.03 | 0.04 | 0.50 | 14.86 | 157.15 | 180.77 | 272.31 | 434.81 | 593.45 | 564.68 | 996.21 |
| Lormetazepam | 0.03 | 0.02 | 1.47 | 2.69 | 17.80 | 34.80 | 103.28 | 211.70 | 318.84 | 515.14 | 907.90 |
| Temazepam | 0.00 | 1.03 | 0.27 | 14.79 | 56.02 | 135.84 | 300.86 | 535.72 | 776.84 | 818.21 | 1726.43 |
| Zopiclone | 0.52 | 2.99 | 9.32 | 242.90 | 1539.36 | 2398.00 | 3222.82 | 4112.16 | 4202.65 | 4341.36 | 6689.06 |
| Zolpidem | 1.16 | 0.95 | 6.03 | 122.73 | 616.42 | 1052.49 | 1924.72 | 2824.57 | 3195.41 | 3362.16 | 4706.89 |
| Antidepressants^1^ | 1.91 | 0.48 | 7.32 | 199.52 | 655.51 | 953.12 | 1400.01 | 1690.17 | 1840.87 | 2019.36 | 4204.58 |
| Antihistamines^2^ | 0.00 | 0.66 | 5.01 | 35.71 | 104.69 | 129.51 | 164.32 | 151.33 | 86.28 | 64.52 | 78.31 |
| Antipsychotics^3^ | 0.19 | 5.36 | 52.28 | 562.43 | 1402.48 | 1753.51 | 2265.37 | 2482.37 | 2541.83 | 2576.03 | 5950.91 |
| Sedatives | 373.82 | 339.71 | 340.67 | 2028.75 | 8754.15 | 12366.66 | 16713.40 | 20812.95 | 21638.16 | 21633.43 | 33519.67 |
| **2016** |  |  |  |  |  |  |  |  |  |  |  |
| Benzodiazepines and z-drugs | 454.79 | 354.61 | 174.78 | 1309.52 | 6982.67 | 9961.34 | 13046.72 | 16204.70 | 17088.53 | 16869.65 | 22860.85 |
| Long-acting benzodiazepines | 434.28 | 344.45 | 144.41 | 657.49 | 3360.77 | 4539.83 | 5245.96 | 5332.81 | 4673.34 | 4007.50 | 4002.16 |
| Clonazepam | 128.67 | 108.48 | 46.32 | 143.19 | 462.92 | 541.45 | 666.98 | 726.35 | 558.76 | 503.72 | 485.97 |
| Diazepam | 45.31 | 26.95 | 30.37 | 319.58 | 2236.27 | 2701.49 | 2615.80 | 2323.66 | 2032.26 | 1691.91 | 1821.53 |
| Chloridiazepoxide | 0.06 | 0.18 | 0.26 | 12.63 | 89.04 | 161.02 | 189.47 | 239.50 | 184.93 | 154.00 | 143.16 |
| Potassium Clorazepate | 0.00 | 0.00 | 0.00 | 0.00 | 1.05 | 0.13 | 4.45 | 2.01 | 6.40 | 0.18 | 1.22 |
| Lorazepam | 0.23 | 0.51 | 7.08 | 30.35 | 87.68 | 104.72 | 166.02 | 221.70 | 252.36 | 253.08 | 378.10 |
| Bromazepam | 0.06 | 0.22 | 0.57 | 18.88 | 77.39 | 142.07 | 296.75 | 650.61 | 955.12 | 950.71 | 1045.31 |
| Clobazam | 253.16 | 205.20 | 64.44 | 109.03 | 77.11 | 101.24 | 148.16 | 129.42 | 117.83 | 88.02 | 100.01 |
| Prazepam | 0.00 | 0.00 | 0.03 | 1.68 | 6.72 | 16.81 | 46.89 | 124.93 | 144.14 | 165.88 | 160.16 |
| Alprazolam | 0.82 | 2.07 | 5.66 | 187.60 | 926.51 | 1147.07 | 1519.34 | 1895.61 | 2014.79 | 2010.49 | 2564.64 |
| Flurazepam | 3.18 | 1.22 | 0.22 | 65.05 | 476.56 | 1004.40 | 1527.85 | 1730.70 | 1531.67 | 1280.02 | 1043.84 |
| Nitrazepam | 3.90 | 2.43 | 2.77 | 6.32 | 11.09 | 13.30 | 46.35 | 56.23 | 97.34 | 123.77 | 246.26 |
| Triazolam | 0.47 | 0.23 | 0.57 | 23.50 | 167.86 | 185.31 | 270.89 | 418.34 | 583.04 | 534.63 | 987.51 |
| Lormetazepam | 0.00 | 0.06 | 1.27 | 2.87 | 19.28 | 29.54 | 90.15 | 202.61 | 311.48 | 472.80 | 872.38 |
| Temazepam | 0.00 | 0.36 | 0.16 | 11.29 | 48.75 | 129.38 | 285.84 | 514.84 | 735.41 | 804.30 | 1595.88 |
| Zopiclone | 16.55 | 4.10 | 10.04 | 255.25 | 1643.55 | 2556.32 | 3250.94 | 4137.08 | 4245.65 | 4399.93 | 6566.53 |
| Zolpidem | 2.15 | 1.91 | 4.09 | 117.28 | 638.36 | 1111.88 | 1897.40 | 2802.95 | 3288.39 | 3403.87 | 4805.90 |
| Antidepressants^1^ | 0.85 | 0.71 | 1.42 | 190.49 | 703.10 | 972.03 | 1402.00 | 1688.84 | 1655.86 | 1955.58 | 4052.25 |
| Antihistamines^2^ | 0.04 | 0.58 | 7.04 | 50.11 | 140.94 | 183.12 | 194.26 | 212.80 | 113.27 | 92.91 | 100.15 |
| Antipsychotics^3^ | 1.69 | 7.78 | 40.06 | 670.05 | 1642.19 | 1848.99 | 2292.57 | 2441.43 | 2378.38 | 2462.65 | 5695.20 |
| Sedatives | 457.37 | 363.68 | 223.29 | 2220.17 | 9468.89 | 12965.47 | 16935.54 | 20547.78 | 21236.03 | 21380.79 | 32708.45 |
| **2017** |  |  |  |  |  |  |  |  |  |  |  |
| Benzodiazepines and z-drugs | 362.50 | 445.38 | 192.82 | 1172.22 | 7213.42 | 10360.44 | 13147.10 | 15812.83 | 16361.07 | 16699.33 | 22173.32 |
| Long-acting benzodiazepines | 343.75 | 433.31 | 171.60 | 588.70 | 3477.56 | 4734.73 | 5378.02 | 5273.74 | 4483.36 | 3957.07 | 3816.66 |
| Clonazepam | 120.22 | 205.18 | 100.95 | 139.00 | 526.76 | 558.12 | 657.42 | 769.93 | 528.33 | 576.25 | 485.73 |
| Diazepam | 33.78 | 18.46 | 32.86 | 290.52 | 2319.29 | 2911.31 | 2772.43 | 2354.32 | 1941.29 | 1670.21 | 1765.37 |
| Chloridiazepoxide | 0.00 | 0.13 | 0.61 | 12.06 | 82.09 | 156.65 | 194.15 | 212.63 | 177.11 | 138.47 | 138.45 |
| Potassium Clorazepate | 0.00 | 0.00 | 0.00 | 0.10 | 0.03 | 1.46 | 1.02 | 6.66 | 4.20 | 2.68 | 1.69 |
| Lorazepam | 0.21 | 0.75 | 6.54 | 25.89 | 98.66 | 112.61 | 155.55 | 231.60 | 238.52 | 256.99 | 360.21 |
| Bromazepam | 0.00 | 0.05 | 0.91 | 16.41 | 73.90 | 143.05 | 275.88 | 590.32 | 900.96 | 946.82 | 999.59 |
| Clobazam | 184.56 | 205.86 | 32.67 | 85.09 | 81.05 | 90.05 | 142.02 | 124.99 | 119.38 | 79.52 | 82.71 |
| Prazepam | 0.00 | 0.02 | 0.10 | 1.17 | 8.38 | 15.95 | 40.79 | 103.76 | 137.31 | 160.42 | 157.28 |
| Alprazolam | 0.95 | 2.01 | 3.34 | 195.56 | 966.21 | 1256.44 | 1497.10 | 1855.05 | 1924.08 | 1989.92 | 2543.85 |
| Flurazepam | 3.73 | 0.07 | 1.14 | 56.09 | 447.41 | 987.36 | 1529.12 | 1650.31 | 1494.34 | 1214.94 | 962.25 |
| Nitrazepam | 1.46 | 3.60 | 3.26 | 4.68 | 12.54 | 13.83 | 41.08 | 51.14 | 81.41 | 114.59 | 223.18 |
| Triazolam | 0.00 | 0.04 | 0.94 | 14.23 | 166.04 | 209.72 | 278.81 | 403.96 | 548.79 | 532.28 | 956.80 |
| Lormetazepam | 0.05 | 0.53 | 0.07 | 5.30 | 17.49 | 22.41 | 90.39 | 191.33 | 284.25 | 409.19 | 800.85 |
| Temazepam | 0.31 | 0.03 | 0.19 | 8.32 | 58.12 | 122.74 | 259.73 | 473.58 | 658.22 | 764.79 | 1462.39 |
| Zopiclone | 15.84 | 6.09 | 6.92 | 211.45 | 1719.82 | 2627.02 | 3332.22 | 4071.64 | 4126.25 | 4369.92 | 6433.73 |
| Zolpidem | 1.39 | 2.54 | 2.28 | 105.66 | 633.96 | 1129.41 | 1878.05 | 2718.59 | 3192.33 | 3469.49 | 4794.80 |
| Antidepressants^1^ | 1.34 | 0.14 | 4.31 | 207.08 | 809.16 | 1079.36 | 1443.96 | 1686.38 | 1606.07 | 1893.57 | 3859.54 |
| Antihistamines^2^ | 0.00 | 0.35 | 5.70 | 69.41 | 191.81 | 233.82 | 253.17 | 262.52 | 149.49 | 136.63 | 126.61 |
| Antipsychotics^3^ | 2.24 | 4.55 | 28.22 | 537.63 | 1402.82 | 1590.45 | 1927.61 | 1992.64 | 1831.16 | 1910.78 | 4271.10 |
| Sedatives | 366.07 | 450.41 | 231.05 | 1986.33 | 9617.21 | 13264.07 | 16771.84 | 19754.38 | 19947.79 | 20640.31 | 30430.57 |
| **2018** |  |  |  |  |  |  |  |  |  |  |  |
| Benzodiazepines and z-drugs | 470.73 | 375.01 | 283.28 | 1230.63 | 7546.52 | 10860.33 | 12795.31 | 16029.52 | 16676.68 | 15982.22 | 21614.65 |
| Long-acting benzodiazepines | 467.94 | 367.11 | 261.78 | 619.94 | 3622.24 | 4927.53 | 5136.47 | 5323.48 | 4523.90 | 3766.43 | 3778.53 |
| Clonazepam | 238.57 | 117.86 | 137.34 | 172.85 | 562.38 | 552.54 | 632.30 | 779.30 | 543.58 | 546.97 | 527.85 |
| Diazepam | 17.96 | 35.93 | 56.22 | 299.78 | 2414.57 | 3123.49 | 2718.61 | 2450.58 | 1949.40 | 1627.41 | 1750.92 |
| Chloridiazepoxide | 0.24 | 0.14 | 0.00 | 16.19 | 92.94 | 151.89 | 182.37 | 225.20 | 196.95 | 115.42 | 123.83 |
| Potassium Clorazepate | 0.00 | 0.00 | 0.00 | 0.00 | 0.20 | 0.24 | 1.20 | 7.59 | 6.32 | 2.46 | 1.99 |
| Lorazepam | 0.28 | 0.92 | 5.69 | 37.70 | 102.91 | 121.36 | 164.94 | 232.13 | 260.89 | 244.07 | 347.14 |
| Bromazepam | 0.35 | 0.06 | 1.63 | 16.44 | 65.84 | 154.32 | 252.17 | 576.04 | 888.68 | 930.30 | 993.80 |
| Clobazam | 209.17 | 210.01 | 63.36 | 65.33 | 113.66 | 90.69 | 124.13 | 120.16 | 121.15 | 80.52 | 77.55 |
| Prazepam | 0.00 | 0.00 | 0.02 | 1.09 | 8.27 | 19.01 | 38.58 | 91.35 | 138.51 | 154.29 | 154.27 |
| Alprazolam | 0.32 | 1.13 | 4.92 | 208.58 | 1116.42 | 1393.76 | 1525.42 | 1919.05 | 2008.89 | 1916.45 | 2433.64 |
| Flurazepam | 0.05 | 0.02 | 0.04 | 60.73 | 418.27 | 977.65 | 1406.44 | 1600.06 | 1481.74 | 1153.25 | 942.50 |
| Nitrazepam | 1.95 | 3.14 | 4.79 | 3.98 | 11.95 | 12.02 | 32.82 | 49.23 | 86.25 | 86.10 | 199.62 |
| Triazolam | 0.05 | 0.04 | 0.56 | 11.07 | 160.58 | 231.28 | 266.68 | 398.70 | 536.22 | 511.28 | 895.52 |
| Lormetazepam | 0.00 | 1.30 | 0.13 | 6.56 | 16.71 | 23.01 | 76.17 | 179.99 | 272.03 | 344.71 | 748.67 |
| Temazepam | 0.00 | 0.03 | 1.01 | 9.77 | 58.25 | 133.11 | 219.04 | 446.44 | 609.25 | 694.52 | 1317.28 |
| Zopiclone | 1.44 | 3.41 | 5.66 | 213.05 | 1734.65 | 2690.81 | 3257.61 | 4199.40 | 4251.90 | 4173.64 | 6324.19 |
| Zolpidem | 0.34 | 1.01 | 1.89 | 107.48 | 668.04 | 1184.32 | 1896.27 | 2753.60 | 3323.58 | 3400.20 | 4774.32 |
| Antidepressants^1^ | 0.57 | 0.80 | 2.19 | 245.18 | 967.72 | 1208.37 | 1523.10 | 1847.26 | 1757.11 | 1897.90 | 4019.08 |
| Antihistamines^2^ | 0.74 | 0.66 | 6.50 | 91.27 | 237.49 | 295.53 | 329.71 | 320.96 | 208.74 | 152.07 | 145.24 |
| Antipsychotics^3^ | 1.18 | 5.81 | 45.53 | 635.81 | 1709.15 | 1806.11 | 2056.92 | 2232.45 | 1996.16 | 1960.52 | 4362.44 |
| Sedatives | 473.22 | 382.28 | 337.49 | 2202.88 | 10460.87 | 14170.35 | 16705.05 | 20430.19 | 20638.69 | 19992.71 | 30141.42 |
| **2019** |  |  |  |  |  |  |  |  |  |  |  |
| Benzodiazepines and z-drugs | 243.03 | 493.81 | 306.17 | 1178.18 | 7353.66 | 10502.34 | 12606.40 | 15763.57 | 16203.93 | 15878.59 | 21651.19 |
| Long-acting benzodiazepines | 241.21 | 488.21 | 289.25 | 591.28 | 3486.00 | 4858.09 | 5072.58 | 5304.60 | 4338.41 | 3669.79 | 3858.90 |
| Clonazepam | 112.65 | 157.37 | 114.03 | 138.66 | 527.39 | 628.81 | 611.31 | 778.20 | 523.16 | 552.21 | 547.62 |
| Diazepam | 21.08 | 23.39 | 74.00 | 308.15 | 2363.12 | 3060.32 | 2783.84 | 2496.23 | 1917.32 | 1599.72 | 1829.93 |
| Chloridiazepoxide | 0.00 | 0.17 | 0.31 | 21.12 | 100.66 | 181.38 | 182.42 | 224.51 | 185.98 | 96.37 | 115.52 |
| Potassium Clorazepate | 0.00 | 0.00 | 0.00 | 0.00 | 0.30 | 0.05 | 1.75 | 7.21 | 9.12 | 2.47 | 1.35 |
| Lorazepam | 0.00 | 0.73 | 2.33 | 50.83 | 112.90 | 125.55 | 163.42 | 218.69 | 263.44 | 248.88 | 360.77 |
| Bromazepam | 0.14 | 0.04 | 0.57 | 18.02 | 68.41 | 145.45 | 257.18 | 520.97 | 815.04 | 920.20 | 985.69 |
| Clobazam | 105.50 | 302.27 | 93.84 | 68.48 | 125.05 | 124.54 | 107.90 | 125.37 | 109.10 | 87.60 | 75.21 |
| Prazepam | 0.00 | 0.00 | 0.04 | 1.64 | 5.93 | 16.23 | 31.86 | 82.43 | 144.49 | 147.89 | 146.16 |
| Alprazolam | 1.14 | 1.24 | 5.64 | 191.11 | 1101.00 | 1328.22 | 1515.86 | 1893.16 | 2015.83 | 1935.29 | 2474.41 |
| Flurazepam | 0.00 | 0.11 | 1.93 | 48.18 | 356.20 | 834.38 | 1321.24 | 1542.07 | 1375.63 | 1099.92 | 946.55 |
| Nitrazepam | 1.98 | 4.90 | 5.11 | 5.05 | 7.35 | 12.39 | 32.27 | 48.57 | 73.60 | 83.62 | 196.56 |
| Triazolam | 0.09 | 0.02 | 0.21 | 8.09 | 138.59 | 205.74 | 271.63 | 413.01 | 482.91 | 519.17 | 886.79 |
| Lormetazepam | 0.00 | 1.05 | 0.00 | 1.61 | 10.64 | 9.31 | 37.93 | 92.78 | 137.69 | 192.51 | 414.57 |
| Temazepam | 0.02 | 0.00 | 1.25 | 10.87 | 58.81 | 106.63 | 188.58 | 417.74 | 555.49 | 666.18 | 1259.13 |
| Zopiclone | 0.34 | 1.49 | 4.24 | 202.24 | 1699.11 | 2591.59 | 3282.41 | 4174.59 | 4305.58 | 4228.92 | 6459.29 |
| Zolpidem | 0.09 | 1.03 | 2.68 | 104.14 | 678.19 | 1131.77 | 1816.81 | 2728.02 | 3289.54 | 3497.63 | 4951.63 |
| Antidepressants^1^ | 0.10 | 0.72 | 1.66 | 255.80 | 1081.83 | 1220.99 | 1589.18 | 1926.10 | 1934.23 | 1982.87 | 4208.37 |
| Antihistamines^2^ | 0.62 | 1.31 | 7.55 | 142.04 | 328.80 | 379.27 | 420.44 | 420.57 | 252.66 | 199.40 | 189.98 |
| Antipsychotics^3^ | 0.24 | 5.76 | 50.85 | 782.02 | 2018.09 | 2046.69 | 2274.03 | 2387.40 | 2065.94 | 2083.26 | 4600.65 |
| Sedatives | 244.00 | 501.61 | 366.23 | 2358.05 | 10782.38 | 14149.29 | 16890.05 | 20497.64 | 20456.77 | 20144.12 | 30650.18 |
| **2020** |  |  |  |  |  |  |  |  |  |  |  |
| Benzodiazepines and z-drugs | 403.11 | 575.67 | 274.50 | 1059.98 | 6236.04 | 10047.93 | 12141.28 | 15690.00 | 15405.46 | 15356.67 | 20939.10 |
| Long-acting benzodiazepines | 401.14 | 571.77 | 257.89 | 552.32 | 2854.91 | 4651.62 | 4896.12 | 5608.99 | 4020.24 | 3547.25 | 3724.43 |
| Clonazepam | 103.74 | 149.12 | 112.53 | 126.24 | 441.98 | 603.68 | 590.79 | 756.10 | 570.06 | 514.04 | 556.31 |
| Diazepam | 45.37 | 14.97 | 57.74 | 267.70 | 1913.98 | 3017.21 | 2762.43 | 2980.20 | 1769.97 | 1595.35 | 1784.94 |
| Chloridiazepoxide | 0.18 | 0.28 | 0.24 | 15.29 | 86.60 | 159.33 | 187.43 | 219.73 | 142.29 | 94.83 | 101.98 |
| Potassium Clorazepate | 0.00 | 0.00 | 0.00 | 0.00 | 0.73 | 0.16 | 0.37 | 6.66 | 9.06 | 2.45 | 1.54 |
| Lorazepam | 0.05 | 1.41 | 3.41 | 50.77 | 114.72 | 127.06 | 165.35 | 220.42 | 247.73 | 259.29 | 358.44 |
| Bromazepam | 0.11 | 0.05 | 0.45 | 13.90 | 69.71 | 125.34 | 228.12 | 476.31 | 769.49 | 877.61 | 966.47 |
| Clobazam | 250.42 | 403.34 | 79.35 | 97.08 | 114.63 | 125.13 | 103.43 | 126.62 | 100.96 | 83.72 | 75.67 |
| Prazepam | 0.00 | 0.02 | 0.03 | 0.95 | 4.75 | 12.08 | 31.08 | 66.51 | 112.46 | 128.40 | 124.66 |
| Alprazolam | 0.65 | 0.95 | 4.35 | 153.99 | 1026.12 | 1237.57 | 1464.95 | 1797.41 | 1978.74 | 1909.29 | 2427.69 |
| Flurazepam | 0.00 | 0.02 | 1.41 | 39.66 | 284.37 | 718.76 | 1192.92 | 1407.85 | 1250.93 | 1048.76 | 898.99 |
| Nitrazepam | 1.43 | 4.02 | 6.59 | 5.40 | 7.88 | 15.26 | 27.67 | 45.33 | 64.49 | 79.70 | 180.34 |
| Triazolam | 0.00 | 0.00 | 0.29 | 7.97 | 121.06 | 199.23 | 277.33 | 394.47 | 440.42 | 540.40 | 855.02 |
| Lormetazepam | 0.00 | 0.00 | 0.30 | 0.06 | 0.43 | 0.33 | 2.52 | 8.01 | 13.96 | 14.00 | 31.28 |
| Temazepam | 0.00 | 0.05 | 1.31 | 5.99 | 62.94 | 95.42 | 174.51 | 374.39 | 490.65 | 601.09 | 1175.09 |
| Zopiclone | 0.61 | 1.04 | 3.96 | 191.01 | 1424.98 | 2499.21 | 3167.14 | 4078.43 | 4194.29 | 4097.57 | 6416.67 |
| Zolpidem | 0.56 | 0.40 | 2.56 | 83.96 | 561.18 | 1112.16 | 1765.24 | 2731.58 | 3249.94 | 3510.16 | 4984.01 |
| Antidepressants^1^ | 0.20 | 0.93 | 1.93 | 238.04 | 1155.82 | 1328.24 | 1675.63 | 2140.61 | 2028.89 | 2124.89 | 4452.06 |
| Antihistamines^2^ | 0.03 | 1.41 | 5.76 | 139.05 | 417.17 | 458.78 | 525.92 | 595.14 | 346.51 | 224.10 | 244.40 |
| Antipsychotics^3^ | 0.08 | 6.30 | 49.16 | 816.42 | 2256.01 | 2325.22 | 2500.46 | 2539.81 | 2109.05 | 2188.95 | 4789.57 |
| Sedatives | 403.41 | 584.31 | 331.36 | 2253.48 | 10065.04 | 14160.17 | 16843.29 | 20965.55 | 19889.92 | 19894.60 | 30425.13 |
| **2021** |  |  |  |  |  |  |  |  |  |  |  |
| Benzodiazepines and z-drugs | 542.30 | 753.99 | 363.64 | 1219.69 | 6816.87 | 10765.25 | 12812.44 | 16912.26 | 15801.20 | 15562.91 | 20907.93 |
| Long-acting benzodiazepines | 541.02 | 750.05 | 348.17 | 711.30 | 3164.59 | 5012.91 | 5280.54 | 6413.70 | 4202.35 | 3509.05 | 3668.34 |
| Clonazepam | 245.81 | 209.47 | 121.07 | 155.49 | 485.86 | 625.02 | 665.01 | 808.45 | 633.81 | 521.58 | 619.52 |
| Diazepam | 37.33 | 26.33 | 42.67 | 328.78 | 2178.92 | 3349.11 | 3052.73 | 3796.35 | 1920.66 | 1644.41 | 1801.60 |
| Chloridiazepoxide | 0.04 | 0.19 | 0.13 | 23.28 | 92.28 | 191.77 | 206.11 | 248.02 | 133.44 | 89.60 | 96.96 |
| Potassium Clorazepate | 0.00 | 0.00 | 0.00 | 0.00 | 2.30 | 0.00 | 0.29 | 3.48 | 4.10 | 6.20 | 1.89 |
| Lorazepam | 0.00 | 1.78 | 4.40 | 65.36 | 144.34 | 143.89 | 171.03 | 236.34 | 280.29 | 251.93 | 366.52 |
| Bromazepam | 0.00 | 0.04 | 1.01 | 11.75 | 78.22 | 117.61 | 208.42 | 458.36 | 744.30 | 820.59 | 945.15 |
| Clobazam | 255.86 | 510.79 | 182.25 | 156.84 | 137.97 | 80.89 | 141.43 | 149.58 | 92.02 | 85.95 | 77.68 |
| Prazepam | 0.00 | 0.02 | 0.00 | 2.19 | 6.11 | 12.67 | 31.15 | 56.78 | 107.83 | 129.70 | 127.34 |
| Alprazolam | 0.19 | 0.90 | 4.01 | 142.41 | 1077.08 | 1356.48 | 1563.72 | 1879.60 | 1965.81 | 1960.80 | 2448.78 |
| Flurazepam | 0.00 | 0.41 | 0.00 | 41.88 | 257.59 | 749.96 | 1173.57 | 1335.40 | 1290.07 | 1000.30 | 875.05 |
| Nitrazepam | 1.97 | 2.85 | 2.05 | 2.84 | 3.55 | 3.49 | 10.26 | 15.63 | 20.41 | 31.30 | 68.30 |
| Triazolam | 0.00 | 0.00 | 0.06 | 10.95 | 117.13 | 212.03 | 270.34 | 389.17 | 460.26 | 543.20 | 847.78 |
| Lormetazepam | 0.00 | 0.00 | 0.00 | 0.00 | 0.00 | 0.05 | 0.15 | 0.27 | 1.38 | 2.03 | 2.88 |
| Temazepam | 0.00 | 0.00 | 0.92 | 5.83 | 57.53 | 90.85 | 174.97 | 355.84 | 492.76 | 585.03 | 1120.06 |
| Zopiclone | 0.80 | 0.40 | 3.26 | 178.64 | 1551.75 | 2677.29 | 3323.36 | 4376.42 | 4301.17 | 4262.10 | 6463.14 |
| Zolpidem | 0.31 | 0.81 | 1.82 | 93.45 | 626.23 | 1154.14 | 1819.92 | 2802.50 | 3352.88 | 3628.19 | 5045.29 |
| Antidepressants^1^ | 0.09 | 2.65 | 0.58 | 280.32 | 1355.74 | 1571.69 | 1885.31 | 2483.63 | 2262.62 | 2325.82 | 5012.18 |
| Antihistamines^2^ | 0.18 | 1.57 | 10.72 | 196.66 | 575.42 | 613.87 | 677.11 | 931.91 | 452.24 | 278.48 | 301.16 |
| Antipsychotics^3^ | 0.00 | 6.80 | 45.94 | 944.96 | 2646.91 | 2728.45 | 2807.85 | 2852.15 | 2470.73 | 2278.21 | 4968.40 |
| Sedatives | 542.58 | 765.01 | 420.88 | 2641.62 | 11394.95 | 15679.25 | 18182.71 | 23179.95 | 20986.79 | 20445.41 | 31189.68 |
| **2022** |  |  |  |  |  |  |  |  |  |  |  |
| Benzodiazepines and z-drugs | 439.38 | 650.94 | 551.11 | 1324.83 | 6327.28 | 10096.19 | 12335.88 | 15300.39 | 14942.72 | 14850.37 | 19700.71 |
| Long-acting benzodiazepines | 438.33 | 646.53 | 535.44 | 851.50 | 3083.26 | 4646.97 | 5130.70 | 5538.08 | 3963.85 | 3355.65 | 3414.49 |
| Clonazepam | 113.93 | 155.96 | 195.31 | 184.59 | 426.57 | 593.08 | 660.71 | 729.03 | 704.98 | 489.47 | 596.91 |
| Diazepam | 35.73 | 26.92 | 39.73 | 330.02 | 2156.98 | 3198.27 | 3075.02 | 3177.10 | 1879.32 | 1646.48 | 1757.91 |
| Chloridiazepoxide | 0.00 | 0.08 | 0.07 | 25.77 | 95.06 | 159.83 | 170.59 | 199.84 | 120.56 | 96.67 | 80.82 |
| Potassium Clorazepate | 0.00 | 0.00 | 0.00 | 0.00 | 0.44 | 0.40 | 0.00 | 4.68 | 4.39 | 3.46 | 2.85 |
| Lorazepam | 0.24 | 2.61 | 6.00 | 67.53 | 133.25 | 160.47 | 185.74 | 209.74 | 263.64 | 257.11 | 350.82 |
| Bromazepam | 0.10 | 0.00 | 1.02 | 10.75 | 70.63 | 123.55 | 183.50 | 415.31 | 681.95 | 758.16 | 888.50 |
| Clobazam | 288.29 | 462.34 | 299.89 | 270.13 | 179.28 | 81.36 | 153.51 | 154.10 | 88.06 | 79.21 | 73.16 |
| Prazepam | 0.00 | 0.00 | 0.00 | 0.89 | 2.49 | 5.79 | 18.78 | 28.23 | 59.48 | 75.27 | 81.15 |
| Alprazolam | 0.25 | 0.57 | 3.53 | 123.27 | 1013.68 | 1332.13 | 1499.65 | 1785.30 | 1838.79 | 1888.89 | 2293.01 |
| Flurazepam | 0.00 | 0.07 | 0.37 | 39.71 | 222.04 | 607.58 | 1050.71 | 1243.77 | 1106.10 | 963.92 | 813.84 |
| Nitrazepam | 0.38 | 1.15 | 0.06 | 0.39 | 0.39 | 0.67 | 1.37 | 1.33 | 0.96 | 1.17 | 7.87 |
| Triazolam | 0.00 | 0.00 | 0.53 | 10.15 | 105.22 | 199.99 | 262.08 | 364.68 | 393.08 | 518.69 | 793.72 |
| Lormetazepam | 0.00 | 0.00 | 0.00 | 0.00 | 0.06 | 0.00 | 0.06 | 0.13 | 0.00 | 1.00 | 0.86 |
| Temazepam | 0.00 | 0.02 | 0.06 | 6.50 | 42.79 | 77.29 | 147.34 | 306.01 | 429.35 | 531.79 | 1000.15 |
| Zopiclone | 0.17 | 0.86 | 2.51 | 168.67 | 1367.78 | 2495.78 | 3184.80 | 4061.30 | 4145.95 | 4073.58 | 6153.49 |
| Zolpidem | 0.28 | 0.36 | 2.01 | 86.45 | 510.61 | 1060.01 | 1742.02 | 2619.84 | 3226.11 | 3465.51 | 4805.66 |
| Antidepressants^1^ | 0.00 | 3.39 | 12.11 | 250.29 | 1258.43 | 1518.53 | 1884.69 | 2441.90 | 2263.95 | 2398.08 | 5294.87 |
| Antihistamines^2^ | 0.04 | 1.83 | 12.47 | 165.51 | 585.70 | 648.20 | 754.80 | 855.76 | 517.09 | 328.08 | 349.56 |
| Antipsychotics^3^ | 0.00 | 5.71 | 36.94 | 809.04 | 2587.17 | 2702.40 | 2866.98 | 2883.64 | 2462.42 | 2190.75 | 4779.06 |
| Sedatives | 439.41 | 661.88 | 612.64 | 2549.67 | 10758.58 | 14965.32 | 17842.35 | 21481.70 | 20186.18 | 19767.29 | 30124.21 |

**Rate of cost per 1,000 GMS population by sex**

|  | **Females** | **Males** |
| --- | --- | --- |
| **2014** |  |  |
| Benzodiazepines and z-drugs | 12093.00 | 8152.77 |
| Long-acting benzodiazepines | 3092.00 | 2811.62 |
| Clonazepam | 401.73 | 375.63 |
| Diazepam | 1421.17 | 1310.30 |
| Chloridiazepoxide | 94.62 | 137.93 |
| Potassium Clorazepate | 1.92 | 1.12 |
| Lorazepam | 158.89 | 123.51 |
| Bromazepam | 554.39 | 206.52 |
| Clobazam | 138.64 | 138.87 |
| Prazepam | 86.38 | 39.95 |
| Alprazolam | 1434.34 | 823.43 |
| Flurazepam | 856.25 | 764.95 |
| Nitrazepam | 91.29 | 42.86 |
| Triazolam | 368.50 | 219.50 |
| Lormetazepam | 302.65 | 121.85 |
| Temazepam | 566.86 | 316.43 |
| Zopiclone | 3446.65 | 2401.88 |
| Zolpidem | 2137.61 | 1108.37 |
| Antidepressants^1^ | 1426.07 | 917.21 |
| Antihistamines^2^ | 69.99 | 54.97 |
| Antipsychotics^3^ | 2246.24 | 1689.26 |
| Sedatives | 15835.29 | 10814.20 |
| **2015** |  |  |
| Benzodiazepines and z-drugs | 11816.87 | 8050.79 |
| Long-acting benzodiazepines | 3218.92 | 2977.65 |
| Clonazepam | 406.04 | 386.09 |
| Diazepam | 1566.58 | 1457.34 |
| Chloridiazepoxide | 90.16 | 135.22 |
| Potassium Clorazepate | 1.91 | 1.25 |
| Lorazepam | 161.65 | 123.15 |
| Bromazepam | 548.25 | 203.90 |
| Clobazam | 111.21 | 132.07 |
| Prazepam | 83.42 | 38.84 |
| Alprazolam | 1466.44 | 832.46 |
| Flurazepam | 873.46 | 787.76 |
| Nitrazepam | 86.15 | 39.09 |
| Triazolam | 381.61 | 226.09 |
| Lormetazepam | 286.66 | 114.45 |
| Temazepam | 537.18 | 301.45 |
| Zopiclone | 3000.21 | 2130.91 |
| Zolpidem | 2181.53 | 1118.68 |
| Antidepressants^1^ | 1549.16 | 1001.91 |
| Antihistamines^2^ | 91.61 | 70.52 |
| Antipsychotics^3^ | 2228.82 | 1689.54 |
| Sedatives | 15686.46 | 10812.75 |
| **2016** |  |  |
| Benzodiazepines and z-drugs | 12037.91 | 8371.31 |
| Long-acting benzodiazepines | 3264.66 | 3116.26 |
| Clonazepam | 420.88 | 424.91 |
| Diazepam | 1606.87 | 1542.96 |
| Chloridiazepoxide | 91.10 | 141.32 |
| Potassium Clorazepate | 1.76 | 0.91 |
| Lorazepam | 165.73 | 126.15 |
| Bromazepam | 541.46 | 202.32 |
| Clobazam | 121.63 | 128.58 |
| Prazepam | 82.63 | 37.15 |
| Alprazolam | 1493.86 | 862.90 |
| Flurazepam | 857.34 | 802.64 |
| Nitrazepam | 82.45 | 37.80 |
| Triazolam | 385.41 | 233.72 |
| Lormetazepam | 280.98 | 113.01 |
| Temazepam | 518.82 | 296.49 |
| Zopiclone | 3087.00 | 2234.84 |
| Zolpidem | 2277.43 | 1171.55 |
| Antidepressants^1^ | 1542.21 | 1020.19 |
| Antihistamines^2^ | 120.92 | 97.59 |
| Antipsychotics^3^ | 2262.79 | 1727.46 |
| Sedatives | 15963.83 | 11216.54 |
| **2017** |  |  |
| Benzodiazepines and z-drugs | 12200.85 | 8496.08 |
| Long-acting benzodiazepines | 3268.23 | 3171.13 |
| Clonazepam | 448.65 | 457.06 |
| Diazepam | 1630.16 | 1588.06 |
| Chloridiazepoxide | 87.98 | 136.74 |
| Potassium Clorazepate | 2.02 | 1.29 |
| Lorazepam | 165.90 | 132.35 |
| Bromazepam | 537.98 | 202.72 |
| Clobazam | 105.26 | 119.96 |
| Prazepam | 81.63 | 35.89 |
| Alprazolam | 1529.03 | 885.42 |
| Flurazepam | 834.12 | 795.85 |
| Nitrazepam | 78.42 | 36.28 |
| Triazolam | 392.71 | 239.63 |
| Lormetazepam | 275.04 | 105.22 |
| Temazepam | 506.47 | 283.47 |
| Zopiclone | 3175.65 | 2267.59 |
| Zolpidem | 2347.06 | 1207.35 |
| Antidepressants^1^ | 1592.29 | 1043.10 |
| Antihistamines^2^ | 159.16 | 123.76 |
| Antipsychotics^3^ | 1843.93 | 1402.48 |
| Sedatives | 15796.23 | 11065.42 |
| **2018** |  |  |
| Benzodiazepines and z-drugs | 12420.74 | 8649.44 |
| Long-acting benzodiazepines | 3289.61 | 3233.82 |
| Clonazepam | 458.41 | 471.46 |
| Diazepam | 1670.71 | 1639.32 |
| Chloridiazepoxide | 82.28 | 143.49 |
| Potassium Clorazepate | 2.25 | 1.45 |
| Lorazepam | 169.34 | 140.61 |
| Bromazepam | 548.20 | 203.84 |
| Clobazam | 96.56 | 132.51 |
| Prazepam | 80.57 | 37.06 |
| Alprazolam | 1593.43 | 912.57 |
| Flurazepam | 826.84 | 775.02 |
| Nitrazepam | 72.00 | 33.51 |
| Triazolam | 388.42 | 238.66 |
| Lormetazepam | 264.63 | 98.71 |
| Temazepam | 482.40 | 263.93 |
| Zopiclone | 3247.88 | 2305.07 |
| Zolpidem | 2435.86 | 1251.91 |
| Antidepressants^1^ | 1747.70 | 1131.86 |
| Antihistamines^2^ | 201.85 | 152.61 |
| Antipsychotics^3^ | 2021.37 | 1540.95 |
| Sedatives | 16391.66 | 11474.86 |
| **2019** |  |  |
| Benzodiazepines and z-drugs | 12454.01 | 8651.71 |
| Long-acting benzodiazepines | 3277.83 | 3234.55 |
| Clonazepam | 460.20 | 474.39 |
| Diazepam | 1686.25 | 1664.00 |
| Chloridiazepoxide | 81.62 | 148.48 |
| Potassium Clorazepate | 2.33 | 1.60 |
| Lorazepam | 172.23 | 149.00 |
| Bromazepam | 545.18 | 197.12 |
| Clobazam | 105.53 | 142.26 |
| Prazepam | 76.91 | 36.36 |
| Alprazolam | 1619.41 | 905.61 |
| Flurazepam | 794.33 | 733.02 |
| Nitrazepam | 70.66 | 34.44 |
| Triazolam | 392.23 | 233.56 |
| Lormetazepam | 149.51 | 52.09 |
| Temazepam | 471.43 | 244.79 |
| Zopiclone | 3311.32 | 2362.38 |
| Zolpidem | 2514.88 | 1272.60 |
| Antidepressants^1^ | 1866.80 | 1208.42 |
| Antihistamines^2^ | 263.16 | 202.03 |
| Antipsychotics^3^ | 2202.89 | 1715.98 |
| Sedatives | 16786.87 | 11778.14 |
| **2020** |  |  |
| Benzodiazepines and z-drugs | 11957.64 | 8336.54 |
| Long-acting benzodiazepines | 3140.76 | 3173.63 |
| Clonazepam | 449.63 | 452.06 |
| Diazepam | 1632.50 | 1707.63 |
| Chloridiazepoxide | 74.29 | 137.78 |
| Potassium Clorazepate | 2.26 | 1.31 |
| Lorazepam | 176.03 | 145.21 |
| Bromazepam | 513.66 | 187.97 |
| Clobazam | 120.90 | 158.72 |
| Prazepam | 64.38 | 30.50 |
| Alprazolam | 1573.20 | 852.42 |
| Flurazepam | 733.66 | 650.93 |
| Nitrazepam | 63.15 | 34.71 |
| Triazolam | 375.34 | 232.01 |
| Lormetazepam | 11.37 | 4.20 |
| Temazepam | 431.93 | 225.59 |
| Zopiclone | 3224.46 | 2280.53 |
| Zolpidem | 2510.89 | 1234.97 |
| Antidepressants^1^ | 1992.04 | 1288.37 |
| Antihistamines^2^ | 325.54 | 265.94 |
| Antipsychotics^3^ | 2363.76 | 1828.12 |
| Sedatives | 16638.98 | 11718.97 |
| **2021** |  |  |
| Benzodiazepines and z-drugs | 12512.14 | 9082.47 |
| Long-acting benzodiazepines | 3301.13 | 3602.08 |
| Clonazepam | 491.58 | 523.43 |
| Diazepam | 1774.58 | 2028.96 |
| Chloridiazepoxide | 79.12 | 153.57 |
| Potassium Clorazepate | 2.20 | 0.97 |
| Lorazepam | 190.24 | 159.05 |
| Bromazepam | 507.56 | 187.23 |
| Clobazam | 139.36 | 191.16 |
| Prazepam | 65.20 | 31.45 |
| Alprazolam | 1658.84 | 900.28 |
| Flurazepam | 724.80 | 659.16 |
| Nitrazepam | 24.30 | 13.38 |
| Triazolam | 384.41 | 241.07 |
| Lormetazepam | 1.05 | 0.44 |
| Temazepam | 426.61 | 228.84 |
| Zopiclone | 3399.07 | 2458.87 |
| Zolpidem | 2643.22 | 1304.61 |
| Antidepressants^1^ | 2294.09 | 1528.30 |
| Antihistamines^2^ | 432.15 | 383.86 |
| Antipsychotics^3^ | 2662.26 | 2027.15 |
| Sedatives | 17900.64 | 13021.78 |
| **2022** |  |  |
| Benzodiazepines and z-drugs | 11817.75 | 8577.70 |
| Long-acting benzodiazepines | 3125.91 | 3351.31 |
| Clonazepam | 469.48 | 503.42 |
| Diazepam | 1736.99 | 1882.81 |
| Chloridiazepoxide | 68.69 | 133.11 |
| Potassium Clorazepate | 2.15 | 0.99 |
| Lorazepam | 181.21 | 164.62 |
| Bromazepam | 468.59 | 178.70 |
| Clobazam | 160.02 | 201.24 |
| Prazepam | 38.42 | 18.34 |
| Alprazolam | 1568.92 | 870.56 |
| Flurazepam | 647.62 | 609.97 |
| Nitrazepam | 2.55 | 1.44 |
| Triazolam | 359.62 | 228.03 |
| Lormetazepam | 0.41 | 0.05 |
| Temazepam | 377.79 | 204.39 |
| Zopiclone | 3229.97 | 2335.70 |
| Zolpidem | 2505.32 | 1244.34 |
| Antidepressants^1^ | 2343.06 | 1567.19 |
| Antihistamines^2^ | 468.45 | 384.38 |
| Antipsychotics^3^ | 2624.16 | 1995.24 |
| Sedatives | 17253.42 | 12524.51 |

**Rate of DDDs per 1,000 GMS populations by age group**

|  | **<5** | **5-11** | **12-15** | **16-24** | **25-34** | **35-44** | **45-54** | **55-64** | **65-69** | **70-74** | **75+** |
| --- | --- | --- | --- | --- | --- | --- | --- | --- | --- | --- | --- |
| **2014** |  |  |  |  |  |  |  |  |  |  |  |
| Benzodiazepines and z-drugs | 228.44 | 236.62 | 325.59 | 3021.20 | 17257.78 | 29897.63 | 41142.81 | 57447.55 | 61257.87 | 59957.60 | 73889.32 |
| Long-acting benzodiazepines | 214.02 | 201.44 | 240.57 | 1226.56 | 7056.56 | 11971.28 | 13128.01 | 15405.40 | 14140.79 | 11816.23 | 10876.05 |
| Clonazepam | 20.60 | 21.72 | 20.04 | 55.77 | 178.04 | 258.64 | 328.47 | 307.66 | 235.00 | 184.40 | 167.45 |
| Diazepam | 18.91 | 43.09 | 49.23 | 822.15 | 5417.58 | 8731.51 | 8439.10 | 9226.53 | 8112.84 | 6543.04 | 5321.11 |
| Chloridiazepoxide | 0.01 | 0.49 | 8.61 | 20.08 | 141.48 | 214.65 | 397.10 | 483.97 | 331.60 | 303.88 | 284.95 |
| Potassium Clorazepate | 0.00 | 0.00 | 0.00 | 0.76 | 0.20 | 0.59 | 7.49 | 5.94 | 3.08 | 2.86 | 1.98 |
| Lorazepam | 0.18 | 4.10 | 10.00 | 72.37 | 189.28 | 242.45 | 441.48 | 809.96 | 1120.57 | 949.04 | 1301.25 |
| Bromazepam | 0.00 | 0.05 | 1.17 | 18.86 | 89.97 | 193.52 | 514.60 | 1225.85 | 1669.96 | 1476.75 | 1342.72 |
| Clobazam | 120.23 | 116.43 | 122.86 | 185.67 | 314.47 | 354.11 | 423.90 | 439.36 | 391.95 | 311.13 | 230.10 |
| Prazepam | 0.04 | 0.10 | 0.03 | 4.64 | 19.35 | 45.15 | 129.30 | 337.60 | 414.18 | 402.99 | 321.29 |
| Alprazolam | 5.21 | 5.74 | 42.33 | 523.26 | 2707.85 | 3683.86 | 4963.92 | 6527.07 | 6424.72 | 5858.95 | 6237.48 |
| Flurazepam | 0.34 | 1.17 | 15.38 | 90.39 | 905.48 | 2223.72 | 3090.54 | 3993.98 | 3779.13 | 2960.94 | 2574.34 |
| Nitrazepam | 53.89 | 18.43 | 24.42 | 47.09 | 79.96 | 142.92 | 312.10 | 610.34 | 873.02 | 1106.99 | 1974.83 |
| Triazolam | 0.09 | 2.20 | 0.10 | 83.97 | 624.28 | 871.61 | 1474.27 | 2260.74 | 2935.93 | 3043.75 | 4397.31 |
| Lormetazepam | 0.25 | 1.25 | 5.68 | 12.35 | 53.53 | 155.55 | 484.19 | 1019.21 | 1684.00 | 2412.82 | 3827.45 |
| Temazepam | 1.21 | 0.30 | 1.39 | 26.26 | 155.88 | 408.24 | 1054.86 | 2015.50 | 3163.99 | 3183.72 | 5896.01 |
| Zopiclone | 1.98 | 14.39 | 15.07 | 677.73 | 4477.64 | 8306.66 | 11760.52 | 16761.90 | 17141.00 | 17745.59 | 23346.25 |
| Zolpidem | 5.49 | 7.15 | 9.29 | 379.86 | 1902.79 | 4064.45 | 7320.97 | 11421.93 | 12976.90 | 13470.75 | 16664.78 |
| Antidepressants^1^ | 1.09 | 1.36 | 2.06 | 188.97 | 622.15 | 952.35 | 1381.42 | 1638.43 | 1655.18 | 1833.61 | 3534.70 |
| Antihistamines^2^ | 0.00 | 6.85 | 20.83 | 175.97 | 437.45 | 595.34 | 779.01 | 824.46 | 502.82 | 398.62 | 403.02 |
| Antipsychotics^3^ | 0.06 | 1.88 | 16.48 | 138.06 | 339.47 | 454.53 | 611.35 | 668.81 | 658.30 | 771.88 | 1525.61 |
| Sedatives | 229.59 | 246.71 | 364.96 | 3524.20 | 18656.84 | 31899.85 | 43914.59 | 60579.25 | 64074.17 | 62961.71 | 79352.65 |
| **2015** |  |  |  |  |  |  |  |  |  |  |  |
| Benzodiazepines and z-drugs | 150.32 | 207.89 | 382.37 | 2972.32 | 17061.99 | 28524.19 | 41386.21 | 55815.74 | 59631.51 | 57589.04 | 69875.13 |
| Long-acting benzodiazepines | 137.14 | 179.88 | 269.86 | 1215.67 | 6978.91 | 11290.04 | 13349.72 | 14600.52 | 13662.09 | 11371.59 | 10026.47 |
| Clonazepam | 8.77 | 16.22 | 16.32 | 60.83 | 189.20 | 248.26 | 333.02 | 316.59 | 243.26 | 180.22 | 161.26 |
| Diazepam | 9.69 | 49.43 | 96.66 | 823.93 | 5538.57 | 8391.40 | 8877.68 | 8886.21 | 8064.35 | 6317.51 | 5098.30 |
| Chloridiazepoxide | 0.09 | 0.15 | 8.61 | 15.10 | 128.86 | 234.49 | 337.59 | 413.97 | 351.90 | 290.43 | 259.53 |
| Potassium Clorazepate | 0.00 | 0.00 | 0.00 | 0.24 | 1.25 | 0.55 | 7.87 | 6.25 | 2.80 | 1.80 | 0.78 |
| Lorazepam | 0.18 | 1.90 | 17.39 | 87.90 | 207.15 | 244.51 | 434.65 | 770.01 | 1024.22 | 942.45 | 1173.81 |
| Bromazepam | 0.01 | 0.04 | 1.02 | 23.01 | 84.16 | 188.77 | 476.42 | 1109.26 | 1569.75 | 1423.77 | 1279.67 |
| Clobazam | 83.50 | 95.87 | 93.19 | 168.51 | 244.15 | 284.66 | 408.10 | 376.49 | 358.28 | 291.63 | 222.49 |
| Prazepam | 0.00 | 0.00 | 0.02 | 4.62 | 17.43 | 47.45 | 112.47 | 321.64 | 383.25 | 370.83 | 305.82 |
| Alprazolam | 3.70 | 7.37 | 41.18 | 448.54 | 2700.03 | 3526.65 | 4866.86 | 6335.78 | 6221.74 | 5737.43 | 6013.39 |
| Flurazepam | 0.10 | 1.57 | 21.42 | 95.12 | 788.11 | 1983.28 | 2998.85 | 3739.62 | 3457.61 | 2909.54 | 2289.09 |
| Nitrazepam | 34.99 | 16.63 | 33.63 | 47.33 | 71.33 | 99.95 | 274.14 | 539.76 | 800.63 | 1009.63 | 1689.20 |
| Triazolam | 0.28 | 0.27 | 2.82 | 65.65 | 668.73 | 885.53 | 1393.29 | 2289.15 | 3014.81 | 2824.72 | 4199.12 |
| Lormetazepam | 0.14 | 0.05 | 8.41 | 10.37 | 57.08 | 139.73 | 438.25 | 898.37 | 1447.75 | 2166.56 | 3488.67 |
| Temazepam | 0.00 | 1.34 | 0.87 | 30.31 | 147.61 | 361.19 | 942.82 | 1815.69 | 2819.85 | 2813.11 | 5210.78 |
| Zopiclone | 2.30 | 13.03 | 26.38 | 713.33 | 4361.12 | 8079.86 | 12057.35 | 16619.74 | 17099.37 | 17307.01 | 22553.62 |
| Zolpidem | 6.56 | 4.01 | 14.44 | 377.54 | 1857.20 | 3807.91 | 7426.85 | 11377.22 | 12771.94 | 13002.39 | 15929.60 |
| Antidepressants^1^ | 1.69 | 0.74 | 8.48 | 243.72 | 733.50 | 1079.86 | 1526.88 | 1841.12 | 1895.49 | 2062.99 | 3845.66 |
| Antihistamines^2^ | 0.00 | 6.29 | 33.28 | 227.55 | 565.58 | 763.10 | 973.08 | 921.27 | 651.37 | 420.27 | 456.64 |
| Antipsychotics^3^ | 0.06 | 3.03 | 19.02 | 183.92 | 399.39 | 503.47 | 661.44 | 727.92 | 745.14 | 757.99 | 1578.47 |
| Sedatives | 152.07 | 217.95 | 443.14 | 3627.51 | 18760.45 | 30870.63 | 44547.61 | 59306.05 | 62923.51 | 60830.29 | 75755.90 |
| **2016** |  |  |  |  |  |  |  |  |  |  |  |
| Benzodiazepines and z-drugs | 179.07 | 204.38 | 275.61 | 3125.08 | 18471.68 | 29623.17 | 41667.91 | 54360.53 | 58868.71 | 57187.21 | 68839.60 |
| Long-acting benzodiazepines | 141.84 | 166.38 | 181.33 | 1257.24 | 7535.52 | 11609.30 | 13567.64 | 13839.39 | 13256.93 | 10987.18 | 9592.79 |
| Clonazepam | 10.49 | 11.30 | 8.33 | 67.69 | 217.80 | 271.87 | 334.87 | 326.19 | 247.95 | 204.76 | 167.47 |
| Diazepam | 38.78 | 28.57 | 43.94 | 850.04 | 6044.75 | 8822.40 | 9161.08 | 8540.01 | 7912.81 | 6146.72 | 4952.81 |
| Chloridiazepoxide | 0.14 | 0.42 | 0.50 | 21.64 | 161.57 | 258.63 | 329.07 | 401.38 | 323.99 | 292.06 | 235.34 |
| Potassium Clorazepate | 0.00 | 0.00 | 0.00 | 0.00 | 1.48 | 0.13 | 6.99 | 2.45 | 8.49 | 0.13 | 1.56 |
| Lorazepam | 0.92 | 1.70 | 24.23 | 106.81 | 238.37 | 271.33 | 453.37 | 763.55 | 890.38 | 886.45 | 1170.82 |
| Bromazepam | 0.04 | 0.26 | 0.57 | 19.78 | 104.24 | 204.96 | 448.50 | 996.75 | 1490.46 | 1433.54 | 1231.70 |
| Clobazam | 65.04 | 100.32 | 84.47 | 175.93 | 243.01 | 259.27 | 369.19 | 342.03 | 359.31 | 251.45 | 215.00 |
| Prazepam | 0.00 | 0.00 | 0.02 | 4.25 | 16.51 | 45.60 | 107.10 | 288.60 | 341.01 | 367.27 | 302.68 |
| Alprazolam | 2.15 | 7.25 | 18.62 | 497.88 | 2945.42 | 3703.54 | 4931.62 | 6137.82 | 6135.02 | 5656.52 | 6006.72 |
| Flurazepam | 4.95 | 2.41 | 0.59 | 100.82 | 765.08 | 1861.45 | 2993.58 | 3492.09 | 3277.21 | 2826.08 | 2120.76 |
| Nitrazepam | 22.44 | 23.35 | 43.49 | 36.87 | 85.31 | 89.95 | 265.78 | 446.64 | 786.15 | 898.71 | 1597.15 |
| Triazolam | 2.14 | 1.69 | 2.56 | 69.97 | 678.54 | 904.07 | 1356.21 | 2199.04 | 2867.50 | 2758.71 | 4097.77 |
| Lormetazepam | 0.00 | 0.22 | 7.33 | 10.10 | 55.25 | 119.52 | 391.07 | 857.65 | 1407.24 | 2005.22 | 3349.07 |
| Temazepam | 0.00 | 0.31 | 0.31 | 26.65 | 132.31 | 366.60 | 848.74 | 1694.20 | 2541.36 | 2775.18 | 4754.73 |
| Zopiclone | 20.19 | 17.45 | 27.08 | 762.19 | 4752.29 | 8496.25 | 12305.38 | 16516.12 | 17063.08 | 17440.30 | 22396.62 |
| Zolpidem | 11.79 | 9.12 | 13.58 | 374.46 | 2029.75 | 3947.60 | 7365.38 | 11356.02 | 13216.74 | 13244.11 | 16239.39 |
| Antidepressants^1^ | 2.24 | 0.79 | 2.67 | 274.90 | 925.34 | 1234.34 | 1721.78 | 2049.10 | 1944.03 | 2256.25 | 4159.33 |
| Antihistamines^2^ | 0.31 | 4.98 | 44.56 | 337.13 | 820.88 | 1023.97 | 1200.50 | 1165.36 | 864.44 | 577.57 | 576.99 |
| Antipsychotics^3^ | 0.20 | 4.04 | 17.61 | 242.26 | 509.71 | 569.72 | 729.89 | 799.36 | 772.80 | 808.69 | 1650.34 |
| Sedatives | 181.81 | 214.19 | 340.45 | 3979.36 | 20727.61 | 32451.20 | 45320.08 | 58374.36 | 62449.98 | 60829.71 | 75226.25 |
| **2017** |  |  |  |  |  |  |  |  |  |  |  |
| Benzodiazepines and z-drugs | 140.91 | 218.63 | 245.72 | 3018.81 | 19449.79 | 31354.06 | 42306.56 | 53092.70 | 56555.12 | 54591.52 | 64997.00 |
| Long-acting benzodiazepines | 108.24 | 168.06 | 179.42 | 1242.23 | 7877.22 | 12339.93 | 13959.18 | 13389.76 | 12640.72 | 10185.76 | 8845.05 |
| Clonazepam | 7.12 | 12.77 | 11.70 | 63.85 | 259.37 | 299.82 | 347.71 | 362.47 | 239.33 | 216.70 | 173.69 |
| Diazepam | 26.64 | 21.73 | 45.96 | 849.96 | 6428.86 | 9557.36 | 9681.78 | 8354.53 | 7589.54 | 5773.95 | 4625.09 |
| Chloridiazepoxide | 0.00 | 0.20 | 0.19 | 22.10 | 143.54 | 254.90 | 344.56 | 371.66 | 301.75 | 253.92 | 220.32 |
| Potassium Clorazepate | 0.00 | 0.00 | 0.00 | 0.08 | 0.03 | 1.92 | 1.18 | 8.60 | 5.90 | 2.93 | 1.94 |
| Lorazepam | 0.74 | 2.14 | 25.44 | 101.54 | 273.32 | 291.55 | 448.52 | 794.33 | 834.45 | 877.92 | 1057.55 |
| Bromazepam | 0.00 | 0.04 | 0.96 | 19.00 | 94.44 | 208.29 | 416.40 | 923.05 | 1391.11 | 1346.79 | 1151.00 |
| Clobazam | 50.79 | 98.15 | 67.38 | 161.88 | 249.75 | 244.94 | 351.90 | 343.15 | 357.52 | 218.72 | 203.50 |
| Prazepam | 0.00 | 0.01 | 0.14 | 3.07 | 23.43 | 41.13 | 97.52 | 240.69 | 313.79 | 333.07 | 289.68 |
| Alprazolam | 1.63 | 6.63 | 7.37 | 500.65 | 3133.39 | 4100.26 | 4892.66 | 6068.23 | 5789.55 | 5445.86 | 5670.50 |
| Flurazepam | 5.94 | 0.18 | 3.31 | 95.87 | 694.90 | 1847.26 | 2885.87 | 3304.13 | 3163.01 | 2607.26 | 1931.59 |
| Nitrazepam | 17.75 | 35.01 | 50.74 | 45.44 | 77.34 | 92.59 | 248.65 | 404.54 | 669.89 | 779.20 | 1399.24 |
| Triazolam | 0.00 | 0.10 | 3.83 | 52.07 | 634.81 | 972.31 | 1350.23 | 2115.74 | 2694.59 | 2629.08 | 3838.89 |
| Lormetazepam | 0.15 | 2.68 | 0.32 | 21.39 | 53.88 | 94.53 | 397.71 | 822.95 | 1264.57 | 1698.49 | 3019.98 |
| Temazepam | 0.70 | 0.12 | 0.55 | 24.16 | 144.30 | 334.84 | 767.14 | 1560.43 | 2263.37 | 2520.02 | 4211.50 |
| Zopiclone | 24.29 | 27.11 | 18.24 | 722.58 | 5192.30 | 8929.66 | 12612.24 | 16321.47 | 16762.71 | 16889.17 | 21391.11 |
| Zolpidem | 5.16 | 11.74 | 9.61 | 335.18 | 2046.12 | 4082.69 | 7462.49 | 11096.73 | 12914.04 | 12998.44 | 15811.43 |
| Antidepressants^1^ | 2.75 | 0.22 | 4.28 | 312.53 | 1097.39 | 1447.99 | 1890.24 | 2192.88 | 1955.71 | 2347.64 | 4210.78 |
| Antihistamines^2^ | 0.00 | 2.52 | 43.91 | 521.34 | 1199.93 | 1366.70 | 1569.47 | 1575.08 | 1114.40 | 786.41 | 747.81 |
| Antipsychotics^3^ | 0.96 | 2.31 | 17.34 | 264.71 | 609.31 | 679.12 | 830.08 | 869.54 | 803.62 | 823.10 | 1682.78 |
| Sedatives | 144.62 | 223.68 | 311.25 | 4117.39 | 22356.42 | 34847.88 | 46596.35 | 57730.19 | 60428.84 | 58548.67 | 71638.37 |
| **2018** |  |  |  |  |  |  |  |  |  |  |  |
| Benzodiazepines and z-drugs | 115.06 | 182.45 | 294.30 | 2960.85 | 20200.11 | 31851.71 | 40673.54 | 51501.84 | 55539.20 | 52321.07 | 62962.61 |
| Long-acting benzodiazepines | 106.46 | 157.45 | 224.52 | 1124.34 | 8129.04 | 12599.21 | 13628.08 | 12982.77 | 12280.58 | 9632.62 | 8486.10 |
| Clonazepam | 12.77 | 8.70 | 12.03 | 67.64 | 278.68 | 320.52 | 338.31 | 369.64 | 260.64 | 213.25 | 183.57 |
| Diazepam | 11.35 | 24.19 | 63.05 | 749.69 | 6737.65 | 9883.12 | 9717.85 | 8241.51 | 7314.18 | 5617.35 | 4517.45 |
| Chloridiazepoxide | 0.75 | 0.26 | 0.00 | 23.87 | 154.37 | 240.55 | 293.98 | 359.35 | 341.32 | 193.98 | 197.23 |
| Potassium Clorazepate | 0.00 | 0.00 | 0.00 | 0.00 | 0.20 | 0.23 | 1.33 | 10.06 | 8.19 | 2.81 | 2.50 |
| Lorazepam | 0.49 | 2.71 | 23.50 | 140.86 | 361.97 | 307.89 | 449.18 | 742.76 | 874.84 | 823.09 | 1007.01 |
| Bromazepam | 0.46 | 0.04 | 2.42 | 21.63 | 84.18 | 198.46 | 362.89 | 848.16 | 1297.26 | 1329.62 | 1130.23 |
| Clobazam | 60.10 | 93.67 | 79.89 | 143.20 | 216.45 | 242.25 | 324.03 | 335.31 | 342.06 | 227.64 | 184.47 |
| Prazepam | 0.00 | 0.00 | 0.04 | 2.16 | 20.81 | 46.07 | 92.74 | 198.09 | 310.24 | 305.91 | 285.05 |
| Alprazolam | 0.61 | 2.28 | 10.08 | 541.04 | 3427.55 | 4299.08 | 4747.64 | 5896.61 | 5644.89 | 5149.45 | 5409.36 |
| Flurazepam | 0.09 | 0.02 | 0.16 | 98.89 | 643.54 | 1782.23 | 2655.13 | 3112.59 | 3029.01 | 2465.86 | 1861.30 |
| Nitrazepam | 21.39 | 30.61 | 69.36 | 38.87 | 77.35 | 84.24 | 204.72 | 356.24 | 674.94 | 605.82 | 1254.54 |
| Triazolam | 0.13 | 0.22 | 2.11 | 47.02 | 653.54 | 1002.39 | 1323.23 | 2025.99 | 2503.77 | 2517.76 | 3682.08 |
| Lormetazepam | 0.00 | 2.78 | 0.32 | 19.19 | 48.24 | 94.01 | 304.81 | 742.56 | 1165.26 | 1408.49 | 2807.84 |
| Temazepam | 0.00 | 0.17 | 4.46 | 25.41 | 143.39 | 334.75 | 654.99 | 1416.98 | 1986.22 | 2365.97 | 3777.31 |
| Zopiclone | 5.43 | 12.56 | 21.68 | 687.61 | 5311.54 | 8908.42 | 12122.70 | 16085.21 | 16675.56 | 16196.48 | 20936.50 |
| Zolpidem | 1.48 | 4.24 | 5.20 | 353.76 | 2040.65 | 4107.49 | 7080.01 | 10760.80 | 13110.81 | 12897.58 | 15726.17 |
| Antidepressants^1^ | 1.28 | 1.54 | 5.38 | 345.02 | 1306.29 | 1600.65 | 2027.78 | 2357.29 | 2221.69 | 2452.29 | 4522.02 |
| Antihistamines^2^ | 8.13 | 4.21 | 51.79 | 620.96 | 1436.61 | 1713.73 | 1963.51 | 1953.97 | 1285.81 | 916.55 | 833.75 |
| Antipsychotics^3^ | 0.70 | 3.45 | 27.02 | 298.79 | 756.28 | 766.67 | 892.60 | 965.58 | 851.29 | 861.96 | 1735.99 |
| Sedatives | 125.16 | 191.66 | 378.49 | 4225.61 | 23699.29 | 35932.76 | 45557.43 | 56778.68 | 59897.99 | 56551.86 | 70054.36 |
| **2019** |  |  |  |  |  |  |  |  |  |  |  |
| Benzodiazepines and z-drugs | 77.03 | 217.53 | 284.63 | 2800.18 | 20005.74 | 31147.37 | 40189.87 | 50940.09 | 54273.89 | 50694.29 | 60659.39 |
| Long-acting benzodiazepines | 69.43 | 199.81 | 232.43 | 1057.57 | 8086.53 | 12456.72 | 13637.21 | 12880.44 | 11764.92 | 9125.95 | 8247.78 |
| Clonazepam | 6.29 | 11.01 | 12.68 | 60.68 | 261.42 | 311.31 | 318.23 | 367.63 | 242.63 | 219.43 | 190.77 |
| Diazepam | 12.07 | 23.43 | 81.33 | 681.21 | 6812.13 | 9938.65 | 9948.84 | 8388.01 | 7097.92 | 5355.24 | 4468.14 |
| Chloridiazepoxide | 0.00 | 0.32 | 0.65 | 27.72 | 165.49 | 280.27 | 289.13 | 341.10 | 321.69 | 167.84 | 181.36 |
| Potassium Clorazepate | 0.00 | 0.00 | 0.00 | 0.00 | 0.31 | 0.08 | 2.07 | 8.76 | 12.78 | 2.84 | 1.77 |
| Lorazepam | 0.00 | 2.01 | 8.61 | 215.69 | 376.92 | 335.08 | 447.83 | 707.25 | 900.39 | 835.43 | 987.93 |
| Bromazepam | 0.18 | 0.04 | 0.67 | 22.32 | 90.66 | 182.16 | 343.45 | 772.36 | 1191.69 | 1287.92 | 1089.78 |
| Clobazam | 26.00 | 112.60 | 71.97 | 147.53 | 212.24 | 257.34 | 293.50 | 301.22 | 335.87 | 224.00 | 170.13 |
| Prazepam | 0.00 | 0.00 | 0.05 | 2.96 | 13.17 | 35.65 | 81.39 | 176.97 | 328.98 | 286.91 | 261.95 |
| Alprazolam | 4.77 | 2.49 | 13.10 | 457.48 | 3438.08 | 4219.34 | 4726.48 | 5760.28 | 5700.23 | 4955.35 | 5267.81 |
| Flurazepam | 0.00 | 0.26 | 3.94 | 80.06 | 563.67 | 1560.45 | 2506.34 | 2952.50 | 2882.29 | 2285.66 | 1801.34 |
| Nitrazepam | 25.07 | 52.20 | 61.79 | 57.41 | 58.08 | 72.96 | 197.70 | 344.25 | 542.75 | 584.03 | 1172.32 |
| Triazolam | 0.41 | 0.04 | 0.82 | 37.04 | 606.81 | 909.04 | 1278.17 | 2141.02 | 2378.11 | 2481.28 | 3532.88 |
| Lormetazepam | 0.00 | 2.31 | 0.00 | 6.32 | 36.01 | 35.56 | 137.40 | 384.24 | 572.36 | 784.11 | 1533.94 |
| Temazepam | 0.03 | 0.00 | 5.26 | 28.61 | 146.30 | 306.47 | 587.52 | 1340.76 | 1788.97 | 2194.82 | 3508.81 |
| Zopiclone | 1.97 | 6.57 | 14.93 | 639.05 | 5175.89 | 8771.38 | 12229.63 | 16133.42 | 16998.54 | 15969.55 | 20740.55 |
| Zolpidem | 0.24 | 4.26 | 8.82 | 336.09 | 2048.55 | 3931.63 | 6802.17 | 10820.32 | 12978.68 | 13059.87 | 15749.90 |
| Antidepressants^1^ | 0.17 | 1.70 | 4.90 | 371.84 | 1454.10 | 1668.21 | 2163.38 | 2563.63 | 2524.43 | 2595.68 | 4762.78 |
| Antihistamines^2^ | 5.97 | 13.29 | 62.35 | 909.10 | 2080.42 | 2321.38 | 2486.26 | 2551.88 | 1570.77 | 1171.45 | 1029.58 |
| Antipsychotics^3^ | 0.16 | 3.78 | 33.56 | 362.08 | 901.91 | 889.87 | 965.99 | 1048.82 | 906.73 | 888.75 | 1784.35 |
| Sedatives | 83.33 | 236.30 | 385.45 | 4443.21 | 24442.17 | 36026.84 | 45805.49 | 57104.42 | 59275.83 | 55350.17 | 68236.10 |
| **2020** |  |  |  |  |  |  |  |  |  |  |  |
| Benzodiazepines and z-drugs | 86.42 | 196.14 | 264.18 | 2369.07 | 17848.51 | 29594.11 | 39186.72 | 49404.17 | 52203.71 | 49104.61 | 58437.76 |
| Long-acting benzodiazepines | 78.72 | 185.59 | 211.21 | 960.70 | 7271.88 | 11701.63 | 13321.52 | 12670.26 | 10773.68 | 8874.16 | 7846.56 |
| Clonazepam | 5.70 | 10.02 | 14.09 | 51.96 | 223.53 | 292.85 | 308.16 | 368.10 | 275.88 | 219.94 | 201.98 |
| Diazepam | 24.65 | 14.84 | 66.06 | 629.61 | 6184.09 | 9557.84 | 9922.84 | 8479.15 | 6499.15 | 5321.58 | 4302.76 |
| Chloridiazepoxide | 0.41 | 0.54 | 0.38 | 19.99 | 130.69 | 254.65 | 265.05 | 319.33 | 243.72 | 175.24 | 160.88 |
| Potassium Clorazepate | 0.00 | 0.00 | 0.00 | 0.00 | 0.88 | 0.16 | 0.44 | 8.43 | 12.90 | 2.86 | 1.91 |
| Lorazepam | 0.03 | 3.64 | 11.49 | 220.34 | 388.51 | 356.02 | 464.15 | 697.50 | 885.46 | 826.56 | 986.71 |
| Bromazepam | 0.07 | 0.05 | 0.29 | 15.99 | 94.67 | 168.14 | 296.40 | 718.36 | 1141.93 | 1210.46 | 1055.64 |
| Clobazam | 36.55 | 110.90 | 51.98 | 130.83 | 195.06 | 249.86 | 286.95 | 286.82 | 313.24 | 222.61 | 176.66 |
| Prazepam | 0.00 | 0.03 | 0.03 | 1.86 | 9.91 | 23.55 | 77.06 | 146.92 | 268.12 | 249.93 | 224.95 |
| Alprazolam | 2.37 | 1.43 | 7.58 | 354.12 | 2985.13 | 3940.33 | 4570.46 | 5463.18 | 5590.35 | 4861.45 | 5116.73 |
| Flurazepam | 0.00 | 0.02 | 3.69 | 61.74 | 460.62 | 1258.89 | 2288.57 | 2725.04 | 2702.70 | 2130.09 | 1716.02 |
| Nitrazepam | 11.42 | 49.25 | 74.98 | 64.71 | 67.11 | 63.84 | 172.46 | 336.46 | 457.99 | 551.90 | 1061.39 |
| Triazolam | 0.00 | 0.00 | 1.56 | 31.49 | 488.09 | 853.61 | 1249.92 | 2067.78 | 2195.95 | 2523.70 | 3400.98 |
| Lormetazepam | 0.00 | 0.00 | 1.42 | 0.15 | 1.45 | 1.16 | 8.49 | 34.01 | 59.24 | 57.86 | 121.18 |
| Temazepam | 0.00 | 0.04 | 4.77 | 16.56 | 150.27 | 294.77 | 554.14 | 1230.77 | 1626.54 | 1990.88 | 3270.74 |
| Zopiclone | 2.69 | 3.88 | 14.05 | 525.60 | 4657.96 | 8462.67 | 12028.86 | 15718.83 | 17090.19 | 15628.18 | 20701.40 |
| Zolpidem | 2.53 | 1.52 | 11.81 | 244.10 | 1810.54 | 3815.78 | 6692.78 | 10803.48 | 12840.36 | 13131.37 | 15937.82 |
| Antidepressants^1^ | 0.46 | 2.21 | 3.74 | 368.19 | 1615.14 | 1856.83 | 2317.10 | 2804.47 | 2822.16 | 2855.86 | 5191.99 |
| Antihistamines^2^ | 0.20 | 12.44 | 49.61 | 1022.53 | 2806.39 | 3087.57 | 3357.15 | 3234.56 | 2081.65 | 1447.86 | 1366.73 |
| Antipsychotics^3^ | 0.02 | 3.61 | 32.79 | 367.84 | 1018.86 | 1005.58 | 1090.92 | 1128.64 | 976.00 | 951.16 | 1862.56 |
| Sedatives | 87.11 | 214.39 | 350.32 | 4127.63 | 23288.90 | 35544.08 | 45951.89 | 56571.84 | 58083.51 | 54359.49 | 66859.05 |
| **2021** |  |  |  |  |  |  |  |  |  |  |  |
| Benzodiazepines and z-drugs | 93.01 | 200.67 | 186.89 | 2400.23 | 17248.79 | 29431.65 | 38455.57 | 48097.46 | 50467.97 | 47440.11 | 55791.72 |
| Long-acting benzodiazepines | 87.35 | 188.54 | 147.74 | 1066.43 | 7113.11 | 11572.87 | 13241.10 | 12410.58 | 10292.65 | 8151.21 | 6865.08 |
| Clonazepam | 13.77 | 13.93 | 11.61 | 55.56 | 238.42 | 296.95 | 315.93 | 361.18 | 294.82 | 213.51 | 214.63 |
| Diazepam | 22.17 | 25.58 | 38.09 | 715.66 | 6070.98 | 9522.80 | 10078.30 | 8655.92 | 6467.21 | 5149.21 | 4126.29 |
| Chloridiazepoxide | 0.04 | 0.28 | 0.15 | 35.15 | 129.82 | 280.99 | 258.50 | 318.77 | 220.08 | 154.01 | 139.78 |
| Potassium Clorazepate | 0.00 | 0.00 | 0.00 | 0.00 | 2.61 | 0.00 | 0.38 | 4.07 | 5.86 | 7.68 | 2.58 |
| Lorazepam | 0.00 | 4.74 | 13.23 | 254.91 | 414.74 | 396.00 | 467.34 | 686.68 | 897.61 | 747.52 | 967.34 |
| Bromazepam | 0.00 | 0.02 | 1.02 | 13.13 | 101.68 | 159.67 | 255.49 | 652.38 | 1072.95 | 1106.13 | 1024.95 |
| Clobazam | 38.25 | 115.20 | 70.08 | 156.18 | 200.93 | 243.33 | 290.95 | 308.10 | 283.81 | 216.67 | 162.44 |
| Prazepam | 0.00 | 0.00 | 0.00 | 3.83 | 10.48 | 26.79 | 69.48 | 122.84 | 234.81 | 239.67 | 217.68 |
| Alprazolam | 0.20 | 2.46 | 5.54 | 279.08 | 2695.20 | 4012.36 | 4440.20 | 5226.06 | 5352.84 | 4737.83 | 4885.43 |
| Flurazepam | 0.00 | 1.06 | 0.00 | 65.30 | 436.53 | 1179.94 | 2170.38 | 2531.15 | 2638.08 | 1981.70 | 1632.63 |
| Nitrazepam | 13.12 | 32.49 | 27.81 | 34.75 | 23.34 | 22.07 | 57.17 | 108.55 | 147.98 | 188.76 | 369.04 |
| Triazolam | 0.00 | 0.00 | 0.28 | 34.86 | 411.55 | 837.32 | 1214.77 | 1889.17 | 2175.91 | 2413.16 | 3234.44 |
| Lormetazepam | 0.00 | 0.00 | 0.00 | 0.00 | 0.00 | 0.13 | 0.53 | 1.29 | 5.95 | 7.82 | 10.07 |
| Temazepam | 0.00 | 0.00 | 3.72 | 10.70 | 131.09 | 273.69 | 525.57 | 1123.00 | 1501.80 | 1823.94 | 2996.09 |
| Zopiclone | 4.46 | 1.95 | 9.92 | 497.34 | 4544.10 | 8460.16 | 11780.21 | 15625.81 | 16588.51 | 15489.83 | 20173.63 |
| Zolpidem | 1.01 | 2.95 | 5.45 | 243.77 | 1837.31 | 3719.46 | 6530.36 | 10482.49 | 12579.75 | 12962.67 | 15634.70 |
| Antidepressants^1^ | 0.24 | 3.72 | 0.78 | 412.96 | 1825.95 | 2088.85 | 2553.58 | 3075.76 | 3116.22 | 3132.35 | 5743.26 |
| Antihistamines^2^ | 1.61 | 12.10 | 76.05 | 1349.32 | 3734.25 | 3976.43 | 4274.73 | 4244.17 | 2648.11 | 1786.52 | 1647.22 |
| Antipsychotics^3^ | 0.00 | 3.89 | 26.27 | 424.77 | 1131.89 | 1131.28 | 1179.85 | 1224.21 | 1114.09 | 968.12 | 1874.63 |
| Sedatives | 94.86 | 220.38 | 289.99 | 4587.28 | 23940.88 | 36628.20 | 46463.73 | 56641.61 | 57346.38 | 53327.10 | 65056.83 |
| **2022** |  |  |  |  |  |  |  |  |  |  |  |
| Benzodiazepines and z-drugs | 71.47 | 148.91 | 198.03 | 2330.32 | 15048.91 | 27401.49 | 36574.96 | 45340.23 | 47607.25 | 45178.67 | 53089.38 |
| Long-acting benzodiazepines | 69.42 | 133.41 | 155.20 | 1057.66 | 6252.96 | 10783.19 | 12622.28 | 11844.51 | 9600.17 | 7718.35 | 6246.80 |
| Clonazepam | 7.02 | 10.93 | 16.21 | 48.21 | 188.80 | 245.54 | 273.03 | 308.50 | 279.85 | 190.62 | 190.89 |
| Diazepam | 17.98 | 18.08 | 40.88 | 720.95 | 5331.16 | 9073.89 | 9879.85 | 8487.91 | 6389.44 | 5090.38 | 4067.94 |
| Chloridiazepoxide | 0.00 | 0.07 | 0.07 | 35.67 | 129.33 | 239.95 | 230.80 | 277.56 | 200.01 | 166.97 | 120.74 |
| Potassium Clorazepate | 0.00 | 0.00 | 0.00 | 0.00 | 0.52 | 0.27 | 0.00 | 5.95 | 6.34 | 4.53 | 3.57 |
| Lorazepam | 0.46 | 8.94 | 16.65 | 256.49 | 411.16 | 417.57 | 495.46 | 630.90 | 825.49 | 728.88 | 932.81 |
| Bromazepam | 0.08 | 0.00 | 1.16 | 12.82 | 82.99 | 165.92 | 232.51 | 571.83 | 989.50 | 1025.93 | 980.87 |
| Clobazam | 41.23 | 93.41 | 96.25 | 183.71 | 235.16 | 231.47 | 281.65 | 335.66 | 262.22 | 210.35 | 166.04 |
| Prazepam | 0.00 | 0.00 | 0.00 | 1.11 | 5.02 | 11.74 | 40.44 | 61.09 | 130.16 | 136.99 | 143.60 |
| Alprazolam | 0.23 | 1.48 | 7.35 | 246.38 | 2423.80 | 3842.75 | 4230.43 | 4953.34 | 4974.33 | 4497.05 | 4605.65 |
| Flurazepam | 0.00 | 0.18 | 0.83 | 65.22 | 360.09 | 975.47 | 1909.87 | 2360.80 | 2322.41 | 1912.03 | 1519.28 |
| Nitrazepam | 3.19 | 10.74 | 0.96 | 2.79 | 2.90 | 4.86 | 6.64 | 7.05 | 9.75 | 6.48 | 34.74 |
| Triazolam | 0.00 | 0.00 | 2.95 | 22.00 | 310.54 | 785.46 | 1158.64 | 1741.51 | 1966.86 | 2249.81 | 3057.37 |
| Lormetazepam | 0.00 | 0.00 | 0.00 | 0.00 | 0.26 | 0.00 | 0.18 | 0.49 | 0.00 | 3.59 | 2.99 |
| Temazepam | 0.00 | 0.01 | 0.07 | 14.97 | 101.49 | 227.36 | 440.20 | 967.77 | 1309.99 | 1651.14 | 2731.44 |
| Zopiclone | 0.53 | 3.88 | 8.52 | 478.27 | 3947.82 | 7864.60 | 11148.55 | 14829.66 | 15796.37 | 14980.17 | 19357.99 |
| Zolpidem | 0.75 | 1.18 | 6.13 | 241.72 | 1517.89 | 3314.66 | 6246.72 | 9800.22 | 12144.54 | 12323.76 | 15173.46 |
| Antidepressants^1^ | 0.00 | 2.61 | 9.97 | 351.67 | 1660.69 | 2021.83 | 2577.28 | 3198.25 | 3132.88 | 3252.95 | 6188.08 |
| Antihistamines^2^ | 0.16 | 11.34 | 91.14 | 1164.89 | 3708.64 | 4194.70 | 4816.65 | 4644.76 | 3048.14 | 2027.84 | 1887.49 |
| Antipsychotics^3^ | 0.00 | 3.62 | 23.54 | 367.28 | 1101.81 | 1130.02 | 1206.63 | 1228.05 | 1075.51 | 930.78 | 1826.21 |
| Sedatives | 71.63 | 166.47 | 322.69 | 4214.17 | 21520.04 | 34748.04 | 45175.52 | 54411.29 | 54863.78 | 51390.23 | 62991.16 |

**Rate of DDDs per 1,000 GMS population by sex**

|  | **Females** | **Males** |
| --- | --- | --- |
| **2014** |  |  |
| Benzodiazepines and z-drugs | 37684.07 | 24839.12 |
| Long-acting benzodiazepines | 8336.46 | 7659.00 |
| Clonazepam | 168.43 | 168.92 |
| Diazepam | 5032.87 | 4892.21 |
| Chloridiazepoxide | 161.07 | 248.16 |
| Potassium Clorazepate | 2.71 | 1.39 |
| Lorazepam | 517.49 | 396.20 |
| Bromazepam | 776.66 | 293.06 |
| Clobazam | 279.73 | 274.41 |
| Prazepam | 188.03 | 84.94 |
| Alprazolam | 4154.71 | 2547.67 |
| Flurazepam | 1840.72 | 1696.66 |
| Nitrazepam | 662.90 | 292.31 |
| Triazolam | 1760.72 | 1075.69 |
| Lormetazepam | 1244.42 | 486.51 |
| Temazepam | 1887.89 | 997.22 |
| Zopiclone | 10972.23 | 7334.91 |
| Zolpidem | 8033.48 | 4048.85 |
| Antidepressants^1^ | 1375.70 | 879.64 |
| Antihistamines^2^ | 461.66 | 328.39 |
| Antipsychotics^3^ | 567.60 | 438.63 |
| Sedatives | 40089.02 | 26485.78 |
| **2015** |  |  |
| Benzodiazepines and z-drugs | 37330.34 | 24683.13 |
| Long-acting benzodiazepines | 8126.93 | 7580.27 |
| Clonazepam | 169.13 | 173.08 |
| Diazepam | 5039.91 | 4956.56 |
| Chloridiazepoxide | 152.23 | 235.62 |
| Potassium Clorazepate | 2.60 | 1.40 |
| Lorazepam | 501.67 | 391.82 |
| Bromazepam | 758.00 | 281.50 |
| Clobazam | 239.57 | 249.58 |
| Prazepam | 180.60 | 83.96 |
| Alprazolam | 4134.09 | 2515.89 |
| Flurazepam | 1743.17 | 1623.71 |
| Nitrazepam | 599.72 | 256.37 |
| Triazolam | 1756.11 | 1089.76 |
| Lormetazepam | 1160.90 | 450.07 |
| Temazepam | 1724.22 | 923.26 |
| Zopiclone | 11084.86 | 7422.11 |
| Zolpidem | 8083.56 | 4028.45 |
| Antidepressants^1^ | 1570.25 | 1010.33 |
| Antihistamines^2^ | 564.46 | 409.06 |
| Antipsychotics^3^ | 618.91 | 487.47 |
| Sedatives | 40083.96 | 26590.00 |
| **2016** |  |  |
| Benzodiazepines and z-drugs | 38115.81 | 25515.88 |
| Long-acting benzodiazepines | 8145.90 | 7761.33 |
| Clonazepam | 177.55 | 188.91 |
| Diazepam | 5142.47 | 5143.65 |
| Chloridiazepoxide | 152.01 | 245.31 |
| Potassium Clorazepate | 2.45 | 1.27 |
| Lorazepam | 506.18 | 410.80 |
| Bromazepam | 751.18 | 283.50 |
| Clobazam | 227.05 | 237.76 |
| Prazepam | 176.60 | 83.20 |
| Alprazolam | 4227.67 | 2627.71 |
| Flurazepam | 1692.50 | 1613.10 |
| Nitrazepam | 575.27 | 248.14 |
| Triazolam | 1754.15 | 1106.08 |
| Lormetazepam | 1140.61 | 441.86 |
| Temazepam | 1645.86 | 890.20 |
| Zopiclone | 11444.87 | 7792.47 |
| Zolpidem | 8499.40 | 4201.92 |
| Antidepressants^1^ | 1760.24 | 1164.90 |
| Antihistamines^2^ | 755.73 | 537.70 |
| Antipsychotics^3^ | 688.25 | 549.02 |
| Sedatives | 41320.04 | 27767.50 |
| **2017** |  |  |
| Benzodiazepines and z-drugs | 38182.32 | 25987.00 |
| Long-acting benzodiazepines | 8021.13 | 7890.70 |
| Clonazepam | 191.78 | 202.04 |
| Diazepam | 5143.76 | 5311.37 |
| Chloridiazepoxide | 143.81 | 241.33 |
| Potassium Clorazepate | 2.60 | 1.53 |
| Lorazepam | 499.24 | 427.83 |
| Bromazepam | 733.22 | 278.52 |
| Clobazam | 215.64 | 231.48 |
| Prazepam | 169.93 | 78.30 |
| Alprazolam | 4244.73 | 2684.10 |
| Flurazepam | 1622.96 | 1583.44 |
| Nitrazepam | 530.63 | 241.22 |
| Triazolam | 1738.58 | 1106.14 |
| Lormetazepam | 1096.18 | 414.79 |
| Temazepam | 1555.59 | 840.78 |
| Zopiclone | 11616.72 | 8028.32 |
| Zolpidem | 8676.93 | 4315.80 |
| Antidepressants^1^ | 1918.46 | 1277.52 |
| Antihistamines^2^ | 1016.20 | 725.69 |
| Antipsychotics^3^ | 758.57 | 607.80 |
| Sedatives | 41875.54 | 28598.01 |
| **2018** |  |  |
| Benzodiazepines and z-drugs | 38075.10 | 26060.79 |
| Long-acting benzodiazepines | 7913.51 | 7908.11 |
| Clonazepam | 198.99 | 211.07 |
| Diazepam | 5146.17 | 5411.15 |
| Chloridiazepoxide | 131.93 | 234.26 |
| Potassium Clorazepate | 2.94 | 1.72 |
| Lorazepam | 510.30 | 435.79 |
| Bromazepam | 722.80 | 270.39 |
| Clobazam | 205.36 | 223.11 |
| Prazepam | 164.31 | 76.96 |
| Alprazolam | 4246.47 | 2697.37 |
| Flurazepam | 1577.51 | 1526.45 |
| Nitrazepam | 486.30 | 223.39 |
| Triazolam | 1726.99 | 1096.65 |
| Lormetazepam | 1032.45 | 380.19 |
| Temazepam | 1459.30 | 784.00 |
| Zopiclone | 11662.04 | 8077.79 |
| Zolpidem | 8801.25 | 4410.50 |
| Antidepressants^1^ | 2137.71 | 1410.83 |
| Antihistamines^2^ | 1243.68 | 877.18 |
| Antipsychotics^3^ | 833.97 | 670.79 |
| Sedatives | 42290.46 | 29019.60 |
| **2019** |  |  |
| Benzodiazepines and z-drugs | 37627.73 | 25961.66 |
| Long-acting benzodiazepines | 7799.19 | 7889.80 |
| Clonazepam | 194.30 | 208.15 |
| Diazepam | 5155.85 | 5489.80 |
| Chloridiazepoxide | 128.88 | 236.10 |
| Potassium Clorazepate | 3.07 | 1.89 |
| Lorazepam | 511.09 | 459.97 |
| Bromazepam | 701.60 | 256.42 |
| Clobazam | 194.45 | 218.55 |
| Prazepam | 155.03 | 73.20 |
| Alprazolam | 4216.56 | 2674.33 |
| Flurazepam | 1509.43 | 1439.70 |
| Nitrazepam | 458.17 | 222.41 |
| Triazolam | 1725.34 | 1070.41 |
| Lormetazepam | 572.42 | 199.10 |
| Temazepam | 1395.51 | 732.78 |
| Zopiclone | 11773.45 | 8224.04 |
| Zolpidem | 8932.57 | 4454.79 |
| Antidepressants^1^ | 2311.38 | 1544.90 |
| Antihistamines^2^ | 1630.29 | 1157.15 |
| Antipsychotics^3^ | 902.46 | 742.53 |
| Sedatives | 42471.85 | 29406.24 |
| **2020** |  |  |
| Benzodiazepines and z-drugs | 36074.15 | 24879.40 |
| Long-acting benzodiazepines | 7400.58 | 7544.63 |
| Clonazepam | 191.70 | 204.25 |
| Diazepam | 4962.29 | 5364.56 |
| Chloridiazepoxide | 114.64 | 211.64 |
| Potassium Clorazepate | 2.96 | 1.64 |
| Lorazepam | 519.15 | 456.89 |
| Bromazepam | 659.23 | 241.08 |
| Clobazam | 190.84 | 207.00 |
| Prazepam | 130.48 | 62.17 |
| Alprazolam | 4046.71 | 2501.02 |
| Flurazepam | 1399.19 | 1274.94 |
| Nitrazepam | 408.48 | 218.43 |
| Triazolam | 1637.86 | 1045.91 |
| Lormetazepam | 45.22 | 16.54 |
| Temazepam | 1283.48 | 684.80 |
| Zopiclone | 11589.18 | 7993.06 |
| Zolpidem | 8892.75 | 4395.49 |
| Antidepressants^1^ | 2534.20 | 1677.51 |
| Antihistamines^2^ | 2121.35 | 1518.47 |
| Antipsychotics^3^ | 983.51 | 795.28 |
| Sedatives | 41713.21 | 28870.66 |
| **2021** |  |  |
| Benzodiazepines and z-drugs | 35508.73 | 24901.54 |
| Long-acting benzodiazepines | 7057.17 | 7502.01 |
| Clonazepam | 198.33 | 213.09 |
| Diazepam | 4946.79 | 5477.83 |
| Chloridiazepoxide | 111.44 | 213.43 |
| Potassium Clorazepate | 2.87 | 1.13 |
| Lorazepam | 526.61 | 468.90 |
| Bromazepam | 634.71 | 231.28 |
| Clobazam | 189.86 | 214.71 |
| Prazepam | 123.70 | 60.74 |
| Alprazolam | 3945.01 | 2475.77 |
| Flurazepam | 1339.68 | 1238.60 |
| Nitrazepam | 144.49 | 82.48 |
| Triazolam | 1594.12 | 1029.83 |
| Lormetazepam | 3.95 | 1.56 |
| Temazepam | 1213.62 | 654.83 |
| Zopiclone | 11605.46 | 8100.87 |
| Zolpidem | 8928.09 | 4436.50 |
| Antidepressants^1^ | 2867.99 | 1901.98 |
| Antihistamines^2^ | 2724.41 | 1963.29 |
| Antipsychotics^3^ | 1068.17 | 858.42 |
| Sedatives | 42169.29 | 29625.23 |
| **2022** |  |  |
| Benzodiazepines and z-drugs | 33461.10 | 23810.96 |
| Long-acting benzodiazepines | 6547.52 | 7142.30 |
| Clonazepam | 173.11 | 184.04 |
| Diazepam | 4790.05 | 5350.86 |
| Chloridiazepoxide | 99.15 | 195.50 |
| Potassium Clorazepate | 2.84 | 1.16 |
| Lorazepam | 511.30 | 466.00 |
| Bromazepam | 587.55 | 220.41 |
| Clobazam | 196.97 | 215.80 |
| Prazepam | 72.25 | 36.32 |
| Alprazolam | 3697.71 | 2386.36 |
| Flurazepam | 1202.29 | 1148.10 |
| Nitrazepam | 10.85 | 10.53 |
| Triazolam | 1488.14 | 967.00 |
| Lormetazepam | 1.51 | 0.11 |
| Temazepam | 1083.51 | 592.08 |
| Zopiclone | 11040.59 | 7773.39 |
| Zolpidem | 8503.26 | 4263.31 |
| Antidepressants^1^ | 2960.04 | 1989.49 |
| Antihistamines^2^ | 2943.45 | 2156.70 |
| Antipsychotics^3^ | 1052.48 | 850.55 |
| Sedatives | 40417.08 | 28807.71 |

**Rate of DDEs per 1,000 GMS population by age group**

|  | **<5** | **5-11** | **12-15** | **16-24** | **25-34** | **35-44** | **45-54** | **55-64** | **65-69** | **70-74** | **75+** |
| --- | --- | --- | --- | --- | --- | --- | --- | --- | --- | --- | --- |
| 2014 |  |  |  |  |  |  |  |  |  |  |  |
| Benzodiazepines and z-drugs | 5123.26 | 5537.32 | 6633.29 | 39507.71 | 197748.7 | 319806.1 | 426507.9 | 566097.6 | 584127.4 | 542488 | 627196.3 |
| Long-acting benzodiazepines | 4962.50 | 5185.47 | 5337.72 | 20671.36 | 99179.80 | 163169.4 | 187418.1 | 209844.9 | 185082.3 | 151261.7 | 133264 |
| Clonazepam | 3296.60 | 3475.96 | 3206.18 | 8923.92 | 28486.92 | 41381.84 | 52555.71 | 49225.85 | 37599.67 | 29504.43 | 26791.40 |
| Diazepam | 189.07 | 430.90 | 492.25 | 8221.45 | 54175.78 | 87315.05 | 84391.02 | 92265.28 | 81128.36 | 65430.41 | 53211.10 |
| Chloridiazepoxide | 0.15 | 5.89 | 103.27 | 240.96 | 1697.77 | 2575.83 | 4765.23 | 5807.68 | 3979.19 | 3646.57 | 3419.36 |
| Potassium Clorazepate | 0.00 | 0.00 | 0.00 | 10.69 | 2.79 | 8.19 | 104.86 | 83.22 | 43.08 | 39.98 | 27.74 |
| Lorazepam | 4.45 | 102.58 | 249.95 | 1809.24 | 4731.97 | 6061.21 | 11036.94 | 20248.98 | 28014.35 | 23725.94 | 32531.34 |
| Bromazepam | 0.00 | 0.85 | 21.01 | 339.43 | 1619.42 | 3483.39 | 9262.88 | 22065.31 | 30059.26 | 26581.44 | 24168.98 |
| Clobazam | 1202.30 | 1164.29 | 1228.61 | 1856.66 | 3144.72 | 3541.15 | 4238.95 | 4393.64 | 3919.49 | 3111.33 | 2301.03 |
| Prazepam | 0.89 | 2.19 | 0.69 | 97.54 | 406.31 | 948.08 | 2715.29 | 7089.70 | 8697.83 | 8462.70 | 6747.12 |
| Alprazolam | 104.21 | 114.75 | 846.59 | 10465.12 | 54157.08 | 73677.17 | 99278.39 | 130541.5 | 128494.4 | 117179 | 124749.6 |
| Flurazepam | 4.03 | 14.07 | 184.61 | 1084.69 | 10865.72 | 26684.67 | 37086.53 | 47927.79 | 45349.60 | 35531.31 | 30892.12 |
| Nitrazepam | 269.46 | 92.16 | 122.12 | 235.45 | 399.79 | 714.59 | 1560.52 | 3051.70 | 4365.10 | 5534.94 | 9874.15 |
| Triazolam | 0.45 | 10.99 | 0.49 | 419.83 | 3121.39 | 4358.07 | 7371.33 | 11303.68 | 14679.66 | 15218.75 | 21986.57 |
| Lormetazepam | 1.70 | 8.36 | 38.09 | 82.77 | 358.67 | 1042.17 | 3244.07 | 6828.68 | 11282.81 | 16165.91 | 25643.94 |
| Temazepam | 12.08 | 3.03 | 13.89 | 262.59 | 1558.80 | 4082.39 | 10548.58 | 20154.96 | 31639.88 | 31837.17 | 58960.09 |
| Zopiclone | 10.40 | 75.53 | 79.11 | 3558.08 | 23507.63 | 43609.98 | 61742.71 | 87999.95 | 89990.23 | 93164.36 | 122567.8 |
| Zolpidem | 27.46 | 35.74 | 46.43 | 1899.29 | 9513.94 | 20322.26 | 36604.84 | 57109.67 | 64884.52 | 67353.76 | 83323.90 |
| **2015** |  |  |  |  |  |  |  |  |  |  |  |
| Benzodiazepines and z-drugs | 2638.15 | 4451.50 | 6605.36 | 39232.20 | 197921.5 | 305076.9 | 426566 | 548358.7 | 565852.1 | 522768.7 | 591651.5 |
| Long-acting benzodiazepines | 2512.16 | 4152.37 | 5038.86 | 21316.18 | 99843.30 | 154598.8 | 190020.8 | 202664.6 | 180952.4 | 146186.5 | 124472.4 |
| Clonazepam | 1403.02 | 2595.58 | 2611.30 | 9732.23 | 30272.11 | 39720.94 | 53282.96 | 50653.93 | 38921.22 | 28834.52 | 25801.88 |
| Diazepam | 96.94 | 494.29 | 966.64 | 8239.31 | 55385.68 | 83913.98 | 88776.85 | 88862.09 | 80643.51 | 63175.10 | 50982.96 |
| Chloridiazepoxide | 1.11 | 1.75 | 103.31 | 181.23 | 1546.28 | 2813.92 | 4051.06 | 4967.59 | 4222.82 | 3485.14 | 3114.33 |
| Potassium Clorazepate | 0.00 | 0.00 | 0.00 | 3.29 | 17.57 | 7.64 | 110.11 | 87.46 | 39.27 | 25.20 | 10.89 |
| Lorazepam | 4.45 | 47.42 | 434.71 | 2197.58 | 5178.75 | 6112.74 | 10866.18 | 19250.37 | 25605.61 | 23561.31 | 29345.22 |
| Bromazepam | 0.27 | 0.81 | 18.36 | 414.22 | 1514.86 | 3397.90 | 8575.58 | 19966.66 | 28255.54 | 25627.88 | 23034.14 |
| Clobazam | 834.96 | 958.70 | 931.93 | 1685.08 | 2441.52 | 2846.63 | 4081.04 | 3764.94 | 3582.83 | 2916.35 | 2224.93 |
| Prazepam | 0.00 | 0.00 | 0.48 | 96.93 | 366.12 | 996.53 | 2361.80 | 6754.45 | 8048.26 | 7787.51 | 6422.31 |
| Alprazolam | 74.05 | 147.33 | 823.54 | 8970.71 | 54000.61 | 70532.92 | 97337.20 | 126715.6 | 124434.8 | 114748.6 | 120267.8 |
| Flurazepam | 1.19 | 18.89 | 257.06 | 1141.45 | 9457.35 | 23799.38 | 35986.26 | 44875.39 | 41491.35 | 34914.46 | 27469.11 |
| Nitrazepam | 174.95 | 83.17 | 168.14 | 236.65 | 356.66 | 499.76 | 1370.69 | 2698.79 | 4003.13 | 5048.16 | 8445.98 |
| Triazolam | 1.38 | 1.33 | 14.08 | 328.23 | 3343.64 | 4427.63 | 6966.45 | 11445.77 | 15074.04 | 14123.59 | 20995.61 |
| Lormetazepam | 0.93 | 0.36 | 56.34 | 69.50 | 382.46 | 936.18 | 2936.26 | 6019.05 | 9699.92 | 14515.94 | 23374.08 |
| Temazepam | 0.00 | 13.43 | 8.73 | 303.12 | 1476.07 | 3611.92 | 9428.24 | 18156.93 | 28198.48 | 28131.14 | 52107.79 |
| Zopiclone | 12.09 | 68.42 | 138.50 | 3744.96 | 22895.87 | 42419.27 | 63301.11 | 87253.63 | 89771.66 | 90861.79 | 118406.5 |
| Zolpidem | 32.82 | 20.03 | 72.22 | 1887.69 | 9285.99 | 19039.55 | 37134.23 | 56886.08 | 63859.70 | 65011.95 | 79648.02 |
| **2016** |  |  |  |  |  |  |  |  |  |  |  |
| Benzodiazepines and z-drugs | 3131.59 | 3589.55 | 4111.53 | 42374.51 | 216567.5 | 319040.4 | 429787.3 | 533020.2 | 553451.6 | 520226.1 | 581939.9 |
| Long-acting benzodiazepines | 2889.19 | 3247.08 | 2848.00 | 22832.83 | 109639.4 | 161166.6 | 192429 | 196060.5 | 176818.7 | 146368.4 | 121110.4 |
| Clonazepam | 1677.67 | 1807.32 | 1333.01 | 10829.99 | 34847.98 | 43499.69 | 53578.89 | 52190.42 | 39672.32 | 32760.90 | 26794.97 |
| Diazepam | 387.85 | 285.74 | 439.37 | 8500.41 | 60447.50 | 88223.97 | 91610.75 | 85400.05 | 79128.13 | 61467.20 | 49528.10 |
| Chloridiazepoxide | 1.66 | 5.09 | 5.97 | 259.67 | 1938.89 | 3103.62 | 3948.82 | 4816.51 | 3887.93 | 3504.74 | 2824.12 |
| Potassium Clorazepate | 0.00 | 0.00 | 0.00 | 0.00 | 20.75 | 1.89 | 97.85 | 34.36 | 118.82 | 1.77 | 21.90 |
| Lorazepam | 22.96 | 42.55 | 605.80 | 2670.13 | 5959.19 | 6783.20 | 11334.21 | 19088.70 | 22259.60 | 22161.15 | 29270.49 |
| Bromazepam | 0.71 | 4.65 | 10.26 | 356.09 | 1876.35 | 3689.28 | 8073.01 | 17941.42 | 26828.20 | 25803.74 | 22170.55 |
| Clobazam | 650.37 | 1003.24 | 844.70 | 1759.30 | 2430.10 | 2592.67 | 3691.86 | 3420.35 | 3593.09 | 2514.50 | 2150.05 |
| Prazepam | 0.00 | 0.00 | 0.49 | 89.26 | 346.63 | 957.65 | 2249.01 | 6060.50 | 7161.12 | 7712.75 | 6356.37 |
| Alprazolam | 43.08 | 145.02 | 372.41 | 9957.69 | 58908.35 | 74070.89 | 98632.33 | 122756.5 | 122700.5 | 113130.4 | 120134.3 |
| Flurazepam | 59.44 | 28.93 | 7.04 | 1209.87 | 9181.01 | 22337.38 | 35922.94 | 41905.10 | 39326.54 | 33913.00 | 25449.10 |
| Nitrazepam | 112.19 | 116.76 | 217.43 | 184.34 | 426.56 | 449.73 | 1328.90 | 2233.21 | 3930.77 | 4493.56 | 7985.76 |
| Triazolam | 10.72 | 8.45 | 12.78 | 349.84 | 3392.69 | 4520.34 | 6781.07 | 10995.18 | 14337.48 | 13793.56 | 20488.84 |
| Lormetazepam | 0.00 | 1.49 | 49.10 | 67.67 | 370.15 | 800.76 | 2620.15 | 5746.24 | 9428.53 | 13434.98 | 22438.79 |
| Temazepam | 0.00 | 3.11 | 3.08 | 266.45 | 1323.07 | 3666.00 | 8487.43 | 16941.96 | 25413.65 | 27751.78 | 47547.32 |
| Zopiclone | 105.98 | 91.61 | 142.18 | 4001.49 | 24949.54 | 44605.33 | 64603.24 | 86709.65 | 89581.17 | 91561.55 | 117582.2 |
| Zolpidem | 58.94 | 45.59 | 67.92 | 1872.32 | 10148.76 | 19737.98 | 36826.88 | 56780.10 | 66083.68 | 66220.55 | 81196.97 |
| **2017** |  |  |  |  |  |  |  |  |  |  |  |
| Benzodiazepines and z-drugs | 2286.41 | 3830.31 | 4275.54 | 41051.03 | 232895.25 | 341754.4063 | 435218.0938 | 526984.25 | 528207.6875 | 498358.1875 | 548476.625 |
| Long-acting benzodiazepines | 2073.92 | 3422.58 | 3304.38 | 22042.81 | 119226.1563 | 172573.7031 | 198042.9063 | 196278.2813 | 169361.8125 | 139865.1719 | 115006.1641 |
| Clonazepam | 1139.58 | 2043.90 | 1872.36 | 10216.19 | 41499.56 | 47971.06 | 55633.09 | 57994.47 | 38292.54 | 34672.72 | 27790.77 |
| Diazepam | 266.38 | 217.33 | 459.58 | 8499.58 | 64288.61 | 95573.63 | 96817.81 | 83545.29 | 75895.38 | 57739.50 | 46250.90 |
| Chloridiazepoxide | 0.00 | 2.42 | 2.31 | 265.18 | 1722.43 | 3058.84 | 4134.76 | 4459.89 | 3620.96 | 3047.10 | 2643.81 |
| Potassium Clorazepate | 0.00 | 0.00 | 0.00 | 1.07 | 0.38 | 26.88 | 16.59 | 120.36 | 82.58 | 41.01 | 27.14 |
| Lorazepam | 18.58 | 53.62 | 635.99 | 2538.46 | 6832.99 | 7288.80 | 11212.92 | 19858.24 | 20861.13 | 21947.91 | 26438.79 |
| Bromazepam | 0.00 | 0.70 | 17.22 | 342.01 | 1700.01 | 3749.28 | 7495.26 | 16614.99 | 25040.07 | 24242.19 | 20717.96 |
| Clobazam | 507.87 | 981.50 | 673.77 | 1618.78 | 2497.52 | 2449.41 | 3518.96 | 3431.49 | 3575.22 | 2187.17 | 2034.95 |
| Prazepam | 0.00 | 0.29 | 2.94 | 64.41 | 492.12 | 863.81 | 2048.00 | 5054.59 | 6589.54 | 6994.54 | 6083.29 |
| Alprazolam | 32.58 | 132.66 | 147.44 | 10012.96 | 62667.88 | 82005.17 | 97853.13 | 121364.6 | 115791 | 108917.1 | 113410 |
| Flurazepam | 71.33 | 2.12 | 39.72 | 1150.43 | 8338.85 | 22167.14 | 34630.43 | 39649.52 | 37956.12 | 31287.13 | 23179.09 |
| Nitrazepam | 88.75 | 175.03 | 253.68 | 227.18 | 386.69 | 462.94 | 1243.27 | 2022.68 | 3349.46 | 3895.99 | 6996.20 |
| Triazolam | 0.00 | 0.52 | 19.13 | 260.36 | 3174.04 | 4861.56 | 6751.17 | 10578.68 | 13472.97 | 13145.38 | 19194.45 |
| Lormetazepam | 1.04 | 17.96 | 2.11 | 143.34 | 360.99 | 633.32 | 2664.62 | 5513.79 | 8472.61 | 11379.89 | 20233.85 |
| Temazepam | 6.97 | 1.24 | 5.46 | 241.63 | 1443.02 | 3348.41 | 7671.39 | 15604.31 | 22633.73 | 25200.16 | 42114.95 |
| Zopiclone | 127.52 | 142.33 | 95.78 | 3793.55 | 27259.57 | 46880.73 | 66214.24 | 85687.73 | 88004.20 | 88668.14 | 112303.3 |
| Zolpidem | 25.79 | 58.70 | 48.03 | 1675.90 | 10230.58 | 20413.44 | 37312.46 | 55483.67 | 64570.21 | 64992.21 | 79057.16 |
| **2018** |  |  |  |  |  |  |  |  |  |  |  |
| Benzodiazepines and z-drugs | 2944.53 | 2950.13 | 4732.82 | 42192.05 | 246760.7969 | 351749.4375 | 419894.25 | 511949.2813 | 520121.375 | 476684.5 | 530636.625 |
| Long-acting benzodiazepines | 2875.34 | 2727.60 | 3702.92 | 21463.89 | 124531.6094 | 178202.3438 | 192927.3281 | 192655.0469 | 168712.875 | 133980.2031 | 113386.4141 |
| Clonazepam | 2043.76 | 1392.56 | 1924.02 | 10821.93 | 44589.27 | 51283.35 | 54129.57 | 59141.77 | 41702.12 | 34119.63 | 29371.32 |
| Diazepam | 113.55 | 241.90 | 630.55 | 7496.92 | 67376.47 | 98831.17 | 97178.53 | 82415.07 | 73141.79 | 56173.53 | 45174.52 |
| Chloridiazepoxide | 8.98 | 3.12 | 0.00 | 286.50 | 1852.39 | 2886.62 | 3527.73 | 4312.16 | 4095.84 | 2327.79 | 2366.70 |
| Potassium Clorazepate | 0.00 | 0.00 | 0.00 | 0.00 | 2.74 | 3.18 | 18.60 | 140.84 | 114.70 | 39.27 | 34.94 |
| Lorazepam | 12.13 | 67.68 | 587.55 | 3521.50 | 9049.20 | 7697.32 | 11229.57 | 18568.88 | 21870.91 | 20577.24 | 25175.24 |
| Bromazepam | 8.36 | 0.70 | 43.54 | 389.36 | 1515.30 | 3572.22 | 6531.96 | 15266.88 | 23350.77 | 23933.13 | 20344.16 |
| Clobazam | 601.03 | 936.72 | 798.87 | 1432.04 | 2164.53 | 2422.49 | 3240.28 | 3353.07 | 3420.62 | 2276.44 | 1844.66 |
| Prazepam | 0.00 | 0.00 | 0.75 | 45.44 | 437.01 | 967.55 | 1947.46 | 4159.88 | 6515.03 | 6424.15 | 5985.98 |
| Alprazolam | 12.13 | 45.58 | 201.68 | 10820.79 | 68551.05 | 85981.58 | 94952.88 | 117932.1953 | 112897.75 | 102989.0938 | 108187.2969 |
| Flurazepam | 1.08 | 0.26 | 1.94 | 1186.70 | 7722.44 | 21386.78 | 31861.57 | 37351.09 | 36348.07 | 29590.29 | 22335.56 |
| Nitrazepam | 106.94 | 153.04 | 346.79 | 194.36 | 386.76 | 421.21 | 1023.59 | 1781.18 | 3374.72 | 3029.10 | 6272.72 |
| Triazolam | 0.65 | 1.08 | 10.55 | 235.12 | 3267.69 | 5011.97 | 6616.14 | 10129.96 | 12518.83 | 12588.78 | 18410.41 |
| Lormetazepam | 0.00 | 18.62 | 2.16 | 128.54 | 323.22 | 629.88 | 2042.20 | 4975.16 | 7807.27 | 9436.91 | 18812.55 |
| Temazepam | 0.00 | 1.72 | 44.57 | 254.11 | 1433.86 | 3347.45 | 6549.94 | 14169.81 | 19862.19 | 23659.70 | 37773.09 |
| Zopiclone | 28.49 | 65.95 | 113.82 | 3609.95 | 27885.61 | 46769.23 | 63644.16 | 84447.33 | 87546.71 | 85031.51 | 109916.6 |
| Zolpidem | 7.42 | 21.21 | 26.02 | 1768.80 | 10203.25 | 20537.44 | 35400.07 | 53803.99 | 65554.07 | 64487.93 | 78630.84 |
| **2019** |  |  |  |  |  |  |  |  |  |  |  |
| Benzodiazepines and z-drugs | 1624.47 | 3562.02 | 4595.66 | 40132.20 | 243363.9688 | 344571.1563 | 412923.4688 | 503074.625 | 507123.6563 | 461251.2813 | 512423.1875 |
| Long-acting benzodiazepines | 1511.85 | 3389.83 | 3927.23 | 19639.38 | 121393.0234 | 174972.4219 | 189613 | 190796.0313 | 161408.1094 | 129328.2031 | 112085.2656 |
| Clonazepam | 1005.81 | 1761.68 | 2028.97 | 9709.51 | 41827.95 | 49809.22 | 50917.25 | 58820.23 | 38821.19 | 35108.73 | 30522.86 |
| Diazepam | 120.65 | 234.27 | 813.33 | 6812.06 | 68121.31 | 99386.53 | 99488.38 | 83880.10 | 70979.22 | 53552.45 | 44681.39 |
| Chloridiazepoxide | 0.00 | 3.85 | 7.82 | 332.68 | 1985.87 | 3363.19 | 3469.52 | 4093.20 | 3860.33 | 2014.11 | 2176.36 |
| Potassium Clorazepate | 0.00 | 0.00 | 0.00 | 0.00 | 4.35 | 1.17 | 29.00 | 122.64 | 178.88 | 39.73 | 24.74 |
| Lorazepam | 0.00 | 50.13 | 215.13 | 5392.18 | 9422.97 | 8377.04 | 11195.63 | 17681.27 | 22509.73 | 20885.86 | 24698.36 |
| Bromazepam | 3.25 | 0.71 | 12.06 | 401.82 | 1631.87 | 3278.86 | 6182.16 | 13902.57 | 21450.34 | 23182.55 | 19616.09 |
| Clobazam | 260.02 | 1125.96 | 719.73 | 1475.29 | 2122.44 | 2573.41 | 2935.03 | 3012.16 | 3358.70 | 2239.96 | 1701.29 |
| Prazepam | 0.00 | 0.00 | 1.12 | 62.06 | 276.63 | 748.64 | 1709.26 | 3716.44 | 6908.53 | 6025.20 | 5500.91 |
| Alprazolam | 95.47 | 49.88 | 262.07 | 9149.61 | 68761.63 | 84386.80 | 94529.55 | 115205.6 | 114004.6 | 99107.03 | 105356.2 |
| Flurazepam | 0.00 | 3.07 | 47.30 | 960.74 | 6764.07 | 18725.45 | 30076.04 | 35429.99 | 34587.48 | 27427.89 | 21616.10 |
| Nitrazepam | 125.37 | 260.99 | 308.97 | 287.04 | 290.41 | 364.81 | 988.52 | 1721.25 | 2713.77 | 2920.14 | 5861.61 |
| Triazolam | 2.04 | 0.19 | 4.11 | 185.22 | 3034.04 | 4545.20 | 6390.86 | 10705.09 | 11890.55 | 12406.39 | 17664.42 |
| Lormetazepam | 0.00 | 15.50 | 0.00 | 42.36 | 241.25 | 238.24 | 920.61 | 2574.39 | 3834.82 | 5253.51 | 10277.37 |
| Temazepam | 0.33 | 0.00 | 52.61 | 286.14 | 1463.01 | 3064.73 | 5875.22 | 13407.60 | 17889.73 | 21948.22 | 35088.11 |
| Zopiclone | 10.33 | 34.50 | 78.37 | 3355.01 | 27173.44 | 46049.73 | 64205.56 | 84700.44 | 89242.34 | 83840.15 | 108887.9 |
| Zolpidem | 1.20 | 21.29 | 44.09 | 1680.47 | 10242.73 | 19658.13 | 34010.87 | 54101.60 | 64893.42 | 65299.37 | 78749.51 |
| **2020** |  |  |  |  |  |  |  |  |  |  |  |
| Benzodiazepines and z-drugs | 1662.61 | 3262.13 | 4501.21 | 34444.02 | 215787.8 | 325376.1 | 401345 | 487021.8 | 491787.7 | 446646 | 494590.9 |
| Long-acting benzodiazepines | 1586.29 | 3113.44 | 3859.31 | 17261.12 | 107208.5 | 163911 | 184533.5 | 187974.5 | 155722 | 126345 | 109691.5 |
| Clonazepam | 912.33 | 1602.50 | 2254.46 | 8313.24 | 35765.52 | 46855.73 | 49305.52 | 58896.60 | 44140.10 | 35191.07 | 32316.61 |
| Diazepam | 246.49 | 148.42 | 660.60 | 6296.15 | 61840.94 | 95578.36 | 99228.40 | 84791.47 | 64991.48 | 53215.78 | 43027.59 |
| Chloridiazepoxide | 4.90 | 6.42 | 4.55 | 239.93 | 1568.23 | 3055.77 | 3180.57 | 3831.99 | 2924.63 | 2102.87 | 1930.60 |
| Potassium Clorazepate | 0.00 | 0.00 | 0.00 | 0.00 | 12.27 | 2.24 | 6.18 | 118.01 | 180.56 | 40.01 | 26.76 |
| Lorazepam | 0.85 | 90.89 | 287.20 | 5508.62 | 9712.68 | 8900.60 | 11603.87 | 17437.44 | 22136.41 | 20664.04 | 24667.71 |
| Bromazepam | 1.27 | 0.84 | 5.18 | 287.87 | 1704.14 | 3026.52 | 5335.23 | 12930.44 | 20554.83 | 21788.26 | 19001.47 |
| Clobazam | 365.49 | 1108.99 | 519.79 | 1308.31 | 1950.62 | 2498.63 | 2869.47 | 2868.21 | 3132.36 | 2226.11 | 1766.63 |
| Prazepam | 0.00 | 0.61 | 0.71 | 39.11 | 208.02 | 494.45 | 1618.20 | 3085.41 | 5630.59 | 5248.54 | 4724.04 |
| Alprazolam | 47.40 | 28.65 | 151.67 | 7082.40 | 59702.62 | 78806.52 | 91409.15 | 109263.6 | 111807.1 | 97228.97 | 102334.6 |
| Flurazepam | 0.00 | 0.26 | 44.33 | 740.85 | 5527.39 | 15106.69 | 27462.84 | 32700.47 | 32432.41 | 25561.10 | 20592.28 |
| Nitrazepam | 57.08 | 246.24 | 374.88 | 323.54 | 335.53 | 319.19 | 862.30 | 1682.32 | 2289.93 | 2759.52 | 5306.94 |
| Triazolam | 0.00 | 0.00 | 7.81 | 157.46 | 2440.46 | 4268.06 | 6249.58 | 10338.92 | 10979.76 | 12618.52 | 17004.92 |
| Lormetazepam | 0.00 | 0.00 | 9.52 | 1.04 | 9.73 | 7.75 | 56.89 | 227.88 | 396.92 | 387.63 | 811.90 |
| Temazepam | 0.00 | 0.37 | 47.75 | 165.58 | 1502.65 | 2947.69 | 5541.37 | 12307.70 | 16265.36 | 19908.75 | 32707.37 |
| Zopiclone | 14.12 | 20.35 | 73.74 | 2759.42 | 24454.30 | 44429.03 | 63151.52 | 82523.85 | 89723.48 | 82047.94 | 108682.4 |
| Zolpidem | 12.67 | 7.59 | 59.04 | 1220.52 | 9052.71 | 19078.89 | 33463.89 | 54017.41 | 64201.79 | 65656.86 | 79689.08 |
| **2021** |  |  |  |  |  |  |  |  |  |  |  |
| Benzodiazepines and z-drugs | 2905.46 | 4008.07 | 3657.71 | 35369.82 | 210550.1 | 326336 | 396055.5 | 473370.2 | 479468.8 | 429380 | 475576.6 |
| Long-acting benzodiazepines | 2873.10 | 3814.92 | 3079.97 | 19067.84 | 108035.5 | 163377.3 | 185138.1 | 184806.8 | 154732.9 | 119532.6 | 104949.9 |
| Clonazepam | 2202.82 | 2228.54 | 1857.38 | 8889.91 | 38146.90 | 47511.89 | 50548.64 | 57788.18 | 47171.98 | 34160.84 | 34340.97 |
| Diazepam | 221.70 | 255.76 | 380.88 | 7156.60 | 60709.76 | 95227.96 | 100783 | 86559.24 | 64672.14 | 51492.09 | 41262.86 |
| Chloridiazepoxide | 0.51 | 3.40 | 1.86 | 421.80 | 1557.82 | 3371.92 | 3101.96 | 3825.30 | 2640.91 | 1848.14 | 1677.42 |
| Potassium Clorazepate | 0.00 | 0.00 | 0.00 | 0.00 | 36.49 | 0.00 | 5.35 | 56.91 | 81.98 | 107.57 | 36.14 |
| Lorazepam | 0.00 | 118.51 | 330.74 | 6372.70 | 10368.53 | 9899.94 | 11683.49 | 17166.90 | 22440.22 | 18688.06 | 24183.43 |
| Bromazepam | 0.00 | 0.35 | 18.30 | 236.38 | 1830.28 | 2874.09 | 4598.89 | 11742.89 | 19313.11 | 19910.36 | 18449.10 |
| Clobazam | 382.48 | 1151.95 | 700.80 | 1561.78 | 2009.31 | 2433.30 | 2909.54 | 3080.97 | 2838.10 | 2166.73 | 1624.39 |
| Prazepam | 0.00 | 0.09 | 0.00 | 80.36 | 220.18 | 562.60 | 1459.18 | 2579.54 | 4930.96 | 5033.01 | 4571.38 |
| Alprazolam | 3.92 | 49.28 | 110.82 | 5581.69 | 53904.04 | 80247.18 | 88804.09 | 104521.1 | 107056.7 | 94756.52 | 97708.52 |
| Flurazepam | 0.00 | 12.71 | 0.00 | 783.63 | 5238.35 | 14159.27 | 26044.57 | 30373.85 | 31656.92 | 23780.37 | 19591.57 |
| Nitrazepam | 65.60 | 162.47 | 139.05 | 173.76 | 116.71 | 110.37 | 285.83 | 542.76 | 739.92 | 943.81 | 1845.20 |
| Triazolam | 0.00 | 0.00 | 1.41 | 174.28 | 2057.76 | 4186.58 | 6073.84 | 9445.86 | 10879.54 | 12065.80 | 16172.19 |
| Lormetazepam | 0.00 | 0.00 | 0.00 | 0.00 | 0.00 | 0.84 | 3.58 | 8.64 | 39.89 | 52.43 | 67.48 |
| Temazepam | 0.00 | 0.00 | 37.16 | 107.03 | 1310.95 | 2736.85 | 5255.67 | 11230.05 | 15018.00 | 18239.35 | 29960.89 |
| Zopiclone | 23.41 | 10.23 | 52.06 | 2611.05 | 23856.53 | 44415.85 | 61846.09 | 82035.53 | 87089.69 | 81321.59 | 105911.6 |
| Zolpidem | 5.03 | 14.77 | 27.25 | 1218.84 | 9186.55 | 18597.31 | 32651.82 | 52412.43 | 62898.75 | 64813.34 | 78173.48 |
| **2022** |  |  |  |  |  |  |  |  |  |  |  |
| Benzodiazepines and z-drugs | 1754.92 | 3199.67 | 4655.90 | 33558.71 | 183004.3 | 301543.1 | 373010.3 | 441034.5 | 448955.1 | 406105.4 | 449008.8 |
| Long-acting benzodiazepines | 1730.93 | 2920.07 | 3980.87 | 18008.79 | 91870.52 | 147199.1 | 171870.3 | 170656.9 | 144432.4 | 111426.5 | 95801.84 |
| Clonazepam | 1122.82 | 1748.47 | 2594.05 | 7714.28 | 30207.25 | 39285.82 | 43684.91 | 49359.45 | 44775.92 | 30498.61 | 30542.58 |
| Diazepam | 179.85 | 180.84 | 408.81 | 7209.49 | 53311.61 | 90738.95 | 98798.46 | 84879.08 | 63894.36 | 50903.77 | 40679.41 |
| Chloridiazepoxide | 0.00 | 0.78 | 0.81 | 428.01 | 1551.91 | 2879.35 | 2769.63 | 3330.73 | 2400.06 | 2003.70 | 1448.83 |
| Potassium Clorazepate | 0.00 | 0.00 | 0.00 | 0.00 | 7.24 | 3.76 | 0.00 | 83.31 | 88.76 | 63.35 | 50.04 |
| Lorazepam | 11.48 | 223.57 | 416.34 | 6412.32 | 10279.12 | 10439.28 | 12386.41 | 15772.47 | 20637.24 | 18222.05 | 23320.23 |
| Bromazepam | 1.38 | 0.00 | 20.79 | 230.81 | 1493.81 | 2986.52 | 4185.22 | 10292.86 | 17811.00 | 18466.81 | 17655.71 |
| Clobazam | 412.31 | 934.09 | 962.47 | 1837.06 | 2351.61 | 2314.72 | 2816.46 | 3356.61 | 2622.19 | 2103.54 | 1660.40 |
| Prazepam | 0.00 | 0.00 | 0.00 | 23.40 | 105.32 | 246.56 | 849.20 | 1282.86 | 2733.45 | 2876.79 | 3015.50 |
| Alprazolam | 4.62 | 29.62 | 147.04 | 4927.54 | 48476.01 | 76854.95 | 84608.51 | 99066.89 | 99486.66 | 89940.95 | 92113.06 |
| Flurazepam | 0.00 | 2.19 | 9.91 | 782.62 | 4321.10 | 11705.64 | 22918.43 | 28329.63 | 27868.92 | 22944.32 | 18231.38 |
| Nitrazepam | 15.95 | 53.69 | 4.82 | 13.94 | 14.48 | 24.29 | 33.22 | 35.24 | 48.75 | 32.41 | 173.70 |
| Triazolam | 0.00 | 0.00 | 14.75 | 110.01 | 1552.71 | 3927.28 | 5793.19 | 8707.53 | 9834.30 | 11249.03 | 15286.87 |
| Lormetazepam | 0.00 | 0.00 | 0.00 | 0.00 | 1.73 | 0.00 | 1.22 | 3.31 | 0.00 | 24.04 | 20.00 |
| Temazepam | 0.00 | 0.13 | 0.69 | 149.68 | 1014.87 | 2273.56 | 4401.98 | 9677.69 | 13099.88 | 16511.36 | 27314.37 |
| Zopiclone | 2.76 | 20.38 | 44.74 | 2510.93 | 20726.06 | 41289.14 | 58529.89 | 77855.71 | 82930.93 | 78645.88 | 101629.4 |
| Zolpidem | 3.75 | 5.90 | 30.66 | 1208.62 | 7589.43 | 16573.29 | 31233.60 | 49001.10 | 60722.68 | 61618.82 | 75867.28 |

**Rate of DDEs per 1,000 GMS population by sex**

|  | **Females** | **Males** |
| --- | --- | --- |
| **2014** |  |  |
| Benzodiazepines and z-drugs | 355201.1 | 248792.3 |
| Long-acting benzodiazepines | 111397.9 | 105296 |
| Clonazepam | 26949.38 | 27027.21 |
| Diazepam | 50328.71 | 48922.06 |
| Chloridiazepoxide | 1932.819 | 2977.944 |
| Potassium Clorazepate | 37.89036 | 19.50631 |
| Lorazepam | 12937.3 | 9904.99 |
| Bromazepam | 13979.85 | 5275.016 |
| Clobazam | 2797.277 | 2744.136 |
| Prazepam | 3948.68 | 1783.753 |
| Alprazolam | 83094.26 | 50953.35 |
| Flurazepam | 22088.69 | 20359.9 |
| Nitrazepam | 3314.492 | 1461.541 |
| Triazolam | 8803.584 | 5378.465 |
| Lormetazepam | 8337.621 | 3259.628 |
| Temazepam | 18878.94 | 9972.212 |
| Zopiclone | 57604.23 | 38508.27 |
| Zolpidem | 40167.4 | 20244.27 |
| **2015** |  |  |
| Benzodiazepines and z-drugs | 350709.5 | 247115.5 |
| Long-acting benzodiazepines | 109428 | 105129.9 |
| Clonazepam | 27060.78 | 27692.08 |
| Diazepam | 50399.14 | 49565.57 |
| Chloridiazepoxide | 1826.749 | 2827.48 |
| Potassium Clorazepate | 36.42591 | 19.61422 |
| Lorazepam | 12541.63 | 9795.516 |
| Bromazepam | 13644.05 | 5067.069 |
| Clobazam | 2395.681 | 2495.835 |
| Prazepam | 3792.579 | 1763.064 |
| Alprazolam | 82681.73 | 50317.81 |
| Flurazepam | 20918.08 | 19484.46 |
| Nitrazepam | 2998.592 | 1281.837 |
| Triazolam | 8780.536 | 5448.788 |
| Lormetazepam | 7778.052 | 3015.499 |
| Temazepam | 17242.17 | 9232.582 |
| Zopiclone | 58195.52 | 38966.1 |
| Zolpidem | 40417.82 | 20142.23 |
| **2016** |  |  |
| Benzodiazepines and z-drugs | 357039.3 | 256586.3 |
| Long-acting benzodiazepines | 110856.2 | 109346 |
| Clonazepam | 28407.64 | 30225.31 |
| Diazepam | 51424.68 | 51436.46 |
| Chloridiazepoxide | 1824.144 | 2943.674 |
| Potassium Clorazepate | 34.30355 | 17.74157 |
| Lorazepam | 12654.58 | 10270.1 |
| Bromazepam | 13521.17 | 5102.998 |
| Clobazam | 2270.528 | 2377.63 |
| Prazepam | 3708.631 | 1747.268 |
| Alprazolam | 84553.33 | 52554.25 |
| Flurazepam | 20309.95 | 19357.2 |
| Nitrazepam | 2876.345 | 1240.702 |
| Triazolam | 8770.746 | 5530.395 |
| Lormetazepam | 7642.078 | 2960.437 |
| Temazepam | 16458.56 | 8902.026 |
| Zopiclone | 60085.59 | 40910.46 |
| Zolpidem | 42497.01 | 21009.6 |
| **2017** |  |  |
| Benzodiazepines and z-drugs | 358277.3 | 262360.3 |
| Long-acting benzodiazepines | 111738.1 | 112523.7 |
| Clonazepam | 30684.65 | 32326.27 |
| Diazepam | 51437.63 | 53113.71 |
| Chloridiazepoxide | 1725.708 | 2895.926 |
| Potassium Clorazepate | 36.44852 | 21.39583 |
| Lorazepam | 12480.93 | 10695.87 |
| Bromazepam | 13198.02 | 5013.441 |
| Clobazam | 2156.445 | 2314.791 |
| Prazepam | 3568.498 | 1644.262 |
| Alprazolam | 84894.59 | 53682.02 |
| Flurazepam | 19475.58 | 19001.28 |
| Nitrazepam | 2653.164 | 1206.102 |
| Triazolam | 8692.913 | 5530.715 |
| Lormetazepam | 7344.407 | 2779.12 |
| Temazepam | 15555.87 | 8407.759 |
| Zopiclone | 60987.79 | 42148.7 |
| Zolpidem | 43384.64 | 21579.01 |
| **2018** |  |  |
| Benzodiazepines and z-drugs | 357864.2 | 264040.4 |
| Long-acting benzodiazepines | 111789.8 | 113999.8 |
| Clonazepam | 31837.95 | 33771.53 |
| Diazepam | 51461.7 | 54111.49 |
| Chloridiazepoxide | 1583.107 | 2811.128 |
| Potassium Clorazepate | 41.15924 | 24.13953 |
| Lorazepam | 12757.43 | 10894.83 |
| Bromazepam | 13010.32 | 4867.033 |
| Clobazam | 2053.57 | 2231.086 |
| Prazepam | 3450.603 | 1616.111 |
| Alprazolam | 84929.39 | 53947.36 |
| Flurazepam | 18930.15 | 18317.35 |
| Nitrazepam | 2431.522 | 1116.944 |
| Triazolam | 8634.927 | 5483.26 |
| Lormetazepam | 6917.401 | 2547.296 |
| Temazepam | 14593.03 | 7839.966 |
| Zopiclone | 61225.72 | 42408.38 |
| Zolpidem | 44006.23 | 22052.51 |
| **2019** |  |  |
| Benzodiazepines and z-drugs | 352468.6 | 262238.8 |
| Long-acting benzodiazepines | 109840.8 | 113173.2 |
| Clonazepam | 31088.63 | 33304.32 |
| Diazepam | 51558.52 | 54897.97 |
| Chloridiazepoxide | 1546.539 | 2833.232 |
| Potassium Clorazepate | 42.9638 | 26.4483 |
| Lorazepam | 12777.33 | 11499.31 |
| Bromazepam | 12628.82 | 4615.571 |
| Clobazam | 1944.494 | 2185.514 |
| Prazepam | 3255.655 | 1537.26 |
| Alprazolam | 84331.16 | 53486.66 |
| Flurazepam | 18113.14 | 17276.38 |
| Nitrazepam | 2290.873 | 1112.034 |
| Triazolam | 8626.72 | 5352.055 |
| Lormetazepam | 3835.184 | 1333.997 |
| Temazepam | 13955.12 | 7327.841 |
| Zopiclone | 61810.6 | 43176.21 |
| Zolpidem | 44662.84 | 22273.96 |
| **2020** |  |  |
| Benzodiazepines and z-drugs | 337605.7 | 250566.8 |
| Long-acting benzodiazepines | 105192.7 | 108655.5 |
| Clonazepam | 30671.56 | 32680.36 |
| Diazepam | 49622.93 | 53645.59 |
| Chloridiazepoxide | 1375.627 | 2539.702 |
| Potassium Clorazepate | 41.47066 | 22.95545 |
| Lorazepam | 12978.67 | 11422.13 |
| Bromazepam | 11866.17 | 4339.429 |
| Clobazam | 1908.397 | 2069.977 |
| Prazepam | 2740.007 | 1305.52 |
| Alprazolam | 80934.21 | 50020.43 |
| Flurazepam | 16790.25 | 15299.24 |
| Nitrazepam | 2042.422 | 1092.172 |
| Triazolam | 8189.292 | 5229.555 |
| Lormetazepam | 302.9777 | 110.7963 |
| Temazepam | 12834.82 | 6847.953 |
| Zopiclone | 60843.21 | 41963.57 |
| Zolpidem | 44463.73 | 21977.43 |
| **2021** |  |  |
| Benzodiazepines and z-drugs | 333065.8 | 251968.5 |
| Long-acting benzodiazepines | 103873.3 | 110147.6 |
| Clonazepam | 31732.92 | 34094.11 |
| Diazepam | 49467.95 | 54778.3 |
| Chloridiazepoxide | 1337.235 | 2561.219 |
| Potassium Clorazepate | 40.24144 | 15.75089 |
| Lorazepam | 13165.26 | 11722.52 |
| Bromazepam | 11424.72 | 4163.052 |
| Clobazam | 1898.638 | 2147.057 |
| Prazepam | 2597.791 | 1275.524 |
| Alprazolam | 78900.16 | 49515.38 |
| Flurazepam | 16076.12 | 14863.23 |
| Nitrazepam | 722.4268 | 412.4052 |
| Triazolam | 7970.614 | 5149.142 |
| Lormetazepam | 26.44139 | 10.43348 |
| Temazepam | 12136.18 | 6548.344 |
| Zopiclone | 60928.69 | 42529.56 |
| Zolpidem | 44640.43 | 22182.48 |
| **2022** |  |  |
| Benzodiazepines and z-drugs | 310875.1 | 238295.2 |
| Long-acting benzodiazepines | 94797.05 | 102067.2 |
| Clonazepam | 27698.22 | 29445.81 |
| Diazepam | 47900.52 | 53508.63 |
| Chloridiazepoxide | 1189.782 | 2345.956 |
| Potassium Clorazepate | 39.7938 | 16.28256 |
| Lorazepam | 12782.61 | 11650.07 |
| Bromazepam | 10575.86 | 3967.327 |
| Clobazam | 1969.661 | 2157.964 |
| Prazepam | 1517.325 | 762.7554 |
| Alprazolam | 73954.23 | 47727.28 |
| Flurazepam | 14427.47 | 13777.16 |
| Nitrazepam | 54.27434 | 52.64322 |
| Triazolam | 7440.688 | 4834.991 |
| Lormetazepam | 10.1132 | 0.726847 |
| Temazepam | 10835.12 | 5920.788 |
| Zopiclone | 57963.09 | 40810.3 |
| Zolpidem | 42516.32 | 21316.56 |

^1^doxepin, trazodone, mirtazapine

^2^promethazine, cyclizine, ketotifen

^3^olanzapine, quetiapine, risperidone

**Supplementary Table 10. Prevalence of any dispensings, initiations (90/180 days), discontinuations (90/180 days), and chronic use (30/90 days) in the GMS population**

**Prevalence of any dispensings in the GMS population**

|  | **2014** | **2015** | **2016** | **2017** | **2018** | **2019** | **2020** | **2021** | **2022** |
| --- | --- | --- | --- | --- | --- | --- | --- | --- | --- |
| Benzodiazepines and z-drugs | 0.182 | 0.178 | 0.183 | 0.184 | 0.183 | 0.181 | 0.165 | 0.169 | 0.164 |
| Long-acting benzodiazepines | 0.072 | 0.070 | 0.072 | 0.072 | 0.072 | 0.071 | 0.064 | 0.066 | 0.063 |
| Clonazepam | 0.005 | 0.006 | 0.006 | 0.006 | 0.007 | 0.007 | 0.006 | 0.007 | 0.006 |
| Diazepam | 0.053 | 0.053 | 0.055 | 0.055 | 0.054 | 0.054 | 0.048 | 0.050 | 0.051 |
| Chloridiazepoxide | 0.005 | 0.005 | 0.005 | 0.005 | 0.005 | 0.005 | 0.005 | 0.005 | 0.004 |
| Potassium Clorazepate | 0.000 | 0.000 | 0.000 | 0.000 | 0.000 | 0.000 | 0.000 | 0.000 | 0.000 |
| Lorazepam | 0.004 | 0.004 | 0.004 | 0.005 | 0.005 | 0.005 | 0.005 | 0.005 | 0.005 |
| Bromazepam | 0.008 | 0.007 | 0.008 | 0.007 | 0.007 | 0.007 | 0.006 | 0.006 | 0.006 |
| Clobazam | 0.002 | 0.001 | 0.001 | 0.001 | 0.001 | 0.001 | 0.001 | 0.001 | 0.001 |
| Prazepam | 0.002 | 0.001 | 0.001 | 0.001 | 0.001 | 0.001 | 0.001 | 0.001 | 0.001 |
| Alprazolam | 0.045 | 0.045 | 0.047 | 0.048 | 0.048 | 0.048 | 0.042 | 0.043 | 0.044 |
| Flurazepam | 0.009 | 0.008 | 0.008 | 0.008 | 0.007 | 0.007 | 0.006 | 0.006 | 0.005 |
| Nitrazepam | 0.002 | 0.001 | 0.001 | 0.001 | 0.001 | 0.001 | 0.001 | 0.001 | 0.000 |
| Triazolam | 0.006 | 0.005 | 0.005 | 0.005 | 0.005 | 0.005 | 0.005 | 0.005 | 0.004 |
| Lormetazepam | 0.004 | 0.004 | 0.004 | 0.004 | 0.003 | 0.003 | 0.000 | 0.000 | 0.000 |
| Temazepam | 0.009 | 0.008 | 0.008 | 0.007 | 0.007 | 0.006 | 0.005 | 0.005 | 0.005 |
| Zopiclone | 0.049 | 0.048 | 0.049 | 0.050 | 0.049 | 0.049 | 0.046 | 0.046 | 0.044 |
| Zolpidem | 0.043 | 0.042 | 0.043 | 0.043 | 0.043 | 0.043 | 0.040 | 0.040 | 0.038 |
| Antidepressants^1^ | 0.015 | 0.017 | 0.019 | 0.021 | 0.023 | 0.024 | 0.025 | 0.027 | 0.027 |
| Antihistamines^2^ | 0.003 | 0.003 | 0.005 | 0.006 | 0.007 | 0.009 | 0.011 | 0.013 | 0.014 |
| Antipsychotics^3^ | 0.016 | 0.017 | 0.019 | 0.021 | 0.023 | 0.025 | 0.026 | 0.028 | 0.028 |
| Sedatives | 0.188 | 0.186 | 0.193 | 0.197 | 0.199 | 0.198 | 0.184 | 0.190 | 0.186 |

**Prevalence of initiations after 90 and 180 days in the GMS population**

|  | **2014** | **2015** | **2016** | **2017** | **2018** | **2019** | **2020** | **2021** | **2022** |
| --- | --- | --- | --- | --- | --- | --- | --- | --- | --- |
| **90 days** |  |  |  |  |  |  |  |  |  |
| Benzodiazepines and z-drugs | 0.025 | 0.063 | 0.072 | 0.073 | 0.074 | 0.073 | 0.066 | 0.068 | 0.066 |
| Long-acting benzodiazepines | 0.054 | 0.082 | 0.089 | 0.090 | 0.089 | 0.088 | 0.077 | 0.078 | 0.077 |
| Clonazepam | 0.025 | 0.063 | 0.071 | 0.072 | 0.071 | 0.071 | 0.063 | 0.064 | 0.064 |
| Diazepam | 0.026 | 0.040 | 0.044 | 0.044 | 0.044 | 0.043 | 0.038 | 0.040 | 0.038 |
| Chloridiazepoxide | 0.013 | 0.033 | 0.037 | 0.038 | 0.038 | 0.037 | 0.033 | 0.035 | 0.034 |
| Potassium Clorazepate | 0.002 | 0.002 | 0.003 | 0.003 | 0.003 | 0.003 | 0.003 | 0.003 | 0.002 |
| Lorazepam | 0.001 | 0.002 | 0.002 | 0.002 | 0.002 | 0.002 | 0.002 | 0.002 | 0.002 |
| Bromazepam | 0.021 | 0.033 | 0.036 | 0.036 | 0.036 | 0.036 | 0.032 | 0.033 | 0.034 |
| Clobazam | 0.011 | 0.028 | 0.032 | 0.032 | 0.032 | 0.032 | 0.028 | 0.029 | 0.031 |
| Prazepam | 0.003 | 0.004 | 0.004 | 0.004 | 0.004 | 0.004 | 0.004 | 0.004 | 0.004 |
| Alprazolam | 0.001 | 0.003 | 0.004 | 0.004 | 0.004 | 0.004 | 0.003 | 0.004 | 0.003 |
| Flurazepam | 0.000 | 0.000 | 0.000 | 0.000 | 0.000 | 0.000 | 0.000 | 0.000 | 0.000 |
| Nitrazepam | 0.000 | 0.000 | 0.000 | 0.000 | 0.000 | 0.000 | 0.000 | 0.000 | 0.000 |
| Triazolam | 0.002 | 0.003 | 0.003 | 0.003 | 0.003 | 0.003 | 0.003 | 0.003 | 0.003 |
| Lormetazepam | 0.001 | 0.002 | 0.002 | 0.002 | 0.002 | 0.002 | 0.003 | 0.003 | 0.003 |
| Temazepam | 0.002 | 0.003 | 0.003 | 0.003 | 0.003 | 0.003 | 0.002 | 0.002 | 0.002 |
| Zopiclone | 0.001 | 0.003 | 0.003 | 0.003 | 0.003 | 0.002 | 0.002 | 0.002 | 0.002 |
| Zolpidem | 0.000 | 0.001 | 0.001 | 0.001 | 0.001 | 0.001 | 0.000 | 0.000 | 0.001 |
| Antidepressants^1^ | 0.000 | 0.000 | 0.000 | 0.000 | 0.000 | 0.000 | 0.000 | 0.000 | 0.001 |
| Antihistamines^2^ | 0.000 | 0.001 | 0.001 | 0.001 | 0.000 | 0.000 | 0.000 | 0.000 | 0.000 |
| Antipsychotics^3^ | 0.000 | 0.000 | 0.000 | 0.000 | 0.000 | 0.000 | 0.000 | 0.000 | 0.000 |
| Sedatives | 0.055 | 0.084 | 0.091 | 0.093 | 0.093 | 0.092 | 0.082 | 0.084 | 0.082 |
| **180 days** |  |  |  |  |  |  |  |  |  |
| Benzodiazepines and z-drugs | 0.054 | 0.082 | 0.089 | 0.090 | 0.089 | 0.088 | 0.077 | 0.078 | 0.077 |
| Long-acting benzodiazepines | 0.025 | 0.063 | 0.071 | 0.072 | 0.071 | 0.071 | 0.063 | 0.064 | 0.064 |
| Clonazepam | 0.001 | 0.002 | 0.002 | 0.002 | 0.002 | 0.002 | 0.002 | 0.002 | 0.002 |
| Diazepam | 0.021 | 0.033 | 0.036 | 0.036 | 0.036 | 0.036 | 0.032 | 0.033 | 0.034 |
| Chloridiazepoxide | 0.011 | 0.028 | 0.032 | 0.032 | 0.032 | 0.032 | 0.028 | 0.029 | 0.031 |
| Potassium Clorazepate | 0.003 | 0.004 | 0.004 | 0.004 | 0.004 | 0.004 | 0.004 | 0.004 | 0.004 |
| Lorazepam | 0.001 | 0.003 | 0.004 | 0.004 | 0.004 | 0.004 | 0.003 | 0.004 | 0.003 |
| Bromazepam | 0.000 | 0.000 | 0.000 | 0.000 | 0.000 | 0.000 | 0.000 | 0.000 | 0.000 |
| Clobazam | 0.000 | 0.000 | 0.000 | 0.000 | 0.000 | 0.000 | 0.000 | 0.000 | 0.000 |
| Prazepam | 0.002 | 0.003 | 0.003 | 0.003 | 0.003 | 0.003 | 0.003 | 0.003 | 0.003 |
| Alprazolam | 0.001 | 0.002 | 0.002 | 0.002 | 0.002 | 0.002 | 0.003 | 0.003 | 0.003 |
| Flurazepam | 0.002 | 0.003 | 0.003 | 0.003 | 0.003 | 0.003 | 0.002 | 0.002 | 0.002 |
| Nitrazepam | 0.001 | 0.003 | 0.003 | 0.003 | 0.003 | 0.002 | 0.002 | 0.002 | 0.002 |
| Triazolam | 0.000 | 0.001 | 0.001 | 0.001 | 0.001 | 0.001 | 0.000 | 0.000 | 0.001 |
| Lormetazepam | 0.000 | 0.000 | 0.000 | 0.000 | 0.000 | 0.000 | 0.000 | 0.000 | 0.001 |
| Temazepam | 0.000 | 0.001 | 0.001 | 0.001 | 0.000 | 0.000 | 0.000 | 0.000 | 0.000 |
| Zopiclone | 0.000 | 0.000 | 0.000 | 0.000 | 0.000 | 0.000 | 0.000 | 0.000 | 0.000 |
| Zolpidem | 0.018 | 0.027 | 0.030 | 0.030 | 0.030 | 0.030 | 0.026 | 0.026 | 0.028 |
| Antidepressants^1^ | 0.004 | 0.009 | 0.010 | 0.010 | 0.011 | 0.012 | 0.012 | 0.013 | 0.012 |
| Antihistamines^2^ | 0.001 | 0.002 | 0.003 | 0.005 | 0.005 | 0.001 | 0.008 | 0.010 | 0.010 |
| Antipsychotics^3^ | 0.001 | 0.002 | 0.003 | 0.004 | 0.005 | 0.007 | 0.007 | 0.009 | 0.009 |
| Sedatives | 0.025 | 0.063 | 0.072 | 0.073 | 0.074 | 0.073 | 0.066 | 0.068 | 0.066 |

**Prevalence of discontinuations after 90 and 180 days in the GMS population**

|  | **2014** | **2015** | **2016** | **2017** | **2018** | **2019** | **2020** | **2021** | **2022** |
| --- | --- | --- | --- | --- | --- | --- | --- | --- | --- |
| **90 days** |  |  |  |  |  |  |  |  |  |
| Sedatives | 0.043 | 0.050 | 0.053 | 0.053 | 0.052 | 0.053 | 0.046 | 0.047 | 0.047 |
| Benzodiazepines and z-drugs | 0.020 | 0.029 | 0.032 | 0.032 | 0.032 | 0.032 | 0.028 | 0.029 | 0.029 |
| Long-acting benzodiazepines | 0.044 | 0.052 | 0.055 | 0.056 | 0.055 | 0.056 | 0.049 | 0.050 | 0.050 |
| Clonazepam | 0.021 | 0.032 | 0.035 | 0.035 | 0.035 | 0.036 | 0.033 | 0.033 | 0.033 |
| Diazepam | 0.024 | 0.030 | 0.032 | 0.032 | 0.031 | 0.032 | 0.028 | 0.029 | 0.029 |
| Chloridiazepoxide | 0.012 | 0.022 | 0.023 | 0.024 | 0.023 | 0.023 | 0.022 | 0.022 | 0.021 |
| Potassium Clorazepate | 0.001 | 0.002 | 0.002 | 0.002 | 0.002 | 0.002 | 0.002 | 0.002 | 0.003 |
| Lorazepam | 0.001 | 0.001 | 0.001 | 0.002 | 0.002 | 0.002 | 0.001 | 0.002 | 0.002 |
| Bromazepam | 0.019 | 0.025 | 0.026 | 0.027 | 0.026 | 0.027 | 0.024 | 0.024 | 0.024 |
| Clobazam | 0.010 | 0.018 | 0.020 | 0.020 | 0.020 | 0.020 | 0.018 | 0.019 | 0.018 |
| Prazepam | 0.002 | 0.003 | 0.003 | 0.003 | 0.003 | 0.003 | 0.003 | 0.003 | 0.003 |
| Alprazolam | 0.001 | 0.002 | 0.003 | 0.003 | 0.003 | 0.003 | 0.003 | 0.002 | 0.002 |
| Flurazepam | 0.000 | 0.000 | 0.000 | 0.000 | 0.000 | 0.000 | 0.000 | 0.000 | 0.000 |
| Nitrazepam | 0.000 | 0.000 | 0.000 | 0.000 | 0.000 | 0.000 | 0.000 | 0.000 | 0.000 |
| Triazolam | 0.002 | 0.002 | 0.002 | 0.002 | 0.002 | 0.002 | 0.002 | 0.002 | 0.002 |
| Lormetazepam | 0.001 | 0.002 | 0.002 | 0.002 | 0.002 | 0.002 | 0.002 | 0.002 | 0.002 |
| Temazepam | 0.002 | 0.003 | 0.003 | 0.003 | 0.003 | 0.002 | 0.002 | 0.002 | 0.002 |
| Zopiclone | 0.001 | 0.002 | 0.002 | 0.002 | 0.002 | 0.002 | 0.002 | 0.001 | 0.001 |
| Zolpidem | 0.000 | 0.000 | 0.000 | 0.000 | 0.000 | 0.000 | 0.000 | 0.000 | 0.000 |
| Antidepressants^1^ | 0.000 | 0.000 | 0.000 | 0.000 | 0.000 | 0.000 | 0.000 | 0.000 | 0.000 |
| Antihistamines^2^ | 0.000 | 0.001 | 0.000 | 0.000 | 0.000 | 0.000 | 0.000 | 0.000 | 0.001 |
| Antipsychotics^3^ | 0.000 | 0.000 | 0.000 | 0.000 | 0.000 | 0.000 | 0.000 | 0.000 | 0.000 |
| **180 days** |  |  |  |  |  |  |  |  |  |
| Sedatives | 0.020 | 0.029 | 0.032 | 0.032 | 0.032 | 0.032 | 0.028 | 0.029 | 0.029 |
| Benzodiazepines and z-drugs | 0.044 | 0.052 | 0.055 | 0.056 | 0.055 | 0.056 | 0.049 | 0.050 | 0.050 |
| Long-acting benzodiazepines | 0.021 | 0.032 | 0.035 | 0.035 | 0.035 | 0.036 | 0.033 | 0.033 | 0.033 |
| Clonazepam | 0.024 | 0.030 | 0.032 | 0.032 | 0.031 | 0.032 | 0.028 | 0.029 | 0.029 |
| Diazepam | 0.012 | 0.022 | 0.023 | 0.024 | 0.023 | 0.023 | 0.022 | 0.022 | 0.021 |
| Chloridiazepoxide | 0.001 | 0.002 | 0.002 | 0.002 | 0.002 | 0.002 | 0.002 | 0.002 | 0.003 |
| Potassium Clorazepate | 0.001 | 0.001 | 0.001 | 0.002 | 0.002 | 0.002 | 0.001 | 0.002 | 0.002 |
| Lorazepam | 0.019 | 0.025 | 0.026 | 0.027 | 0.026 | 0.027 | 0.024 | 0.024 | 0.024 |
| Bromazepam | 0.010 | 0.018 | 0.020 | 0.020 | 0.020 | 0.020 | 0.018 | 0.019 | 0.018 |
| Clobazam | 0.002 | 0.003 | 0.003 | 0.003 | 0.003 | 0.003 | 0.003 | 0.003 | 0.003 |
| Prazepam | 0.001 | 0.002 | 0.003 | 0.003 | 0.003 | 0.003 | 0.003 | 0.002 | 0.002 |
| Alprazolam | 0.000 | 0.000 | 0.000 | 0.000 | 0.000 | 0.000 | 0.000 | 0.000 | 0.000 |
| Flurazepam | 0.000 | 0.000 | 0.000 | 0.000 | 0.000 | 0.000 | 0.000 | 0.000 | 0.000 |
| Nitrazepam | 0.002 | 0.002 | 0.002 | 0.002 | 0.002 | 0.002 | 0.002 | 0.002 | 0.002 |
| Triazolam | 0.001 | 0.002 | 0.002 | 0.002 | 0.002 | 0.002 | 0.002 | 0.002 | 0.002 |
| Lormetazepam | 0.002 | 0.003 | 0.003 | 0.003 | 0.003 | 0.002 | 0.002 | 0.002 | 0.002 |
| Temazepam | 0.001 | 0.002 | 0.002 | 0.002 | 0.002 | 0.002 | 0.002 | 0.001 | 0.001 |
| Zopiclone | 0.000 | 0.000 | 0.000 | 0.000 | 0.000 | 0.000 | 0.000 | 0.000 | 0.000 |
| Zolpidem | 0.000 | 0.000 | 0.000 | 0.000 | 0.000 | 0.000 | 0.000 | 0.000 | 0.000 |
| Antidepressants^1^ | 0.000 | 0.001 | 0.000 | 0.000 | 0.000 | 0.000 | 0.000 | 0.000 | 0.001 |
| Antihistamines^2^ | 0.000 | 0.000 | 0.000 | 0.000 | 0.000 | 0.000 | 0.000 | 0.000 | 0.000 |
| Antipsychotics^3^ | 0.015 | 0.020 | 0.022 | 0.022 | 0.022 | 0.022 | 0.019 | 0.019 | 0.020 |

**Prevalence of chronic use (30 and 90 days) in the GMS population**

|  | **2014** | **2015** | **2016** | **2017** | **2018** | **2019** | **2020** | **2021** | **2022** |
| --- | --- | --- | --- | --- | --- | --- | --- | --- | --- |
| **30 days** |  |  |  |  |  |  |  |  |  |
| Sedatives | 0.122 | 0.122 | 0.127 | 0.130 | 0.133 | 0.134 | 0.131 | 0.135 | 0.130 |
| Benzodiazepines and z-drugs | 0.097 | 0.100 | 0.104 | 0.108 | 0.111 | 0.112 | 0.111 | 0.114 | 0.110 |
| Long-acting benzodiazepines | 0.114 | 0.112 | 0.115 | 0.116 | 0.117 | 0.116 | 0.111 | 0.112 | 0.106 |
| Clonazepam | 0.004 | 0.004 | 0.004 | 0.005 | 0.005 | 0.005 | 0.005 | 0.005 | 0.005 |
| Diazepam | 0.003 | 0.003 | 0.004 | 0.004 | 0.004 | 0.004 | 0.004 | 0.005 | 0.004 |
| Chloridiazepoxide | 0.022 | 0.022 | 0.023 | 0.023 | 0.023 | 0.023 | 0.022 | 0.023 | 0.023 |
| Potassium Clorazepate | 0.015 | 0.016 | 0.016 | 0.017 | 0.017 | 0.017 | 0.016 | 0.017 | 0.017 |
| Lorazepam | 0.002 | 0.002 | 0.002 | 0.002 | 0.002 | 0.002 | 0.002 | 0.002 | 0.002 |
| Bromazepam | 0.001 | 0.001 | 0.001 | 0.001 | 0.001 | 0.001 | 0.001 | 0.001 | 0.001 |
| Clobazam | 0.000 | 0.000 | 0.000 | 0.000 | 0.000 | 0.000 | 0.000 | 0.000 | 0.000 |
| Prazepam | 0.000 | 0.000 | 0.000 | 0.000 | 0.000 | 0.000 | 0.000 | 0.000 | 0.000 |
| Alprazolam | 0.003 | 0.003 | 0.003 | 0.003 | 0.003 | 0.003 | 0.003 | 0.003 | 0.003 |
| Flurazepam | 0.002 | 0.002 | 0.002 | 0.002 | 0.002 | 0.002 | 0.002 | 0.002 | 0.002 |
| Nitrazepam | 0.005 | 0.005 | 0.005 | 0.005 | 0.005 | 0.005 | 0.004 | 0.004 | 0.004 |
| Triazolam | 0.004 | 0.004 | 0.004 | 0.004 | 0.004 | 0.004 | 0.004 | 0.004 | 0.003 |
| Lormetazepam | 0.003 | 0.003 | 0.003 | 0.002 | 0.002 | 0.002 | 0.000 | 0.000 | 0.000 |
| Temazepam | 0.002 | 0.002 | 0.002 | 0.002 | 0.002 | 0.002 | 0.000 | 0.000 | 0.000 |
| Zopiclone | 0.006 | 0.006 | 0.005 | 0.005 | 0.005 | 0.005 | 0.004 | 0.004 | 0.004 |
| Zolpidem | 0.005 | 0.005 | 0.005 | 0.004 | 0.004 | 0.004 | 0.004 | 0.004 | 0.003 |
| Antidepressants^1^ | 0.031 | 0.031 | 0.032 | 0.033 | 0.033 | 0.034 | 0.033 | 0.033 | 0.032 |
| Antihistamines^2^ | 0.024 | 0.026 | 0.027 | 0.028 | 0.028 | 0.028 | 0.028 | 0.028 | 0.027 |
| Antipsychotics^3^ | 0.025 | 0.024 | 0.025 | 0.026 | 0.026 | 0.027 | 0.026 | 0.027 | 0.026 |
| **90 days** |  |  |  |  |  |  |  |  |  |
| Sedatives | 0.097 | 0.100 | 0.104 | 0.108 | 0.111 | 0.112 | 0.111 | 0.114 | 0.110 |
| Benzodiazepines and z-drugs | 0.114 | 0.112 | 0.115 | 0.116 | 0.117 | 0.116 | 0.111 | 0.112 | 0.106 |
| Long-acting benzodiazepines | 0.089 | 0.091 | 0.093 | 0.095 | 0.096 | 0.096 | 0.092 | 0.093 | 0.089 |
| Clonazepam | 0.003 | 0.003 | 0.004 | 0.004 | 0.004 | 0.004 | 0.004 | 0.005 | 0.004 |
| Diazepam | 0.022 | 0.022 | 0.023 | 0.023 | 0.023 | 0.023 | 0.022 | 0.023 | 0.023 |
| Chloridiazepoxide | 0.015 | 0.016 | 0.016 | 0.017 | 0.017 | 0.017 | 0.016 | 0.017 | 0.017 |
| Potassium Clorazepate | 0.002 | 0.002 | 0.002 | 0.002 | 0.002 | 0.002 | 0.002 | 0.002 | 0.002 |
| Lorazepam | 0.001 | 0.001 | 0.001 | 0.001 | 0.001 | 0.001 | 0.001 | 0.001 | 0.001 |
| Bromazepam | 0.000 | 0.000 | 0.000 | 0.000 | 0.000 | 0.000 | 0.000 | 0.000 | 0.000 |
| Clobazam | 0.000 | 0.000 | 0.000 | 0.000 | 0.000 | 0.000 | 0.000 | 0.000 | 0.000 |
| Prazepam | 0.003 | 0.003 | 0.003 | 0.003 | 0.003 | 0.003 | 0.003 | 0.003 | 0.003 |
| Alprazolam | 0.002 | 0.002 | 0.002 | 0.002 | 0.002 | 0.002 | 0.002 | 0.002 | 0.002 |
| Flurazepam | 0.005 | 0.005 | 0.005 | 0.005 | 0.005 | 0.005 | 0.004 | 0.004 | 0.004 |
| Nitrazepam | 0.004 | 0.004 | 0.004 | 0.004 | 0.004 | 0.004 | 0.004 | 0.004 | 0.003 |
| Triazolam | 0.001 | 0.001 | 0.001 | 0.001 | 0.001 | 0.001 | 0.001 | 0.001 | 0.001 |
| Lormetazepam | 0.001 | 0.001 | 0.001 | 0.001 | 0.001 | 0.001 | 0.001 | 0.001 | 0.001 |
| Temazepam | 0.001 | 0.001 | 0.001 | 0.001 | 0.001 | 0.001 | 0.001 | 0.001 | 0.001 |
| Zopiclone | 0.024 | 0.026 | 0.027 | 0.028 | 0.028 | 0.028 | 0.028 | 0.028 | 0.027 |
| Zolpidem | 0.025 | 0.024 | 0.025 | 0.026 | 0.026 | 0.027 | 0.026 | 0.027 | 0.026 |
| Antidepressants^1^ | 0.019 | 0.020 | 0.020 | 0.021 | 0.022 | 0.022 | 0.022 | 0.023 | 0.022 |
| Antihistamines^2^ | 0.010 | 0.011 | 0.013 | 0.014 | 0.016 | 0.017 | 0.018 | 0.020 | 0.020 |
| Antipsychotics^3^ | 0.007 | 0.008 | 0.010 | 0.011 | 0.012 | 0.013 | 0.014 | 0.016 | 0.016 |

^1^doxepin, trazodone, mirtazapine

^2^promethazine, cyclizine, ketotifen

^3^olanzapine, quetiapine, risperidone

**Supplementary Table 11. Prevalence of any dispensings, initiations (90/180 days), discontinuations (90/180 days), chronic use (30/90 days), and dispensings at a dose equivalent to >40mg diazepam per day in the GMS population, by age group and sex**

**Prevalence of any dispensings in the GMS population by age group**

|  | **<5** | **5-11** | **12-15** | **16-24** | **25-34** | **35-44** | **45-54** | **55-64** | **65-69** | **70-74** | **75+** |
| --- | --- | --- | --- | --- | --- | --- | --- | --- | --- | --- | --- |
| **2014** |  |  |  |  |  |  |  |  |  |  |  |
| Benzodiazepines and z-drugs | 0.003 | 0.003 | 0.006 | 0.060 | 0.156 | 0.195 | 0.241 | 0.286 | 0.299 | 0.307 | 0.382 |
| Long-acting benzodiazepines | 0.002 | 0.002 | 0.004 | 0.029 | 0.082 | 0.103 | 0.114 | 0.118 | 0.107 | 0.099 | 0.099 |
| Clonazepam | 0.000 | 0.000 | 0.000 | 0.002 | 0.006 | 0.007 | 0.009 | 0.009 | 0.008 | 0.008 | 0.008 |
| Diazepam | 0.002 | 0.002 | 0.003 | 0.024 | 0.068 | 0.083 | 0.085 | 0.083 | 0.074 | 0.069 | 0.066 |
| Chloridiazepoxide | 0.000 | 0.000 | 0.000 | 0.001 | 0.006 | 0.009 | 0.013 | 0.011 | 0.006 | 0.005 | 0.004 |
| Potassium Clorazepate | 0.000 | 0.000 | 0.000 | 0.000 | 0.000 | 0.000 | 0.000 | 0.000 | 0.000 | 0.000 | 0.000 |
| Lorazepam | 0.000 | 0.000 | 0.000 | 0.001 | 0.003 | 0.003 | 0.005 | 0.007 | 0.007 | 0.007 | 0.013 |
| Bromazepam | 0.000 | 0.000 | 0.000 | 0.002 | 0.004 | 0.005 | 0.008 | 0.014 | 0.018 | 0.018 | 0.019 |
| Clobazam | 0.001 | 0.001 | 0.001 | 0.001 | 0.002 | 0.002 | 0.002 | 0.002 | 0.002 | 0.002 | 0.002 |
| Prazepam | 0.000 | 0.000 | 0.000 | 0.000 | 0.001 | 0.001 | 0.002 | 0.003 | 0.004 | 0.004 | 0.003 |
| Alprazolam | 0.000 | 0.000 | 0.001 | 0.017 | 0.046 | 0.052 | 0.061 | 0.071 | 0.073 | 0.072 | 0.086 |
| Flurazepam | 0.000 | 0.000 | 0.000 | 0.001 | 0.006 | 0.010 | 0.015 | 0.019 | 0.018 | 0.015 | 0.015 |
| Nitrazepam | 0.000 | 0.000 | 0.000 | 0.000 | 0.000 | 0.001 | 0.001 | 0.002 | 0.003 | 0.003 | 0.007 |
| Triazolam | 0.000 | 0.000 | 0.000 | 0.001 | 0.003 | 0.003 | 0.006 | 0.009 | 0.011 | 0.012 | 0.017 |
| Lormetazepam | 0.000 | 0.000 | 0.000 | 0.000 | 0.001 | 0.002 | 0.003 | 0.006 | 0.008 | 0.011 | 0.016 |
| Temazepam | 0.000 | 0.000 | 0.000 | 0.001 | 0.003 | 0.004 | 0.008 | 0.013 | 0.016 | 0.018 | 0.033 |
| Zopiclone | 0.000 | 0.000 | 0.001 | 0.012 | 0.035 | 0.047 | 0.063 | 0.079 | 0.080 | 0.085 | 0.114 |
| Zolpidem | 0.000 | 0.000 | 0.000 | 0.011 | 0.029 | 0.041 | 0.056 | 0.071 | 0.076 | 0.081 | 0.100 |
| Antidepressants^1^ | 0.000 | 0.000 | 0.000 | 0.006 | 0.013 | 0.016 | 0.020 | 0.021 | 0.021 | 0.022 | 0.039 |
| Antihistamines^2^ | 0.000 | 0.000 | 0.000 | 0.002 | 0.004 | 0.004 | 0.005 | 0.004 | 0.003 | 0.003 | 0.003 |
| Antipsychotics^3^ | 0.000 | 0.000 | 0.001 | 0.007 | 0.014 | 0.015 | 0.018 | 0.018 | 0.017 | 0.020 | 0.049 |
| Sedatives | 0.003 | 0.003 | 0.007 | 0.066 | 0.165 | 0.203 | 0.250 | 0.295 | 0.307 | 0.315 | 0.395 |
| **2015** |  |  |  |  |  |  |  |  |  |  |  |
| Benzodiazepines and z-drugs | 0.002 | 0.003 | 0.006 | 0.064 | 0.156 | 0.191 | 0.232 | 0.274 | 0.289 | 0.290 | 0.356 |
| Long-acting benzodiazepines | 0.002 | 0.002 | 0.004 | 0.029 | 0.082 | 0.101 | 0.110 | 0.112 | 0.105 | 0.094 | 0.092 |
| Clonazepam | 0.000 | 0.000 | 0.000 | 0.003 | 0.006 | 0.007 | 0.009 | 0.009 | 0.008 | 0.007 | 0.008 |
| Diazepam | 0.002 | 0.001 | 0.004 | 0.024 | 0.068 | 0.082 | 0.083 | 0.080 | 0.074 | 0.065 | 0.063 |
| Chloridiazepoxide | 0.000 | 0.000 | 0.000 | 0.001 | 0.006 | 0.009 | 0.012 | 0.010 | 0.007 | 0.005 | 0.003 |
| Potassium Clorazepate | 0.000 | 0.000 | 0.000 | 0.000 | 0.000 | 0.000 | 0.000 | 0.000 | 0.000 | 0.000 | 0.000 |
| Lorazepam | 0.000 | 0.000 | 0.000 | 0.002 | 0.003 | 0.003 | 0.005 | 0.006 | 0.007 | 0.007 | 0.012 |
| Bromazepam | 0.000 | 0.000 | 0.000 | 0.002 | 0.004 | 0.005 | 0.008 | 0.012 | 0.017 | 0.017 | 0.018 |
| Clobazam | 0.000 | 0.001 | 0.000 | 0.001 | 0.001 | 0.002 | 0.002 | 0.002 | 0.002 | 0.002 | 0.002 |
| Prazepam | 0.000 | 0.000 | 0.000 | 0.000 | 0.001 | 0.001 | 0.001 | 0.003 | 0.003 | 0.003 | 0.003 |
| Alprazolam | 0.000 | 0.000 | 0.001 | 0.019 | 0.048 | 0.052 | 0.060 | 0.070 | 0.070 | 0.070 | 0.083 |
| Flurazepam | 0.000 | 0.000 | 0.000 | 0.001 | 0.006 | 0.009 | 0.013 | 0.017 | 0.016 | 0.014 | 0.013 |
| Nitrazepam | 0.000 | 0.000 | 0.000 | 0.000 | 0.000 | 0.001 | 0.001 | 0.002 | 0.002 | 0.003 | 0.005 |
| Triazolam | 0.000 | 0.000 | 0.000 | 0.001 | 0.003 | 0.003 | 0.005 | 0.008 | 0.010 | 0.010 | 0.015 |
| Lormetazepam | 0.000 | 0.000 | 0.000 | 0.000 | 0.001 | 0.002 | 0.003 | 0.005 | 0.007 | 0.009 | 0.014 |
| Temazepam | 0.000 | 0.000 | 0.000 | 0.001 | 0.002 | 0.004 | 0.007 | 0.011 | 0.014 | 0.016 | 0.028 |
| Zopiclone | 0.000 | 0.000 | 0.001 | 0.013 | 0.034 | 0.045 | 0.061 | 0.076 | 0.078 | 0.081 | 0.108 |
| Zolpidem | 0.000 | 0.000 | 0.000 | 0.012 | 0.028 | 0.038 | 0.053 | 0.068 | 0.072 | 0.075 | 0.093 |
| Antidepressants^1^ | 0.000 | 0.000 | 0.000 | 0.007 | 0.015 | 0.017 | 0.021 | 0.023 | 0.022 | 0.024 | 0.042 |
| Antihistamines^2^ | 0.000 | 0.000 | 0.000 | 0.003 | 0.005 | 0.005 | 0.005 | 0.005 | 0.003 | 0.003 | 0.003 |
| Antipsychotics^3^ | 0.000 | 0.000 | 0.001 | 0.009 | 0.016 | 0.016 | 0.019 | 0.018 | 0.018 | 0.020 | 0.051 |
| Sedatives | 0.002 | 0.003 | 0.007 | 0.071 | 0.167 | 0.201 | 0.243 | 0.284 | 0.298 | 0.300 | 0.374 |
| **2016** |  |  |  |  |  |  |  |  |  |  |  |
| Benzodiazepines and z-drugs | 0.002 | 0.003 | 0.006 | 0.063 | 0.167 | 0.196 | 0.232 | 0.273 | 0.288 | 0.289 | 0.354 |
| Long-acting benzodiazepines | 0.002 | 0.002 | 0.004 | 0.029 | 0.088 | 0.104 | 0.111 | 0.113 | 0.105 | 0.094 | 0.090 |
| Clonazepam | 0.000 | 0.000 | 0.000 | 0.003 | 0.007 | 0.008 | 0.009 | 0.010 | 0.009 | 0.008 | 0.008 |
| Diazepam | 0.001 | 0.001 | 0.003 | 0.024 | 0.073 | 0.085 | 0.085 | 0.082 | 0.075 | 0.066 | 0.062 |
| Chloridiazepoxide | 0.000 | 0.000 | 0.000 | 0.001 | 0.006 | 0.009 | 0.012 | 0.010 | 0.007 | 0.005 | 0.003 |
| Potassium Clorazepate | 0.000 | 0.000 | 0.000 | 0.000 | 0.000 | 0.000 | 0.000 | 0.000 | 0.000 | 0.000 | 0.000 |
| Lorazepam | 0.000 | 0.000 | 0.000 | 0.002 | 0.004 | 0.004 | 0.005 | 0.006 | 0.007 | 0.007 | 0.011 |
| Bromazepam | 0.000 | 0.000 | 0.000 | 0.002 | 0.005 | 0.005 | 0.007 | 0.012 | 0.016 | 0.016 | 0.017 |
| Clobazam | 0.000 | 0.001 | 0.000 | 0.001 | 0.001 | 0.001 | 0.002 | 0.002 | 0.002 | 0.002 | 0.001 |
| Prazepam | 0.000 | 0.000 | 0.000 | 0.000 | 0.001 | 0.001 | 0.001 | 0.002 | 0.003 | 0.003 | 0.003 |
| Alprazolam | 0.000 | 0.000 | 0.001 | 0.019 | 0.053 | 0.055 | 0.061 | 0.070 | 0.072 | 0.071 | 0.084 |
| Flurazepam | 0.000 | 0.000 | 0.000 | 0.001 | 0.005 | 0.009 | 0.012 | 0.016 | 0.015 | 0.014 | 0.012 |
| Nitrazepam | 0.000 | 0.000 | 0.000 | 0.000 | 0.000 | 0.000 | 0.001 | 0.001 | 0.002 | 0.003 | 0.005 |
| Triazolam | 0.000 | 0.000 | 0.000 | 0.001 | 0.003 | 0.003 | 0.005 | 0.008 | 0.010 | 0.010 | 0.015 |
| Lormetazepam | 0.000 | 0.000 | 0.000 | 0.000 | 0.001 | 0.002 | 0.003 | 0.004 | 0.006 | 0.008 | 0.013 |
| Temazepam | 0.000 | 0.000 | 0.000 | 0.001 | 0.002 | 0.004 | 0.006 | 0.010 | 0.013 | 0.015 | 0.026 |
| Zopiclone | 0.000 | 0.000 | 0.000 | 0.013 | 0.037 | 0.047 | 0.062 | 0.075 | 0.079 | 0.080 | 0.108 |
| Zolpidem | 0.000 | 0.000 | 0.000 | 0.011 | 0.030 | 0.038 | 0.052 | 0.067 | 0.073 | 0.076 | 0.094 |
| Antidepressants^1^ | 0.000 | 0.000 | 0.000 | 0.007 | 0.019 | 0.020 | 0.023 | 0.024 | 0.023 | 0.026 | 0.046 |
| Antihistamines^2^ | 0.000 | 0.000 | 0.000 | 0.004 | 0.007 | 0.007 | 0.007 | 0.006 | 0.004 | 0.004 | 0.004 |
| Antipsychotics^3^ | 0.000 | 0.000 | 0.001 | 0.011 | 0.020 | 0.018 | 0.020 | 0.020 | 0.020 | 0.021 | 0.053 |
| Sedatives | 0.002 | 0.003 | 0.006 | 0.072 | 0.181 | 0.209 | 0.245 | 0.285 | 0.299 | 0.301 | 0.376 |
| **2017** |  |  |  |  |  |  |  |  |  |  |  |
| Benzodiazepines and z-drugs | 0.002 | 0.003 | 0.006 | 0.058 | 0.171 | 0.206 | 0.236 | 0.269 | 0.278 | 0.279 | 0.339 |
| Long-acting benzodiazepines | 0.002 | 0.002 | 0.004 | 0.027 | 0.090 | 0.109 | 0.114 | 0.111 | 0.100 | 0.090 | 0.085 |
| Clonazepam | 0.000 | 0.000 | 0.000 | 0.003 | 0.008 | 0.008 | 0.009 | 0.010 | 0.008 | 0.009 | 0.008 |
| Diazepam | 0.001 | 0.001 | 0.003 | 0.022 | 0.075 | 0.089 | 0.088 | 0.081 | 0.071 | 0.063 | 0.059 |
| Chloridiazepoxide | 0.000 | 0.000 | 0.000 | 0.001 | 0.006 | 0.010 | 0.012 | 0.010 | 0.007 | 0.004 | 0.003 |
| Potassium Clorazepate | 0.000 | 0.000 | 0.000 | 0.000 | 0.000 | 0.000 | 0.000 | 0.000 | 0.000 | 0.000 | 0.000 |
| Lorazepam | 0.000 | 0.000 | 0.000 | 0.002 | 0.004 | 0.004 | 0.005 | 0.006 | 0.006 | 0.006 | 0.010 |
| Bromazepam | 0.000 | 0.000 | 0.000 | 0.001 | 0.004 | 0.005 | 0.007 | 0.011 | 0.015 | 0.016 | 0.016 |
| Clobazam | 0.000 | 0.000 | 0.000 | 0.001 | 0.002 | 0.001 | 0.002 | 0.002 | 0.002 | 0.001 | 0.001 |
| Prazepam | 0.000 | 0.000 | 0.000 | 0.000 | 0.001 | 0.001 | 0.001 | 0.002 | 0.003 | 0.003 | 0.003 |
| Alprazolam | 0.000 | 0.000 | 0.001 | 0.018 | 0.055 | 0.058 | 0.061 | 0.069 | 0.069 | 0.068 | 0.081 |
| Flurazepam | 0.000 | 0.000 | 0.000 | 0.001 | 0.005 | 0.009 | 0.012 | 0.015 | 0.014 | 0.013 | 0.011 |
| Nitrazepam | 0.000 | 0.000 | 0.000 | 0.000 | 0.000 | 0.000 | 0.001 | 0.001 | 0.002 | 0.002 | 0.004 |
| Triazolam | 0.000 | 0.000 | 0.000 | 0.001 | 0.003 | 0.004 | 0.005 | 0.007 | 0.009 | 0.009 | 0.014 |
| Lormetazepam | 0.000 | 0.000 | 0.000 | 0.000 | 0.001 | 0.001 | 0.002 | 0.004 | 0.006 | 0.007 | 0.012 |
| Temazepam | 0.000 | 0.000 | 0.000 | 0.001 | 0.002 | 0.003 | 0.006 | 0.009 | 0.012 | 0.013 | 0.023 |
| Zopiclone | 0.000 | 0.000 | 0.000 | 0.011 | 0.037 | 0.049 | 0.062 | 0.075 | 0.076 | 0.077 | 0.103 |
| Zolpidem | 0.000 | 0.000 | 0.000 | 0.010 | 0.028 | 0.038 | 0.053 | 0.065 | 0.071 | 0.074 | 0.091 |
| Antidepressants^1^ | 0.000 | 0.000 | 0.000 | 0.008 | 0.021 | 0.023 | 0.025 | 0.026 | 0.023 | 0.027 | 0.047 |
| Antihistamines^2^ | 0.000 | 0.000 | 0.000 | 0.006 | 0.011 | 0.010 | 0.010 | 0.009 | 0.007 | 0.006 | 0.006 |
| Antipsychotics^3^ | 0.000 | 0.000 | 0.001 | 0.011 | 0.024 | 0.021 | 0.023 | 0.022 | 0.020 | 0.022 | 0.054 |
| Sedatives | 0.002 | 0.003 | 0.006 | 0.068 | 0.190 | 0.221 | 0.253 | 0.284 | 0.290 | 0.292 | 0.364 |
| **2018** |  |  |  |  |  |  |  |  |  |  |  |
| Benzodiazepines and z-drugs | 0.002 | 0.003 | 0.006 | 0.058 | 0.174 | 0.206 | 0.225 | 0.260 | 0.276 | 0.272 | 0.331 |
| Long-acting benzodiazepines | 0.002 | 0.002 | 0.004 | 0.027 | 0.092 | 0.111 | 0.109 | 0.107 | 0.101 | 0.087 | 0.084 |
| Clonazepam | 0.000 | 0.000 | 0.000 | 0.003 | 0.009 | 0.008 | 0.009 | 0.010 | 0.009 | 0.008 | 0.009 |
| Diazepam | 0.001 | 0.001 | 0.003 | 0.022 | 0.076 | 0.091 | 0.084 | 0.078 | 0.073 | 0.062 | 0.058 |
| Chloridiazepoxide | 0.000 | 0.000 | 0.000 | 0.001 | 0.007 | 0.010 | 0.011 | 0.010 | 0.007 | 0.004 | 0.003 |
| Potassium Clorazepate | 0.000 | 0.000 | 0.000 | 0.000 | 0.000 | 0.000 | 0.000 | 0.000 | 0.000 | 0.000 | 0.000 |
| Lorazepam | 0.000 | 0.000 | 0.000 | 0.002 | 0.005 | 0.004 | 0.005 | 0.006 | 0.007 | 0.006 | 0.010 |
| Bromazepam | 0.000 | 0.000 | 0.000 | 0.001 | 0.004 | 0.005 | 0.006 | 0.010 | 0.014 | 0.016 | 0.016 |
| Clobazam | 0.000 | 0.000 | 0.000 | 0.001 | 0.002 | 0.002 | 0.002 | 0.002 | 0.002 | 0.001 | 0.001 |
| Prazepam | 0.000 | 0.000 | 0.000 | 0.000 | 0.001 | 0.001 | 0.001 | 0.002 | 0.003 | 0.003 | 0.003 |
| Alprazolam | 0.000 | 0.000 | 0.001 | 0.018 | 0.056 | 0.060 | 0.060 | 0.067 | 0.068 | 0.066 | 0.077 |
| Flurazepam | 0.000 | 0.000 | 0.000 | 0.001 | 0.005 | 0.008 | 0.011 | 0.014 | 0.014 | 0.012 | 0.010 |
| Nitrazepam | 0.000 | 0.000 | 0.000 | 0.000 | 0.000 | 0.000 | 0.001 | 0.001 | 0.002 | 0.002 | 0.004 |
| Triazolam | 0.000 | 0.000 | 0.000 | 0.001 | 0.003 | 0.003 | 0.005 | 0.007 | 0.009 | 0.009 | 0.013 |
| Lormetazepam | 0.000 | 0.000 | 0.000 | 0.001 | 0.001 | 0.001 | 0.002 | 0.003 | 0.005 | 0.006 | 0.011 |
| Temazepam | 0.000 | 0.000 | 0.000 | 0.001 | 0.002 | 0.003 | 0.005 | 0.008 | 0.010 | 0.012 | 0.020 |
| Zopiclone | 0.000 | 0.000 | 0.000 | 0.011 | 0.038 | 0.048 | 0.058 | 0.072 | 0.075 | 0.076 | 0.101 |
| Zolpidem | 0.000 | 0.000 | 0.000 | 0.009 | 0.028 | 0.038 | 0.049 | 0.063 | 0.071 | 0.071 | 0.090 |
| Antidepressants^1^ | 0.000 | 0.000 | 0.000 | 0.008 | 0.025 | 0.025 | 0.027 | 0.028 | 0.026 | 0.027 | 0.050 |
| Antihistamines^2^ | 0.000 | 0.000 | 0.000 | 0.006 | 0.013 | 0.012 | 0.012 | 0.010 | 0.007 | 0.006 | 0.006 |
| Antipsychotics^3^ | 0.000 | 0.000 | 0.001 | 0.013 | 0.030 | 0.024 | 0.025 | 0.024 | 0.022 | 0.022 | 0.055 |
| Sedatives | 0.002 | 0.003 | 0.007 | 0.069 | 0.197 | 0.225 | 0.243 | 0.277 | 0.290 | 0.286 | 0.361 |
| **2019** |  |  |  |  |  |  |  |  |  |  |  |
| Benzodiazepines and z-drugs | 0.002 | 0.002 | 0.005 | 0.057 | 0.171 | 0.201 | 0.222 | 0.255 | 0.275 | 0.265 | 0.321 |
| Long-acting benzodiazepines | 0.001 | 0.002 | 0.003 | 0.026 | 0.090 | 0.108 | 0.108 | 0.107 | 0.100 | 0.085 | 0.081 |
| Clonazepam | 0.000 | 0.000 | 0.000 | 0.003 | 0.008 | 0.009 | 0.009 | 0.010 | 0.009 | 0.008 | 0.009 |
| Diazepam | 0.001 | 0.001 | 0.003 | 0.021 | 0.075 | 0.088 | 0.084 | 0.079 | 0.073 | 0.060 | 0.055 |
| Chloridiazepoxide | 0.000 | 0.000 | 0.000 | 0.001 | 0.007 | 0.011 | 0.011 | 0.010 | 0.006 | 0.004 | 0.002 |
| Potassium Clorazepate | 0.000 | 0.000 | 0.000 | 0.000 | 0.000 | 0.000 | 0.000 | 0.000 | 0.000 | 0.000 | 0.000 |
| Lorazepam | 0.000 | 0.000 | 0.000 | 0.002 | 0.004 | 0.005 | 0.005 | 0.006 | 0.007 | 0.006 | 0.010 |
| Bromazepam | 0.000 | 0.000 | 0.000 | 0.001 | 0.004 | 0.005 | 0.006 | 0.009 | 0.013 | 0.015 | 0.015 |
| Clobazam | 0.000 | 0.000 | 0.000 | 0.001 | 0.001 | 0.002 | 0.002 | 0.002 | 0.002 | 0.001 | 0.001 |
| Prazepam | 0.000 | 0.000 | 0.000 | 0.000 | 0.001 | 0.001 | 0.001 | 0.002 | 0.003 | 0.002 | 0.002 |
| Alprazolam | 0.000 | 0.000 | 0.001 | 0.019 | 0.058 | 0.059 | 0.060 | 0.066 | 0.069 | 0.065 | 0.076 |
| Flurazepam | 0.000 | 0.000 | 0.000 | 0.001 | 0.004 | 0.008 | 0.010 | 0.012 | 0.013 | 0.011 | 0.010 |
| Nitrazepam | 0.000 | 0.000 | 0.000 | 0.000 | 0.000 | 0.000 | 0.001 | 0.001 | 0.002 | 0.002 | 0.004 |
| Triazolam | 0.000 | 0.000 | 0.000 | 0.000 | 0.003 | 0.003 | 0.005 | 0.007 | 0.008 | 0.008 | 0.013 |
| Lormetazepam | 0.000 | 0.000 | 0.000 | 0.000 | 0.001 | 0.001 | 0.001 | 0.002 | 0.004 | 0.005 | 0.009 |
| Temazepam | 0.000 | 0.000 | 0.000 | 0.001 | 0.002 | 0.003 | 0.004 | 0.007 | 0.010 | 0.011 | 0.019 |
| Zopiclone | 0.000 | 0.000 | 0.000 | 0.010 | 0.036 | 0.046 | 0.057 | 0.071 | 0.077 | 0.075 | 0.100 |
| Zolpidem | 0.000 | 0.000 | 0.000 | 0.009 | 0.027 | 0.036 | 0.047 | 0.062 | 0.072 | 0.072 | 0.091 |
| Antidepressants^1^ | 0.000 | 0.000 | 0.000 | 0.008 | 0.027 | 0.025 | 0.028 | 0.029 | 0.029 | 0.030 | 0.052 |
| Antihistamines^2^ | 0.000 | 0.000 | 0.000 | 0.008 | 0.019 | 0.016 | 0.015 | 0.013 | 0.010 | 0.008 | 0.007 |
| Antipsychotics^3^ | 0.000 | 0.000 | 0.001 | 0.015 | 0.033 | 0.028 | 0.027 | 0.026 | 0.023 | 0.023 | 0.057 |
| Sedatives | 0.002 | 0.003 | 0.006 | 0.069 | 0.197 | 0.221 | 0.242 | 0.274 | 0.291 | 0.281 | 0.353 |
| **2020** |  |  |  |  |  |  |  |  |  |  |  |
| Benzodiazepines and z-drugs | 0.002 | 0.002 | 0.004 | 0.043 | 0.147 | 0.178 | 0.202 | 0.233 | 0.251 | 0.247 | 0.303 |
| Long-acting benzodiazepines | 0.001 | 0.002 | 0.003 | 0.020 | 0.078 | 0.097 | 0.100 | 0.097 | 0.089 | 0.079 | 0.076 |
| Clonazepam | 0.000 | 0.000 | 0.000 | 0.002 | 0.007 | 0.008 | 0.009 | 0.010 | 0.009 | 0.009 | 0.009 |
| Diazepam | 0.001 | 0.001 | 0.002 | 0.016 | 0.064 | 0.079 | 0.078 | 0.071 | 0.063 | 0.056 | 0.052 |
| Chloridiazepoxide | 0.000 | 0.000 | 0.000 | 0.001 | 0.006 | 0.009 | 0.011 | 0.009 | 0.006 | 0.003 | 0.002 |
| Potassium Clorazepate | 0.000 | 0.000 | 0.000 | 0.000 | 0.000 | 0.000 | 0.000 | 0.000 | 0.000 | 0.000 | 0.000 |
| Lorazepam | 0.000 | 0.000 | 0.000 | 0.002 | 0.004 | 0.004 | 0.005 | 0.006 | 0.006 | 0.006 | 0.011 |
| Bromazepam | 0.000 | 0.000 | 0.000 | 0.001 | 0.003 | 0.004 | 0.005 | 0.008 | 0.012 | 0.013 | 0.014 |
| Clobazam | 0.000 | 0.000 | 0.000 | 0.001 | 0.001 | 0.001 | 0.001 | 0.001 | 0.001 | 0.001 | 0.001 |
| Prazepam | 0.000 | 0.000 | 0.000 | 0.000 | 0.000 | 0.001 | 0.001 | 0.001 | 0.002 | 0.002 | 0.002 |
| Alprazolam | 0.000 | 0.000 | 0.001 | 0.013 | 0.048 | 0.050 | 0.052 | 0.057 | 0.061 | 0.060 | 0.071 |
| Flurazepam | 0.000 | 0.000 | 0.000 | 0.001 | 0.003 | 0.006 | 0.009 | 0.011 | 0.012 | 0.010 | 0.009 |
| Nitrazepam | 0.000 | 0.000 | 0.000 | 0.000 | 0.000 | 0.000 | 0.000 | 0.001 | 0.001 | 0.002 | 0.003 |
| Triazolam | 0.000 | 0.000 | 0.000 | 0.000 | 0.002 | 0.003 | 0.004 | 0.007 | 0.007 | 0.008 | 0.012 |
| Lormetazepam | 0.000 | 0.000 | 0.000 | 0.000 | 0.000 | 0.000 | 0.000 | 0.000 | 0.001 | 0.001 | 0.002 |
| Temazepam | 0.000 | 0.000 | 0.000 | 0.000 | 0.002 | 0.003 | 0.004 | 0.006 | 0.008 | 0.010 | 0.017 |
| Zopiclone | 0.000 | 0.000 | 0.000 | 0.008 | 0.031 | 0.041 | 0.052 | 0.066 | 0.073 | 0.071 | 0.096 |
| Zolpidem | 0.000 | 0.000 | 0.000 | 0.006 | 0.023 | 0.032 | 0.043 | 0.058 | 0.066 | 0.069 | 0.086 |
| Antidepressants^1^ | 0.000 | 0.000 | 0.000 | 0.007 | 0.026 | 0.025 | 0.028 | 0.029 | 0.030 | 0.031 | 0.055 |
| Antihistamines^2^ | 0.000 | 0.000 | 0.000 | 0.008 | 0.021 | 0.019 | 0.018 | 0.016 | 0.012 | 0.009 | 0.009 |
| Antipsychotics^3^ | 0.000 | 0.000 | 0.001 | 0.014 | 0.034 | 0.029 | 0.028 | 0.027 | 0.024 | 0.024 | 0.061 |
| Sedatives | 0.002 | 0.002 | 0.005 | 0.055 | 0.174 | 0.201 | 0.226 | 0.254 | 0.269 | 0.264 | 0.339 |
| **2021** |  |  |  |  |  |  |  |  |  |  |  |
| Benzodiazepines and z-drugs | 0.001 | 0.002 | 0.005 | 0.048 | 0.150 | 0.183 | 0.205 | 0.232 | 0.252 | 0.242 | 0.297 |
| Long-acting benzodiazepines | 0.001 | 0.002 | 0.003 | 0.023 | 0.080 | 0.101 | 0.103 | 0.097 | 0.091 | 0.078 | 0.074 |
| Clonazepam | 0.000 | 0.000 | 0.000 | 0.003 | 0.008 | 0.008 | 0.009 | 0.010 | 0.010 | 0.008 | 0.010 |
| Diazepam | 0.001 | 0.001 | 0.002 | 0.018 | 0.066 | 0.082 | 0.080 | 0.072 | 0.065 | 0.056 | 0.051 |
| Chloridiazepoxide | 0.000 | 0.000 | 0.000 | 0.001 | 0.006 | 0.010 | 0.011 | 0.009 | 0.006 | 0.003 | 0.002 |
| Potassium Clorazepate | 0.000 | 0.000 | 0.000 | 0.000 | 0.000 | 0.000 | 0.000 | 0.000 | 0.000 | 0.000 | 0.000 |
| Lorazepam | 0.000 | 0.000 | 0.000 | 0.003 | 0.005 | 0.005 | 0.005 | 0.006 | 0.007 | 0.006 | 0.011 |
| Bromazepam | 0.000 | 0.000 | 0.000 | 0.001 | 0.003 | 0.004 | 0.004 | 0.007 | 0.011 | 0.012 | 0.013 |
| Clobazam | 0.001 | 0.001 | 0.000 | 0.001 | 0.001 | 0.001 | 0.001 | 0.002 | 0.001 | 0.001 | 0.001 |
| Prazepam | 0.000 | 0.000 | 0.000 | 0.000 | 0.000 | 0.001 | 0.001 | 0.001 | 0.002 | 0.002 | 0.002 |
| Alprazolam | 0.000 | 0.000 | 0.001 | 0.015 | 0.048 | 0.052 | 0.053 | 0.057 | 0.060 | 0.058 | 0.069 |
| Flurazepam | 0.000 | 0.000 | 0.000 | 0.001 | 0.004 | 0.006 | 0.009 | 0.011 | 0.012 | 0.010 | 0.009 |
| Nitrazepam | 0.000 | 0.000 | 0.000 | 0.000 | 0.000 | 0.000 | 0.000 | 0.001 | 0.001 | 0.001 | 0.003 |
| Triazolam | 0.000 | 0.000 | 0.000 | 0.000 | 0.002 | 0.003 | 0.004 | 0.006 | 0.007 | 0.008 | 0.011 |
| Lormetazepam | 0.000 | 0.000 | 0.000 | 0.000 | 0.000 | 0.000 | 0.000 | 0.000 | 0.000 | 0.000 | 0.000 |
| Temazepam | 0.000 | 0.000 | 0.000 | 0.000 | 0.002 | 0.002 | 0.004 | 0.006 | 0.008 | 0.009 | 0.016 |
| Zopiclone | 0.000 | 0.000 | 0.000 | 0.008 | 0.031 | 0.041 | 0.052 | 0.065 | 0.072 | 0.070 | 0.095 |
| Zolpidem | 0.000 | 0.000 | 0.000 | 0.006 | 0.023 | 0.031 | 0.042 | 0.056 | 0.066 | 0.067 | 0.084 |
| Antidepressants^1^ | 0.000 | 0.000 | 0.000 | 0.008 | 0.029 | 0.028 | 0.030 | 0.033 | 0.033 | 0.033 | 0.059 |
| Antihistamines^2^ | 0.000 | 0.000 | 0.001 | 0.010 | 0.026 | 0.023 | 0.022 | 0.019 | 0.014 | 0.011 | 0.011 |
| Antipsychotics^3^ | 0.000 | 0.000 | 0.001 | 0.016 | 0.039 | 0.033 | 0.031 | 0.029 | 0.027 | 0.025 | 0.061 |
| Sedatives | 0.001 | 0.002 | 0.005 | 0.061 | 0.184 | 0.211 | 0.232 | 0.257 | 0.273 | 0.261 | 0.336 |
| **2022** |  |  |  |  |  |  |  |  |  |  |  |
| Benzodiazepines and z-drugs | 0.002 | 0.002 | 0.004 | 0.042 | 0.137 | 0.175 | 0.200 | 0.226 | 0.243 | 0.238 | 0.290 |
| Long-acting benzodiazepines | 0.001 | 0.002 | 0.003 | 0.020 | 0.072 | 0.095 | 0.099 | 0.095 | 0.088 | 0.077 | 0.071 |
| Clonazepam | 0.000 | 0.000 | 0.000 | 0.002 | 0.007 | 0.008 | 0.009 | 0.009 | 0.010 | 0.008 | 0.009 |
| Diazepam | 0.001 | 0.001 | 0.002 | 0.017 | 0.062 | 0.080 | 0.080 | 0.074 | 0.068 | 0.059 | 0.054 |
| Chloridiazepoxide | 0.000 | 0.000 | 0.000 | 0.001 | 0.005 | 0.009 | 0.010 | 0.008 | 0.005 | 0.004 | 0.002 |
| Potassium Clorazepate | 0.000 | 0.000 | 0.000 | 0.000 | 0.000 | 0.000 | 0.000 | 0.000 | 0.000 | 0.000 | 0.000 |
| Lorazepam | 0.000 | 0.000 | 0.001 | 0.003 | 0.005 | 0.005 | 0.006 | 0.006 | 0.007 | 0.007 | 0.011 |
| Bromazepam | 0.000 | 0.000 | 0.000 | 0.001 | 0.003 | 0.004 | 0.004 | 0.007 | 0.010 | 0.012 | 0.013 |
| Clobazam | 0.000 | 0.001 | 0.000 | 0.001 | 0.002 | 0.002 | 0.002 | 0.002 | 0.001 | 0.001 | 0.001 |
| Prazepam | 0.000 | 0.000 | 0.000 | 0.000 | 0.000 | 0.000 | 0.001 | 0.001 | 0.002 | 0.002 | 0.002 |
| Alprazolam | 0.000 | 0.000 | 0.001 | 0.014 | 0.048 | 0.054 | 0.056 | 0.060 | 0.063 | 0.061 | 0.070 |
| Flurazepam | 0.000 | 0.000 | 0.000 | 0.001 | 0.003 | 0.005 | 0.008 | 0.010 | 0.010 | 0.009 | 0.008 |
| Nitrazepam | 0.000 | 0.000 | 0.000 | 0.000 | 0.000 | 0.000 | 0.000 | 0.000 | 0.000 | 0.000 | 0.000 |
| Triazolam | 0.000 | 0.000 | 0.000 | 0.000 | 0.002 | 0.003 | 0.004 | 0.006 | 0.006 | 0.007 | 0.011 |
| Lormetazepam | 0.000 | 0.000 | 0.000 | 0.000 | 0.000 | 0.000 | 0.000 | 0.000 | 0.000 | 0.000 | 0.000 |
| Temazepam | 0.000 | 0.000 | 0.000 | 0.000 | 0.001 | 0.002 | 0.003 | 0.005 | 0.007 | 0.008 | 0.014 |
| Zopiclone | 0.000 | 0.000 | 0.000 | 0.007 | 0.027 | 0.039 | 0.049 | 0.062 | 0.069 | 0.067 | 0.092 |
| Zolpidem | 0.000 | 0.000 | 0.000 | 0.005 | 0.018 | 0.027 | 0.039 | 0.052 | 0.062 | 0.065 | 0.082 |
| Antidepressants^1^ | 0.000 | 0.000 | 0.000 | 0.006 | 0.025 | 0.027 | 0.029 | 0.032 | 0.032 | 0.034 | 0.060 |
| Antihistamines^2^ | 0.000 | 0.000 | 0.001 | 0.008 | 0.024 | 0.024 | 0.024 | 0.020 | 0.016 | 0.012 | 0.012 |
| Antipsychotics^3^ | 0.000 | 0.000 | 0.001 | 0.013 | 0.037 | 0.034 | 0.031 | 0.030 | 0.026 | 0.025 | 0.059 |
| Sedatives | 0.002 | 0.002 | 0.005 | 0.052 | 0.168 | 0.203 | 0.227 | 0.251 | 0.264 | 0.257 | 0.329 |

**Prevalence of any dispensings in the GMS population by sex**

|  | **Females** | **Males** |
| --- | --- | --- |
| **2014** |  |  |
| Benzodiazepines and z-drugs | 0.22 | 0.14 |
| Long-acting benzodiazepines | 0.08 | 0.06 |
| Clonazepam | 0.01 | 0.01 |
| Diazepam | 0.06 | 0.04 |
| Chloridiazepoxide | 0.00 | 0.01 |
| Potassium Clorazepate | 0.00 | 0.00 |
| Lorazepam | 0.00 | 0.00 |
| Bromazepam | 0.01 | 0.00 |
| Clobazam | 0.00 | 0.00 |
| Prazepam | 0.00 | 0.00 |
| Alprazolam | 0.06 | 0.03 |
| Flurazepam | 0.01 | 0.01 |
| Nitrazepam | 0.00 | 0.00 |
| Triazolam | 0.01 | 0.00 |
| Lormetazepam | 0.01 | 0.00 |
| Temazepam | 0.01 | 0.01 |
| Zopiclone | 0.06 | 0.04 |
| Zolpidem | 0.05 | 0.03 |
| Antidepressants^1^ | 0.02 | 0.01 |
| Antihistamines^2^ | 0.00 | 0.00 |
| Antipsychotics^3^ | 0.02 | 0.01 |
| Sedatives | 0.23 | 0.15 |
| **2015** |  |  |
| Benzodiazepines and z-drugs | 0.21 | 0.14 |
| Long-acting benzodiazepines | 0.08 | 0.06 |
| Clonazepam | 0.01 | 0.01 |
| Diazepam | 0.06 | 0.04 |
| Chloridiazepoxide | 0.00 | 0.01 |
| Potassium Clorazepate | 0.00 | 0.00 |
| Lorazepam | 0.00 | 0.00 |
| Bromazepam | 0.01 | 0.00 |
| Clobazam | 0.00 | 0.00 |
| Prazepam | 0.00 | 0.00 |
| Alprazolam | 0.06 | 0.03 |
| Flurazepam | 0.01 | 0.01 |
| Nitrazepam | 0.00 | 0.00 |
| Triazolam | 0.01 | 0.00 |
| Lormetazepam | 0.01 | 0.00 |
| Temazepam | 0.01 | 0.01 |
| Zopiclone | 0.06 | 0.04 |
| Zolpidem | 0.05 | 0.03 |
| Antidepressants^1^ | 0.02 | 0.01 |
| Antihistamines^2^ | 0.00 | 0.00 |
| Antipsychotics^3^ | 0.02 | 0.02 |
| Sedatives | 0.22 | 0.14 |
| **2016** |  |  |
| Benzodiazepines and z-drugs | 0.22 | 0.14 |
| Long-acting benzodiazepines | 0.08 | 0.06 |
| Clonazepam | 0.01 | 0.01 |
| Diazepam | 0.06 | 0.04 |
| Chloridiazepoxide | 0.00 | 0.01 |
| Potassium Clorazepate | 0.00 | 0.00 |
| Lorazepam | 0.00 | 0.00 |
| Bromazepam | 0.01 | 0.00 |
| Clobazam | 0.00 | 0.00 |
| Prazepam | 0.00 | 0.00 |
| Alprazolam | 0.06 | 0.03 |
| Flurazepam | 0.01 | 0.01 |
| Nitrazepam | 0.00 | 0.00 |
| Triazolam | 0.01 | 0.00 |
| Lormetazepam | 0.01 | 0.00 |
| Temazepam | 0.01 | 0.01 |
| Zopiclone | 0.06 | 0.04 |
| Zolpidem | 0.05 | 0.03 |
| Antidepressants^1^ | 0.02 | 0.02 |
| Antihistamines^2^ | 0.01 | 0.00 |
| Antipsychotics^3^ | 0.02 | 0.02 |
| Sedatives | 0.23 | 0.15 |
| **2017** |  |  |
| Benzodiazepines and z-drugs | 0.22 | 0.14 |
| Long-acting benzodiazepines | 0.08 | 0.06 |
| Clonazepam | 0.01 | 0.01 |
| Diazepam | 0.06 | 0.04 |
| Chloridiazepoxide | 0.00 | 0.01 |
| Potassium Clorazepate | 0.00 | 0.00 |
| Lorazepam | 0.00 | 0.00 |
| Bromazepam | 0.01 | 0.00 |
| Clobazam | 0.00 | 0.00 |
| Prazepam | 0.00 | 0.00 |
| Alprazolam | 0.06 | 0.03 |
| Flurazepam | 0.01 | 0.01 |
| Nitrazepam | 0.00 | 0.00 |
| Triazolam | 0.01 | 0.00 |
| Lormetazepam | 0.00 | 0.00 |
| Temazepam | 0.01 | 0.01 |
| Zopiclone | 0.06 | 0.04 |
| Zolpidem | 0.05 | 0.03 |
| Antidepressants^1^ | 0.02 | 0.02 |
| Antihistamines^2^ | 0.01 | 0.00 |
| Antipsychotics^3^ | 0.02 | 0.02 |
| Sedatives | 0.23 | 0.15 |
| **2018** |  |  |
| Benzodiazepines and z-drugs | 0.22 | 0.14 |
| Long-acting benzodiazepines | 0.08 | 0.06 |
| Clonazepam | 0.01 | 0.01 |
| Diazepam | 0.06 | 0.04 |
| Chloridiazepoxide | 0.00 | 0.01 |
| Potassium Clorazepate | 0.00 | 0.00 |
| Lorazepam | 0.00 | 0.00 |
| Bromazepam | 0.01 | 0.00 |
| Clobazam | 0.00 | 0.00 |
| Prazepam | 0.00 | 0.00 |
| Alprazolam | 0.06 | 0.03 |
| Flurazepam | 0.01 | 0.01 |
| Nitrazepam | 0.00 | 0.00 |
| Triazolam | 0.01 | 0.00 |
| Lormetazepam | 0.00 | 0.00 |
| Temazepam | 0.01 | 0.00 |
| Zopiclone | 0.06 | 0.04 |
| Zolpidem | 0.05 | 0.03 |
| Antidepressants^1^ | 0.03 | 0.02 |
| Antihistamines^2^ | 0.01 | 0.01 |
| Antipsychotics^3^ | 0.03 | 0.02 |
| Sedatives | 0.24 | 0.15 |
| **2019** |  |  |
| Benzodiazepines and z-drugs | 0.22 | 0.14 |
| Long-acting benzodiazepines | 0.08 | 0.06 |
| Clonazepam | 0.01 | 0.01 |
| Diazepam | 0.06 | 0.04 |
| Chloridiazepoxide | 0.00 | 0.01 |
| Potassium Clorazepate | 0.00 | 0.00 |
| Lorazepam | 0.01 | 0.00 |
| Bromazepam | 0.01 | 0.00 |
| Clobazam | 0.00 | 0.00 |
| Prazepam | 0.00 | 0.00 |
| Alprazolam | 0.06 | 0.03 |
| Flurazepam | 0.01 | 0.01 |
| Nitrazepam | 0.00 | 0.00 |
| Triazolam | 0.01 | 0.00 |
| Lormetazepam | 0.00 | 0.00 |
| Temazepam | 0.01 | 0.00 |
| Zopiclone | 0.06 | 0.04 |
| Zolpidem | 0.05 | 0.03 |
| Antidepressants^1^ | 0.03 | 0.02 |
| Antihistamines^2^ | 0.01 | 0.01 |
| Antipsychotics^3^ | 0.03 | 0.02 |
| Sedatives | 0.24 | 0.15 |
| **2020** |  |  |
| Benzodiazepines and z-drugs | 0.20 | 0.13 |
| Long-acting benzodiazepines | 0.07 | 0.05 |
| Clonazepam | 0.01 | 0.01 |
| Diazepam | 0.06 | 0.04 |
| Chloridiazepoxide | 0.00 | 0.01 |
| Potassium Clorazepate | 0.00 | 0.00 |
| Lorazepam | 0.00 | 0.00 |
| Bromazepam | 0.01 | 0.00 |
| Clobazam | 0.00 | 0.00 |
| Prazepam | 0.00 | 0.00 |
| Alprazolam | 0.06 | 0.03 |
| Flurazepam | 0.01 | 0.01 |
| Nitrazepam | 0.00 | 0.00 |
| Triazolam | 0.01 | 0.00 |
| Lormetazepam | 0.00 | 0.00 |
| Temazepam | 0.01 | 0.00 |
| Zopiclone | 0.05 | 0.04 |
| Zolpidem | 0.05 | 0.03 |
| Antidepressants^1^ | 0.03 | 0.02 |
| Antihistamines^2^ | 0.01 | 0.01 |
| Antipsychotics^3^ | 0.03 | 0.02 |
| Sedatives | 0.22 | 0.14 |
| **2021** |  |  |
| Benzodiazepines and z-drugs | 0.20 | 0.13 |
| Long-acting benzodiazepines | 0.07 | 0.06 |
| Clonazepam | 0.01 | 0.01 |
| Diazepam | 0.06 | 0.04 |
| Chloridiazepoxide | 0.00 | 0.01 |
| Potassium Clorazepate | 0.00 | 0.00 |
| Lorazepam | 0.01 | 0.00 |
| Bromazepam | 0.01 | 0.00 |
| Clobazam | 0.00 | 0.00 |
| Prazepam | 0.00 | 0.00 |
| Alprazolam | 0.06 | 0.03 |
| Flurazepam | 0.01 | 0.01 |
| Nitrazepam | 0.00 | 0.00 |
| Triazolam | 0.01 | 0.00 |
| Lormetazepam | 0.00 | 0.00 |
| Temazepam | 0.01 | 0.00 |
| Zopiclone | 0.05 | 0.04 |
| Zolpidem | 0.05 | 0.03 |
| Antidepressants^1^ | 0.03 | 0.02 |
| Antihistamines^2^ | 0.02 | 0.01 |
| Antipsychotics^3^ | 0.03 | 0.02 |
| Sedatives | 0.23 | 0.15 |
| **2022** |  |  |
| Benzodiazepines and z-drugs | 0.20 | 0.13 |
| Long-acting benzodiazepines | 0.07 | 0.05 |
| Clonazepam | 0.01 | 0.01 |
| Diazepam | 0.06 | 0.04 |
| Chloridiazepoxide | 0.00 | 0.01 |
| Potassium Clorazepate | 0.00 | 0.00 |
| Lorazepam | 0.01 | 0.01 |
| Bromazepam | 0.01 | 0.00 |
| Clobazam | 0.00 | 0.00 |
| Prazepam | 0.00 | 0.00 |
| Alprazolam | 0.06 | 0.03 |
| Flurazepam | 0.01 | 0.01 |
| Nitrazepam | 0.00 | 0.00 |
| Triazolam | 0.01 | 0.00 |
| Lormetazepam | 0.00 | 0.00 |
| Temazepam | 0.01 | 0.00 |
| Zopiclone | 0.05 | 0.04 |
| Zolpidem | 0.05 | 0.03 |
| Antidepressants^1^ | 0.03 | 0.02 |
| Antihistamines^2^ | 0.02 | 0.01 |
| Antipsychotics^3^ | 0.03 | 0.02 |
| Sedatives | 0.22 | 0.14 |

**Prevalence of initiations after 90 and 180 days in the GMS population by age group**

|  | **<5** | **5-11** | **12-15** | **16-24** | **25-34** | **35-44** | **45-54** | **55-64** | **65-69** | **70-74** | **75+** |
| --- | --- | --- | --- | --- | --- | --- | --- | --- | --- | --- | --- |
| 2014 |  |  |  |  |  |  |  |  |  |  |  |
| Sedative (90) | 0.001 | 0.001 | 0.002 | 0.021 | 0.053 | 0.066 | 0.080 | 0.088 | 0.086 | 0.089 | 0.099 |
| Sedative (180) | 0.000 | 0.000 | 0.001 | 0.010 | 0.023 | 0.029 | 0.036 | 0.041 | 0.041 | 0.041 | 0.047 |
| Benzodiazepines and z-drugs (90) | 0.001 | 0.001 | 0.002 | 0.019 | 0.051 | 0.065 | 0.079 | 0.087 | 0.086 | 0.089 | 0.100 |
| Benzodiazepines and z-drugs after (180) | 0.000 | 0.000 | 0.001 | 0.009 | 0.022 | 0.029 | 0.036 | 0.041 | 0.041 | 0.042 | 0.048 |
| Long-acting benzodiazepines (90) | 0.001 | 0.001 | 0.001 | 0.009 | 0.028 | 0.037 | 0.043 | 0.044 | 0.039 | 0.037 | 0.036 |
| Long-acting benzodiazepines (180) | 0.000 | 0.000 | 0.001 | 0.004 | 0.014 | 0.019 | 0.022 | 0.023 | 0.021 | 0.020 | 0.019 |
| Clonazepam (90) | 0.000 | 0.000 | 0.000 | 0.001 | 0.002 | 0.002 | 0.003 | 0.003 | 0.002 | 0.002 | 0.002 |
| Clonazepam (180) | 0.000 | 0.000 | 0.000 | 0.000 | 0.001 | 0.001 | 0.001 | 0.001 | 0.001 | 0.001 | 0.001 |
| Diazepam (90) | 0.000 | 0.000 | 0.001 | 0.008 | 0.024 | 0.031 | 0.035 | 0.035 | 0.031 | 0.030 | 0.029 |
| Diazepam (180) | 0.000 | 0.000 | 0.000 | 0.004 | 0.012 | 0.016 | 0.018 | 0.019 | 0.017 | 0.017 | 0.016 |
| Chlordiazepoxide (90) | 0.000 | 0.000 | 0.000 | 0.000 | 0.003 | 0.004 | 0.006 | 0.006 | 0.003 | 0.002 | 0.001 |
| Chlordiazepoxide (180) | 0.000 | 0.000 | 0.000 | 0.000 | 0.001 | 0.002 | 0.003 | 0.003 | 0.002 | 0.001 | 0.001 |
| Clorazepate (90) | 0.000 | 0.000 | 0.000 | 0.000 | 0.000 | 0.000 | 0.000 | 0.000 | 0.000 | 0.000 | 0.000 |
| Clorazepate (180) | 0.000 | 0.000 | 0.000 | 0.000 | 0.000 | 0.000 | 0.000 | 0.000 | 0.000 | 0.000 | 0.000 |
| Lorazepam (90) | 0.000 | 0.000 | 0.000 | 0.001 | 0.001 | 0.002 | 0.002 | 0.003 | 0.003 | 0.002 | 0.005 |
| Lorazepam (180) | 0.000 | 0.000 | 0.000 | 0.000 | 0.001 | 0.001 | 0.001 | 0.001 | 0.002 | 0.001 | 0.003 |
| Bromazepam (90) | 0.000 | 0.000 | 0.000 | 0.001 | 0.001 | 0.002 | 0.003 | 0.004 | 0.005 | 0.005 | 0.005 |
| Bromazepam (180) | 0.000 | 0.000 | 0.000 | 0.000 | 0.001 | 0.001 | 0.002 | 0.002 | 0.003 | 0.002 | 0.002 |
| Clobazam (90) | 0.000 | 0.000 | 0.000 | 0.000 | 0.001 | 0.000 | 0.001 | 0.001 | 0.001 | 0.001 | 0.001 |
| Clobazam (180) | 0.000 | 0.000 | 0.000 | 0.000 | 0.000 | 0.000 | 0.000 | 0.000 | 0.000 | 0.000 | 0.000 |
| Prazepam (90) | 0.000 | 0.000 | 0.000 | 0.000 | 0.000 | 0.000 | 0.001 | 0.001 | 0.001 | 0.001 | 0.001 |
| Prazepam (180) | 0.000 | 0.000 | 0.000 | 0.000 | 0.000 | 0.000 | 0.000 | 0.000 | 0.000 | 0.000 | 0.000 |
| Alprazolam (90) | 0.000 | 0.000 | 0.000 | 0.006 | 0.017 | 0.021 | 0.025 | 0.029 | 0.030 | 0.030 | 0.035 |
| Alprazolam (180) | 0.000 | 0.000 | 0.000 | 0.003 | 0.008 | 0.010 | 0.013 | 0.015 | 0.016 | 0.016 | 0.019 |
| Flurazepam (90) | 0.000 | 0.000 | 0.000 | 0.000 | 0.002 | 0.003 | 0.004 | 0.004 | 0.004 | 0.003 | 0.003 |
| Flurazepam (180) | 0.000 | 0.000 | 0.000 | 0.000 | 0.001 | 0.001 | 0.002 | 0.002 | 0.002 | 0.001 | 0.002 |
| Nitrazepam (90) | 0.000 | 0.000 | 0.000 | 0.000 | 0.000 | 0.000 | 0.000 | 0.001 | 0.001 | 0.001 | 0.001 |
| Nitrazepam (180) | 0.000 | 0.000 | 0.000 | 0.000 | 0.000 | 0.000 | 0.000 | 0.000 | 0.000 | 0.000 | 0.001 |
| Triazolam (90) | 0.000 | 0.000 | 0.000 | 0.000 | 0.001 | 0.001 | 0.001 | 0.002 | 0.002 | 0.003 | 0.003 |
| Triazolam (180) | 0.000 | 0.000 | 0.000 | 0.000 | 0.000 | 0.000 | 0.001 | 0.001 | 0.001 | 0.001 | 0.001 |
| Lormetazepam (90) | 0.000 | 0.000 | 0.000 | 0.000 | 0.000 | 0.001 | 0.001 | 0.002 | 0.002 | 0.003 | 0.004 |
| Lormetazepam (180) | 0.000 | 0.000 | 0.000 | 0.000 | 0.000 | 0.000 | 0.001 | 0.001 | 0.001 | 0.001 | 0.002 |
| Temazepam (90) | 0.000 | 0.000 | 0.000 | 0.000 | 0.001 | 0.002 | 0.003 | 0.004 | 0.005 | 0.005 | 0.008 |
| Temazepam (180) | 0.000 | 0.000 | 0.000 | 0.000 | 0.001 | 0.001 | 0.002 | 0.002 | 0.002 | 0.002 | 0.004 |
| Zopiclone (90) | 0.000 | 0.000 | 0.000 | 0.004 | 0.012 | 0.016 | 0.020 | 0.025 | 0.024 | 0.026 | 0.033 |
| Zopiclone (180) | 0.000 | 0.000 | 0.000 | 0.002 | 0.006 | 0.008 | 0.010 | 0.013 | 0.013 | 0.013 | 0.017 |
| Zolpidem (90) | 0.000 | 0.000 | 0.000 | 0.004 | 0.011 | 0.015 | 0.021 | 0.024 | 0.025 | 0.027 | 0.031 |
| Zolpidem (180) | 0.000 | 0.000 | 0.000 | 0.002 | 0.005 | 0.008 | 0.010 | 0.013 | 0.013 | 0.013 | 0.016 |
| Antidepressant (90) | 0.000 | 0.000 | 0.000 | 0.002 | 0.005 | 0.007 | 0.009 | 0.009 | 0.009 | 0.009 | 0.014 |
| Antidepressant (180) | 0.000 | 0.000 | 0.000 | 0.001 | 0.003 | 0.004 | 0.005 | 0.005 | 0.005 | 0.006 | 0.008 |
| Antihistamines (90) | 0.000 | 0.000 | 0.000 | 0.001 | 0.002 | 0.002 | 0.002 | 0.002 | 0.002 | 0.001 | 0.002 |
| Antihistamines (180) | 0.000 | 0.000 | 0.000 | 0.000 | 0.001 | 0.001 | 0.001 | 0.001 | 0.001 | 0.001 | 0.001 |
| Antipsychotics (90) | 0.000 | 0.000 | 0.000 | 0.003 | 0.006 | 0.006 | 0.007 | 0.006 | 0.006 | 0.007 | 0.017 |
| Antipsychotics (180) | 0.000 | 0.000 | 0.000 | 0.001 | 0.003 | 0.003 | 0.004 | 0.003 | 0.003 | 0.004 | 0.010 |
| 2015 |  |  |  |  |  |  |  |  |  |  |  |
| Sedative (90) | 0.001 | 0.001 | 0.003 | 0.042 | 0.099 | 0.112 | 0.124 | 0.123 | 0.117 | 0.120 | 0.127 |
| Sedative (180) | 0.001 | 0.001 | 0.002 | 0.033 | 0.076 | 0.085 | 0.093 | 0.092 | 0.087 | 0.090 | 0.095 |
| Benzodiazepines and z-drugs (90) | 0.001 | 0.001 | 0.003 | 0.039 | 0.095 | 0.108 | 0.121 | 0.122 | 0.116 | 0.118 | 0.128 |
| Benzodiazepines and z-drugs after (180) | 0.001 | 0.001 | 0.002 | 0.031 | 0.074 | 0.084 | 0.093 | 0.092 | 0.087 | 0.089 | 0.096 |
| Long-acting benzodiazepines (90) | 0.001 | 0.001 | 0.002 | 0.018 | 0.052 | 0.062 | 0.066 | 0.062 | 0.054 | 0.049 | 0.047 |
| Long-acting benzodiazepines (180) | 0.001 | 0.001 | 0.002 | 0.015 | 0.042 | 0.051 | 0.055 | 0.051 | 0.045 | 0.040 | 0.039 |
| Clonazepam (90) | 0.000 | 0.000 | 0.000 | 0.001 | 0.003 | 0.003 | 0.004 | 0.004 | 0.003 | 0.003 | 0.003 |
| Clonazepam (180) | 0.000 | 0.000 | 0.000 | 0.001 | 0.003 | 0.003 | 0.003 | 0.003 | 0.003 | 0.002 | 0.003 |
| Diazepam (90) | 0.001 | 0.001 | 0.002 | 0.015 | 0.045 | 0.052 | 0.054 | 0.049 | 0.044 | 0.040 | 0.038 |
| Diazepam (180) | 0.000 | 0.000 | 0.001 | 0.013 | 0.037 | 0.044 | 0.046 | 0.042 | 0.037 | 0.034 | 0.032 |
| Chlordiazepoxide (90) | 0.000 | 0.000 | 0.000 | 0.001 | 0.004 | 0.007 | 0.009 | 0.008 | 0.005 | 0.003 | 0.001 |
| Chlordiazepoxide (180) | 0.000 | 0.000 | 0.000 | 0.001 | 0.004 | 0.006 | 0.008 | 0.007 | 0.004 | 0.003 | 0.001 |
| Clorazepate (90) | 0.000 | 0.000 | 0.000 | 0.000 | 0.000 | 0.000 | 0.000 | 0.000 | 0.000 | 0.000 | 0.000 |
| Clorazepate (180) | 0.000 | 0.000 | 0.000 | 0.000 | 0.000 | 0.000 | 0.000 | 0.000 | 0.000 | 0.000 | 0.000 |
| Lorazepam (90) | 0.000 | 0.000 | 0.000 | 0.001 | 0.002 | 0.002 | 0.003 | 0.003 | 0.003 | 0.003 | 0.007 |
| Lorazepam (180) | 0.000 | 0.000 | 0.000 | 0.001 | 0.002 | 0.002 | 0.003 | 0.003 | 0.003 | 0.003 | 0.006 |
| Bromazepam (90) | 0.000 | 0.000 | 0.000 | 0.001 | 0.003 | 0.003 | 0.004 | 0.005 | 0.006 | 0.007 | 0.006 |
| Bromazepam (180) | 0.000 | 0.000 | 0.000 | 0.001 | 0.002 | 0.003 | 0.003 | 0.004 | 0.004 | 0.005 | 0.005 |
| Clobazam (90) | 0.000 | 0.000 | 0.000 | 0.000 | 0.001 | 0.001 | 0.001 | 0.001 | 0.001 | 0.001 | 0.001 |
| Clobazam (180) | 0.000 | 0.000 | 0.000 | 0.000 | 0.001 | 0.000 | 0.001 | 0.001 | 0.001 | 0.001 | 0.000 |
| Prazepam (90) | 0.000 | 0.000 | 0.000 | 0.000 | 0.001 | 0.001 | 0.001 | 0.001 | 0.001 | 0.001 | 0.001 |
| Prazepam (180) | 0.000 | 0.000 | 0.000 | 0.000 | 0.000 | 0.000 | 0.001 | 0.001 | 0.001 | 0.001 | 0.001 |
| Alprazolam (90) | 0.000 | 0.000 | 0.000 | 0.012 | 0.032 | 0.034 | 0.037 | 0.040 | 0.040 | 0.040 | 0.047 |
| Alprazolam (180) | 0.000 | 0.000 | 0.000 | 0.010 | 0.027 | 0.028 | 0.031 | 0.033 | 0.033 | 0.033 | 0.040 |
| Flurazepam (90) | 0.000 | 0.000 | 0.000 | 0.001 | 0.003 | 0.004 | 0.005 | 0.005 | 0.005 | 0.004 | 0.004 |
| Flurazepam (180) | 0.000 | 0.000 | 0.000 | 0.001 | 0.003 | 0.003 | 0.004 | 0.004 | 0.003 | 0.003 | 0.003 |
| Nitrazepam (90) | 0.000 | 0.000 | 0.000 | 0.000 | 0.000 | 0.000 | 0.001 | 0.001 | 0.001 | 0.001 | 0.002 |
| Nitrazepam (180) | 0.000 | 0.000 | 0.000 | 0.000 | 0.000 | 0.000 | 0.000 | 0.000 | 0.000 | 0.001 | 0.001 |
| Triazolam (90) | 0.000 | 0.000 | 0.000 | 0.000 | 0.001 | 0.001 | 0.002 | 0.003 | 0.003 | 0.003 | 0.004 |
| Triazolam (180) | 0.000 | 0.000 | 0.000 | 0.000 | 0.001 | 0.001 | 0.002 | 0.002 | 0.002 | 0.002 | 0.003 |
| Lormetazepam (90) | 0.000 | 0.000 | 0.000 | 0.000 | 0.001 | 0.001 | 0.002 | 0.002 | 0.003 | 0.003 | 0.005 |
| Lormetazepam (180) | 0.000 | 0.000 | 0.000 | 0.000 | 0.001 | 0.001 | 0.001 | 0.002 | 0.002 | 0.003 | 0.004 |
| Temazepam (90) | 0.000 | 0.000 | 0.000 | 0.001 | 0.002 | 0.002 | 0.004 | 0.005 | 0.005 | 0.006 | 0.009 |
| Temazepam (180) | 0.000 | 0.000 | 0.000 | 0.001 | 0.001 | 0.002 | 0.003 | 0.004 | 0.004 | 0.005 | 0.007 |
| Zopiclone (90) | 0.000 | 0.000 | 0.000 | 0.008 | 0.021 | 0.025 | 0.031 | 0.034 | 0.033 | 0.035 | 0.043 |
| Zopiclone (180) | 0.000 | 0.000 | 0.000 | 0.007 | 0.017 | 0.020 | 0.025 | 0.027 | 0.026 | 0.028 | 0.035 |
| Zolpidem (90) | 0.000 | 0.000 | 0.000 | 0.007 | 0.019 | 0.024 | 0.031 | 0.034 | 0.033 | 0.034 | 0.040 |
| Zolpidem (180) | 0.000 | 0.000 | 0.000 | 0.006 | 0.016 | 0.020 | 0.025 | 0.027 | 0.026 | 0.027 | 0.032 |
| Antidepressant (90) | 0.000 | 0.000 | 0.000 | 0.005 | 0.011 | 0.012 | 0.013 | 0.013 | 0.012 | 0.014 | 0.021 |
| Antidepressant (180) | 0.000 | 0.000 | 0.000 | 0.004 | 0.009 | 0.010 | 0.011 | 0.011 | 0.011 | 0.012 | 0.019 |
| Antihistamines (90) | 0.000 | 0.000 | 0.000 | 0.002 | 0.004 | 0.004 | 0.004 | 0.003 | 0.002 | 0.002 | 0.003 |
| Antihistamines (180) | 0.000 | 0.000 | 0.000 | 0.002 | 0.003 | 0.003 | 0.003 | 0.003 | 0.002 | 0.002 | 0.002 |
| Antipsychotics (90) | 0.000 | 0.000 | 0.000 | 0.005 | 0.010 | 0.009 | 0.010 | 0.009 | 0.008 | 0.009 | 0.022 |
| Antipsychotics (180) | 0.000 | 0.000 | 0.000 | 0.004 | 0.008 | 0.008 | 0.008 | 0.008 | 0.007 | 0.008 | 0.019 |
| 2016 |  |  |  |  |  |  |  |  |  |  |  |
| Sedative (90) | 0.001 | 0.001 | 0.003 | 0.047 | 0.119 | 0.125 | 0.131 | 0.130 | 0.121 | 0.121 | 0.130 |
| Sedative (180) | 0.001 | 0.001 | 0.003 | 0.039 | 0.097 | 0.102 | 0.104 | 0.101 | 0.091 | 0.091 | 0.099 |
| Benzodiazepines and z-drugs (90) | 0.001 | 0.001 | 0.003 | 0.042 | 0.113 | 0.121 | 0.128 | 0.128 | 0.119 | 0.119 | 0.130 |
| Benzodiazepines and z-drugs after (180) | 0.001 | 0.001 | 0.003 | 0.036 | 0.094 | 0.099 | 0.102 | 0.100 | 0.091 | 0.091 | 0.099 |
| Long-acting benzodiazepines (90) | 0.001 | 0.001 | 0.002 | 0.020 | 0.061 | 0.069 | 0.070 | 0.066 | 0.056 | 0.051 | 0.047 |
| Long-acting benzodiazepines (180) | 0.001 | 0.001 | 0.002 | 0.018 | 0.053 | 0.060 | 0.060 | 0.057 | 0.048 | 0.042 | 0.039 |
| Clonazepam (90) | 0.000 | 0.000 | 0.000 | 0.002 | 0.004 | 0.004 | 0.004 | 0.004 | 0.004 | 0.003 | 0.003 |
| Clonazepam (180) | 0.000 | 0.000 | 0.000 | 0.001 | 0.003 | 0.003 | 0.003 | 0.003 | 0.003 | 0.003 | 0.003 |
| Diazepam (90) | 0.001 | 0.001 | 0.002 | 0.017 | 0.053 | 0.059 | 0.058 | 0.053 | 0.046 | 0.041 | 0.038 |
| Diazepam (180) | 0.001 | 0.001 | 0.001 | 0.015 | 0.047 | 0.052 | 0.051 | 0.047 | 0.040 | 0.036 | 0.033 |
| Chlordiazepoxide (90) | 0.000 | 0.000 | 0.000 | 0.001 | 0.005 | 0.008 | 0.010 | 0.008 | 0.005 | 0.003 | 0.002 |
| Chlordiazepoxide (180) | 0.000 | 0.000 | 0.000 | 0.001 | 0.004 | 0.007 | 0.008 | 0.007 | 0.004 | 0.003 | 0.001 |
| Clorazepate (90) | 0.000 | 0.000 | 0.000 | 0.000 | 0.000 | 0.000 | 0.000 | 0.000 | 0.000 | 0.000 | 0.000 |
| Clorazepate (180) | 0.000 | 0.000 | 0.000 | 0.000 | 0.000 | 0.000 | 0.000 | 0.000 | 0.000 | 0.000 | 0.000 |
| Lorazepam (90) | 0.000 | 0.000 | 0.000 | 0.001 | 0.003 | 0.003 | 0.003 | 0.003 | 0.004 | 0.003 | 0.006 |
| Lorazepam (180) | 0.000 | 0.000 | 0.000 | 0.001 | 0.002 | 0.002 | 0.003 | 0.003 | 0.003 | 0.003 | 0.005 |
| Bromazepam (90) | 0.000 | 0.000 | 0.000 | 0.001 | 0.003 | 0.003 | 0.004 | 0.005 | 0.006 | 0.006 | 0.006 |
| Bromazepam (180) | 0.000 | 0.000 | 0.000 | 0.001 | 0.003 | 0.003 | 0.003 | 0.004 | 0.005 | 0.004 | 0.005 |
| Clobazam (90) | 0.000 | 0.000 | 0.000 | 0.000 | 0.001 | 0.001 | 0.001 | 0.001 | 0.001 | 0.001 | 0.001 |
| Clobazam (180) | 0.000 | 0.000 | 0.000 | 0.000 | 0.001 | 0.001 | 0.001 | 0.001 | 0.001 | 0.000 | 0.000 |
| Prazepam (90) | 0.000 | 0.000 | 0.000 | 0.000 | 0.000 | 0.001 | 0.001 | 0.001 | 0.001 | 0.001 | 0.001 |
| Prazepam (180) | 0.000 | 0.000 | 0.000 | 0.000 | 0.000 | 0.000 | 0.000 | 0.001 | 0.001 | 0.001 | 0.001 |
| Alprazolam (90) | 0.000 | 0.000 | 0.001 | 0.014 | 0.039 | 0.039 | 0.039 | 0.042 | 0.041 | 0.041 | 0.048 |
| Alprazolam (180) | 0.000 | 0.000 | 0.001 | 0.012 | 0.034 | 0.033 | 0.033 | 0.035 | 0.034 | 0.035 | 0.040 |
| Flurazepam (90) | 0.000 | 0.000 | 0.000 | 0.001 | 0.003 | 0.003 | 0.004 | 0.005 | 0.004 | 0.004 | 0.004 |
| Flurazepam (180) | 0.000 | 0.000 | 0.000 | 0.001 | 0.003 | 0.003 | 0.003 | 0.004 | 0.003 | 0.003 | 0.003 |
| Nitrazepam (90) | 0.000 | 0.000 | 0.000 | 0.000 | 0.000 | 0.000 | 0.000 | 0.001 | 0.001 | 0.001 | 0.001 |
| Nitrazepam (180) | 0.000 | 0.000 | 0.000 | 0.000 | 0.000 | 0.000 | 0.000 | 0.000 | 0.000 | 0.001 | 0.001 |
| Triazolam (90) | 0.000 | 0.000 | 0.000 | 0.000 | 0.001 | 0.001 | 0.002 | 0.003 | 0.003 | 0.003 | 0.004 |
| Triazolam (180) | 0.000 | 0.000 | 0.000 | 0.000 | 0.001 | 0.001 | 0.002 | 0.002 | 0.002 | 0.002 | 0.003 |
| Lormetazepam (90) | 0.000 | 0.000 | 0.000 | 0.000 | 0.001 | 0.001 | 0.002 | 0.002 | 0.003 | 0.003 | 0.004 |
| Lormetazepam (180) | 0.000 | 0.000 | 0.000 | 0.000 | 0.001 | 0.001 | 0.001 | 0.002 | 0.002 | 0.003 | 0.003 |
| Temazepam (90) | 0.000 | 0.000 | 0.000 | 0.001 | 0.002 | 0.003 | 0.004 | 0.004 | 0.005 | 0.006 | 0.008 |
| Temazepam (180) | 0.000 | 0.000 | 0.000 | 0.001 | 0.002 | 0.002 | 0.003 | 0.004 | 0.004 | 0.004 | 0.006 |
| Zopiclone (90) | 0.000 | 0.000 | 0.000 | 0.009 | 0.024 | 0.027 | 0.032 | 0.034 | 0.033 | 0.034 | 0.042 |
| Zopiclone (180) | 0.000 | 0.000 | 0.000 | 0.008 | 0.021 | 0.023 | 0.027 | 0.028 | 0.027 | 0.028 | 0.035 |
| Zolpidem (90) | 0.000 | 0.000 | 0.000 | 0.008 | 0.022 | 0.025 | 0.030 | 0.034 | 0.033 | 0.035 | 0.040 |
| Zolpidem (180) | 0.000 | 0.000 | 0.000 | 0.007 | 0.019 | 0.022 | 0.025 | 0.028 | 0.026 | 0.028 | 0.033 |
| Antidepressant (90) | 0.000 | 0.000 | 0.000 | 0.006 | 0.014 | 0.013 | 0.014 | 0.014 | 0.013 | 0.014 | 0.023 |
| Antidepressant (180) | 0.000 | 0.000 | 0.000 | 0.005 | 0.012 | 0.012 | 0.012 | 0.012 | 0.011 | 0.013 | 0.020 |
| Antihistamines (90) | 0.000 | 0.000 | 0.000 | 0.003 | 0.006 | 0.005 | 0.005 | 0.005 | 0.003 | 0.003 | 0.003 |
| Antihistamines (180) | 0.000 | 0.000 | 0.000 | 0.003 | 0.005 | 0.005 | 0.005 | 0.004 | 0.003 | 0.003 | 0.003 |
| Antipsychotics (90) | 0.000 | 0.000 | 0.000 | 0.007 | 0.013 | 0.010 | 0.011 | 0.009 | 0.009 | 0.010 | 0.023 |
| Antipsychotics (180) | 0.000 | 0.000 | 0.000 | 0.005 | 0.011 | 0.009 | 0.009 | 0.008 | 0.008 | 0.008 | 0.020 |
| 2017 |  |  |  |  |  |  |  |  |  |  |  |
| Sedative (90) | 0.001 | 0.002 | 0.003 | 0.046 | 0.127 | 0.134 | 0.135 | 0.130 | 0.117 | 0.115 | 0.126 |
| Sedative (180) | 0.001 | 0.001 | 0.003 | 0.039 | 0.105 | 0.110 | 0.107 | 0.101 | 0.090 | 0.088 | 0.095 |
| Benzodiazepines and z-drugs (90) | 0.001 | 0.001 | 0.003 | 0.041 | 0.118 | 0.129 | 0.130 | 0.127 | 0.115 | 0.113 | 0.124 |
| Benzodiazepines and z-drugs after (180) | 0.001 | 0.001 | 0.003 | 0.036 | 0.099 | 0.107 | 0.105 | 0.100 | 0.089 | 0.087 | 0.095 |
| Long-acting benzodiazepines (90) | 0.001 | 0.001 | 0.002 | 0.019 | 0.064 | 0.073 | 0.072 | 0.065 | 0.053 | 0.048 | 0.044 |
| Long-acting benzodiazepines (180) | 0.001 | 0.001 | 0.002 | 0.017 | 0.057 | 0.064 | 0.062 | 0.056 | 0.045 | 0.041 | 0.037 |
| Clonazepam (90) | 0.000 | 0.000 | 0.000 | 0.002 | 0.004 | 0.004 | 0.004 | 0.004 | 0.003 | 0.003 | 0.003 |
| Clonazepam (180) | 0.000 | 0.000 | 0.000 | 0.001 | 0.004 | 0.003 | 0.003 | 0.003 | 0.002 | 0.003 | 0.003 |
| Diazepam (90) | 0.000 | 0.001 | 0.002 | 0.016 | 0.055 | 0.062 | 0.060 | 0.053 | 0.043 | 0.040 | 0.037 |
| Diazepam (180) | 0.000 | 0.001 | 0.002 | 0.015 | 0.049 | 0.055 | 0.053 | 0.047 | 0.038 | 0.035 | 0.032 |
| Chlordiazepoxide (90) | 0.000 | 0.000 | 0.000 | 0.001 | 0.006 | 0.009 | 0.010 | 0.008 | 0.005 | 0.003 | 0.002 |
| Chlordiazepoxide (180) | 0.000 | 0.000 | 0.000 | 0.001 | 0.005 | 0.007 | 0.008 | 0.007 | 0.004 | 0.002 | 0.001 |
| Clorazepate (90) | 0.000 | 0.000 | 0.000 | 0.000 | 0.000 | 0.000 | 0.000 | 0.000 | 0.000 | 0.000 | 0.000 |
| Clorazepate (180) | 0.000 | 0.000 | 0.000 | 0.000 | 0.000 | 0.000 | 0.000 | 0.000 | 0.000 | 0.000 | 0.000 |
| Lorazepam (90) | 0.000 | 0.000 | 0.000 | 0.001 | 0.003 | 0.003 | 0.003 | 0.004 | 0.003 | 0.003 | 0.006 |
| Lorazepam (180) | 0.000 | 0.000 | 0.000 | 0.001 | 0.002 | 0.002 | 0.003 | 0.003 | 0.003 | 0.003 | 0.005 |
| Bromazepam (90) | 0.000 | 0.000 | 0.000 | 0.001 | 0.003 | 0.003 | 0.004 | 0.005 | 0.005 | 0.006 | 0.006 |
| Bromazepam (180) | 0.000 | 0.000 | 0.000 | 0.001 | 0.003 | 0.003 | 0.003 | 0.004 | 0.004 | 0.004 | 0.004 |
| Clobazam (90) | 0.000 | 0.000 | 0.000 | 0.000 | 0.001 | 0.001 | 0.001 | 0.001 | 0.001 | 0.001 | 0.000 |
| Clobazam (180) | 0.000 | 0.000 | 0.000 | 0.000 | 0.001 | 0.001 | 0.000 | 0.001 | 0.001 | 0.000 | 0.000 |
| Prazepam (90) | 0.000 | 0.000 | 0.000 | 0.000 | 0.001 | 0.000 | 0.001 | 0.001 | 0.001 | 0.001 | 0.001 |
| Prazepam (180) | 0.000 | 0.000 | 0.000 | 0.000 | 0.000 | 0.000 | 0.000 | 0.001 | 0.001 | 0.001 | 0.001 |
| Alprazolam (90) | 0.000 | 0.000 | 0.001 | 0.014 | 0.042 | 0.042 | 0.039 | 0.041 | 0.039 | 0.039 | 0.046 |
| Alprazolam (180) | 0.000 | 0.000 | 0.000 | 0.012 | 0.037 | 0.036 | 0.033 | 0.034 | 0.033 | 0.033 | 0.039 |
| Flurazepam (90) | 0.000 | 0.000 | 0.000 | 0.001 | 0.003 | 0.004 | 0.004 | 0.004 | 0.003 | 0.003 | 0.003 |
| Flurazepam (180) | 0.000 | 0.000 | 0.000 | 0.001 | 0.003 | 0.003 | 0.003 | 0.003 | 0.003 | 0.003 | 0.002 |
| Nitrazepam (90) | 0.000 | 0.000 | 0.000 | 0.000 | 0.000 | 0.000 | 0.000 | 0.000 | 0.001 | 0.001 | 0.001 |
| Nitrazepam (180) | 0.000 | 0.000 | 0.000 | 0.000 | 0.000 | 0.000 | 0.000 | 0.000 | 0.001 | 0.001 | 0.001 |
| Triazolam (90) | 0.000 | 0.000 | 0.000 | 0.000 | 0.001 | 0.001 | 0.002 | 0.003 | 0.003 | 0.002 | 0.003 |
| Triazolam (180) | 0.000 | 0.000 | 0.000 | 0.000 | 0.001 | 0.001 | 0.001 | 0.002 | 0.002 | 0.002 | 0.003 |
| Lormetazepam (90) | 0.000 | 0.000 | 0.000 | 0.000 | 0.001 | 0.001 | 0.001 | 0.002 | 0.002 | 0.003 | 0.004 |
| Lormetazepam (180) | 0.000 | 0.000 | 0.000 | 0.000 | 0.001 | 0.001 | 0.001 | 0.002 | 0.002 | 0.002 | 0.003 |
| Temazepam (90) | 0.000 | 0.000 | 0.000 | 0.001 | 0.002 | 0.002 | 0.003 | 0.004 | 0.004 | 0.005 | 0.007 |
| Temazepam (180) | 0.000 | 0.000 | 0.000 | 0.000 | 0.002 | 0.002 | 0.003 | 0.003 | 0.003 | 0.004 | 0.005 |
| Zopiclone (90) | 0.000 | 0.000 | 0.000 | 0.008 | 0.024 | 0.027 | 0.031 | 0.033 | 0.031 | 0.032 | 0.040 |
| Zopiclone (180) | 0.000 | 0.000 | 0.000 | 0.007 | 0.021 | 0.023 | 0.025 | 0.027 | 0.025 | 0.026 | 0.033 |
| Zolpidem (90) | 0.000 | 0.000 | 0.000 | 0.007 | 0.021 | 0.025 | 0.031 | 0.032 | 0.032 | 0.033 | 0.038 |
| Zolpidem (180) | 0.000 | 0.000 | 0.000 | 0.007 | 0.018 | 0.022 | 0.026 | 0.026 | 0.025 | 0.026 | 0.031 |
| Antidepressant (90) | 0.000 | 0.000 | 0.000 | 0.006 | 0.016 | 0.015 | 0.016 | 0.014 | 0.013 | 0.014 | 0.022 |
| Antidepressant (180) | 0.000 | 0.000 | 0.000 | 0.005 | 0.014 | 0.013 | 0.014 | 0.012 | 0.011 | 0.013 | 0.020 |
| Antihistamines (90) | 0.000 | 0.000 | 0.000 | 0.004 | 0.009 | 0.008 | 0.007 | 0.006 | 0.005 | 0.004 | 0.004 |
| Antihistamines (180) | 0.000 | 0.000 | 0.000 | 0.004 | 0.008 | 0.007 | 0.007 | 0.006 | 0.005 | 0.004 | 0.004 |
| Antipsychotics (90) | 0.000 | 0.000 | 0.000 | 0.007 | 0.015 | 0.012 | 0.012 | 0.011 | 0.009 | 0.010 | 0.023 |
| Antipsychotics (180) | 0.000 | 0.000 | 0.000 | 0.006 | 0.013 | 0.010 | 0.010 | 0.009 | 0.008 | 0.009 | 0.020 |
| 2018 |  |  |  |  |  |  |  |  |  |  |  |
| Sedative (90) | 0.001 | 0.001 | 0.004 | 0.047 | 0.132 | 0.136 | 0.130 | 0.125 | 0.117 | 0.114 | 0.123 |
| Sedative (180) | 0.001 | 0.001 | 0.003 | 0.040 | 0.110 | 0.111 | 0.104 | 0.098 | 0.090 | 0.088 | 0.094 |
| Benzodiazepines and z-drugs (90) | 0.001 | 0.001 | 0.003 | 0.041 | 0.121 | 0.129 | 0.125 | 0.122 | 0.113 | 0.112 | 0.121 |
| Benzodiazepines and z-drugs after (180) | 0.001 | 0.001 | 0.003 | 0.036 | 0.103 | 0.107 | 0.102 | 0.097 | 0.088 | 0.086 | 0.093 |
| Long-acting benzodiazepines (90) | 0.001 | 0.001 | 0.002 | 0.019 | 0.066 | 0.074 | 0.069 | 0.063 | 0.055 | 0.047 | 0.044 |
| Long-acting benzodiazepines (180) | 0.001 | 0.001 | 0.002 | 0.017 | 0.058 | 0.065 | 0.060 | 0.054 | 0.047 | 0.039 | 0.037 |
| Clonazepam (90) | 0.000 | 0.000 | 0.000 | 0.002 | 0.005 | 0.004 | 0.004 | 0.004 | 0.003 | 0.003 | 0.004 |
| Clonazepam (180) | 0.000 | 0.000 | 0.000 | 0.002 | 0.004 | 0.003 | 0.003 | 0.003 | 0.002 | 0.002 | 0.003 |
| Diazepam (90) | 0.001 | 0.001 | 0.002 | 0.016 | 0.056 | 0.063 | 0.057 | 0.051 | 0.046 | 0.039 | 0.036 |
| Diazepam (180) | 0.001 | 0.001 | 0.002 | 0.015 | 0.051 | 0.056 | 0.051 | 0.045 | 0.041 | 0.034 | 0.031 |
| Chlordiazepoxide (90) | 0.000 | 0.000 | 0.000 | 0.001 | 0.006 | 0.009 | 0.010 | 0.008 | 0.005 | 0.003 | 0.001 |
| Chlordiazepoxide (180) | 0.000 | 0.000 | 0.000 | 0.001 | 0.005 | 0.007 | 0.008 | 0.007 | 0.004 | 0.002 | 0.001 |
| Clorazepate (90) | 0.000 | 0.000 | 0.000 | 0.000 | 0.000 | 0.000 | 0.000 | 0.000 | 0.000 | 0.000 | 0.000 |
| Clorazepate (180) | 0.000 | 0.000 | 0.000 | 0.000 | 0.000 | 0.000 | 0.000 | 0.000 | 0.000 | 0.000 | 0.000 |
| Lorazepam (90) | 0.000 | 0.000 | 0.000 | 0.001 | 0.003 | 0.003 | 0.003 | 0.004 | 0.004 | 0.003 | 0.005 |
| Lorazepam (180) | 0.000 | 0.000 | 0.000 | 0.001 | 0.003 | 0.003 | 0.003 | 0.003 | 0.003 | 0.003 | 0.005 |
| Bromazepam (90) | 0.000 | 0.000 | 0.000 | 0.001 | 0.003 | 0.003 | 0.004 | 0.005 | 0.005 | 0.006 | 0.005 |
| Bromazepam (180) | 0.000 | 0.000 | 0.000 | 0.001 | 0.003 | 0.003 | 0.003 | 0.004 | 0.004 | 0.004 | 0.004 |
| Clobazam (90) | 0.000 | 0.000 | 0.000 | 0.001 | 0.001 | 0.001 | 0.001 | 0.001 | 0.000 | 0.001 | 0.000 |
| Clobazam (180) | 0.000 | 0.000 | 0.000 | 0.000 | 0.001 | 0.001 | 0.001 | 0.001 | 0.000 | 0.000 | 0.000 |
| Prazepam (90) | 0.000 | 0.000 | 0.000 | 0.000 | 0.000 | 0.000 | 0.001 | 0.001 | 0.001 | 0.001 | 0.001 |
| Prazepam (180) | 0.000 | 0.000 | 0.000 | 0.000 | 0.000 | 0.000 | 0.001 | 0.001 | 0.001 | 0.001 | 0.001 |
| Alprazolam (90) | 0.000 | 0.000 | 0.001 | 0.014 | 0.043 | 0.042 | 0.039 | 0.040 | 0.039 | 0.038 | 0.044 |
| Alprazolam (180) | 0.000 | 0.000 | 0.001 | 0.012 | 0.038 | 0.036 | 0.033 | 0.033 | 0.033 | 0.032 | 0.037 |
| Flurazepam (90) | 0.000 | 0.000 | 0.000 | 0.001 | 0.003 | 0.003 | 0.004 | 0.004 | 0.004 | 0.003 | 0.003 |
| Flurazepam (180) | 0.000 | 0.000 | 0.000 | 0.001 | 0.003 | 0.003 | 0.003 | 0.003 | 0.003 | 0.002 | 0.002 |
| Nitrazepam (90) | 0.000 | 0.000 | 0.000 | 0.000 | 0.000 | 0.000 | 0.000 | 0.000 | 0.000 | 0.001 | 0.001 |
| Nitrazepam (180) | 0.000 | 0.000 | 0.000 | 0.000 | 0.000 | 0.000 | 0.000 | 0.000 | 0.000 | 0.000 | 0.001 |
| Triazolam (90) | 0.000 | 0.000 | 0.000 | 0.000 | 0.001 | 0.001 | 0.002 | 0.002 | 0.002 | 0.002 | 0.003 |
| Triazolam (180) | 0.000 | 0.000 | 0.000 | 0.000 | 0.001 | 0.001 | 0.001 | 0.002 | 0.002 | 0.002 | 0.002 |
| Lormetazepam (90) | 0.000 | 0.000 | 0.000 | 0.000 | 0.001 | 0.001 | 0.001 | 0.001 | 0.002 | 0.003 | 0.003 |
| Lormetazepam (180) | 0.000 | 0.000 | 0.000 | 0.000 | 0.001 | 0.001 | 0.001 | 0.001 | 0.002 | 0.002 | 0.002 |
| Temazepam (90) | 0.000 | 0.000 | 0.000 | 0.000 | 0.002 | 0.002 | 0.002 | 0.003 | 0.004 | 0.004 | 0.006 |
| Temazepam (180) | 0.000 | 0.000 | 0.000 | 0.000 | 0.001 | 0.002 | 0.002 | 0.003 | 0.003 | 0.003 | 0.004 |
| Zopiclone (90) | 0.000 | 0.000 | 0.000 | 0.008 | 0.025 | 0.027 | 0.029 | 0.031 | 0.030 | 0.031 | 0.038 |
| Zopiclone (180) | 0.000 | 0.000 | 0.000 | 0.007 | 0.021 | 0.022 | 0.024 | 0.025 | 0.024 | 0.025 | 0.032 |
| Zolpidem (90) | 0.000 | 0.000 | 0.000 | 0.007 | 0.021 | 0.024 | 0.028 | 0.030 | 0.030 | 0.031 | 0.036 |
| Zolpidem (180) | 0.000 | 0.000 | 0.000 | 0.006 | 0.019 | 0.021 | 0.023 | 0.025 | 0.024 | 0.025 | 0.029 |
| Antidepressant (90) | 0.000 | 0.000 | 0.000 | 0.006 | 0.018 | 0.016 | 0.016 | 0.016 | 0.014 | 0.015 | 0.024 |
| Antidepressant (180) | 0.000 | 0.000 | 0.000 | 0.006 | 0.016 | 0.014 | 0.014 | 0.014 | 0.012 | 0.013 | 0.021 |
| Antihistamines (90) | 0.000 | 0.000 | 0.000 | 0.005 | 0.011 | 0.009 | 0.008 | 0.008 | 0.005 | 0.004 | 0.005 |
| Antihistamines (180) | 0.000 | 0.000 | 0.000 | 0.004 | 0.010 | 0.008 | 0.008 | 0.007 | 0.005 | 0.004 | 0.004 |
| Antipsychotics (90) | 0.000 | 0.000 | 0.001 | 0.008 | 0.019 | 0.014 | 0.012 | 0.011 | 0.010 | 0.009 | 0.023 |
| Antipsychotics (180) | 0.000 | 0.000 | 0.000 | 0.007 | 0.016 | 0.012 | 0.010 | 0.009 | 0.008 | 0.008 | 0.021 |
| 2019 |  |  |  |  |  |  |  |  |  |  |  |
| Sedative (90) | 0.001 | 0.001 | 0.003 | 0.048 | 0.132 | 0.134 | 0.131 | 0.125 | 0.118 | 0.111 | 0.120 |
| Sedative (180) | 0.001 | 0.001 | 0.003 | 0.040 | 0.109 | 0.109 | 0.105 | 0.098 | 0.091 | 0.086 | 0.092 |
| Benzodiazepines and z-drugs (90) | 0.001 | 0.001 | 0.003 | 0.042 | 0.120 | 0.126 | 0.124 | 0.120 | 0.115 | 0.108 | 0.117 |
| Benzodiazepines and z-drugs after (180) | 0.001 | 0.001 | 0.003 | 0.036 | 0.101 | 0.104 | 0.101 | 0.096 | 0.089 | 0.084 | 0.090 |
| Long-acting benzodiazepines (90) | 0.001 | 0.001 | 0.002 | 0.019 | 0.065 | 0.072 | 0.070 | 0.063 | 0.056 | 0.046 | 0.042 |
| Long-acting benzodiazepines (180) | 0.000 | 0.001 | 0.002 | 0.017 | 0.057 | 0.063 | 0.061 | 0.055 | 0.047 | 0.039 | 0.035 |
| Clonazepam (90) | 0.000 | 0.000 | 0.000 | 0.002 | 0.004 | 0.004 | 0.003 | 0.004 | 0.003 | 0.003 | 0.004 |
| Clonazepam (180) | 0.000 | 0.000 | 0.000 | 0.001 | 0.004 | 0.003 | 0.003 | 0.003 | 0.003 | 0.003 | 0.003 |
| Diazepam (90) | 0.000 | 0.001 | 0.002 | 0.016 | 0.056 | 0.061 | 0.058 | 0.052 | 0.047 | 0.038 | 0.034 |
| Diazepam (180) | 0.000 | 0.001 | 0.001 | 0.014 | 0.050 | 0.053 | 0.051 | 0.046 | 0.041 | 0.033 | 0.030 |
| Chlordiazepoxide (90) | 0.000 | 0.000 | 0.000 | 0.001 | 0.006 | 0.009 | 0.010 | 0.008 | 0.005 | 0.003 | 0.001 |
| Chlordiazepoxide (180) | 0.000 | 0.000 | 0.000 | 0.001 | 0.005 | 0.008 | 0.009 | 0.007 | 0.004 | 0.002 | 0.001 |
| Clorazepate (90) | 0.000 | 0.000 | 0.000 | 0.000 | 0.000 | 0.000 | 0.000 | 0.000 | 0.000 | 0.000 | 0.000 |
| Clorazepate (180) | 0.000 | 0.000 | 0.000 | 0.000 | 0.000 | 0.000 | 0.000 | 0.000 | 0.000 | 0.000 | 0.000 |
| Lorazepam (90) | 0.000 | 0.000 | 0.000 | 0.002 | 0.003 | 0.003 | 0.003 | 0.004 | 0.004 | 0.003 | 0.005 |
| Lorazepam (180) | 0.000 | 0.000 | 0.000 | 0.001 | 0.003 | 0.003 | 0.003 | 0.003 | 0.003 | 0.003 | 0.005 |
| Bromazepam (90) | 0.000 | 0.000 | 0.000 | 0.001 | 0.003 | 0.003 | 0.003 | 0.004 | 0.005 | 0.005 | 0.005 |
| Bromazepam (180) | 0.000 | 0.000 | 0.000 | 0.001 | 0.003 | 0.003 | 0.003 | 0.003 | 0.004 | 0.004 | 0.003 |
| Clobazam (90) | 0.000 | 0.000 | 0.000 | 0.001 | 0.001 | 0.001 | 0.001 | 0.001 | 0.001 | 0.000 | 0.000 |
| Clobazam (180) | 0.000 | 0.000 | 0.000 | 0.000 | 0.001 | 0.001 | 0.001 | 0.001 | 0.001 | 0.000 | 0.000 |
| Prazepam (90) | 0.000 | 0.000 | 0.000 | 0.000 | 0.000 | 0.000 | 0.000 | 0.001 | 0.001 | 0.001 | 0.001 |
| Prazepam (180) | 0.000 | 0.000 | 0.000 | 0.000 | 0.000 | 0.000 | 0.000 | 0.000 | 0.001 | 0.000 | 0.000 |
| Alprazolam (90) | 0.000 | 0.000 | 0.001 | 0.015 | 0.044 | 0.042 | 0.039 | 0.040 | 0.040 | 0.038 | 0.044 |
| Alprazolam (180) | 0.000 | 0.000 | 0.001 | 0.013 | 0.039 | 0.035 | 0.033 | 0.033 | 0.033 | 0.032 | 0.037 |
| Flurazepam (90) | 0.000 | 0.000 | 0.000 | 0.001 | 0.003 | 0.003 | 0.003 | 0.003 | 0.003 | 0.003 | 0.003 |
| Flurazepam (180) | 0.000 | 0.000 | 0.000 | 0.001 | 0.002 | 0.002 | 0.003 | 0.003 | 0.002 | 0.002 | 0.002 |
| Nitrazepam (90) | 0.000 | 0.000 | 0.000 | 0.000 | 0.000 | 0.000 | 0.000 | 0.000 | 0.001 | 0.001 | 0.001 |
| Nitrazepam (180) | 0.000 | 0.000 | 0.000 | 0.000 | 0.000 | 0.000 | 0.000 | 0.000 | 0.000 | 0.001 | 0.001 |
| Triazolam (90) | 0.000 | 0.000 | 0.000 | 0.000 | 0.001 | 0.001 | 0.002 | 0.002 | 0.002 | 0.002 | 0.003 |
| Triazolam (180) | 0.000 | 0.000 | 0.000 | 0.000 | 0.001 | 0.001 | 0.001 | 0.002 | 0.002 | 0.002 | 0.003 |
| Lormetazepam (90) | 0.000 | 0.000 | 0.000 | 0.000 | 0.000 | 0.000 | 0.001 | 0.001 | 0.001 | 0.002 | 0.002 |
| Lormetazepam (180) | 0.000 | 0.000 | 0.000 | 0.000 | 0.000 | 0.000 | 0.000 | 0.001 | 0.001 | 0.001 | 0.001 |
| Temazepam (90) | 0.000 | 0.000 | 0.000 | 0.000 | 0.002 | 0.002 | 0.002 | 0.003 | 0.003 | 0.004 | 0.006 |
| Temazepam (180) | 0.000 | 0.000 | 0.000 | 0.000 | 0.001 | 0.002 | 0.002 | 0.003 | 0.003 | 0.003 | 0.005 |
| Zopiclone (90) | 0.000 | 0.000 | 0.000 | 0.008 | 0.024 | 0.026 | 0.028 | 0.030 | 0.030 | 0.031 | 0.039 |
| Zopiclone (180) | 0.000 | 0.000 | 0.000 | 0.007 | 0.021 | 0.022 | 0.023 | 0.025 | 0.025 | 0.026 | 0.032 |
| Zolpidem (90) | 0.000 | 0.000 | 0.000 | 0.007 | 0.020 | 0.023 | 0.027 | 0.030 | 0.032 | 0.032 | 0.038 |
| Zolpidem (180) | 0.000 | 0.000 | 0.000 | 0.006 | 0.018 | 0.020 | 0.023 | 0.024 | 0.025 | 0.026 | 0.031 |
| Antidepressant (90) | 0.000 | 0.000 | 0.000 | 0.006 | 0.020 | 0.016 | 0.017 | 0.016 | 0.015 | 0.015 | 0.025 |
| Antidepressant (180) | 0.000 | 0.000 | 0.000 | 0.006 | 0.018 | 0.014 | 0.015 | 0.014 | 0.014 | 0.014 | 0.022 |
| Antihistamines (90) | 0.000 | 0.000 | 0.000 | 0.007 | 0.015 | 0.012 | 0.011 | 0.009 | 0.007 | 0.005 | 0.006 |
| Antihistamines (180) | 0.000 | 0.000 | 0.000 | 0.006 | 0.014 | 0.011 | 0.010 | 0.009 | 0.006 | 0.005 | 0.005 |
| Antipsychotics (90) | 0.000 | 0.000 | 0.000 | 0.010 | 0.021 | 0.016 | 0.013 | 0.012 | 0.010 | 0.010 | 0.024 |
| Antipsychotics (180) | 0.000 | 0.000 | 0.000 | 0.008 | 0.018 | 0.013 | 0.011 | 0.010 | 0.008 | 0.008 | 0.022 |
| 2020 |  |  |  |  |  |  |  |  |  |  |  |
| Sedative (90) | 0.001 | 0.001 | 0.003 | 0.037 | 0.116 | 0.120 | 0.118 | 0.110 | 0.104 | 0.099 | 0.111 |
| Sedative (180) | 0.000 | 0.001 | 0.002 | 0.031 | 0.096 | 0.097 | 0.095 | 0.087 | 0.081 | 0.076 | 0.087 |
| Benzodiazepines and z-drugs (90) | 0.001 | 0.001 | 0.003 | 0.031 | 0.102 | 0.111 | 0.111 | 0.105 | 0.100 | 0.096 | 0.107 |
| Benzodiazepines and z-drugs after (180) | 0.000 | 0.001 | 0.002 | 0.027 | 0.088 | 0.092 | 0.091 | 0.084 | 0.078 | 0.074 | 0.084 |
| Long-acting benzodiazepines (90) | 0.000 | 0.001 | 0.002 | 0.015 | 0.057 | 0.064 | 0.064 | 0.056 | 0.048 | 0.041 | 0.038 |
| Long-acting benzodiazepines (180) | 0.000 | 0.001 | 0.001 | 0.013 | 0.050 | 0.056 | 0.056 | 0.048 | 0.041 | 0.035 | 0.032 |
| Clonazepam (90) | 0.000 | 0.000 | 0.000 | 0.001 | 0.004 | 0.003 | 0.003 | 0.004 | 0.003 | 0.003 | 0.004 |
| Clonazepam (180) | 0.000 | 0.000 | 0.000 | 0.001 | 0.003 | 0.003 | 0.003 | 0.003 | 0.003 | 0.003 | 0.003 |
| Diazepam (90) | 0.000 | 0.001 | 0.001 | 0.012 | 0.049 | 0.055 | 0.052 | 0.045 | 0.039 | 0.035 | 0.031 |
| Diazepam (180) | 0.000 | 0.000 | 0.001 | 0.011 | 0.043 | 0.049 | 0.047 | 0.040 | 0.035 | 0.030 | 0.027 |
| Chlordiazepoxide (90) | 0.000 | 0.000 | 0.000 | 0.001 | 0.006 | 0.008 | 0.009 | 0.008 | 0.005 | 0.002 | 0.001 |
| Chlordiazepoxide (180) | 0.000 | 0.000 | 0.000 | 0.001 | 0.005 | 0.007 | 0.008 | 0.006 | 0.004 | 0.002 | 0.001 |
| Clorazepate (90) | 0.000 | 0.000 | 0.000 | 0.000 | 0.000 | 0.000 | 0.000 | 0.000 | 0.000 | 0.000 | 0.000 |
| Clorazepate (180) | 0.000 | 0.000 | 0.000 | 0.000 | 0.000 | 0.000 | 0.000 | 0.000 | 0.000 | 0.000 | 0.000 |
| Lorazepam (90) | 0.000 | 0.000 | 0.000 | 0.002 | 0.003 | 0.003 | 0.003 | 0.004 | 0.003 | 0.003 | 0.006 |
| Lorazepam (180) | 0.000 | 0.000 | 0.000 | 0.001 | 0.003 | 0.003 | 0.003 | 0.003 | 0.003 | 0.003 | 0.005 |
| Bromazepam (90) | 0.000 | 0.000 | 0.000 | 0.001 | 0.002 | 0.003 | 0.003 | 0.003 | 0.004 | 0.004 | 0.005 |
| Bromazepam (180) | 0.000 | 0.000 | 0.000 | 0.001 | 0.002 | 0.002 | 0.003 | 0.002 | 0.003 | 0.003 | 0.003 |
| Clobazam (90) | 0.000 | 0.000 | 0.000 | 0.000 | 0.001 | 0.001 | 0.001 | 0.001 | 0.000 | 0.000 | 0.000 |
| Clobazam (180) | 0.000 | 0.000 | 0.000 | 0.000 | 0.001 | 0.001 | 0.000 | 0.000 | 0.000 | 0.000 | 0.000 |
| Prazepam (90) | 0.000 | 0.000 | 0.000 | 0.000 | 0.000 | 0.000 | 0.000 | 0.000 | 0.001 | 0.001 | 0.001 |
| Prazepam (180) | 0.000 | 0.000 | 0.000 | 0.000 | 0.000 | 0.000 | 0.000 | 0.000 | 0.000 | 0.000 | 0.000 |
| Alprazolam (90) | 0.000 | 0.000 | 0.000 | 0.010 | 0.036 | 0.035 | 0.033 | 0.032 | 0.032 | 0.033 | 0.040 |
| Alprazolam (180) | 0.000 | 0.000 | 0.000 | 0.009 | 0.032 | 0.030 | 0.027 | 0.027 | 0.027 | 0.028 | 0.035 |
| Flurazepam (90) | 0.000 | 0.000 | 0.000 | 0.000 | 0.002 | 0.002 | 0.003 | 0.003 | 0.003 | 0.002 | 0.002 |
| Flurazepam (180) | 0.000 | 0.000 | 0.000 | 0.000 | 0.002 | 0.002 | 0.002 | 0.002 | 0.002 | 0.002 | 0.002 |
| Nitrazepam (90) | 0.000 | 0.000 | 0.000 | 0.000 | 0.000 | 0.000 | 0.000 | 0.000 | 0.000 | 0.000 | 0.001 |
| Nitrazepam (180) | 0.000 | 0.000 | 0.000 | 0.000 | 0.000 | 0.000 | 0.000 | 0.000 | 0.000 | 0.000 | 0.000 |
| Triazolam (90) | 0.000 | 0.000 | 0.000 | 0.000 | 0.001 | 0.001 | 0.001 | 0.002 | 0.002 | 0.002 | 0.003 |
| Triazolam (180) | 0.000 | 0.000 | 0.000 | 0.000 | 0.001 | 0.001 | 0.001 | 0.001 | 0.001 | 0.001 | 0.002 |
| Lormetazepam (90) | 0.000 | 0.000 | 0.000 | 0.000 | 0.000 | 0.000 | 0.000 | 0.000 | 0.000 | 0.000 | 0.000 |
| Lormetazepam (180) | 0.000 | 0.000 | 0.000 | 0.000 | 0.000 | 0.000 | 0.000 | 0.000 | 0.000 | 0.000 | 0.000 |
| Temazepam (90) | 0.000 | 0.000 | 0.000 | 0.000 | 0.001 | 0.002 | 0.002 | 0.003 | 0.003 | 0.003 | 0.005 |
| Temazepam (180) | 0.000 | 0.000 | 0.000 | 0.000 | 0.001 | 0.001 | 0.002 | 0.002 | 0.003 | 0.002 | 0.004 |
| Zopiclone (90) | 0.000 | 0.000 | 0.000 | 0.006 | 0.021 | 0.023 | 0.024 | 0.027 | 0.028 | 0.027 | 0.035 |
| Zopiclone (180) | 0.000 | 0.000 | 0.000 | 0.005 | 0.018 | 0.020 | 0.020 | 0.022 | 0.023 | 0.023 | 0.029 |
| Zolpidem (90) | 0.000 | 0.000 | 0.000 | 0.005 | 0.017 | 0.020 | 0.024 | 0.026 | 0.027 | 0.028 | 0.033 |
| Zolpidem (180) | 0.000 | 0.000 | 0.000 | 0.005 | 0.016 | 0.018 | 0.021 | 0.022 | 0.022 | 0.022 | 0.027 |
| Antidepressant (90) | 0.000 | 0.000 | 0.000 | 0.006 | 0.019 | 0.017 | 0.016 | 0.016 | 0.015 | 0.016 | 0.025 |
| Antidepressant (180) | 0.000 | 0.000 | 0.000 | 0.005 | 0.017 | 0.015 | 0.014 | 0.014 | 0.013 | 0.014 | 0.023 |
| Antihistamines (90) | 0.000 | 0.000 | 0.000 | 0.006 | 0.017 | 0.014 | 0.013 | 0.011 | 0.008 | 0.007 | 0.007 |
| Antihistamines (180) | 0.000 | 0.000 | 0.000 | 0.005 | 0.015 | 0.013 | 0.012 | 0.010 | 0.008 | 0.006 | 0.006 |
| Antipsychotics (90) | 0.000 | 0.000 | 0.000 | 0.009 | 0.021 | 0.016 | 0.013 | 0.012 | 0.011 | 0.011 | 0.026 |
| Antipsychotics (180) | 0.000 | 0.000 | 0.000 | 0.007 | 0.018 | 0.014 | 0.011 | 0.010 | 0.009 | 0.010 | 0.024 |
| 2021 |  |  |  |  |  |  |  |  |  |  |  |
| Sedative (90) | 0.001 | 0.001 | 0.003 | 0.039 | 0.120 | 0.125 | 0.119 | 0.110 | 0.103 | 0.095 | 0.107 |
| Sedative (180) | 0.000 | 0.001 | 0.003 | 0.033 | 0.100 | 0.103 | 0.098 | 0.089 | 0.081 | 0.075 | 0.085 |
| Benzodiazepines and z-drugs (90) | 0.001 | 0.001 | 0.003 | 0.034 | 0.105 | 0.115 | 0.112 | 0.105 | 0.098 | 0.092 | 0.103 |
| Benzodiazepines and z-drugs after (180) | 0.000 | 0.001 | 0.003 | 0.030 | 0.091 | 0.098 | 0.093 | 0.086 | 0.078 | 0.072 | 0.082 |
| Long-acting benzodiazepines (90) | 0.000 | 0.001 | 0.002 | 0.016 | 0.059 | 0.068 | 0.065 | 0.057 | 0.049 | 0.041 | 0.037 |
| Long-acting benzodiazepines (180) | 0.000 | 0.001 | 0.002 | 0.015 | 0.052 | 0.060 | 0.057 | 0.050 | 0.042 | 0.035 | 0.032 |
| Clonazepam (90) | 0.000 | 0.000 | 0.000 | 0.001 | 0.004 | 0.003 | 0.003 | 0.003 | 0.003 | 0.003 | 0.004 |
| Clonazepam (180) | 0.000 | 0.000 | 0.000 | 0.001 | 0.004 | 0.003 | 0.003 | 0.003 | 0.003 | 0.002 | 0.003 |
| Diazepam (90) | 0.000 | 0.001 | 0.001 | 0.014 | 0.050 | 0.058 | 0.053 | 0.047 | 0.040 | 0.034 | 0.031 |
| Diazepam (180) | 0.000 | 0.001 | 0.001 | 0.012 | 0.045 | 0.052 | 0.048 | 0.042 | 0.036 | 0.030 | 0.027 |
| Chlordiazepoxide (90) | 0.000 | 0.000 | 0.000 | 0.001 | 0.005 | 0.009 | 0.010 | 0.008 | 0.005 | 0.003 | 0.001 |
| Chlordiazepoxide (180) | 0.000 | 0.000 | 0.000 | 0.001 | 0.005 | 0.008 | 0.008 | 0.007 | 0.004 | 0.002 | 0.001 |
| Clorazepate (90) | 0.000 | 0.000 | 0.000 | 0.000 | 0.000 | 0.000 | 0.000 | 0.000 | 0.000 | 0.000 | 0.000 |
| Clorazepate (180) | 0.000 | 0.000 | 0.000 | 0.000 | 0.000 | 0.000 | 0.000 | 0.000 | 0.000 | 0.000 | 0.000 |
| Lorazepam (90) | 0.000 | 0.000 | 0.000 | 0.002 | 0.003 | 0.003 | 0.003 | 0.004 | 0.003 | 0.003 | 0.006 |
| Lorazepam (180) | 0.000 | 0.000 | 0.000 | 0.002 | 0.003 | 0.003 | 0.003 | 0.003 | 0.003 | 0.003 | 0.006 |
| Bromazepam (90) | 0.000 | 0.000 | 0.000 | 0.001 | 0.002 | 0.002 | 0.002 | 0.003 | 0.003 | 0.004 | 0.004 |
| Bromazepam (180) | 0.000 | 0.000 | 0.000 | 0.001 | 0.002 | 0.002 | 0.002 | 0.002 | 0.003 | 0.003 | 0.003 |
| Clobazam (90) | 0.000 | 0.000 | 0.000 | 0.001 | 0.001 | 0.001 | 0.001 | 0.001 | 0.000 | 0.000 | 0.000 |
| Clobazam (180) | 0.000 | 0.000 | 0.000 | 0.000 | 0.001 | 0.001 | 0.001 | 0.000 | 0.000 | 0.000 | 0.000 |
| Prazepam (90) | 0.000 | 0.000 | 0.000 | 0.000 | 0.000 | 0.000 | 0.000 | 0.001 | 0.001 | 0.001 | 0.000 |
| Prazepam (180) | 0.000 | 0.000 | 0.000 | 0.000 | 0.000 | 0.000 | 0.000 | 0.000 | 0.000 | 0.000 | 0.000 |
| Alprazolam (90) | 0.000 | 0.000 | 0.001 | 0.011 | 0.037 | 0.036 | 0.034 | 0.033 | 0.032 | 0.031 | 0.038 |
| Alprazolam (180) | 0.000 | 0.000 | 0.000 | 0.010 | 0.033 | 0.031 | 0.029 | 0.027 | 0.027 | 0.026 | 0.033 |
| Flurazepam (90) | 0.000 | 0.000 | 0.000 | 0.001 | 0.002 | 0.002 | 0.003 | 0.003 | 0.003 | 0.002 | 0.003 |
| Flurazepam (180) | 0.000 | 0.000 | 0.000 | 0.001 | 0.002 | 0.002 | 0.002 | 0.002 | 0.002 | 0.002 | 0.002 |
| Nitrazepam (90) | 0.000 | 0.000 | 0.000 | 0.000 | 0.000 | 0.000 | 0.000 | 0.000 | 0.000 | 0.000 | 0.000 |
| Nitrazepam (180) | 0.000 | 0.000 | 0.000 | 0.000 | 0.000 | 0.000 | 0.000 | 0.000 | 0.000 | 0.000 | 0.000 |
| Triazolam (90) | 0.000 | 0.000 | 0.000 | 0.000 | 0.001 | 0.001 | 0.001 | 0.001 | 0.001 | 0.002 | 0.002 |
| Triazolam (180) | 0.000 | 0.000 | 0.000 | 0.000 | 0.001 | 0.001 | 0.001 | 0.001 | 0.001 | 0.001 | 0.002 |
| Lormetazepam (90) | 0.000 | 0.000 | 0.000 | 0.000 | 0.000 | 0.000 | 0.000 | 0.000 | 0.000 | 0.000 | 0.000 |
| Lormetazepam (180) | 0.000 | 0.000 | 0.000 | 0.000 | 0.000 | 0.000 | 0.000 | 0.000 | 0.000 | 0.000 | 0.000 |
| Temazepam (90) | 0.000 | 0.000 | 0.000 | 0.000 | 0.001 | 0.001 | 0.002 | 0.002 | 0.003 | 0.003 | 0.005 |
| Temazepam (180) | 0.000 | 0.000 | 0.000 | 0.000 | 0.001 | 0.001 | 0.002 | 0.002 | 0.002 | 0.003 | 0.004 |
| Zopiclone (90) | 0.000 | 0.000 | 0.000 | 0.006 | 0.020 | 0.022 | 0.023 | 0.025 | 0.027 | 0.025 | 0.033 |
| Zopiclone (180) | 0.000 | 0.000 | 0.000 | 0.006 | 0.018 | 0.019 | 0.020 | 0.021 | 0.022 | 0.021 | 0.028 |
| Zolpidem (90) | 0.000 | 0.000 | 0.000 | 0.005 | 0.016 | 0.020 | 0.023 | 0.025 | 0.025 | 0.025 | 0.030 |
| Zolpidem (180) | 0.000 | 0.000 | 0.000 | 0.004 | 0.015 | 0.018 | 0.019 | 0.021 | 0.020 | 0.020 | 0.025 |
| Antidepressant (90) | 0.000 | 0.000 | 0.000 | 0.006 | 0.020 | 0.018 | 0.017 | 0.017 | 0.016 | 0.016 | 0.026 |
| Antidepressant (180) | 0.000 | 0.000 | 0.000 | 0.005 | 0.018 | 0.016 | 0.015 | 0.015 | 0.014 | 0.015 | 0.024 |
| Antihistamines (90) | 0.000 | 0.000 | 0.001 | 0.007 | 0.020 | 0.017 | 0.016 | 0.013 | 0.010 | 0.008 | 0.007 |
| Antihistamines (180) | 0.000 | 0.000 | 0.000 | 0.006 | 0.019 | 0.015 | 0.014 | 0.011 | 0.009 | 0.007 | 0.007 |
| Antipsychotics (90) | 0.000 | 0.000 | 0.000 | 0.009 | 0.023 | 0.017 | 0.014 | 0.012 | 0.011 | 0.011 | 0.025 |
| Antipsychotics (180) | 0.000 | 0.000 | 0.000 | 0.008 | 0.019 | 0.015 | 0.012 | 0.010 | 0.010 | 0.009 | 0.023 |
| 2022 |  |  |  |  |  |  |  |  |  |  |  |
| Sedative (90) | 0.000 | 0.001 | 0.003 | 0.033 | 0.109 | 0.118 | 0.117 | 0.110 | 0.103 | 0.099 | 0.107 |
| Sedative (180) | 0.000 | 0.001 | 0.002 | 0.027 | 0.090 | 0.096 | 0.096 | 0.089 | 0.081 | 0.078 | 0.085 |
| Benzodiazepines and z-drugs (90) | 0.000 | 0.001 | 0.002 | 0.029 | 0.097 | 0.109 | 0.110 | 0.105 | 0.099 | 0.095 | 0.103 |
| Benzodiazepines and z-drugs after (180) | 0.000 | 0.001 | 0.002 | 0.026 | 0.084 | 0.092 | 0.093 | 0.087 | 0.079 | 0.076 | 0.083 |
| Long-acting benzodiazepines (90) | 0.000 | 0.001 | 0.002 | 0.014 | 0.052 | 0.063 | 0.062 | 0.056 | 0.048 | 0.042 | 0.037 |
| Long-acting benzodiazepines (180) | 0.000 | 0.001 | 0.002 | 0.013 | 0.047 | 0.055 | 0.055 | 0.049 | 0.042 | 0.036 | 0.032 |
| Clonazepam (90) | 0.000 | 0.000 | 0.000 | 0.001 | 0.004 | 0.003 | 0.003 | 0.003 | 0.003 | 0.003 | 0.003 |
| Clonazepam (180) | 0.000 | 0.000 | 0.000 | 0.001 | 0.003 | 0.003 | 0.003 | 0.002 | 0.003 | 0.002 | 0.003 |
| Diazepam (90) | 0.000 | 0.001 | 0.001 | 0.013 | 0.047 | 0.056 | 0.054 | 0.049 | 0.044 | 0.038 | 0.034 |
| Diazepam (180) | 0.000 | 0.001 | 0.001 | 0.011 | 0.042 | 0.050 | 0.049 | 0.044 | 0.039 | 0.034 | 0.031 |
| Chlordiazepoxide (90) | 0.000 | 0.000 | 0.000 | 0.001 | 0.005 | 0.008 | 0.009 | 0.007 | 0.004 | 0.003 | 0.001 |
| Chlordiazepoxide (180) | 0.000 | 0.000 | 0.000 | 0.001 | 0.004 | 0.007 | 0.008 | 0.006 | 0.004 | 0.002 | 0.001 |
| Clorazepate (90) | 0.000 | 0.000 | 0.000 | 0.000 | 0.000 | 0.000 | 0.000 | 0.000 | 0.000 | 0.000 | 0.000 |
| Clorazepate (180) | 0.000 | 0.000 | 0.000 | 0.000 | 0.000 | 0.000 | 0.000 | 0.000 | 0.000 | 0.000 | 0.000 |
| Lorazepam (90) | 0.000 | 0.000 | 0.000 | 0.002 | 0.004 | 0.004 | 0.004 | 0.004 | 0.004 | 0.004 | 0.007 |
| Lorazepam (180) | 0.000 | 0.000 | 0.000 | 0.002 | 0.003 | 0.003 | 0.003 | 0.004 | 0.004 | 0.003 | 0.006 |
| Bromazepam (90) | 0.000 | 0.000 | 0.000 | 0.001 | 0.002 | 0.002 | 0.002 | 0.003 | 0.004 | 0.004 | 0.004 |
| Bromazepam (180) | 0.000 | 0.000 | 0.000 | 0.000 | 0.002 | 0.002 | 0.002 | 0.002 | 0.003 | 0.003 | 0.003 |
| Clobazam (90) | 0.000 | 0.000 | 0.000 | 0.001 | 0.001 | 0.001 | 0.001 | 0.001 | 0.001 | 0.000 | 0.001 |
| Clobazam (180) | 0.000 | 0.000 | 0.000 | 0.001 | 0.001 | 0.001 | 0.001 | 0.001 | 0.001 | 0.000 | 0.000 |
| Prazepam (90) | 0.000 | 0.000 | 0.000 | 0.000 | 0.000 | 0.000 | 0.000 | 0.000 | 0.000 | 0.000 | 0.000 |
| Prazepam (180) | 0.000 | 0.000 | 0.000 | 0.000 | 0.000 | 0.000 | 0.000 | 0.000 | 0.000 | 0.000 | 0.000 |
| Alprazolam (90) | 0.000 | 0.000 | 0.000 | 0.010 | 0.037 | 0.038 | 0.037 | 0.036 | 0.036 | 0.036 | 0.040 |
| Alprazolam (180) | 0.000 | 0.000 | 0.000 | 0.009 | 0.033 | 0.033 | 0.032 | 0.031 | 0.030 | 0.031 | 0.035 |
| Flurazepam (90) | 0.000 | 0.000 | 0.000 | 0.000 | 0.002 | 0.002 | 0.002 | 0.002 | 0.002 | 0.002 | 0.002 |
| Flurazepam (180) | 0.000 | 0.000 | 0.000 | 0.000 | 0.001 | 0.002 | 0.002 | 0.002 | 0.001 | 0.002 | 0.002 |
| Nitrazepam (90) | 0.000 | 0.000 | 0.000 | 0.000 | 0.000 | 0.000 | 0.000 | 0.000 | 0.000 | 0.000 | 0.000 |
| Nitrazepam (180) | 0.000 | 0.000 | 0.000 | 0.000 | 0.000 | 0.000 | 0.000 | 0.000 | 0.000 | 0.000 | 0.000 |
| Triazolam (90) | 0.000 | 0.000 | 0.000 | 0.000 | 0.001 | 0.001 | 0.001 | 0.001 | 0.001 | 0.001 | 0.002 |
| Triazolam (180) | 0.000 | 0.000 | 0.000 | 0.000 | 0.001 | 0.001 | 0.001 | 0.001 | 0.001 | 0.001 | 0.001 |
| Lormetazepam (90) | 0.000 | 0.000 | 0.000 | 0.000 | 0.000 | 0.000 | 0.000 | 0.000 | 0.000 | 0.000 | 0.000 |
| Lormetazepam (180) | 0.000 | 0.000 | 0.000 | 0.000 | 0.000 | 0.000 | 0.000 | 0.000 | 0.000 | 0.000 | 0.000 |
| Temazepam (90) | 0.000 | 0.000 | 0.000 | 0.000 | 0.001 | 0.001 | 0.001 | 0.002 | 0.003 | 0.003 | 0.004 |
| Temazepam (180) | 0.000 | 0.000 | 0.000 | 0.000 | 0.001 | 0.001 | 0.001 | 0.001 | 0.002 | 0.002 | 0.003 |
| Zopiclone (90) | 0.000 | 0.000 | 0.000 | 0.005 | 0.017 | 0.020 | 0.022 | 0.024 | 0.025 | 0.025 | 0.032 |
| Zopiclone (180) | 0.000 | 0.000 | 0.000 | 0.004 | 0.015 | 0.017 | 0.019 | 0.020 | 0.021 | 0.021 | 0.027 |
| Zolpidem (90) | 0.000 | 0.000 | 0.000 | 0.004 | 0.013 | 0.017 | 0.021 | 0.022 | 0.024 | 0.025 | 0.029 |
| Zolpidem (180) | 0.000 | 0.000 | 0.000 | 0.003 | 0.012 | 0.015 | 0.017 | 0.018 | 0.019 | 0.020 | 0.024 |
| Antidepressant (90) | 0.000 | 0.000 | 0.000 | 0.004 | 0.016 | 0.016 | 0.016 | 0.016 | 0.015 | 0.017 | 0.026 |
| Antidepressant (180) | 0.000 | 0.000 | 0.000 | 0.004 | 0.014 | 0.014 | 0.014 | 0.014 | 0.013 | 0.015 | 0.023 |
| Antihistamines (90) | 0.000 | 0.000 | 0.000 | 0.006 | 0.018 | 0.017 | 0.016 | 0.013 | 0.011 | 0.009 | 0.008 |
| Antihistamines (180) | 0.000 | 0.000 | 0.000 | 0.005 | 0.016 | 0.015 | 0.014 | 0.012 | 0.010 | 0.008 | 0.008 |
| Antipsychotics (90) | 0.000 | 0.000 | 0.000 | 0.008 | 0.021 | 0.017 | 0.014 | 0.012 | 0.010 | 0.010 | 0.024 |
| Antipsychotics (180) | 0.000 | 0.000 | 0.000 | 0.006 | 0.017 | 0.014 | 0.012 | 0.010 | 0.009 | 0.009 | 0.021 |

**Prevalence of initiations after 90 and 180 days in the GMS population by sex**

|  | F | M |
| --- | --- | --- |
| 2014 |  |  |
| Sedative (90) | 0.067 | 0.042 |
| Sedative (180) | 0.030 | 0.020 |
| Benzodiazepines and z-drugs (90) | 0.066 | 0.041 |
| Benzodiazepines and z-drugs after (180) | 0.030 | 0.020 |
| Long-acting benzodiazepines (90) | 0.031 | 0.021 |
| Long-acting benzodiazepines (180) | 0.016 | 0.011 |
| Clonazepam (90) | 0.002 | 0.001 |
| Clonazepam (180) | 0.001 | 0.001 |
| Diazepam (90) | 0.026 | 0.016 |
| Diazepam (180) | 0.014 | 0.008 |
| Chlordiazepoxide (90) | 0.002 | 0.003 |
| Chlordiazepoxide (180) | 0.001 | 0.002 |
| Clorazepate (90) | 0.000 | 0.000 |
| Clorazepate (180) | 0.000 | 0.000 |
| Lorazepam (90) | 0.002 | 0.002 |
| Lorazepam (180) | 0.001 | 0.001 |
| Bromazepam (90) | 0.003 | 0.001 |
| Bromazepam (180) | 0.002 | 0.001 |
| Clobazam (90) | 0.000 | 0.000 |
| Clobazam (180) | 0.000 | 0.000 |
| Prazepam (90) | 0.001 | 0.000 |
| Prazepam (180) | 0.000 | 0.000 |
| Alprazolam (90) | 0.024 | 0.011 |
| Alprazolam (180) | 0.012 | 0.006 |
| Flurazepam (90) | 0.002 | 0.002 |
| Flurazepam (180) | 0.001 | 0.001 |
| Nitrazepam (90) | 0.000 | 0.000 |
| Nitrazepam (180) | 0.000 | 0.000 |
| Triazolam (90) | 0.002 | 0.001 |
| Triazolam (180) | 0.001 | 0.000 |
| Lormetazepam (90) | 0.002 | 0.001 |
| Lormetazepam (180) | 0.001 | 0.000 |
| Temazepam (90) | 0.003 | 0.002 |
| Temazepam (180) | 0.001 | 0.001 |
| Zopiclone (90) | 0.018 | 0.012 |
| Zopiclone (180) | 0.009 | 0.006 |
| Zolpidem (90) | 0.018 | 0.011 |
| Zolpidem (180) | 0.009 | 0.005 |
| Antidepressant (90) | 0.008 | 0.005 |
| Antidepressant (180) | 0.004 | 0.003 |
| Antihistamines (90) | 0.002 | 0.001 |
| Antihistamines (180) | 0.001 | 0.001 |
| Antipsychotics (90) | 0.006 | 0.005 |
| Antipsychotics (180) | 0.004 | 0.003 |
| 2015 |  |  |
| Sedative (90) | 0.100 | 0.065 |
| Sedative (180) | 0.075 | 0.049 |
| Benzodiazepines and z-drugs (90) | 0.099 | 0.064 |
| Benzodiazepines and z-drugs after (180) | 0.075 | 0.049 |
| Long-acting benzodiazepines (90) | 0.047 | 0.033 |
| Long-acting benzodiazepines (180) | 0.039 | 0.027 |
| Clonazepam (90) | 0.003 | 0.002 |
| Clonazepam (180) | 0.002 | 0.002 |
| Diazepam (90) | 0.041 | 0.025 |
| Diazepam (180) | 0.034 | 0.021 |
| Chlordiazepoxide (90) | 0.002 | 0.005 |
| Chlordiazepoxide (180) | 0.002 | 0.004 |
| Clorazepate (90) | 0.000 | 0.000 |
| Clorazepate (180) | 0.000 | 0.000 |
| Lorazepam (90) | 0.003 | 0.002 |
| Lorazepam (180) | 0.002 | 0.002 |
| Bromazepam (90) | 0.004 | 0.002 |
| Bromazepam (180) | 0.003 | 0.002 |
| Clobazam (90) | 0.001 | 0.000 |
| Clobazam (180) | 0.000 | 0.000 |
| Prazepam (90) | 0.001 | 0.000 |
| Prazepam (180) | 0.001 | 0.000 |
| Alprazolam (90) | 0.037 | 0.017 |
| Alprazolam (180) | 0.030 | 0.014 |
| Flurazepam (90) | 0.003 | 0.003 |
| Flurazepam (180) | 0.002 | 0.002 |
| Nitrazepam (90) | 0.001 | 0.000 |
| Nitrazepam (180) | 0.000 | 0.000 |
| Triazolam (90) | 0.002 | 0.001 |
| Triazolam (180) | 0.002 | 0.001 |
| Lormetazepam (90) | 0.002 | 0.001 |
| Lormetazepam (180) | 0.002 | 0.001 |
| Temazepam (90) | 0.004 | 0.003 |
| Temazepam (180) | 0.003 | 0.002 |
| Zopiclone (90) | 0.026 | 0.018 |
| Zopiclone (180) | 0.021 | 0.014 |
| Zolpidem (90) | 0.026 | 0.016 |
| Zolpidem (180) | 0.021 | 0.013 |
| Antidepressant (90) | 0.012 | 0.008 |
| Antidepressant (180) | 0.010 | 0.007 |
| Antihistamines (90) | 0.003 | 0.002 |
| Antihistamines (180) | 0.002 | 0.002 |
| Antipsychotics (90) | 0.009 | 0.008 |
| Antipsychotics (180) | 0.008 | 0.006 |
| 2016 |  |  |
| Sedative (90) | 0.108 | 0.072 |
| Sedative (180) | 0.085 | 0.057 |
| Benzodiazepines and z-drugs (90) | 0.106 | 0.069 |
| Benzodiazepines and z-drugs after (180) | 0.084 | 0.055 |
| Long-acting benzodiazepines (90) | 0.051 | 0.036 |
| Long-acting benzodiazepines (180) | 0.043 | 0.030 |
| Clonazepam (90) | 0.003 | 0.002 |
| Clonazepam (180) | 0.002 | 0.002 |
| Diazepam (90) | 0.044 | 0.027 |
| Diazepam (180) | 0.039 | 0.024 |
| Chlordiazepoxide (90) | 0.003 | 0.006 |
| Chlordiazepoxide (180) | 0.002 | 0.005 |
| Clorazepate (90) | 0.000 | 0.000 |
| Clorazepate (180) | 0.000 | 0.000 |
| Lorazepam (90) | 0.003 | 0.002 |
| Lorazepam (180) | 0.002 | 0.002 |
| Bromazepam (90) | 0.005 | 0.002 |
| Bromazepam (180) | 0.004 | 0.002 |
| Clobazam (90) | 0.001 | 0.000 |
| Clobazam (180) | 0.000 | 0.000 |
| Prazepam (90) | 0.001 | 0.000 |
| Prazepam (180) | 0.001 | 0.000 |
| Alprazolam (90) | 0.040 | 0.018 |
| Alprazolam (180) | 0.034 | 0.016 |
| Flurazepam (90) | 0.003 | 0.003 |
| Flurazepam (180) | 0.002 | 0.002 |
| Nitrazepam (90) | 0.001 | 0.000 |
| Nitrazepam (180) | 0.000 | 0.000 |
| Triazolam (90) | 0.002 | 0.001 |
| Triazolam (180) | 0.002 | 0.001 |
| Lormetazepam (90) | 0.002 | 0.001 |
| Lormetazepam (180) | 0.002 | 0.001 |
| Temazepam (90) | 0.004 | 0.003 |
| Temazepam (180) | 0.003 | 0.002 |
| Zopiclone (90) | 0.027 | 0.019 |
| Zopiclone (180) | 0.022 | 0.016 |
| Zolpidem (90) | 0.027 | 0.016 |
| Zolpidem (180) | 0.023 | 0.014 |
| Antidepressant (90) | 0.013 | 0.009 |
| Antidepressant (180) | 0.012 | 0.008 |
| Antihistamines (90) | 0.004 | 0.003 |
| Antihistamines (180) | 0.004 | 0.002 |
| Antipsychotics (90) | 0.010 | 0.009 |
| Antipsychotics (180) | 0.009 | 0.007 |
| 2017 |  |  |
| Sedative (90) | 0.110 | 0.073 |
| Sedative (180) | 0.086 | 0.059 |
| Benzodiazepines and z-drugs (90) | 0.107 | 0.070 |
| Benzodiazepines and z-drugs after (180) | 0.084 | 0.057 |
| Long-acting benzodiazepines (90) | 0.051 | 0.036 |
| Long-acting benzodiazepines (180) | 0.044 | 0.031 |
| Clonazepam (90) | 0.003 | 0.002 |
| Clonazepam (180) | 0.002 | 0.002 |
| Diazepam (90) | 0.045 | 0.027 |
| Diazepam (180) | 0.039 | 0.024 |
| Chlordiazepoxide (90) | 0.003 | 0.006 |
| Chlordiazepoxide (180) | 0.002 | 0.005 |
| Clorazepate (90) | 0.000 | 0.000 |
| Clorazepate (180) | 0.000 | 0.000 |
| Lorazepam (90) | 0.003 | 0.003 |
| Lorazepam (180) | 0.002 | 0.002 |
| Bromazepam (90) | 0.004 | 0.002 |
| Bromazepam (180) | 0.003 | 0.002 |
| Clobazam (90) | 0.001 | 0.000 |
| Clobazam (180) | 0.000 | 0.000 |
| Prazepam (90) | 0.001 | 0.000 |
| Prazepam (180) | 0.000 | 0.000 |
| Alprazolam (90) | 0.040 | 0.019 |
| Alprazolam (180) | 0.034 | 0.016 |
| Flurazepam (90) | 0.003 | 0.003 |
| Flurazepam (180) | 0.002 | 0.002 |
| Nitrazepam (90) | 0.000 | 0.000 |
| Nitrazepam (180) | 0.000 | 0.000 |
| Triazolam (90) | 0.002 | 0.001 |
| Triazolam (180) | 0.001 | 0.001 |
| Lormetazepam (90) | 0.002 | 0.001 |
| Lormetazepam (180) | 0.001 | 0.001 |
| Temazepam (90) | 0.003 | 0.002 |
| Temazepam (180) | 0.002 | 0.002 |
| Zopiclone (90) | 0.026 | 0.019 |
| Zopiclone (180) | 0.022 | 0.016 |
| Zolpidem (90) | 0.027 | 0.016 |
| Zolpidem (180) | 0.022 | 0.014 |
| Antidepressant (90) | 0.014 | 0.009 |
| Antidepressant (180) | 0.012 | 0.008 |
| Antihistamines (90) | 0.006 | 0.004 |
| Antihistamines (180) | 0.005 | 0.003 |
| Antipsychotics (90) | 0.011 | 0.009 |
| Antipsychotics (180) | 0.010 | 0.008 |
| 2018 |  |  |
| Sedative (90) | 0.110 | 0.073 |
| Sedative (180) | 0.087 | 0.058 |
| Benzodiazepines and z-drugs (90) | 0.106 | 0.069 |
| Benzodiazepines and z-drugs after (180) | 0.085 | 0.056 |
| Long-acting benzodiazepines (90) | 0.051 | 0.035 |
| Long-acting benzodiazepines (180) | 0.044 | 0.030 |
| Clonazepam (90) | 0.003 | 0.002 |
| Clonazepam (180) | 0.002 | 0.002 |
| Diazepam (90) | 0.044 | 0.027 |
| Diazepam (180) | 0.039 | 0.024 |
| Chlordiazepoxide (90) | 0.003 | 0.006 |
| Chlordiazepoxide (180) | 0.002 | 0.005 |
| Clorazepate (90) | 0.000 | 0.000 |
| Clorazepate (180) | 0.000 | 0.000 |
| Lorazepam (90) | 0.003 | 0.003 |
| Lorazepam (180) | 0.003 | 0.002 |
| Bromazepam (90) | 0.004 | 0.002 |
| Bromazepam (180) | 0.003 | 0.002 |
| Clobazam (90) | 0.001 | 0.000 |
| Clobazam (180) | 0.000 | 0.000 |
| Prazepam (90) | 0.001 | 0.000 |
| Prazepam (180) | 0.000 | 0.000 |
| Alprazolam (90) | 0.040 | 0.018 |
| Alprazolam (180) | 0.034 | 0.016 |
| Flurazepam (90) | 0.002 | 0.002 |
| Flurazepam (180) | 0.002 | 0.002 |
| Nitrazepam (90) | 0.000 | 0.000 |
| Nitrazepam (180) | 0.000 | 0.000 |
| Triazolam (90) | 0.002 | 0.001 |
| Triazolam (180) | 0.001 | 0.001 |
| Lormetazepam (90) | 0.002 | 0.001 |
| Lormetazepam (180) | 0.001 | 0.001 |
| Temazepam (90) | 0.003 | 0.002 |
| Temazepam (180) | 0.002 | 0.002 |
| Zopiclone (90) | 0.025 | 0.018 |
| Zopiclone (180) | 0.021 | 0.015 |
| Zolpidem (90) | 0.026 | 0.016 |
| Zolpidem (180) | 0.021 | 0.013 |
| Antidepressant (90) | 0.015 | 0.010 |
| Antidepressant (180) | 0.013 | 0.009 |
| Antihistamines (90) | 0.006 | 0.004 |
| Antihistamines (180) | 0.006 | 0.004 |
| Antipsychotics (90) | 0.012 | 0.010 |
| Antipsychotics (180) | 0.011 | 0.009 |
| 2019 |  |  |
| Sedative (90) | 0.109 | 0.072 |
| Sedative (180) | 0.086 | 0.058 |
| Benzodiazepines and z-drugs (90) | 0.105 | 0.068 |
| Benzodiazepines and z-drugs after (180) | 0.083 | 0.056 |
| Long-acting benzodiazepines (90) | 0.050 | 0.035 |
| Long-acting benzodiazepines (180) | 0.043 | 0.030 |
| Clonazepam (90) | 0.003 | 0.002 |
| Clonazepam (180) | 0.002 | 0.002 |
| Diazepam (90) | 0.044 | 0.026 |
| Diazepam (180) | 0.039 | 0.023 |
| Chlordiazepoxide (90) | 0.003 | 0.006 |
| Chlordiazepoxide (180) | 0.002 | 0.006 |
| Clorazepate (90) | 0.000 | 0.000 |
| Clorazepate (180) | 0.000 | 0.000 |
| Lorazepam (90) | 0.003 | 0.003 |
| Lorazepam (180) | 0.003 | 0.002 |
| Bromazepam (90) | 0.004 | 0.002 |
| Bromazepam (180) | 0.003 | 0.001 |
| Clobazam (90) | 0.001 | 0.000 |
| Clobazam (180) | 0.000 | 0.000 |
| Prazepam (90) | 0.001 | 0.000 |
| Prazepam (180) | 0.000 | 0.000 |
| Alprazolam (90) | 0.040 | 0.018 |
| Alprazolam (180) | 0.034 | 0.016 |
| Flurazepam (90) | 0.002 | 0.002 |
| Flurazepam (180) | 0.002 | 0.002 |
| Nitrazepam (90) | 0.000 | 0.000 |
| Nitrazepam (180) | 0.000 | 0.000 |
| Triazolam (90) | 0.002 | 0.001 |
| Triazolam (180) | 0.001 | 0.001 |
| Lormetazepam (90) | 0.001 | 0.001 |
| Lormetazepam (180) | 0.001 | 0.000 |
| Temazepam (90) | 0.003 | 0.002 |
| Temazepam (180) | 0.002 | 0.002 |
| Zopiclone (90) | 0.026 | 0.018 |
| Zopiclone (180) | 0.021 | 0.015 |
| Zolpidem (90) | 0.026 | 0.016 |
| Zolpidem (180) | 0.022 | 0.013 |
| Antidepressant (90) | 0.016 | 0.011 |
| Antidepressant (180) | 0.014 | 0.010 |
| Antihistamines (90) | 0.008 | 0.006 |
| Antihistamines (180) | 0.008 | 0.005 |
| Antipsychotics (90) | 0.013 | 0.011 |
| Antipsychotics (180) | 0.011 | 0.009 |
| 2020 |  |  |
| Sedative (90) | 0.098 | 0.064 |
| Sedative (180) | 0.077 | 0.052 |
| Benzodiazepines and z-drugs (90) | 0.092 | 0.060 |
| Benzodiazepines and z-drugs after (180) | 0.074 | 0.050 |
| Long-acting benzodiazepines (90) | 0.045 | 0.031 |
| Long-acting benzodiazepines (180) | 0.039 | 0.027 |
| Clonazepam (90) | 0.003 | 0.002 |
| Clonazepam (180) | 0.002 | 0.002 |
| Diazepam (90) | 0.039 | 0.023 |
| Diazepam (180) | 0.035 | 0.021 |
| Chlordiazepoxide (90) | 0.003 | 0.006 |
| Chlordiazepoxide (180) | 0.002 | 0.005 |
| Clorazepate (90) | 0.000 | 0.000 |
| Clorazepate (180) | 0.000 | 0.000 |
| Lorazepam (90) | 0.003 | 0.003 |
| Lorazepam (180) | 0.003 | 0.002 |
| Bromazepam (90) | 0.003 | 0.001 |
| Bromazepam (180) | 0.003 | 0.001 |
| Clobazam (90) | 0.000 | 0.000 |
| Clobazam (180) | 0.000 | 0.000 |
| Prazepam (90) | 0.000 | 0.000 |
| Prazepam (180) | 0.000 | 0.000 |
| Alprazolam (90) | 0.034 | 0.015 |
| Alprazolam (180) | 0.029 | 0.013 |
| Flurazepam (90) | 0.002 | 0.002 |
| Flurazepam (180) | 0.001 | 0.001 |
| Nitrazepam (90) | 0.000 | 0.000 |
| Nitrazepam (180) | 0.000 | 0.000 |
| Triazolam (90) | 0.001 | 0.001 |
| Triazolam (180) | 0.001 | 0.001 |
| Lormetazepam (90) | 0.000 | 0.000 |
| Lormetazepam (180) | 0.000 | 0.000 |
| Temazepam (90) | 0.002 | 0.002 |
| Temazepam (180) | 0.002 | 0.001 |
| Zopiclone (90) | 0.022 | 0.016 |
| Zopiclone (180) | 0.019 | 0.014 |
| Zolpidem (90) | 0.023 | 0.014 |
| Zolpidem (180) | 0.019 | 0.012 |
| Antidepressant (90) | 0.016 | 0.010 |
| Antidepressant (180) | 0.014 | 0.009 |
| Antihistamines (90) | 0.010 | 0.006 |
| Antihistamines (180) | 0.009 | 0.006 |
| Antipsychotics (90) | 0.014 | 0.011 |
| Antipsychotics (180) | 0.012 | 0.009 |
| 2021 |  |  |
| Sedative (90) | 0.099 | 0.065 |
| Sedative (180) | 0.080 | 0.053 |
| Benzodiazepines and z-drugs (90) | 0.093 | 0.060 |
| Benzodiazepines and z-drugs after (180) | 0.076 | 0.051 |
| Long-acting benzodiazepines (90) | 0.046 | 0.032 |
| Long-acting benzodiazepines (180) | 0.040 | 0.028 |
| Clonazepam (90) | 0.003 | 0.002 |
| Clonazepam (180) | 0.003 | 0.002 |
| Diazepam (90) | 0.040 | 0.024 |
| Diazepam (180) | 0.036 | 0.022 |
| Chlordiazepoxide (90) | 0.003 | 0.006 |
| Chlordiazepoxide (180) | 0.002 | 0.005 |
| Clorazepate (90) | 0.000 | 0.000 |
| Clorazepate (180) | 0.000 | 0.000 |
| Lorazepam (90) | 0.003 | 0.003 |
| Lorazepam (180) | 0.003 | 0.003 |
| Bromazepam (90) | 0.003 | 0.001 |
| Bromazepam (180) | 0.002 | 0.001 |
| Clobazam (90) | 0.001 | 0.000 |
| Clobazam (180) | 0.000 | 0.000 |
| Prazepam (90) | 0.000 | 0.000 |
| Prazepam (180) | 0.000 | 0.000 |
| Alprazolam (90) | 0.035 | 0.015 |
| Alprazolam (180) | 0.030 | 0.013 |
| Flurazepam (90) | 0.002 | 0.002 |
| Flurazepam (180) | 0.002 | 0.002 |
| Nitrazepam (90) | 0.000 | 0.000 |
| Nitrazepam (180) | 0.000 | 0.000 |
| Triazolam (90) | 0.001 | 0.001 |
| Triazolam (180) | 0.001 | 0.001 |
| Lormetazepam (90) | 0.000 | 0.000 |
| Lormetazepam (180) | 0.000 | 0.000 |
| Temazepam (90) | 0.002 | 0.002 |
| Temazepam (180) | 0.002 | 0.001 |
| Zopiclone (90) | 0.022 | 0.015 |
| Zopiclone (180) | 0.018 | 0.013 |
| Zolpidem (90) | 0.022 | 0.013 |
| Zolpidem (180) | 0.018 | 0.011 |
| Antidepressant (90) | 0.017 | 0.011 |
| Antidepressant (180) | 0.015 | 0.010 |
| Antihistamines (90) | 0.012 | 0.007 |
| Antihistamines (180) | 0.011 | 0.007 |
| Antipsychotics (90) | 0.014 | 0.011 |
| Antipsychotics (180) | 0.012 | 0.010 |
| 2022 |  |  |
| Sedative (90) | 0.097 | 0.064 |
| Sedative (180) | 0.078 | 0.052 |
| Benzodiazepines and z-drugs (90) | 0.092 | 0.060 |
| Benzodiazepines and z-drugs after (180) | 0.075 | 0.050 |
| Long-acting benzodiazepines (90) | 0.044 | 0.031 |
| Long-acting benzodiazepines (180) | 0.039 | 0.027 |
| Clonazepam (90) | 0.003 | 0.002 |
| Clonazepam (180) | 0.002 | 0.002 |
| Diazepam (90) | 0.041 | 0.025 |
| Diazepam (180) | 0.037 | 0.023 |
| Chlordiazepoxide (90) | 0.002 | 0.006 |
| Chlordiazepoxide (180) | 0.002 | 0.005 |
| Clorazepate (90) | 0.000 | 0.000 |
| Clorazepate (180) | 0.000 | 0.000 |
| Lorazepam (90) | 0.004 | 0.003 |
| Lorazepam (180) | 0.003 | 0.003 |
| Bromazepam (90) | 0.003 | 0.001 |
| Bromazepam (180) | 0.002 | 0.001 |
| Clobazam (90) | 0.001 | 0.001 |
| Clobazam (180) | 0.001 | 0.000 |
| Prazepam (90) | 0.000 | 0.000 |
| Prazepam (180) | 0.000 | 0.000 |
| Alprazolam (90) | 0.037 | 0.017 |
| Alprazolam (180) | 0.032 | 0.015 |
| Flurazepam (90) | 0.001 | 0.001 |
| Flurazepam (180) | 0.001 | 0.001 |
| Nitrazepam (90) | 0.000 | 0.000 |
| Nitrazepam (180) | 0.000 | 0.000 |
| Triazolam (90) | 0.001 | 0.001 |
| Triazolam (180) | 0.001 | 0.001 |
| Lormetazepam (90) | 0.000 | 0.000 |
| Lormetazepam (180) | 0.000 | 0.000 |
| Temazepam (90) | 0.002 | 0.001 |
| Temazepam (180) | 0.001 | 0.001 |
| Zopiclone (90) | 0.020 | 0.015 |
| Zopiclone (180) | 0.017 | 0.013 |
| Zolpidem (90) | 0.020 | 0.012 |
| Zolpidem (180) | 0.016 | 0.010 |
| Antidepressant (90) | 0.016 | 0.011 |
| Antidepressant (180) | 0.014 | 0.009 |
| Antihistamines (90) | 0.012 | 0.007 |
| Antihistamines (180) | 0.011 | 0.007 |
| Antipsychotics (90) | 0.014 | 0.011 |
| Antipsychotics (180) | 0.012 | 0.009 |

**Prevalence of discontinuations after 90 and 180 days in the GMS population by age group**

|  | <5 | 5-11 | 12-15 | 16-24 | 25-34 | 35-44 | 45-54 | 55-64 | 65-69 | 70-74 | 75+ |
| --- | --- | --- | --- | --- | --- | --- | --- | --- | --- | --- | --- |
| 2014 |  |  |  |  |  |  |  |  |  |  |  |
| Sedative (90) | 0.001 | 0.000 | 0.001 | 0.016 | 0.039 | 0.050 | 0.063 | 0.071 | 0.068 | 0.072 | 0.074 |
| Sedative (180) | 0.000 | 0.000 | 0.001 | 0.007 | 0.018 | 0.023 | 0.030 | 0.034 | 0.033 | 0.033 | 0.034 |
| Benzodiazepines and z-drugs (90) | 0.001 | 0.000 | 0.001 | 0.016 | 0.040 | 0.051 | 0.064 | 0.071 | 0.069 | 0.073 | 0.080 |
| Benzodiazepines and z-drugs after (180) | 0.000 | 0.000 | 0.001 | 0.008 | 0.018 | 0.024 | 0.031 | 0.034 | 0.034 | 0.035 | 0.037 |
| Long-acting benzodiazepines (90) | 0.000 | 0.000 | 0.001 | 0.009 | 0.025 | 0.033 | 0.040 | 0.040 | 0.036 | 0.034 | 0.033 |
| Long-acting benzodiazepines (180) | 0.000 | 0.000 | 0.000 | 0.004 | 0.012 | 0.017 | 0.020 | 0.021 | 0.019 | 0.018 | 0.017 |
| Clonazepam (90) | 0.000 | 0.000 | 0.000 | 0.001 | 0.002 | 0.002 | 0.002 | 0.002 | 0.002 | 0.002 | 0.002 |
| Clonazepam (180) | 0.000 | 0.000 | 0.000 | 0.000 | 0.001 | 0.001 | 0.001 | 0.001 | 0.001 | 0.001 | 0.001 |
| Diazepam (90) | 0.000 | 0.000 | 0.001 | 0.008 | 0.022 | 0.028 | 0.032 | 0.032 | 0.029 | 0.028 | 0.026 |
| Diazepam (180) | 0.000 | 0.000 | 0.000 | 0.004 | 0.011 | 0.015 | 0.017 | 0.017 | 0.015 | 0.015 | 0.014 |
| Chlordiazepoxide (90) | 0.000 | 0.000 | 0.000 | 0.000 | 0.002 | 0.004 | 0.006 | 0.005 | 0.003 | 0.002 | 0.001 |
| Chlordiazepoxide (180) | 0.000 | 0.000 | 0.000 | 0.000 | 0.001 | 0.002 | 0.003 | 0.003 | 0.002 | 0.001 | 0.001 |
| Clorazepate (90) | 0.000 | 0.000 | 0.000 | 0.000 | 0.000 | 0.000 | 0.000 | 0.000 | 0.000 | 0.000 | 0.000 |
| Clorazepate (180) | 0.000 | 0.000 | 0.000 | 0.000 | 0.000 | 0.000 | 0.000 | 0.000 | 0.000 | 0.000 | 0.000 |
| Lorazepam (90) | 0.000 | 0.000 | 0.000 | 0.000 | 0.001 | 0.002 | 0.002 | 0.002 | 0.002 | 0.002 | 0.004 |
| Lorazepam (180) | 0.000 | 0.000 | 0.000 | 0.000 | 0.001 | 0.001 | 0.001 | 0.001 | 0.001 | 0.001 | 0.002 |
| Bromazepam (90) | 0.000 | 0.000 | 0.000 | 0.001 | 0.001 | 0.002 | 0.003 | 0.003 | 0.004 | 0.005 | 0.005 |
| Bromazepam (180) | 0.000 | 0.000 | 0.000 | 0.000 | 0.001 | 0.001 | 0.001 | 0.002 | 0.002 | 0.002 | 0.002 |
| Clobazam (90) | 0.000 | 0.000 | 0.000 | 0.000 | 0.001 | 0.000 | 0.001 | 0.001 | 0.001 | 0.001 | 0.000 |
| Clobazam (180) | 0.000 | 0.000 | 0.000 | 0.000 | 0.000 | 0.000 | 0.000 | 0.000 | 0.000 | 0.000 | 0.000 |
| Prazepam (90) | 0.000 | 0.000 | 0.000 | 0.000 | 0.000 | 0.000 | 0.001 | 0.001 | 0.001 | 0.001 | 0.001 |
| Prazepam (180) | 0.000 | 0.000 | 0.000 | 0.000 | 0.000 | 0.000 | 0.000 | 0.000 | 0.000 | 0.001 | 0.000 |
| Alprazolam (90) | 0.000 | 0.000 | 0.000 | 0.005 | 0.014 | 0.018 | 0.021 | 0.025 | 0.025 | 0.026 | 0.028 |
| Alprazolam (180) | 0.000 | 0.000 | 0.000 | 0.002 | 0.007 | 0.009 | 0.011 | 0.013 | 0.013 | 0.014 | 0.014 |
| Flurazepam (90) | 0.000 | 0.000 | 0.000 | 0.000 | 0.002 | 0.003 | 0.003 | 0.004 | 0.004 | 0.003 | 0.004 |
| Flurazepam (180) | 0.000 | 0.000 | 0.000 | 0.000 | 0.001 | 0.001 | 0.002 | 0.002 | 0.002 | 0.002 | 0.002 |
| Nitrazepam (90) | 0.000 | 0.000 | 0.000 | 0.000 | 0.000 | 0.000 | 0.000 | 0.001 | 0.001 | 0.001 | 0.001 |
| Nitrazepam (180) | 0.000 | 0.000 | 0.000 | 0.000 | 0.000 | 0.000 | 0.000 | 0.000 | 0.000 | 0.000 | 0.001 |
| Triazolam (90) | 0.000 | 0.000 | 0.000 | 0.000 | 0.001 | 0.001 | 0.001 | 0.002 | 0.002 | 0.002 | 0.003 |
| Triazolam (180) | 0.000 | 0.000 | 0.000 | 0.000 | 0.000 | 0.000 | 0.001 | 0.001 | 0.001 | 0.001 | 0.002 |
| Lormetazepam (90) | 0.000 | 0.000 | 0.000 | 0.000 | 0.001 | 0.001 | 0.001 | 0.002 | 0.002 | 0.003 | 0.004 |
| Lormetazepam (180) | 0.000 | 0.000 | 0.000 | 0.000 | 0.000 | 0.000 | 0.001 | 0.001 | 0.001 | 0.001 | 0.002 |
| Temazepam (90) | 0.000 | 0.000 | 0.000 | 0.000 | 0.001 | 0.002 | 0.003 | 0.004 | 0.004 | 0.005 | 0.007 |
| Temazepam (180) | 0.000 | 0.000 | 0.000 | 0.000 | 0.001 | 0.001 | 0.002 | 0.002 | 0.002 | 0.003 | 0.004 |
| Zopiclone (90) | 0.000 | 0.000 | 0.000 | 0.004 | 0.010 | 0.014 | 0.018 | 0.021 | 0.019 | 0.021 | 0.026 |
| Zopiclone (180) | 0.000 | 0.000 | 0.000 | 0.002 | 0.006 | 0.007 | 0.010 | 0.011 | 0.010 | 0.011 | 0.013 |
| Zolpidem (90) | 0.000 | 0.000 | 0.000 | 0.004 | 0.011 | 0.014 | 0.019 | 0.022 | 0.022 | 0.023 | 0.027 |
| Zolpidem (180) | 0.000 | 0.000 | 0.000 | 0.002 | 0.006 | 0.008 | 0.010 | 0.012 | 0.012 | 0.012 | 0.014 |
| Antidepressant (90) | 0.000 | 0.000 | 0.000 | 0.002 | 0.005 | 0.006 | 0.008 | 0.008 | 0.007 | 0.007 | 0.010 |
| Antidepressant (180) | 0.000 | 0.000 | 0.000 | 0.001 | 0.003 | 0.003 | 0.004 | 0.004 | 0.004 | 0.004 | 0.006 |
| Antihistamines (90) | 0.000 | 0.000 | 0.000 | 0.001 | 0.002 | 0.001 | 0.002 | 0.002 | 0.001 | 0.001 | 0.001 |
| Antihistamines (180) | 0.000 | 0.000 | 0.000 | 0.000 | 0.001 | 0.001 | 0.001 | 0.001 | 0.001 | 0.001 | 0.001 |
| Antipsychotics (90) | 0.000 | 0.000 | 0.000 | 0.002 | 0.005 | 0.005 | 0.006 | 0.005 | 0.004 | 0.005 | 0.009 |
| Antipsychotics (180) | 0.000 | 0.000 | 0.000 | 0.001 | 0.003 | 0.003 | 0.003 | 0.003 | 0.002 | 0.003 | 0.005 |
| 2015 |  |  |  |  |  |  |  |  |  |  |  |
| Sedative (90) | 0.001 | 0.000 | 0.002 | 0.019 | 0.050 | 0.061 | 0.074 | 0.080 | 0.076 | 0.078 | 0.080 |
| Sedative (180) | 0.000 | 0.000 | 0.001 | 0.011 | 0.029 | 0.037 | 0.045 | 0.047 | 0.045 | 0.045 | 0.045 |
| Benzodiazepines and z-drugs (90) | 0.001 | 0.000 | 0.002 | 0.019 | 0.052 | 0.064 | 0.076 | 0.082 | 0.078 | 0.080 | 0.087 |
| Benzodiazepines and z-drugs after (180) | 0.000 | 0.000 | 0.001 | 0.012 | 0.032 | 0.040 | 0.048 | 0.050 | 0.048 | 0.048 | 0.052 |
| Long-acting benzodiazepines (90) | 0.001 | 0.000 | 0.001 | 0.011 | 0.034 | 0.043 | 0.050 | 0.049 | 0.044 | 0.040 | 0.039 |
| Long-acting benzodiazepines (180) | 0.000 | 0.000 | 0.001 | 0.007 | 0.024 | 0.031 | 0.037 | 0.036 | 0.032 | 0.029 | 0.028 |
| Clonazepam (90) | 0.000 | 0.000 | 0.000 | 0.001 | 0.002 | 0.003 | 0.003 | 0.003 | 0.003 | 0.002 | 0.002 |
| Clonazepam (180) | 0.000 | 0.000 | 0.000 | 0.001 | 0.002 | 0.002 | 0.002 | 0.002 | 0.002 | 0.002 | 0.002 |
| Diazepam (90) | 0.000 | 0.000 | 0.001 | 0.009 | 0.029 | 0.038 | 0.041 | 0.040 | 0.036 | 0.033 | 0.031 |
| Diazepam (180) | 0.000 | 0.000 | 0.001 | 0.006 | 0.021 | 0.027 | 0.031 | 0.030 | 0.027 | 0.024 | 0.023 |
| Chlordiazepoxide (90) | 0.000 | 0.000 | 0.000 | 0.001 | 0.003 | 0.005 | 0.007 | 0.007 | 0.004 | 0.003 | 0.001 |
| Chlordiazepoxide (180) | 0.000 | 0.000 | 0.000 | 0.000 | 0.002 | 0.004 | 0.006 | 0.006 | 0.003 | 0.002 | 0.001 |
| Clorazepate (90) | 0.000 | 0.000 | 0.000 | 0.000 | 0.000 | 0.000 | 0.000 | 0.000 | 0.000 | 0.000 | 0.000 |
| Clorazepate (180) | 0.000 | 0.000 | 0.000 | 0.000 | 0.000 | 0.000 | 0.000 | 0.000 | 0.000 | 0.000 | 0.000 |
| Lorazepam (90) | 0.000 | 0.000 | 0.000 | 0.001 | 0.002 | 0.002 | 0.003 | 0.003 | 0.003 | 0.002 | 0.005 |
| Lorazepam (180) | 0.000 | 0.000 | 0.000 | 0.001 | 0.001 | 0.002 | 0.002 | 0.002 | 0.002 | 0.002 | 0.004 |
| Bromazepam (90) | 0.000 | 0.000 | 0.000 | 0.001 | 0.002 | 0.002 | 0.003 | 0.004 | 0.005 | 0.006 | 0.006 |
| Bromazepam (180) | 0.000 | 0.000 | 0.000 | 0.001 | 0.002 | 0.002 | 0.002 | 0.003 | 0.004 | 0.004 | 0.004 |
| Clobazam (90) | 0.000 | 0.000 | 0.000 | 0.000 | 0.001 | 0.001 | 0.001 | 0.001 | 0.001 | 0.001 | 0.001 |
| Clobazam (180) | 0.000 | 0.000 | 0.000 | 0.000 | 0.000 | 0.001 | 0.000 | 0.001 | 0.000 | 0.000 | 0.000 |
| Prazepam (90) | 0.000 | 0.000 | 0.000 | 0.000 | 0.000 | 0.000 | 0.001 | 0.001 | 0.001 | 0.001 | 0.001 |
| Prazepam (180) | 0.000 | 0.000 | 0.000 | 0.000 | 0.000 | 0.000 | 0.000 | 0.001 | 0.001 | 0.001 | 0.001 |
| Alprazolam (90) | 0.000 | 0.000 | 0.000 | 0.007 | 0.022 | 0.024 | 0.028 | 0.031 | 0.031 | 0.031 | 0.035 |
| Alprazolam (180) | 0.000 | 0.000 | 0.000 | 0.005 | 0.016 | 0.018 | 0.021 | 0.023 | 0.023 | 0.022 | 0.025 |
| Flurazepam (90) | 0.000 | 0.000 | 0.000 | 0.001 | 0.003 | 0.003 | 0.004 | 0.005 | 0.004 | 0.004 | 0.004 |
| Flurazepam (180) | 0.000 | 0.000 | 0.000 | 0.001 | 0.002 | 0.003 | 0.003 | 0.004 | 0.003 | 0.003 | 0.003 |
| Nitrazepam (90) | 0.000 | 0.000 | 0.000 | 0.000 | 0.000 | 0.000 | 0.001 | 0.001 | 0.001 | 0.001 | 0.001 |
| Nitrazepam (180) | 0.000 | 0.000 | 0.000 | 0.000 | 0.000 | 0.000 | 0.000 | 0.000 | 0.000 | 0.001 | 0.001 |
| Triazolam (90) | 0.000 | 0.000 | 0.000 | 0.000 | 0.001 | 0.001 | 0.002 | 0.002 | 0.002 | 0.003 | 0.003 |
| Triazolam (180) | 0.000 | 0.000 | 0.000 | 0.000 | 0.001 | 0.001 | 0.001 | 0.002 | 0.001 | 0.002 | 0.002 |
| Lormetazepam (90) | 0.000 | 0.000 | 0.000 | 0.000 | 0.001 | 0.001 | 0.001 | 0.002 | 0.002 | 0.003 | 0.004 |
| Lormetazepam (180) | 0.000 | 0.000 | 0.000 | 0.000 | 0.001 | 0.001 | 0.001 | 0.002 | 0.002 | 0.002 | 0.003 |
| Temazepam (90) | 0.000 | 0.000 | 0.000 | 0.000 | 0.001 | 0.002 | 0.003 | 0.004 | 0.005 | 0.005 | 0.007 |
| Temazepam (180) | 0.000 | 0.000 | 0.000 | 0.000 | 0.001 | 0.002 | 0.003 | 0.003 | 0.004 | 0.004 | 0.005 |
| Zopiclone (90) | 0.000 | 0.000 | 0.000 | 0.006 | 0.016 | 0.019 | 0.023 | 0.026 | 0.024 | 0.026 | 0.030 |
| Zopiclone (180) | 0.000 | 0.000 | 0.000 | 0.004 | 0.012 | 0.015 | 0.017 | 0.019 | 0.017 | 0.017 | 0.021 |
| Zolpidem (90) | 0.000 | 0.000 | 0.000 | 0.005 | 0.015 | 0.018 | 0.024 | 0.027 | 0.025 | 0.026 | 0.030 |
| Zolpidem (180) | 0.000 | 0.000 | 0.000 | 0.004 | 0.011 | 0.014 | 0.018 | 0.019 | 0.018 | 0.019 | 0.022 |
| Antidepressant (90) | 0.000 | 0.000 | 0.000 | 0.003 | 0.009 | 0.009 | 0.011 | 0.010 | 0.009 | 0.010 | 0.014 |
| Antidepressant (180) | 0.000 | 0.000 | 0.000 | 0.003 | 0.007 | 0.007 | 0.009 | 0.009 | 0.008 | 0.008 | 0.010 |
| Antihistamines (90) | 0.000 | 0.000 | 0.000 | 0.001 | 0.003 | 0.003 | 0.003 | 0.003 | 0.002 | 0.002 | 0.002 |
| Antihistamines (180) | 0.000 | 0.000 | 0.000 | 0.001 | 0.002 | 0.002 | 0.002 | 0.002 | 0.002 | 0.001 | 0.002 |
| Antipsychotics (90) | 0.000 | 0.000 | 0.000 | 0.004 | 0.008 | 0.007 | 0.008 | 0.007 | 0.006 | 0.006 | 0.012 |
| Antipsychotics (180) | 0.000 | 0.000 | 0.000 | 0.003 | 0.006 | 0.006 | 0.006 | 0.006 | 0.004 | 0.005 | 0.009 |
| 2016 |  |  |  |  |  |  |  |  |  |  |  |
| Sedative (90) | 0.001 | 0.001 | 0.001 | 0.019 | 0.056 | 0.065 | 0.076 | 0.082 | 0.080 | 0.080 | 0.082 |
| Sedative (180) | 0.000 | 0.000 | 0.001 | 0.011 | 0.034 | 0.040 | 0.047 | 0.050 | 0.047 | 0.047 | 0.047 |
| Benzodiazepines and z-drugs (90) | 0.001 | 0.001 | 0.001 | 0.020 | 0.059 | 0.068 | 0.078 | 0.084 | 0.081 | 0.082 | 0.089 |
| Benzodiazepines and z-drugs after (180) | 0.000 | 0.000 | 0.001 | 0.013 | 0.038 | 0.044 | 0.051 | 0.053 | 0.049 | 0.050 | 0.054 |
| Long-acting benzodiazepines (90) | 0.000 | 0.000 | 0.001 | 0.011 | 0.039 | 0.046 | 0.050 | 0.051 | 0.045 | 0.041 | 0.038 |
| Long-acting benzodiazepines (180) | 0.000 | 0.000 | 0.001 | 0.008 | 0.028 | 0.034 | 0.038 | 0.037 | 0.033 | 0.030 | 0.028 |
| Clonazepam (90) | 0.000 | 0.000 | 0.000 | 0.001 | 0.003 | 0.003 | 0.003 | 0.003 | 0.003 | 0.002 | 0.002 |
| Clonazepam (180) | 0.000 | 0.000 | 0.000 | 0.001 | 0.002 | 0.002 | 0.002 | 0.002 | 0.002 | 0.001 | 0.002 |
| Diazepam (90) | 0.000 | 0.000 | 0.001 | 0.010 | 0.034 | 0.040 | 0.042 | 0.042 | 0.037 | 0.034 | 0.031 |
| Diazepam (180) | 0.000 | 0.000 | 0.001 | 0.007 | 0.025 | 0.030 | 0.032 | 0.032 | 0.028 | 0.026 | 0.023 |
| Chlordiazepoxide (90) | 0.000 | 0.000 | 0.000 | 0.001 | 0.004 | 0.006 | 0.008 | 0.007 | 0.005 | 0.003 | 0.001 |
| Chlordiazepoxide (180) | 0.000 | 0.000 | 0.000 | 0.001 | 0.003 | 0.005 | 0.006 | 0.005 | 0.004 | 0.002 | 0.001 |
| Clorazepate (90) | 0.000 | 0.000 | 0.000 | 0.000 | 0.000 | 0.000 | 0.000 | 0.000 | 0.000 | 0.000 | 0.000 |
| Clorazepate (180) | 0.000 | 0.000 | 0.000 | 0.000 | 0.000 | 0.000 | 0.000 | 0.000 | 0.000 | 0.000 | 0.000 |
| Lorazepam (90) | 0.000 | 0.000 | 0.000 | 0.001 | 0.002 | 0.002 | 0.003 | 0.003 | 0.003 | 0.003 | 0.004 |
| Lorazepam (180) | 0.000 | 0.000 | 0.000 | 0.001 | 0.002 | 0.002 | 0.002 | 0.002 | 0.002 | 0.002 | 0.003 |
| Bromazepam (90) | 0.000 | 0.000 | 0.000 | 0.001 | 0.002 | 0.003 | 0.003 | 0.005 | 0.005 | 0.005 | 0.005 |
| Bromazepam (180) | 0.000 | 0.000 | 0.000 | 0.001 | 0.002 | 0.002 | 0.002 | 0.003 | 0.004 | 0.004 | 0.004 |
| Clobazam (90) | 0.000 | 0.000 | 0.000 | 0.000 | 0.001 | 0.000 | 0.001 | 0.001 | 0.001 | 0.001 | 0.001 |
| Clobazam (180) | 0.000 | 0.000 | 0.000 | 0.000 | 0.000 | 0.000 | 0.000 | 0.000 | 0.000 | 0.000 | 0.000 |
| Prazepam (90) | 0.000 | 0.000 | 0.000 | 0.000 | 0.000 | 0.000 | 0.001 | 0.001 | 0.001 | 0.001 | 0.001 |
| Prazepam (180) | 0.000 | 0.000 | 0.000 | 0.000 | 0.000 | 0.000 | 0.000 | 0.001 | 0.001 | 0.001 | 0.001 |
| Alprazolam (90) | 0.000 | 0.000 | 0.000 | 0.008 | 0.025 | 0.027 | 0.029 | 0.032 | 0.032 | 0.032 | 0.036 |
| Alprazolam (180) | 0.000 | 0.000 | 0.000 | 0.006 | 0.019 | 0.020 | 0.022 | 0.023 | 0.024 | 0.023 | 0.026 |
| Flurazepam (90) | 0.000 | 0.000 | 0.000 | 0.001 | 0.003 | 0.003 | 0.004 | 0.004 | 0.004 | 0.003 | 0.004 |
| Flurazepam (180) | 0.000 | 0.000 | 0.000 | 0.001 | 0.002 | 0.003 | 0.003 | 0.003 | 0.003 | 0.003 | 0.003 |
| Nitrazepam (90) | 0.000 | 0.000 | 0.000 | 0.000 | 0.000 | 0.000 | 0.000 | 0.000 | 0.001 | 0.001 | 0.001 |
| Nitrazepam (180) | 0.000 | 0.000 | 0.000 | 0.000 | 0.000 | 0.000 | 0.000 | 0.000 | 0.000 | 0.001 | 0.001 |
| Triazolam (90) | 0.000 | 0.000 | 0.000 | 0.000 | 0.001 | 0.001 | 0.002 | 0.002 | 0.002 | 0.002 | 0.003 |
| Triazolam (180) | 0.000 | 0.000 | 0.000 | 0.000 | 0.001 | 0.001 | 0.001 | 0.001 | 0.001 | 0.002 | 0.002 |
| Lormetazepam (90) | 0.000 | 0.000 | 0.000 | 0.000 | 0.001 | 0.001 | 0.001 | 0.002 | 0.002 | 0.003 | 0.004 |
| Lormetazepam (180) | 0.000 | 0.000 | 0.000 | 0.000 | 0.001 | 0.001 | 0.001 | 0.001 | 0.002 | 0.002 | 0.003 |
| Temazepam (90) | 0.000 | 0.000 | 0.000 | 0.000 | 0.002 | 0.002 | 0.003 | 0.004 | 0.004 | 0.005 | 0.007 |
| Temazepam (180) | 0.000 | 0.000 | 0.000 | 0.000 | 0.001 | 0.002 | 0.003 | 0.003 | 0.003 | 0.004 | 0.005 |
| Zopiclone (90) | 0.000 | 0.000 | 0.000 | 0.006 | 0.018 | 0.019 | 0.024 | 0.027 | 0.025 | 0.026 | 0.032 |
| Zopiclone (180) | 0.000 | 0.000 | 0.000 | 0.004 | 0.014 | 0.015 | 0.018 | 0.020 | 0.018 | 0.018 | 0.023 |
| Zolpidem (90) | 0.000 | 0.000 | 0.000 | 0.005 | 0.016 | 0.019 | 0.023 | 0.026 | 0.026 | 0.026 | 0.031 |
| Zolpidem (180) | 0.000 | 0.000 | 0.000 | 0.004 | 0.013 | 0.015 | 0.018 | 0.019 | 0.018 | 0.018 | 0.022 |
| Antidepressant (90) | 0.000 | 0.000 | 0.000 | 0.004 | 0.011 | 0.010 | 0.012 | 0.011 | 0.011 | 0.011 | 0.016 |
| Antidepressant (180) | 0.000 | 0.000 | 0.000 | 0.003 | 0.009 | 0.009 | 0.009 | 0.009 | 0.009 | 0.009 | 0.012 |
| Antihistamines (90) | 0.000 | 0.000 | 0.000 | 0.002 | 0.004 | 0.004 | 0.004 | 0.003 | 0.002 | 0.002 | 0.002 |
| Antihistamines (180) | 0.000 | 0.000 | 0.000 | 0.002 | 0.003 | 0.003 | 0.003 | 0.003 | 0.002 | 0.002 | 0.002 |
| Antipsychotics (90) | 0.000 | 0.000 | 0.000 | 0.004 | 0.010 | 0.008 | 0.008 | 0.007 | 0.007 | 0.006 | 0.013 |
| Antipsychotics (180) | 0.000 | 0.000 | 0.000 | 0.003 | 0.008 | 0.006 | 0.006 | 0.006 | 0.005 | 0.005 | 0.009 |
| 2017 |  |  |  |  |  |  |  |  |  |  |  |
| Sedative (90) | 0.001 | 0.001 | 0.001 | 0.018 | 0.057 | 0.068 | 0.076 | 0.081 | 0.077 | 0.076 | 0.079 |
| Sedative (180) | 0.000 | 0.000 | 0.001 | 0.011 | 0.036 | 0.042 | 0.047 | 0.050 | 0.046 | 0.045 | 0.046 |
| Benzodiazepines and z-drugs (90) | 0.001 | 0.001 | 0.001 | 0.019 | 0.060 | 0.071 | 0.079 | 0.084 | 0.079 | 0.078 | 0.086 |
| Benzodiazepines and z-drugs after (180) | 0.000 | 0.000 | 0.001 | 0.013 | 0.040 | 0.046 | 0.051 | 0.053 | 0.049 | 0.048 | 0.054 |
| Long-acting benzodiazepines (90) | 0.000 | 0.000 | 0.001 | 0.011 | 0.039 | 0.048 | 0.052 | 0.051 | 0.042 | 0.038 | 0.037 |
| Long-acting benzodiazepines (180) | 0.000 | 0.000 | 0.001 | 0.008 | 0.030 | 0.036 | 0.039 | 0.039 | 0.032 | 0.029 | 0.029 |
| Clonazepam (90) | 0.000 | 0.000 | 0.000 | 0.001 | 0.003 | 0.003 | 0.003 | 0.003 | 0.002 | 0.002 | 0.002 |
| Clonazepam (180) | 0.000 | 0.000 | 0.000 | 0.001 | 0.002 | 0.002 | 0.002 | 0.002 | 0.002 | 0.002 | 0.002 |
| Diazepam (90) | 0.000 | 0.000 | 0.001 | 0.009 | 0.035 | 0.041 | 0.044 | 0.042 | 0.036 | 0.032 | 0.031 |
| Diazepam (180) | 0.000 | 0.000 | 0.001 | 0.006 | 0.027 | 0.031 | 0.033 | 0.033 | 0.028 | 0.025 | 0.024 |
| Chlordiazepoxide (90) | 0.000 | 0.000 | 0.000 | 0.001 | 0.004 | 0.006 | 0.008 | 0.007 | 0.004 | 0.003 | 0.001 |
| Chlordiazepoxide (180) | 0.000 | 0.000 | 0.000 | 0.000 | 0.003 | 0.005 | 0.006 | 0.005 | 0.003 | 0.002 | 0.001 |
| Clorazepate (90) | 0.000 | 0.000 | 0.000 | 0.000 | 0.000 | 0.000 | 0.000 | 0.000 | 0.000 | 0.000 | 0.000 |
| Clorazepate (180) | 0.000 | 0.000 | 0.000 | 0.000 | 0.000 | 0.000 | 0.000 | 0.000 | 0.000 | 0.000 | 0.000 |
| Lorazepam (90) | 0.000 | 0.000 | 0.000 | 0.001 | 0.002 | 0.002 | 0.003 | 0.003 | 0.003 | 0.002 | 0.004 |
| Lorazepam (180) | 0.000 | 0.000 | 0.000 | 0.001 | 0.002 | 0.002 | 0.002 | 0.002 | 0.002 | 0.002 | 0.003 |
| Bromazepam (90) | 0.000 | 0.000 | 0.000 | 0.001 | 0.002 | 0.003 | 0.003 | 0.004 | 0.005 | 0.005 | 0.005 |
| Bromazepam (180) | 0.000 | 0.000 | 0.000 | 0.001 | 0.002 | 0.002 | 0.002 | 0.003 | 0.003 | 0.003 | 0.004 |
| Clobazam (90) | 0.000 | 0.000 | 0.000 | 0.000 | 0.001 | 0.000 | 0.001 | 0.001 | 0.001 | 0.001 | 0.000 |
| Clobazam (180) | 0.000 | 0.000 | 0.000 | 0.000 | 0.000 | 0.000 | 0.001 | 0.000 | 0.000 | 0.000 | 0.000 |
| Prazepam (90) | 0.000 | 0.000 | 0.000 | 0.000 | 0.000 | 0.000 | 0.001 | 0.001 | 0.001 | 0.001 | 0.001 |
| Prazepam (180) | 0.000 | 0.000 | 0.000 | 0.000 | 0.000 | 0.000 | 0.000 | 0.001 | 0.000 | 0.001 | 0.001 |
| Alprazolam (90) | 0.000 | 0.000 | 0.000 | 0.008 | 0.027 | 0.028 | 0.029 | 0.032 | 0.032 | 0.031 | 0.035 |
| Alprazolam (180) | 0.000 | 0.000 | 0.000 | 0.006 | 0.021 | 0.022 | 0.022 | 0.024 | 0.023 | 0.023 | 0.026 |
| Flurazepam (90) | 0.000 | 0.000 | 0.000 | 0.001 | 0.003 | 0.003 | 0.003 | 0.004 | 0.003 | 0.003 | 0.003 |
| Flurazepam (180) | 0.000 | 0.000 | 0.000 | 0.000 | 0.002 | 0.003 | 0.003 | 0.003 | 0.003 | 0.002 | 0.002 |
| Nitrazepam (90) | 0.000 | 0.000 | 0.000 | 0.000 | 0.000 | 0.000 | 0.000 | 0.000 | 0.000 | 0.001 | 0.001 |
| Nitrazepam (180) | 0.000 | 0.000 | 0.000 | 0.000 | 0.000 | 0.000 | 0.000 | 0.000 | 0.000 | 0.000 | 0.001 |
| Triazolam (90) | 0.000 | 0.000 | 0.000 | 0.000 | 0.001 | 0.001 | 0.002 | 0.002 | 0.002 | 0.002 | 0.003 |
| Triazolam (180) | 0.000 | 0.000 | 0.000 | 0.000 | 0.001 | 0.001 | 0.001 | 0.002 | 0.002 | 0.001 | 0.002 |
| Lormetazepam (90) | 0.000 | 0.000 | 0.000 | 0.000 | 0.001 | 0.001 | 0.001 | 0.002 | 0.002 | 0.003 | 0.004 |
| Lormetazepam (180) | 0.000 | 0.000 | 0.000 | 0.000 | 0.001 | 0.001 | 0.001 | 0.001 | 0.002 | 0.002 | 0.002 |
| Temazepam (90) | 0.000 | 0.000 | 0.000 | 0.000 | 0.002 | 0.002 | 0.003 | 0.003 | 0.004 | 0.004 | 0.006 |
| Temazepam (180) | 0.000 | 0.000 | 0.000 | 0.000 | 0.001 | 0.002 | 0.002 | 0.003 | 0.003 | 0.003 | 0.005 |
| Zopiclone (90) | 0.000 | 0.000 | 0.000 | 0.005 | 0.018 | 0.020 | 0.023 | 0.026 | 0.024 | 0.024 | 0.029 |
| Zopiclone (180) | 0.000 | 0.000 | 0.000 | 0.004 | 0.014 | 0.015 | 0.018 | 0.019 | 0.018 | 0.017 | 0.021 |
| Zolpidem (90) | 0.000 | 0.000 | 0.000 | 0.005 | 0.016 | 0.019 | 0.023 | 0.025 | 0.025 | 0.025 | 0.030 |
| Zolpidem (180) | 0.000 | 0.000 | 0.000 | 0.004 | 0.013 | 0.015 | 0.018 | 0.019 | 0.017 | 0.018 | 0.021 |
| Antidepressant (90) | 0.000 | 0.000 | 0.000 | 0.004 | 0.012 | 0.012 | 0.012 | 0.012 | 0.010 | 0.010 | 0.015 |
| Antidepressant (180) | 0.000 | 0.000 | 0.000 | 0.003 | 0.010 | 0.009 | 0.010 | 0.010 | 0.008 | 0.008 | 0.012 |
| Antihistamines (90) | 0.000 | 0.000 | 0.000 | 0.002 | 0.006 | 0.006 | 0.005 | 0.005 | 0.004 | 0.003 | 0.003 |
| Antihistamines (180) | 0.000 | 0.000 | 0.000 | 0.002 | 0.005 | 0.004 | 0.004 | 0.004 | 0.003 | 0.002 | 0.002 |
| Antipsychotics (90) | 0.000 | 0.000 | 0.000 | 0.005 | 0.011 | 0.009 | 0.009 | 0.008 | 0.006 | 0.006 | 0.012 |
| Antipsychotics (180) | 0.000 | 0.000 | 0.000 | 0.004 | 0.009 | 0.007 | 0.007 | 0.006 | 0.005 | 0.005 | 0.009 |
| 2018 |  |  |  |  |  |  |  |  |  |  |  |
| Sedative (90) | 0.000 | 0.000 | 0.001 | 0.018 | 0.059 | 0.068 | 0.073 | 0.078 | 0.074 | 0.074 | 0.077 |
| Sedative (180) | 0.000 | 0.000 | 0.001 | 0.011 | 0.037 | 0.042 | 0.045 | 0.047 | 0.045 | 0.043 | 0.045 |
| Benzodiazepines and z-drugs (90) | 0.000 | 0.000 | 0.002 | 0.020 | 0.062 | 0.071 | 0.076 | 0.080 | 0.076 | 0.076 | 0.084 |
| Benzodiazepines and z-drugs after (180) | 0.000 | 0.000 | 0.001 | 0.013 | 0.042 | 0.046 | 0.050 | 0.051 | 0.048 | 0.047 | 0.052 |
| Long-acting benzodiazepines (90) | 0.000 | 0.000 | 0.001 | 0.010 | 0.041 | 0.048 | 0.049 | 0.048 | 0.043 | 0.038 | 0.035 |
| Long-acting benzodiazepines (180) | 0.000 | 0.000 | 0.001 | 0.007 | 0.030 | 0.036 | 0.037 | 0.036 | 0.032 | 0.028 | 0.027 |
| Clonazepam (90) | 0.000 | 0.000 | 0.000 | 0.001 | 0.003 | 0.003 | 0.003 | 0.003 | 0.002 | 0.002 | 0.002 |
| Clonazepam (180) | 0.000 | 0.000 | 0.000 | 0.001 | 0.003 | 0.003 | 0.002 | 0.002 | 0.002 | 0.002 | 0.002 |
| Diazepam (90) | 0.000 | 0.000 | 0.001 | 0.009 | 0.036 | 0.041 | 0.041 | 0.040 | 0.036 | 0.032 | 0.030 |
| Diazepam (180) | 0.000 | 0.000 | 0.001 | 0.006 | 0.027 | 0.031 | 0.032 | 0.030 | 0.028 | 0.024 | 0.023 |
| Chlordiazepoxide (90) | 0.000 | 0.000 | 0.000 | 0.001 | 0.004 | 0.006 | 0.007 | 0.007 | 0.004 | 0.002 | 0.001 |
| Chlordiazepoxide (180) | 0.000 | 0.000 | 0.000 | 0.000 | 0.003 | 0.005 | 0.006 | 0.005 | 0.003 | 0.002 | 0.001 |
| Clorazepate (90) | 0.000 | 0.000 | 0.000 | 0.000 | 0.000 | 0.000 | 0.000 | 0.000 | 0.000 | 0.000 | 0.000 |
| Clorazepate (180) | 0.000 | 0.000 | 0.000 | 0.000 | 0.000 | 0.000 | 0.000 | 0.000 | 0.000 | 0.000 | 0.000 |
| Lorazepam (90) | 0.000 | 0.000 | 0.000 | 0.001 | 0.003 | 0.002 | 0.003 | 0.003 | 0.002 | 0.002 | 0.004 |
| Lorazepam (180) | 0.000 | 0.000 | 0.000 | 0.001 | 0.002 | 0.002 | 0.002 | 0.002 | 0.002 | 0.002 | 0.003 |
| Bromazepam (90) | 0.000 | 0.000 | 0.000 | 0.001 | 0.002 | 0.003 | 0.003 | 0.004 | 0.004 | 0.005 | 0.005 |
| Bromazepam (180) | 0.000 | 0.000 | 0.000 | 0.000 | 0.002 | 0.002 | 0.002 | 0.003 | 0.003 | 0.003 | 0.003 |
| Clobazam (90) | 0.000 | 0.000 | 0.000 | 0.000 | 0.001 | 0.001 | 0.001 | 0.001 | 0.000 | 0.000 | 0.000 |
| Clobazam (180) | 0.000 | 0.000 | 0.000 | 0.000 | 0.000 | 0.000 | 0.000 | 0.001 | 0.000 | 0.000 | 0.000 |
| Prazepam (90) | 0.000 | 0.000 | 0.000 | 0.000 | 0.000 | 0.000 | 0.000 | 0.001 | 0.001 | 0.001 | 0.001 |
| Prazepam (180) | 0.000 | 0.000 | 0.000 | 0.000 | 0.000 | 0.000 | 0.000 | 0.000 | 0.000 | 0.001 | 0.001 |
| Alprazolam (90) | 0.000 | 0.000 | 0.000 | 0.008 | 0.027 | 0.029 | 0.028 | 0.031 | 0.030 | 0.030 | 0.033 |
| Alprazolam (180) | 0.000 | 0.000 | 0.000 | 0.006 | 0.021 | 0.022 | 0.021 | 0.023 | 0.022 | 0.022 | 0.024 |
| Flurazepam (90) | 0.000 | 0.000 | 0.000 | 0.001 | 0.003 | 0.003 | 0.003 | 0.003 | 0.003 | 0.003 | 0.003 |
| Flurazepam (180) | 0.000 | 0.000 | 0.000 | 0.000 | 0.002 | 0.002 | 0.003 | 0.003 | 0.002 | 0.002 | 0.002 |
| Nitrazepam (90) | 0.000 | 0.000 | 0.000 | 0.000 | 0.000 | 0.000 | 0.000 | 0.000 | 0.001 | 0.000 | 0.001 |
| Nitrazepam (180) | 0.000 | 0.000 | 0.000 | 0.000 | 0.000 | 0.000 | 0.000 | 0.000 | 0.000 | 0.000 | 0.001 |
| Triazolam (90) | 0.000 | 0.000 | 0.000 | 0.000 | 0.001 | 0.001 | 0.001 | 0.002 | 0.002 | 0.002 | 0.003 |
| Triazolam (180) | 0.000 | 0.000 | 0.000 | 0.000 | 0.001 | 0.001 | 0.001 | 0.001 | 0.001 | 0.001 | 0.002 |
| Lormetazepam (90) | 0.000 | 0.000 | 0.000 | 0.000 | 0.001 | 0.001 | 0.001 | 0.001 | 0.002 | 0.002 | 0.003 |
| Lormetazepam (180) | 0.000 | 0.000 | 0.000 | 0.000 | 0.001 | 0.001 | 0.001 | 0.001 | 0.001 | 0.001 | 0.002 |
| Temazepam (90) | 0.000 | 0.000 | 0.000 | 0.000 | 0.001 | 0.002 | 0.002 | 0.003 | 0.003 | 0.003 | 0.005 |
| Temazepam (180) | 0.000 | 0.000 | 0.000 | 0.000 | 0.001 | 0.001 | 0.002 | 0.002 | 0.002 | 0.003 | 0.004 |
| Zopiclone (90) | 0.000 | 0.000 | 0.000 | 0.005 | 0.018 | 0.020 | 0.021 | 0.024 | 0.023 | 0.023 | 0.028 |
| Zopiclone (180) | 0.000 | 0.000 | 0.000 | 0.004 | 0.015 | 0.016 | 0.017 | 0.018 | 0.016 | 0.016 | 0.020 |
| Zolpidem (90) | 0.000 | 0.000 | 0.000 | 0.005 | 0.015 | 0.018 | 0.021 | 0.024 | 0.023 | 0.023 | 0.027 |
| Zolpidem (180) | 0.000 | 0.000 | 0.000 | 0.004 | 0.013 | 0.015 | 0.016 | 0.017 | 0.017 | 0.017 | 0.020 |
| Antidepressant (90) | 0.000 | 0.000 | 0.000 | 0.004 | 0.014 | 0.012 | 0.013 | 0.012 | 0.010 | 0.010 | 0.015 |
| Antidepressant (180) | 0.000 | 0.000 | 0.000 | 0.003 | 0.011 | 0.010 | 0.011 | 0.010 | 0.008 | 0.008 | 0.012 |
| Antihistamines (90) | 0.000 | 0.000 | 0.000 | 0.003 | 0.008 | 0.007 | 0.006 | 0.005 | 0.004 | 0.004 | 0.004 |
| Antihistamines (180) | 0.000 | 0.000 | 0.000 | 0.003 | 0.007 | 0.006 | 0.005 | 0.004 | 0.004 | 0.003 | 0.003 |
| Antipsychotics (90) | 0.000 | 0.000 | 0.000 | 0.005 | 0.013 | 0.010 | 0.009 | 0.008 | 0.006 | 0.006 | 0.012 |
| Antipsychotics (180) | 0.000 | 0.000 | 0.000 | 0.004 | 0.010 | 0.008 | 0.007 | 0.006 | 0.005 | 0.005 | 0.009 |
| 2019 |  |  |  |  |  |  |  |  |  |  |  |
| Sedative (90) | 0.000 | 0.001 | 0.001 | 0.018 | 0.060 | 0.067 | 0.074 | 0.078 | 0.077 | 0.073 | 0.075 |
| Sedative (180) | 0.000 | 0.000 | 0.001 | 0.011 | 0.037 | 0.041 | 0.045 | 0.047 | 0.046 | 0.043 | 0.043 |
| Benzodiazepines and z-drugs (90) | 0.000 | 0.001 | 0.001 | 0.020 | 0.065 | 0.072 | 0.077 | 0.080 | 0.079 | 0.075 | 0.082 |
| Benzodiazepines and z-drugs after (180) | 0.000 | 0.000 | 0.001 | 0.014 | 0.044 | 0.047 | 0.051 | 0.052 | 0.049 | 0.046 | 0.050 |
| Long-acting benzodiazepines (90) | 0.000 | 0.000 | 0.001 | 0.011 | 0.042 | 0.049 | 0.050 | 0.048 | 0.044 | 0.037 | 0.034 |
| Long-acting benzodiazepines (180) | 0.000 | 0.000 | 0.001 | 0.008 | 0.031 | 0.037 | 0.038 | 0.036 | 0.032 | 0.027 | 0.025 |
| Clonazepam (90) | 0.000 | 0.000 | 0.000 | 0.001 | 0.004 | 0.003 | 0.003 | 0.003 | 0.003 | 0.002 | 0.002 |
| Clonazepam (180) | 0.000 | 0.000 | 0.000 | 0.001 | 0.003 | 0.002 | 0.002 | 0.002 | 0.002 | 0.001 | 0.002 |
| Diazepam (90) | 0.000 | 0.000 | 0.001 | 0.009 | 0.036 | 0.042 | 0.042 | 0.040 | 0.037 | 0.031 | 0.028 |
| Diazepam (180) | 0.000 | 0.000 | 0.001 | 0.007 | 0.028 | 0.032 | 0.032 | 0.031 | 0.028 | 0.023 | 0.021 |
| Chlordiazepoxide (90) | 0.000 | 0.000 | 0.000 | 0.001 | 0.004 | 0.007 | 0.008 | 0.007 | 0.004 | 0.002 | 0.001 |
| Chlordiazepoxide (180) | 0.000 | 0.000 | 0.000 | 0.001 | 0.003 | 0.005 | 0.006 | 0.005 | 0.003 | 0.002 | 0.001 |
| Clorazepate (90) | 0.000 | 0.000 | 0.000 | 0.000 | 0.000 | 0.000 | 0.000 | 0.000 | 0.000 | 0.000 | 0.000 |
| Clorazepate (180) | 0.000 | 0.000 | 0.000 | 0.000 | 0.000 | 0.000 | 0.000 | 0.000 | 0.000 | 0.000 | 0.000 |
| Lorazepam (90) | 0.000 | 0.000 | 0.000 | 0.001 | 0.003 | 0.003 | 0.003 | 0.003 | 0.003 | 0.002 | 0.003 |
| Lorazepam (180) | 0.000 | 0.000 | 0.000 | 0.001 | 0.002 | 0.002 | 0.002 | 0.002 | 0.002 | 0.002 | 0.003 |
| Bromazepam (90) | 0.000 | 0.000 | 0.000 | 0.001 | 0.002 | 0.002 | 0.003 | 0.003 | 0.004 | 0.004 | 0.004 |
| Bromazepam (180) | 0.000 | 0.000 | 0.000 | 0.001 | 0.002 | 0.002 | 0.002 | 0.003 | 0.003 | 0.003 | 0.003 |
| Clobazam (90) | 0.000 | 0.000 | 0.000 | 0.000 | 0.001 | 0.001 | 0.001 | 0.001 | 0.001 | 0.000 | 0.000 |
| Clobazam (180) | 0.000 | 0.000 | 0.000 | 0.000 | 0.000 | 0.000 | 0.001 | 0.000 | 0.000 | 0.000 | 0.000 |
| Prazepam (90) | 0.000 | 0.000 | 0.000 | 0.000 | 0.000 | 0.000 | 0.000 | 0.000 | 0.001 | 0.001 | 0.001 |
| Prazepam (180) | 0.000 | 0.000 | 0.000 | 0.000 | 0.000 | 0.000 | 0.000 | 0.000 | 0.001 | 0.000 | 0.001 |
| Alprazolam (90) | 0.000 | 0.000 | 0.000 | 0.008 | 0.029 | 0.029 | 0.029 | 0.031 | 0.031 | 0.030 | 0.033 |
| Alprazolam (180) | 0.000 | 0.000 | 0.000 | 0.006 | 0.023 | 0.022 | 0.021 | 0.023 | 0.022 | 0.022 | 0.024 |
| Flurazepam (90) | 0.000 | 0.000 | 0.000 | 0.001 | 0.002 | 0.003 | 0.003 | 0.003 | 0.003 | 0.002 | 0.003 |
| Flurazepam (180) | 0.000 | 0.000 | 0.000 | 0.000 | 0.002 | 0.002 | 0.002 | 0.002 | 0.002 | 0.002 | 0.002 |
| Nitrazepam (90) | 0.000 | 0.000 | 0.000 | 0.000 | 0.000 | 0.000 | 0.000 | 0.000 | 0.001 | 0.000 | 0.001 |
| Nitrazepam (180) | 0.000 | 0.000 | 0.000 | 0.000 | 0.000 | 0.000 | 0.000 | 0.000 | 0.000 | 0.000 | 0.000 |
| Triazolam (90) | 0.000 | 0.000 | 0.000 | 0.000 | 0.001 | 0.001 | 0.001 | 0.002 | 0.002 | 0.002 | 0.002 |
| Triazolam (180) | 0.000 | 0.000 | 0.000 | 0.000 | 0.001 | 0.001 | 0.001 | 0.001 | 0.001 | 0.001 | 0.002 |
| Lormetazepam (90) | 0.000 | 0.000 | 0.000 | 0.000 | 0.001 | 0.001 | 0.001 | 0.002 | 0.002 | 0.004 | 0.006 |
| Lormetazepam (180) | 0.000 | 0.000 | 0.000 | 0.000 | 0.001 | 0.001 | 0.001 | 0.002 | 0.002 | 0.003 | 0.004 |
| Temazepam (90) | 0.000 | 0.000 | 0.000 | 0.000 | 0.001 | 0.002 | 0.002 | 0.003 | 0.003 | 0.003 | 0.005 |
| Temazepam (180) | 0.000 | 0.000 | 0.000 | 0.000 | 0.001 | 0.001 | 0.002 | 0.002 | 0.002 | 0.002 | 0.003 |
| Zopiclone (90) | 0.000 | 0.000 | 0.000 | 0.005 | 0.018 | 0.020 | 0.021 | 0.023 | 0.023 | 0.023 | 0.027 |
| Zopiclone (180) | 0.000 | 0.000 | 0.000 | 0.004 | 0.015 | 0.015 | 0.016 | 0.018 | 0.017 | 0.016 | 0.019 |
| Zolpidem (90) | 0.000 | 0.000 | 0.000 | 0.004 | 0.015 | 0.018 | 0.021 | 0.023 | 0.024 | 0.024 | 0.029 |
| Zolpidem (180) | 0.000 | 0.000 | 0.000 | 0.003 | 0.013 | 0.014 | 0.016 | 0.017 | 0.017 | 0.016 | 0.020 |
| Antidepressant (90) | 0.000 | 0.000 | 0.000 | 0.004 | 0.015 | 0.013 | 0.013 | 0.013 | 0.011 | 0.011 | 0.016 |
| Antidepressant (180) | 0.000 | 0.000 | 0.000 | 0.004 | 0.013 | 0.011 | 0.010 | 0.011 | 0.009 | 0.009 | 0.012 |
| Antihistamines (90) | 0.000 | 0.000 | 0.000 | 0.004 | 0.010 | 0.008 | 0.007 | 0.006 | 0.005 | 0.004 | 0.004 |
| Antihistamines (180) | 0.000 | 0.000 | 0.000 | 0.003 | 0.008 | 0.006 | 0.006 | 0.005 | 0.004 | 0.003 | 0.003 |
| Antipsychotics (90) | 0.000 | 0.000 | 0.000 | 0.006 | 0.014 | 0.011 | 0.010 | 0.008 | 0.007 | 0.006 | 0.012 |
| Antipsychotics (180) | 0.000 | 0.000 | 0.000 | 0.004 | 0.011 | 0.009 | 0.007 | 0.006 | 0.005 | 0.005 | 0.008 |
| 2020 |  |  |  |  |  |  |  |  |  |  |  |
| Sedative (90) | 0.000 | 0.000 | 0.001 | 0.014 | 0.051 | 0.059 | 0.065 | 0.067 | 0.065 | 0.062 | 0.065 |
| Sedative (180) | 0.000 | 0.000 | 0.001 | 0.008 | 0.033 | 0.037 | 0.041 | 0.042 | 0.040 | 0.038 | 0.039 |
| Benzodiazepines and z-drugs (90) | 0.000 | 0.000 | 0.001 | 0.016 | 0.058 | 0.064 | 0.069 | 0.070 | 0.067 | 0.065 | 0.072 |
| Benzodiazepines and z-drugs after (180) | 0.000 | 0.000 | 0.001 | 0.012 | 0.042 | 0.044 | 0.047 | 0.047 | 0.044 | 0.042 | 0.047 |
| Long-acting benzodiazepines (90) | 0.000 | 0.000 | 0.001 | 0.008 | 0.037 | 0.044 | 0.045 | 0.043 | 0.038 | 0.033 | 0.031 |
| Long-acting benzodiazepines (180) | 0.000 | 0.000 | 0.001 | 0.006 | 0.028 | 0.033 | 0.035 | 0.033 | 0.030 | 0.026 | 0.024 |
| Clonazepam (90) | 0.000 | 0.000 | 0.000 | 0.001 | 0.003 | 0.003 | 0.003 | 0.003 | 0.002 | 0.002 | 0.002 |
| Clonazepam (180) | 0.000 | 0.000 | 0.000 | 0.001 | 0.003 | 0.002 | 0.002 | 0.002 | 0.002 | 0.002 | 0.002 |
| Diazepam (90) | 0.000 | 0.000 | 0.001 | 0.007 | 0.032 | 0.037 | 0.038 | 0.035 | 0.032 | 0.028 | 0.026 |
| Diazepam (180) | 0.000 | 0.000 | 0.000 | 0.005 | 0.025 | 0.029 | 0.030 | 0.028 | 0.026 | 0.022 | 0.020 |
| Chlordiazepoxide (90) | 0.000 | 0.000 | 0.000 | 0.001 | 0.004 | 0.006 | 0.007 | 0.006 | 0.004 | 0.002 | 0.001 |
| Chlordiazepoxide (180) | 0.000 | 0.000 | 0.000 | 0.000 | 0.003 | 0.005 | 0.006 | 0.005 | 0.003 | 0.002 | 0.001 |
| Clorazepate (90) | 0.000 | 0.000 | 0.000 | 0.000 | 0.000 | 0.000 | 0.000 | 0.000 | 0.000 | 0.000 | 0.000 |
| Clorazepate (180) | 0.000 | 0.000 | 0.000 | 0.000 | 0.000 | 0.000 | 0.000 | 0.000 | 0.000 | 0.000 | 0.000 |
| Lorazepam (90) | 0.000 | 0.000 | 0.000 | 0.001 | 0.003 | 0.003 | 0.002 | 0.003 | 0.002 | 0.002 | 0.003 |
| Lorazepam (180) | 0.000 | 0.000 | 0.000 | 0.001 | 0.002 | 0.002 | 0.002 | 0.002 | 0.002 | 0.002 | 0.002 |
| Bromazepam (90) | 0.000 | 0.000 | 0.000 | 0.000 | 0.002 | 0.002 | 0.002 | 0.003 | 0.003 | 0.004 | 0.004 |
| Bromazepam (180) | 0.000 | 0.000 | 0.000 | 0.000 | 0.002 | 0.002 | 0.002 | 0.002 | 0.002 | 0.003 | 0.003 |
| Clobazam (90) | 0.000 | 0.000 | 0.000 | 0.000 | 0.000 | 0.000 | 0.000 | 0.000 | 0.000 | 0.000 | 0.000 |
| Clobazam (180) | 0.000 | 0.000 | 0.000 | 0.000 | 0.000 | 0.000 | 0.000 | 0.000 | 0.000 | 0.000 | 0.000 |
| Prazepam (90) | 0.000 | 0.000 | 0.000 | 0.000 | 0.000 | 0.000 | 0.000 | 0.001 | 0.001 | 0.001 | 0.001 |
| Prazepam (180) | 0.000 | 0.000 | 0.000 | 0.000 | 0.000 | 0.000 | 0.000 | 0.000 | 0.001 | 0.000 | 0.001 |
| Alprazolam (90) | 0.000 | 0.000 | 0.000 | 0.006 | 0.025 | 0.025 | 0.025 | 0.026 | 0.025 | 0.026 | 0.030 |
| Alprazolam (180) | 0.000 | 0.000 | 0.000 | 0.005 | 0.021 | 0.020 | 0.020 | 0.021 | 0.020 | 0.020 | 0.023 |
| Flurazepam (90) | 0.000 | 0.000 | 0.000 | 0.000 | 0.002 | 0.002 | 0.003 | 0.003 | 0.002 | 0.002 | 0.002 |
| Flurazepam (180) | 0.000 | 0.000 | 0.000 | 0.000 | 0.002 | 0.002 | 0.002 | 0.002 | 0.002 | 0.002 | 0.002 |
| Nitrazepam (90) | 0.000 | 0.000 | 0.000 | 0.000 | 0.000 | 0.000 | 0.000 | 0.000 | 0.000 | 0.000 | 0.001 |
| Nitrazepam (180) | 0.000 | 0.000 | 0.000 | 0.000 | 0.000 | 0.000 | 0.000 | 0.000 | 0.000 | 0.000 | 0.001 |
| Triazolam (90) | 0.000 | 0.000 | 0.000 | 0.000 | 0.001 | 0.001 | 0.001 | 0.001 | 0.001 | 0.001 | 0.002 |
| Triazolam (180) | 0.000 | 0.000 | 0.000 | 0.000 | 0.001 | 0.001 | 0.001 | 0.001 | 0.001 | 0.001 | 0.002 |
| Lormetazepam (90) | 0.000 | 0.000 | 0.000 | 0.000 | 0.000 | 0.000 | 0.000 | 0.001 | 0.001 | 0.002 | 0.003 |
| Lormetazepam (180) | 0.000 | 0.000 | 0.000 | 0.000 | 0.000 | 0.000 | 0.000 | 0.001 | 0.002 | 0.002 | 0.004 |
| Temazepam (90) | 0.000 | 0.000 | 0.000 | 0.000 | 0.001 | 0.001 | 0.002 | 0.002 | 0.002 | 0.003 | 0.004 |
| Temazepam (180) | 0.000 | 0.000 | 0.000 | 0.000 | 0.001 | 0.001 | 0.001 | 0.002 | 0.002 | 0.002 | 0.003 |
| Zopiclone (90) | 0.000 | 0.000 | 0.000 | 0.004 | 0.016 | 0.017 | 0.019 | 0.020 | 0.021 | 0.020 | 0.024 |
| Zopiclone (180) | 0.000 | 0.000 | 0.000 | 0.004 | 0.013 | 0.014 | 0.015 | 0.015 | 0.015 | 0.015 | 0.018 |
| Zolpidem (90) | 0.000 | 0.000 | 0.000 | 0.004 | 0.013 | 0.016 | 0.018 | 0.020 | 0.021 | 0.021 | 0.025 |
| Zolpidem (180) | 0.000 | 0.000 | 0.000 | 0.003 | 0.011 | 0.013 | 0.014 | 0.015 | 0.015 | 0.015 | 0.019 |
| Antidepressant (90) | 0.000 | 0.000 | 0.000 | 0.004 | 0.013 | 0.012 | 0.012 | 0.011 | 0.011 | 0.010 | 0.015 |
| Antidepressant (180) | 0.000 | 0.000 | 0.000 | 0.003 | 0.011 | 0.010 | 0.010 | 0.009 | 0.008 | 0.008 | 0.011 |
| Antihistamines (90) | 0.000 | 0.000 | 0.000 | 0.004 | 0.011 | 0.010 | 0.009 | 0.008 | 0.006 | 0.005 | 0.005 |
| Antihistamines (180) | 0.000 | 0.000 | 0.000 | 0.003 | 0.009 | 0.008 | 0.007 | 0.006 | 0.005 | 0.004 | 0.004 |
| Antipsychotics (90) | 0.000 | 0.000 | 0.000 | 0.005 | 0.014 | 0.011 | 0.010 | 0.008 | 0.007 | 0.006 | 0.012 |
| Antipsychotics (180) | 0.000 | 0.000 | 0.000 | 0.004 | 0.011 | 0.009 | 0.008 | 0.007 | 0.006 | 0.005 | 0.008 |
| 2021 |  |  |  |  |  |  |  |  |  |  |  |
| Sedative (90) | 0.000 | 0.000 | 0.001 | 0.015 | 0.054 | 0.062 | 0.067 | 0.068 | 0.066 | 0.060 | 0.065 |
| Sedative (180) | 0.000 | 0.000 | 0.001 | 0.009 | 0.034 | 0.039 | 0.042 | 0.042 | 0.040 | 0.035 | 0.038 |
| Benzodiazepines and z-drugs (90) | 0.000 | 0.000 | 0.001 | 0.018 | 0.059 | 0.067 | 0.070 | 0.070 | 0.068 | 0.063 | 0.072 |
| Benzodiazepines and z-drugs after (180) | 0.000 | 0.000 | 0.001 | 0.013 | 0.042 | 0.046 | 0.048 | 0.047 | 0.044 | 0.039 | 0.046 |
| Long-acting benzodiazepines (90) | 0.000 | 0.000 | 0.001 | 0.010 | 0.038 | 0.045 | 0.047 | 0.044 | 0.038 | 0.033 | 0.032 |
| Long-acting benzodiazepines (180) | 0.000 | 0.000 | 0.001 | 0.007 | 0.029 | 0.034 | 0.035 | 0.033 | 0.028 | 0.024 | 0.024 |
| Clonazepam (90) | 0.000 | 0.000 | 0.000 | 0.001 | 0.003 | 0.003 | 0.003 | 0.003 | 0.002 | 0.002 | 0.002 |
| Clonazepam (180) | 0.000 | 0.000 | 0.000 | 0.001 | 0.003 | 0.002 | 0.002 | 0.002 | 0.002 | 0.002 | 0.002 |
| Diazepam (90) | 0.000 | 0.000 | 0.001 | 0.008 | 0.033 | 0.039 | 0.039 | 0.036 | 0.032 | 0.027 | 0.026 |
| Diazepam (180) | 0.000 | 0.000 | 0.001 | 0.006 | 0.026 | 0.030 | 0.030 | 0.028 | 0.024 | 0.020 | 0.019 |
| Chlordiazepoxide (90) | 0.000 | 0.000 | 0.000 | 0.001 | 0.004 | 0.006 | 0.007 | 0.006 | 0.004 | 0.002 | 0.001 |
| Chlordiazepoxide (180) | 0.000 | 0.000 | 0.000 | 0.000 | 0.003 | 0.005 | 0.006 | 0.005 | 0.003 | 0.002 | 0.001 |
| Clorazepate (90) | 0.000 | 0.000 | 0.000 | 0.000 | 0.000 | 0.000 | 0.000 | 0.000 | 0.000 | 0.000 | 0.000 |
| Clorazepate (180) | 0.000 | 0.000 | 0.000 | 0.000 | 0.000 | 0.000 | 0.000 | 0.000 | 0.000 | 0.000 | 0.000 |
| Lorazepam (90) | 0.000 | 0.000 | 0.000 | 0.001 | 0.003 | 0.003 | 0.003 | 0.003 | 0.002 | 0.002 | 0.004 |
| Lorazepam (180) | 0.000 | 0.000 | 0.000 | 0.001 | 0.002 | 0.002 | 0.002 | 0.002 | 0.002 | 0.002 | 0.003 |
| Bromazepam (90) | 0.000 | 0.000 | 0.000 | 0.000 | 0.002 | 0.002 | 0.002 | 0.002 | 0.003 | 0.003 | 0.004 |
| Bromazepam (180) | 0.000 | 0.000 | 0.000 | 0.000 | 0.001 | 0.002 | 0.001 | 0.002 | 0.002 | 0.002 | 0.003 |
| Clobazam (90) | 0.000 | 0.000 | 0.000 | 0.000 | 0.001 | 0.000 | 0.000 | 0.000 | 0.000 | 0.000 | 0.000 |
| Clobazam (180) | 0.000 | 0.000 | 0.000 | 0.000 | 0.000 | 0.000 | 0.000 | 0.000 | 0.000 | 0.000 | 0.000 |
| Prazepam (90) | 0.000 | 0.000 | 0.000 | 0.000 | 0.000 | 0.000 | 0.000 | 0.000 | 0.000 | 0.000 | 0.000 |
| Prazepam (180) | 0.000 | 0.000 | 0.000 | 0.000 | 0.000 | 0.000 | 0.000 | 0.000 | 0.000 | 0.000 | 0.000 |
| Alprazolam (90) | 0.000 | 0.000 | 0.000 | 0.007 | 0.024 | 0.025 | 0.025 | 0.024 | 0.024 | 0.024 | 0.028 |
| Alprazolam (180) | 0.000 | 0.000 | 0.000 | 0.005 | 0.019 | 0.020 | 0.018 | 0.018 | 0.018 | 0.018 | 0.021 |
| Flurazepam (90) | 0.000 | 0.000 | 0.000 | 0.000 | 0.002 | 0.002 | 0.002 | 0.003 | 0.002 | 0.002 | 0.002 |
| Flurazepam (180) | 0.000 | 0.000 | 0.000 | 0.000 | 0.002 | 0.002 | 0.002 | 0.002 | 0.002 | 0.002 | 0.002 |
| Nitrazepam (90) | 0.000 | 0.000 | 0.000 | 0.000 | 0.000 | 0.000 | 0.000 | 0.001 | 0.001 | 0.001 | 0.002 |
| Nitrazepam (180) | 0.000 | 0.000 | 0.000 | 0.000 | 0.000 | 0.000 | 0.000 | 0.001 | 0.001 | 0.001 | 0.002 |
| Triazolam (90) | 0.000 | 0.000 | 0.000 | 0.000 | 0.001 | 0.001 | 0.001 | 0.001 | 0.001 | 0.002 | 0.002 |
| Triazolam (180) | 0.000 | 0.000 | 0.000 | 0.000 | 0.001 | 0.001 | 0.001 | 0.001 | 0.001 | 0.001 | 0.001 |
| Lormetazepam (90) | 0.000 | 0.000 | 0.000 | 0.000 | 0.000 | 0.000 | 0.000 | 0.000 | 0.000 | 0.000 | 0.000 |
| Lormetazepam (180) | 0.000 | 0.000 | 0.000 | 0.000 | 0.000 | 0.000 | 0.000 | 0.000 | 0.000 | 0.000 | 0.000 |
| Temazepam (90) | 0.000 | 0.000 | 0.000 | 0.000 | 0.001 | 0.001 | 0.002 | 0.002 | 0.002 | 0.002 | 0.004 |
| Temazepam (180) | 0.000 | 0.000 | 0.000 | 0.000 | 0.001 | 0.001 | 0.001 | 0.002 | 0.002 | 0.002 | 0.003 |
| Zopiclone (90) | 0.000 | 0.000 | 0.000 | 0.004 | 0.016 | 0.017 | 0.018 | 0.019 | 0.021 | 0.019 | 0.024 |
| Zopiclone (180) | 0.000 | 0.000 | 0.000 | 0.004 | 0.013 | 0.014 | 0.014 | 0.015 | 0.015 | 0.013 | 0.017 |
| Zolpidem (90) | 0.000 | 0.000 | 0.000 | 0.004 | 0.013 | 0.016 | 0.018 | 0.020 | 0.019 | 0.020 | 0.023 |
| Zolpidem (180) | 0.000 | 0.000 | 0.000 | 0.003 | 0.011 | 0.013 | 0.014 | 0.015 | 0.014 | 0.014 | 0.017 |
| Antidepressant (90) | 0.000 | 0.000 | 0.000 | 0.004 | 0.015 | 0.014 | 0.012 | 0.012 | 0.012 | 0.012 | 0.016 |
| Antidepressant (180) | 0.000 | 0.000 | 0.000 | 0.003 | 0.012 | 0.012 | 0.010 | 0.010 | 0.009 | 0.009 | 0.013 |
| Antihistamines (90) | 0.000 | 0.000 | 0.000 | 0.005 | 0.014 | 0.012 | 0.011 | 0.010 | 0.008 | 0.006 | 0.006 |
| Antihistamines (180) | 0.000 | 0.000 | 0.000 | 0.004 | 0.012 | 0.009 | 0.009 | 0.008 | 0.006 | 0.005 | 0.005 |
| Antipsychotics (90) | 0.000 | 0.000 | 0.000 | 0.006 | 0.016 | 0.012 | 0.010 | 0.009 | 0.008 | 0.006 | 0.013 |
| Antipsychotics (180) | 0.000 | 0.000 | 0.000 | 0.004 | 0.013 | 0.010 | 0.008 | 0.007 | 0.006 | 0.005 | 0.009 |
| 2022 |  |  |  |  |  |  |  |  |  |  |  |
| Sedative (90) | 0.000 | 0.000 | 0.001 | 0.013 | 0.050 | 0.058 | 0.066 | 0.068 | 0.068 | 0.065 | 0.066 |
| Sedative (180) | 0.000 | 0.000 | 0.000 | 0.008 | 0.031 | 0.036 | 0.041 | 0.042 | 0.041 | 0.039 | 0.039 |
| Benzodiazepines and z-drugs (90) | 0.000 | 0.000 | 0.001 | 0.016 | 0.054 | 0.063 | 0.069 | 0.071 | 0.069 | 0.068 | 0.073 |
| Benzodiazepines and z-drugs after (180) | 0.000 | 0.000 | 0.001 | 0.011 | 0.039 | 0.043 | 0.047 | 0.046 | 0.044 | 0.043 | 0.048 |
| Long-acting benzodiazepines (90) | 0.000 | 0.000 | 0.001 | 0.009 | 0.034 | 0.043 | 0.045 | 0.043 | 0.040 | 0.035 | 0.031 |
| Long-acting benzodiazepines (180) | 0.000 | 0.000 | 0.001 | 0.006 | 0.026 | 0.032 | 0.035 | 0.032 | 0.029 | 0.026 | 0.024 |
| Clonazepam (90) | 0.000 | 0.000 | 0.000 | 0.001 | 0.004 | 0.004 | 0.003 | 0.004 | 0.003 | 0.003 | 0.003 |
| Clonazepam (180) | 0.000 | 0.000 | 0.000 | 0.001 | 0.003 | 0.002 | 0.002 | 0.002 | 0.002 | 0.002 | 0.002 |
| Diazepam (90) | 0.000 | 0.000 | 0.001 | 0.007 | 0.029 | 0.036 | 0.038 | 0.036 | 0.033 | 0.029 | 0.026 |
| Diazepam (180) | 0.000 | 0.000 | 0.000 | 0.005 | 0.023 | 0.028 | 0.029 | 0.027 | 0.025 | 0.022 | 0.020 |
| Chlordiazepoxide (90) | 0.000 | 0.000 | 0.000 | 0.001 | 0.003 | 0.006 | 0.007 | 0.006 | 0.004 | 0.002 | 0.001 |
| Chlordiazepoxide (180) | 0.000 | 0.000 | 0.000 | 0.000 | 0.003 | 0.004 | 0.005 | 0.005 | 0.003 | 0.002 | 0.001 |
| Clorazepate (90) | 0.000 | 0.000 | 0.000 | 0.000 | 0.000 | 0.000 | 0.000 | 0.000 | 0.000 | 0.000 | 0.000 |
| Clorazepate (180) | 0.000 | 0.000 | 0.000 | 0.000 | 0.000 | 0.000 | 0.000 | 0.000 | 0.000 | 0.000 | 0.000 |
| Lorazepam (90) | 0.000 | 0.000 | 0.000 | 0.001 | 0.003 | 0.003 | 0.003 | 0.003 | 0.002 | 0.002 | 0.003 |
| Lorazepam (180) | 0.000 | 0.000 | 0.000 | 0.001 | 0.002 | 0.002 | 0.002 | 0.002 | 0.002 | 0.002 | 0.002 |
| Bromazepam (90) | 0.000 | 0.000 | 0.000 | 0.000 | 0.002 | 0.001 | 0.002 | 0.002 | 0.003 | 0.003 | 0.004 |
| Bromazepam (180) | 0.000 | 0.000 | 0.000 | 0.000 | 0.001 | 0.001 | 0.001 | 0.002 | 0.002 | 0.002 | 0.002 |
| Clobazam (90) | 0.000 | 0.000 | 0.000 | 0.000 | 0.001 | 0.000 | 0.000 | 0.000 | 0.000 | 0.000 | 0.000 |
| Clobazam (180) | 0.000 | 0.000 | 0.000 | 0.000 | 0.000 | 0.000 | 0.000 | 0.000 | 0.000 | 0.000 | 0.000 |
| Prazepam (90) | 0.000 | 0.000 | 0.000 | 0.000 | 0.000 | 0.000 | 0.001 | 0.001 | 0.001 | 0.001 | 0.001 |
| Prazepam (180) | 0.000 | 0.000 | 0.000 | 0.000 | 0.000 | 0.000 | 0.000 | 0.000 | 0.000 | 0.000 | 0.000 |
| Alprazolam (90) | 0.000 | 0.000 | 0.000 | 0.006 | 0.024 | 0.025 | 0.025 | 0.027 | 0.027 | 0.027 | 0.029 |
| Alprazolam (180) | 0.000 | 0.000 | 0.000 | 0.004 | 0.018 | 0.019 | 0.019 | 0.019 | 0.019 | 0.019 | 0.021 |
| Flurazepam (90) | 0.000 | 0.000 | 0.000 | 0.000 | 0.001 | 0.002 | 0.002 | 0.002 | 0.002 | 0.002 | 0.002 |
| Flurazepam (180) | 0.000 | 0.000 | 0.000 | 0.000 | 0.001 | 0.001 | 0.002 | 0.002 | 0.001 | 0.001 | 0.001 |
| Nitrazepam (90) | 0.000 | 0.000 | 0.000 | 0.000 | 0.000 | 0.000 | 0.000 | 0.000 | 0.000 | 0.000 | 0.000 |
| Nitrazepam (180) | 0.000 | 0.000 | 0.000 | 0.000 | 0.000 | 0.000 | 0.000 | 0.000 | 0.000 | 0.000 | 0.000 |
| Triazolam (90) | 0.000 | 0.000 | 0.000 | 0.000 | 0.001 | 0.001 | 0.001 | 0.001 | 0.001 | 0.001 | 0.002 |
| Triazolam (180) | 0.000 | 0.000 | 0.000 | 0.000 | 0.001 | 0.001 | 0.001 | 0.001 | 0.001 | 0.001 | 0.001 |
| Lormetazepam (90) | 0.000 | 0.000 | 0.000 | 0.000 | 0.000 | 0.000 | 0.000 | 0.000 | 0.000 | 0.000 | 0.000 |
| Lormetazepam (180) | 0.000 | 0.000 | 0.000 | 0.000 | 0.000 | 0.000 | 0.000 | 0.000 | 0.000 | 0.000 | 0.000 |
| Temazepam (90) | 0.000 | 0.000 | 0.000 | 0.000 | 0.001 | 0.001 | 0.001 | 0.002 | 0.002 | 0.002 | 0.003 |
| Temazepam (180) | 0.000 | 0.000 | 0.000 | 0.000 | 0.001 | 0.001 | 0.001 | 0.001 | 0.002 | 0.002 | 0.002 |
| Zopiclone (90) | 0.000 | 0.000 | 0.000 | 0.003 | 0.012 | 0.015 | 0.017 | 0.018 | 0.019 | 0.018 | 0.023 |
| Zopiclone (180) | 0.000 | 0.000 | 0.000 | 0.003 | 0.010 | 0.012 | 0.013 | 0.013 | 0.014 | 0.013 | 0.017 |
| Zolpidem (90) | 0.000 | 0.000 | 0.000 | 0.003 | 0.010 | 0.013 | 0.016 | 0.018 | 0.018 | 0.019 | 0.023 |
| Zolpidem (180) | 0.000 | 0.000 | 0.000 | 0.002 | 0.009 | 0.011 | 0.012 | 0.013 | 0.013 | 0.013 | 0.017 |
| Antidepressant (90) | 0.000 | 0.000 | 0.000 | 0.003 | 0.013 | 0.012 | 0.013 | 0.013 | 0.012 | 0.012 | 0.017 |
| Antidepressant (180) | 0.000 | 0.000 | 0.000 | 0.003 | 0.011 | 0.010 | 0.010 | 0.010 | 0.009 | 0.009 | 0.013 |
| Antihistamines (90) | 0.000 | 0.000 | 0.000 | 0.005 | 0.013 | 0.013 | 0.012 | 0.010 | 0.008 | 0.007 | 0.007 |
| Antihistamines (180) | 0.000 | 0.000 | 0.000 | 0.004 | 0.011 | 0.010 | 0.009 | 0.008 | 0.006 | 0.006 | 0.005 |
| Antipsychotics (90) | 0.000 | 0.000 | 0.000 | 0.005 | 0.015 | 0.012 | 0.010 | 0.009 | 0.007 | 0.007 | 0.012 |
| Antipsychotics (180) | 0.000 | 0.000 | 0.000 | 0.004 | 0.012 | 0.010 | 0.008 | 0.007 | 0.006 | 0.005 | 0.008 |

**Prevalence of discontinuations after 90 and 180 days in the GMS population by sex**

|  | F | M |
| --- | --- | --- |
| 2014 |  |  |
| Sedative (90) | 0.052 | 0.032 |
| Sedative (180) | 0.024 | 0.015 |
| Benzodiazepines and z-drugs (90) | 0.053 | 0.033 |
| Benzodiazepines and z-drugs after (180) | 0.025 | 0.016 |
| Long-acting benzodiazepines (90) | 0.028 | 0.018 |
| Long-acting benzodiazepines (180) | 0.015 | 0.010 |
| Clonazepam (90) | 0.001 | 0.001 |
| Clonazepam (180) | 0.001 | 0.001 |
| Diazepam (90) | 0.024 | 0.014 |
| Diazepam (180) | 0.013 | 0.007 |
| Chlordiazepoxide (90) | 0.002 | 0.003 |
| Chlordiazepoxide (180) | 0.001 | 0.002 |
| Clorazepate (90) | 0.000 | 0.000 |
| Clorazepate (180) | 0.000 | 0.000 |
| Lorazepam (90) | 0.002 | 0.001 |
| Lorazepam (180) | 0.001 | 0.001 |
| Bromazepam (90) | 0.003 | 0.001 |
| Bromazepam (180) | 0.001 | 0.001 |
| Clobazam (90) | 0.000 | 0.000 |
| Clobazam (180) | 0.000 | 0.000 |
| Prazepam (90) | 0.001 | 0.000 |
| Prazepam (180) | 0.000 | 0.000 |
| Alprazolam (90) | 0.021 | 0.009 |
| Alprazolam (180) | 0.010 | 0.005 |
| Flurazepam (90) | 0.002 | 0.002 |
| Flurazepam (180) | 0.001 | 0.001 |
| Nitrazepam (90) | 0.000 | 0.000 |
| Nitrazepam (180) | 0.000 | 0.000 |
| Triazolam (90) | 0.001 | 0.001 |
| Triazolam (180) | 0.001 | 0.000 |
| Lormetazepam (90) | 0.002 | 0.001 |
| Lormetazepam (180) | 0.001 | 0.000 |
| Temazepam (90) | 0.003 | 0.002 |
| Temazepam (180) | 0.002 | 0.001 |
| Zopiclone (90) | 0.015 | 0.010 |
| Zopiclone (180) | 0.008 | 0.005 |
| Zolpidem (90) | 0.016 | 0.010 |
| Zolpidem (180) | 0.009 | 0.005 |
| Antidepressant (90) | 0.006 | 0.004 |
| Antidepressant (180) | 0.003 | 0.002 |
| Antihistamines (90) | 0.001 | 0.001 |
| Antihistamines (180) | 0.001 | 0.000 |
| Antipsychotics (90) | 0.005 | 0.004 |
| Antipsychotics (180) | 0.002 | 0.002 |
| 2015 |  |  |
| Sedative (90) | 0.061 | 0.037 |
| Sedative (180) | 0.036 | 0.022 |
| Benzodiazepines and z-drugs (90) | 0.063 | 0.039 |
| Benzodiazepines and z-drugs after (180) | 0.039 | 0.024 |
| Long-acting benzodiazepines (90) | 0.036 | 0.023 |
| Long-acting benzodiazepines (180) | 0.026 | 0.017 |
| Clonazepam (90) | 0.002 | 0.002 |
| Clonazepam (180) | 0.002 | 0.001 |
| Diazepam (90) | 0.031 | 0.018 |
| Diazepam (180) | 0.023 | 0.013 |
| Chlordiazepoxide (90) | 0.002 | 0.004 |
| Chlordiazepoxide (180) | 0.002 | 0.003 |
| Clorazepate (90) | 0.000 | 0.000 |
| Clorazepate (180) | 0.000 | 0.000 |
| Lorazepam (90) | 0.002 | 0.002 |
| Lorazepam (180) | 0.002 | 0.001 |
| Bromazepam (90) | 0.004 | 0.002 |
| Bromazepam (180) | 0.003 | 0.001 |
| Clobazam (90) | 0.001 | 0.000 |
| Clobazam (180) | 0.000 | 0.000 |
| Prazepam (90) | 0.001 | 0.000 |
| Prazepam (180) | 0.001 | 0.000 |
| Alprazolam (90) | 0.027 | 0.012 |
| Alprazolam (180) | 0.020 | 0.009 |
| Flurazepam (90) | 0.003 | 0.002 |
| Flurazepam (180) | 0.002 | 0.002 |
| Nitrazepam (90) | 0.001 | 0.000 |
| Nitrazepam (180) | 0.000 | 0.000 |
| Triazolam (90) | 0.002 | 0.001 |
| Triazolam (180) | 0.001 | 0.001 |
| Lormetazepam (90) | 0.002 | 0.001 |
| Lormetazepam (180) | 0.001 | 0.001 |
| Temazepam (90) | 0.003 | 0.002 |
| Temazepam (180) | 0.003 | 0.002 |
| Zopiclone (90) | 0.019 | 0.013 |
| Zopiclone (180) | 0.014 | 0.009 |
| Zolpidem (90) | 0.020 | 0.012 |
| Zolpidem (180) | 0.015 | 0.009 |
| Antidepressant (90) | 0.009 | 0.006 |
| Antidepressant (180) | 0.007 | 0.005 |
| Antihistamines (90) | 0.002 | 0.001 |
| Antihistamines (180) | 0.002 | 0.001 |
| Antipsychotics (90) | 0.006 | 0.005 |
| Antipsychotics (180) | 0.005 | 0.004 |
| 2016 |  |  |
| Sedative (90) | 0.064 | 0.040 |
| Sedative (180) | 0.039 | 0.024 |
| Benzodiazepines and z-drugs (90) | 0.067 | 0.041 |
| Benzodiazepines and z-drugs after (180) | 0.042 | 0.026 |
| Long-acting benzodiazepines (90) | 0.038 | 0.025 |
| Long-acting benzodiazepines (180) | 0.028 | 0.018 |
| Clonazepam (90) | 0.002 | 0.002 |
| Clonazepam (180) | 0.002 | 0.001 |
| Diazepam (90) | 0.033 | 0.019 |
| Diazepam (180) | 0.025 | 0.014 |
| Chlordiazepoxide (90) | 0.002 | 0.004 |
| Chlordiazepoxide (180) | 0.002 | 0.003 |
| Clorazepate (90) | 0.000 | 0.000 |
| Clorazepate (180) | 0.000 | 0.000 |
| Lorazepam (90) | 0.002 | 0.002 |
| Lorazepam (180) | 0.002 | 0.001 |
| Bromazepam (90) | 0.004 | 0.002 |
| Bromazepam (180) | 0.003 | 0.001 |
| Clobazam (90) | 0.000 | 0.000 |
| Clobazam (180) | 0.000 | 0.000 |
| Prazepam (90) | 0.001 | 0.000 |
| Prazepam (180) | 0.000 | 0.000 |
| Alprazolam (90) | 0.029 | 0.013 |
| Alprazolam (180) | 0.022 | 0.010 |
| Flurazepam (90) | 0.003 | 0.002 |
| Flurazepam (180) | 0.002 | 0.002 |
| Nitrazepam (90) | 0.000 | 0.000 |
| Nitrazepam (180) | 0.000 | 0.000 |
| Triazolam (90) | 0.002 | 0.001 |
| Triazolam (180) | 0.001 | 0.001 |
| Lormetazepam (90) | 0.002 | 0.001 |
| Lormetazepam (180) | 0.001 | 0.001 |
| Temazepam (90) | 0.003 | 0.002 |
| Temazepam (180) | 0.002 | 0.002 |
| Zopiclone (90) | 0.021 | 0.013 |
| Zopiclone (180) | 0.015 | 0.010 |
| Zolpidem (90) | 0.021 | 0.012 |
| Zolpidem (180) | 0.015 | 0.009 |
| Antidepressant (90) | 0.010 | 0.007 |
| Antidepressant (180) | 0.008 | 0.005 |
| Antihistamines (90) | 0.003 | 0.002 |
| Antihistamines (180) | 0.002 | 0.002 |
| Antipsychotics (90) | 0.007 | 0.006 |
| Antipsychotics (180) | 0.005 | 0.004 |
| 2017 |  |  |
| Sedative (90) | 0.064 | 0.040 |
| Sedative (180) | 0.039 | 0.024 |
| Benzodiazepines and z-drugs (90) | 0.067 | 0.042 |
| Benzodiazepines and z-drugs after (180) | 0.043 | 0.027 |
| Long-acting benzodiazepines (90) | 0.038 | 0.025 |
| Long-acting benzodiazepines (180) | 0.029 | 0.018 |
| Clonazepam (90) | 0.002 | 0.002 |
| Clonazepam (180) | 0.002 | 0.001 |
| Diazepam (90) | 0.033 | 0.019 |
| Diazepam (180) | 0.026 | 0.014 |
| Chlordiazepoxide (90) | 0.002 | 0.005 |
| Chlordiazepoxide (180) | 0.002 | 0.004 |
| Clorazepate (90) | 0.000 | 0.000 |
| Clorazepate (180) | 0.000 | 0.000 |
| Lorazepam (90) | 0.002 | 0.002 |
| Lorazepam (180) | 0.002 | 0.002 |
| Bromazepam (90) | 0.004 | 0.002 |
| Bromazepam (180) | 0.003 | 0.001 |
| Clobazam (90) | 0.000 | 0.000 |
| Clobazam (180) | 0.000 | 0.000 |
| Prazepam (90) | 0.001 | 0.000 |
| Prazepam (180) | 0.000 | 0.000 |
| Alprazolam (90) | 0.030 | 0.013 |
| Alprazolam (180) | 0.022 | 0.010 |
| Flurazepam (90) | 0.002 | 0.002 |
| Flurazepam (180) | 0.002 | 0.002 |
| Nitrazepam (90) | 0.000 | 0.000 |
| Nitrazepam (180) | 0.000 | 0.000 |
| Triazolam (90) | 0.002 | 0.001 |
| Triazolam (180) | 0.001 | 0.001 |
| Lormetazepam (90) | 0.002 | 0.001 |
| Lormetazepam (180) | 0.001 | 0.001 |
| Temazepam (90) | 0.003 | 0.002 |
| Temazepam (180) | 0.002 | 0.001 |
| Zopiclone (90) | 0.020 | 0.014 |
| Zopiclone (180) | 0.015 | 0.010 |
| Zolpidem (90) | 0.021 | 0.012 |
| Zolpidem (180) | 0.016 | 0.009 |
| Antidepressant (90) | 0.010 | 0.007 |
| Antidepressant (180) | 0.008 | 0.006 |
| Antihistamines (90) | 0.004 | 0.003 |
| Antihistamines (180) | 0.003 | 0.002 |
| Antipsychotics (90) | 0.007 | 0.006 |
| Antipsychotics (180) | 0.006 | 0.005 |
| 2018 |  |  |
| Sedative (90) | 0.064 | 0.039 |
| Sedative (180) | 0.038 | 0.024 |
| Benzodiazepines and z-drugs (90) | 0.067 | 0.041 |
| Benzodiazepines and z-drugs after (180) | 0.043 | 0.027 |
| Long-acting benzodiazepines (90) | 0.037 | 0.024 |
| Long-acting benzodiazepines (180) | 0.028 | 0.018 |
| Clonazepam (90) | 0.002 | 0.002 |
| Clonazepam (180) | 0.002 | 0.001 |
| Diazepam (90) | 0.033 | 0.019 |
| Diazepam (180) | 0.025 | 0.014 |
| Chlordiazepoxide (90) | 0.002 | 0.004 |
| Chlordiazepoxide (180) | 0.002 | 0.004 |
| Clorazepate (90) | 0.000 | 0.000 |
| Clorazepate (180) | 0.000 | 0.000 |
| Lorazepam (90) | 0.002 | 0.002 |
| Lorazepam (180) | 0.002 | 0.002 |
| Bromazepam (90) | 0.003 | 0.002 |
| Bromazepam (180) | 0.002 | 0.001 |
| Clobazam (90) | 0.000 | 0.000 |
| Clobazam (180) | 0.000 | 0.000 |
| Prazepam (90) | 0.001 | 0.000 |
| Prazepam (180) | 0.000 | 0.000 |
| Alprazolam (90) | 0.029 | 0.013 |
| Alprazolam (180) | 0.022 | 0.010 |
| Flurazepam (90) | 0.002 | 0.002 |
| Flurazepam (180) | 0.002 | 0.002 |
| Nitrazepam (90) | 0.000 | 0.000 |
| Nitrazepam (180) | 0.000 | 0.000 |
| Triazolam (90) | 0.001 | 0.001 |
| Triazolam (180) | 0.001 | 0.001 |
| Lormetazepam (90) | 0.001 | 0.001 |
| Lormetazepam (180) | 0.001 | 0.001 |
| Temazepam (90) | 0.002 | 0.002 |
| Temazepam (180) | 0.002 | 0.001 |
| Zopiclone (90) | 0.019 | 0.013 |
| Zopiclone (180) | 0.015 | 0.010 |
| Zolpidem (90) | 0.020 | 0.011 |
| Zolpidem (180) | 0.015 | 0.009 |
| Antidepressant (90) | 0.011 | 0.007 |
| Antidepressant (180) | 0.009 | 0.006 |
| Antihistamines (90) | 0.005 | 0.003 |
| Antihistamines (180) | 0.004 | 0.003 |
| Antipsychotics (90) | 0.008 | 0.006 |
| Antipsychotics (180) | 0.006 | 0.005 |
| 2019 |  |  |
| Sedative (90) | 0.064 | 0.039 |
| Sedative (180) | 0.039 | 0.024 |
| Benzodiazepines and z-drugs (90) | 0.068 | 0.042 |
| Benzodiazepines and z-drugs after (180) | 0.043 | 0.027 |
| Long-acting benzodiazepines (90) | 0.038 | 0.024 |
| Long-acting benzodiazepines (180) | 0.028 | 0.018 |
| Clonazepam (90) | 0.002 | 0.002 |
| Clonazepam (180) | 0.002 | 0.001 |
| Diazepam (90) | 0.033 | 0.019 |
| Diazepam (180) | 0.025 | 0.014 |
| Chlordiazepoxide (90) | 0.002 | 0.005 |
| Chlordiazepoxide (180) | 0.002 | 0.004 |
| Clorazepate (90) | 0.000 | 0.000 |
| Clorazepate (180) | 0.000 | 0.000 |
| Lorazepam (90) | 0.002 | 0.002 |
| Lorazepam (180) | 0.002 | 0.002 |
| Bromazepam (90) | 0.003 | 0.001 |
| Bromazepam (180) | 0.002 | 0.001 |
| Clobazam (90) | 0.000 | 0.000 |
| Clobazam (180) | 0.000 | 0.000 |
| Prazepam (90) | 0.001 | 0.000 |
| Prazepam (180) | 0.000 | 0.000 |
| Alprazolam (90) | 0.030 | 0.013 |
| Alprazolam (180) | 0.022 | 0.010 |
| Flurazepam (90) | 0.002 | 0.002 |
| Flurazepam (180) | 0.002 | 0.002 |
| Nitrazepam (90) | 0.000 | 0.000 |
| Nitrazepam (180) | 0.000 | 0.000 |
| Triazolam (90) | 0.001 | 0.001 |
| Triazolam (180) | 0.001 | 0.001 |
| Lormetazepam (90) | 0.003 | 0.001 |
| Lormetazepam (180) | 0.002 | 0.001 |
| Temazepam (90) | 0.002 | 0.002 |
| Temazepam (180) | 0.002 | 0.001 |
| Zopiclone (90) | 0.019 | 0.013 |
| Zopiclone (180) | 0.014 | 0.010 |
| Zolpidem (90) | 0.020 | 0.012 |
| Zolpidem (180) | 0.015 | 0.009 |
| Antidepressant (90) | 0.012 | 0.008 |
| Antidepressant (180) | 0.009 | 0.006 |
| Antihistamines (90) | 0.006 | 0.004 |
| Antihistamines (180) | 0.005 | 0.003 |
| Antipsychotics (90) | 0.008 | 0.007 |
| Antipsychotics (180) | 0.006 | 0.005 |
| 2020 |  |  |
| Sedative (90) | 0.056 | 0.034 |
| Sedative (180) | 0.034 | 0.021 |
| Benzodiazepines and z-drugs (90) | 0.059 | 0.036 |
| Benzodiazepines and z-drugs after (180) | 0.040 | 0.025 |
| Long-acting benzodiazepines (90) | 0.033 | 0.022 |
| Long-acting benzodiazepines (180) | 0.026 | 0.016 |
| Clonazepam (90) | 0.002 | 0.002 |
| Clonazepam (180) | 0.002 | 0.001 |
| Diazepam (90) | 0.030 | 0.016 |
| Diazepam (180) | 0.023 | 0.013 |
| Chlordiazepoxide (90) | 0.002 | 0.004 |
| Chlordiazepoxide (180) | 0.002 | 0.003 |
| Clorazepate (90) | 0.000 | 0.000 |
| Clorazepate (180) | 0.000 | 0.000 |
| Lorazepam (90) | 0.002 | 0.002 |
| Lorazepam (180) | 0.002 | 0.001 |
| Bromazepam (90) | 0.003 | 0.001 |
| Bromazepam (180) | 0.002 | 0.001 |
| Clobazam (90) | 0.000 | 0.000 |
| Clobazam (180) | 0.000 | 0.000 |
| Prazepam (90) | 0.000 | 0.000 |
| Prazepam (180) | 0.000 | 0.000 |
| Alprazolam (90) | 0.026 | 0.011 |
| Alprazolam (180) | 0.020 | 0.009 |
| Flurazepam (90) | 0.002 | 0.002 |
| Flurazepam (180) | 0.001 | 0.001 |
| Nitrazepam (90) | 0.000 | 0.000 |
| Nitrazepam (180) | 0.000 | 0.000 |
| Triazolam (90) | 0.001 | 0.001 |
| Triazolam (180) | 0.001 | 0.001 |
| Lormetazepam (90) | 0.001 | 0.000 |
| Lormetazepam (180) | 0.002 | 0.001 |
| Temazepam (90) | 0.002 | 0.001 |
| Temazepam (180) | 0.002 | 0.001 |
| Zopiclone (90) | 0.017 | 0.011 |
| Zopiclone (180) | 0.013 | 0.009 |
| Zolpidem (90) | 0.017 | 0.010 |
| Zolpidem (180) | 0.013 | 0.008 |
| Antidepressant (90) | 0.011 | 0.007 |
| Antidepressant (180) | 0.009 | 0.006 |
| Antihistamines (90) | 0.007 | 0.004 |
| Antihistamines (180) | 0.005 | 0.003 |
| Antipsychotics (90) | 0.008 | 0.006 |
| Antipsychotics (180) | 0.006 | 0.005 |
| 2021 |  |  |
| Sedative (90) | 0.057 | 0.035 |
| Sedative (180) | 0.035 | 0.021 |
| Benzodiazepines and z-drugs (90) | 0.061 | 0.037 |
| Benzodiazepines and z-drugs after (180) | 0.041 | 0.025 |
| Long-acting benzodiazepines (90) | 0.035 | 0.022 |
| Long-acting benzodiazepines (180) | 0.027 | 0.016 |
| Clonazepam (90) | 0.002 | 0.002 |
| Clonazepam (180) | 0.002 | 0.001 |
| Diazepam (90) | 0.030 | 0.017 |
| Diazepam (180) | 0.023 | 0.013 |
| Chlordiazepoxide (90) | 0.002 | 0.004 |
| Chlordiazepoxide (180) | 0.002 | 0.003 |
| Clorazepate (90) | 0.000 | 0.000 |
| Clorazepate (180) | 0.000 | 0.000 |
| Lorazepam (90) | 0.002 | 0.002 |
| Lorazepam (180) | 0.002 | 0.002 |
| Bromazepam (90) | 0.003 | 0.001 |
| Bromazepam (180) | 0.002 | 0.001 |
| Clobazam (90) | 0.000 | 0.000 |
| Clobazam (180) | 0.000 | 0.000 |
| Prazepam (90) | 0.000 | 0.000 |
| Prazepam (180) | 0.000 | 0.000 |
| Alprazolam (90) | 0.025 | 0.011 |
| Alprazolam (180) | 0.019 | 0.008 |
| Flurazepam (90) | 0.002 | 0.002 |
| Flurazepam (180) | 0.001 | 0.001 |
| Nitrazepam (90) | 0.001 | 0.000 |
| Nitrazepam (180) | 0.001 | 0.000 |
| Triazolam (90) | 0.001 | 0.001 |
| Triazolam (180) | 0.001 | 0.001 |
| Lormetazepam (90) | 0.000 | 0.000 |
| Lormetazepam (180) | 0.000 | 0.000 |
| Temazepam (90) | 0.002 | 0.001 |
| Temazepam (180) | 0.002 | 0.001 |
| Zopiclone (90) | 0.017 | 0.011 |
| Zopiclone (180) | 0.013 | 0.008 |
| Zolpidem (90) | 0.017 | 0.010 |
| Zolpidem (180) | 0.013 | 0.008 |
| Antidepressant (90) | 0.012 | 0.008 |
| Antidepressant (180) | 0.010 | 0.006 |
| Antihistamines (90) | 0.008 | 0.005 |
| Antihistamines (180) | 0.007 | 0.004 |
| Antipsychotics (90) | 0.009 | 0.007 |
| Antipsychotics (180) | 0.007 | 0.005 |
| 2022 |  |  |
| Sedative (90) | 0.057 | 0.034 |
| Sedative (180) | 0.035 | 0.021 |
| Benzodiazepines and z-drugs (90) | 0.061 | 0.037 |
| Benzodiazepines and z-drugs after (180) | 0.040 | 0.025 |
| Long-acting benzodiazepines (90) | 0.034 | 0.022 |
| Long-acting benzodiazepines (180) | 0.026 | 0.016 |
| Clonazepam (90) | 0.003 | 0.002 |
| Clonazepam (180) | 0.002 | 0.001 |
| Diazepam (90) | 0.030 | 0.017 |
| Diazepam (180) | 0.023 | 0.013 |
| Chlordiazepoxide (90) | 0.002 | 0.004 |
| Chlordiazepoxide (180) | 0.002 | 0.003 |
| Clorazepate (90) | 0.000 | 0.000 |
| Clorazepate (180) | 0.000 | 0.000 |
| Lorazepam (90) | 0.002 | 0.002 |
| Lorazepam (180) | 0.002 | 0.001 |
| Bromazepam (90) | 0.002 | 0.001 |
| Bromazepam (180) | 0.002 | 0.001 |
| Clobazam (90) | 0.000 | 0.000 |
| Clobazam (180) | 0.000 | 0.000 |
| Prazepam (90) | 0.001 | 0.000 |
| Prazepam (180) | 0.000 | 0.000 |
| Alprazolam (90) | 0.027 | 0.011 |
| Alprazolam (180) | 0.019 | 0.008 |
| Flurazepam (90) | 0.001 | 0.001 |
| Flurazepam (180) | 0.001 | 0.001 |
| Nitrazepam (90) | 0.000 | 0.000 |
| Nitrazepam (180) | 0.000 | 0.000 |
| Triazolam (90) | 0.001 | 0.001 |
| Triazolam (180) | 0.001 | 0.000 |
| Lormetazepam (90) | 0.000 | 0.000 |
| Lormetazepam (180) | 0.000 | 0.000 |
| Temazepam (90) | 0.002 | 0.001 |
| Temazepam (180) | 0.001 | 0.001 |
| Zopiclone (90) | 0.016 | 0.011 |
| Zopiclone (180) | 0.012 | 0.008 |
| Zolpidem (90) | 0.016 | 0.009 |
| Zolpidem (180) | 0.012 | 0.007 |
| Antidepressant (90) | 0.012 | 0.008 |
| Antidepressant (180) | 0.010 | 0.006 |
| Antihistamines (90) | 0.009 | 0.006 |
| Antihistamines (180) | 0.007 | 0.004 |
| Antipsychotics (90) | 0.009 | 0.007 |
| Antipsychotics (180) | 0.007 | 0.005 |

**Prevalence of chronic use >30/>90 days by age group**

|  | <5 | 5-11 | 12-15 | 16-24 | 25-34 | 35-44 | 45-54 | 55-64 | 65-69 | 70-74 | 75+ |
| --- | --- | --- | --- | --- | --- | --- | --- | --- | --- | --- | --- |
| 2014 |  |  |  |  |  |  |  |  |  |  |  |
| Sedative (30) | 0.001 | 0.001 | 0.002 | 0.025 | 0.076 | 0.105 | 0.147 | 0.198 | 0.218 | 0.227 | 0.315 |
| Sedative (90) | 0.001 | 0.001 | 0.001 | 0.013 | 0.048 | 0.074 | 0.111 | 0.158 | 0.181 | 0.189 | 0.272 |
| Benzodiazepines and z-drugs (30) | 0.001 | 0.001 | 0.002 | 0.020 | 0.067 | 0.096 | 0.136 | 0.187 | 0.208 | 0.218 | 0.295 |
| Benzodiazepines and z-drugs after (90) | 0.001 | 0.001 | 0.001 | 0.010 | 0.042 | 0.066 | 0.101 | 0.149 | 0.172 | 0.180 | 0.251 |
| Long-acting benzodiazepines (30) | 0.001 | 0.001 | 0.001 | 0.009 | 0.033 | 0.045 | 0.055 | 0.065 | 0.063 | 0.057 | 0.061 |
| Long-acting benzodiazepines (90) | 0.001 | 0.001 | 0.001 | 0.005 | 0.020 | 0.030 | 0.038 | 0.048 | 0.049 | 0.045 | 0.048 |
| Clonazepam (30) | 0.000 | 0.000 | 0.000 | 0.001 | 0.004 | 0.005 | 0.007 | 0.007 | 0.006 | 0.006 | 0.006 |
| Clonazepam (90) | 0.000 | 0.000 | 0.000 | 0.001 | 0.003 | 0.004 | 0.006 | 0.006 | 0.005 | 0.005 | 0.005 |
| Diazepam (30) | 0.000 | 0.000 | 0.001 | 0.006 | 0.023 | 0.031 | 0.034 | 0.038 | 0.036 | 0.033 | 0.034 |
| Diazepam (90) | 0.000 | 0.000 | 0.000 | 0.003 | 0.014 | 0.020 | 0.022 | 0.027 | 0.027 | 0.024 | 0.025 |
| Chlordiazepoxide (30) | 0.000 | 0.000 | 0.000 | 0.000 | 0.002 | 0.004 | 0.006 | 0.005 | 0.003 | 0.002 | 0.002 |
| Chlordiazepoxide (90) | 0.000 | 0.000 | 0.000 | 0.000 | 0.001 | 0.001 | 0.002 | 0.002 | 0.001 | 0.002 | 0.002 |
| Clorazepate (30) | 0.000 | 0.000 | 0.000 | 0.000 | 0.000 | 0.000 | 0.000 | 0.000 | 0.000 | 0.000 | 0.000 |
| Clorazepate (90) | 0.000 | 0.000 | 0.000 | 0.000 | 0.000 | 0.000 | 0.000 | 0.000 | 0.000 | 0.000 | 0.000 |
| Lorazepam (30) | 0.000 | 0.000 | 0.000 | 0.001 | 0.001 | 0.002 | 0.003 | 0.004 | 0.005 | 0.005 | 0.008 |
| Lorazepam (90) | 0.000 | 0.000 | 0.000 | 0.000 | 0.001 | 0.001 | 0.002 | 0.003 | 0.004 | 0.004 | 0.006 |
| Bromazepam (30) | 0.000 | 0.000 | 0.000 | 0.000 | 0.001 | 0.002 | 0.004 | 0.009 | 0.013 | 0.013 | 0.014 |
| Bromazepam (90) | 0.000 | 0.000 | 0.000 | 0.000 | 0.001 | 0.001 | 0.003 | 0.007 | 0.011 | 0.011 | 0.012 |
| Clobazam (30) | 0.000 | 0.001 | 0.001 | 0.001 | 0.001 | 0.001 | 0.002 | 0.002 | 0.002 | 0.001 | 0.001 |
| Clobazam (90) | 0.000 | 0.000 | 0.000 | 0.001 | 0.001 | 0.001 | 0.001 | 0.001 | 0.001 | 0.001 | 0.001 |
| Prazepam (30) | 0.000 | 0.000 | 0.000 | 0.000 | 0.000 | 0.000 | 0.001 | 0.002 | 0.002 | 0.003 | 0.002 |
| Prazepam (90) | 0.000 | 0.000 | 0.000 | 0.000 | 0.000 | 0.000 | 0.001 | 0.002 | 0.002 | 0.002 | 0.002 |
| Alprazolam (30) | 0.000 | 0.000 | 0.000 | 0.004 | 0.015 | 0.020 | 0.027 | 0.037 | 0.040 | 0.039 | 0.052 |
| Alprazolam (90) | 0.000 | 0.000 | 0.000 | 0.002 | 0.008 | 0.012 | 0.018 | 0.027 | 0.029 | 0.029 | 0.039 |
| Flurazepam (30) | 0.000 | 0.000 | 0.000 | 0.001 | 0.004 | 0.008 | 0.011 | 0.015 | 0.015 | 0.012 | 0.012 |
| Flurazepam (90) | 0.000 | 0.000 | 0.000 | 0.000 | 0.003 | 0.006 | 0.009 | 0.012 | 0.012 | 0.010 | 0.010 |
| Nitrazepam (30) | 0.000 | 0.000 | 0.000 | 0.000 | 0.000 | 0.000 | 0.001 | 0.001 | 0.002 | 0.003 | 0.005 |
| Nitrazepam (90) | 0.000 | 0.000 | 0.000 | 0.000 | 0.000 | 0.000 | 0.001 | 0.001 | 0.002 | 0.002 | 0.005 |
| Triazolam (30) | 0.000 | 0.000 | 0.000 | 0.000 | 0.002 | 0.002 | 0.004 | 0.006 | 0.008 | 0.009 | 0.014 |
| Triazolam (90) | 0.000 | 0.000 | 0.000 | 0.000 | 0.001 | 0.002 | 0.003 | 0.005 | 0.007 | 0.008 | 0.012 |
| Lormetazepam (30) | 0.000 | 0.000 | 0.000 | 0.000 | 0.000 | 0.001 | 0.002 | 0.003 | 0.005 | 0.007 | 0.012 |
| Lormetazepam (90) | 0.000 | 0.000 | 0.000 | 0.000 | 0.000 | 0.000 | 0.001 | 0.002 | 0.004 | 0.006 | 0.010 |
| Temazepam (30) | 0.000 | 0.000 | 0.000 | 0.000 | 0.001 | 0.002 | 0.004 | 0.008 | 0.012 | 0.013 | 0.027 |
| Temazepam (90) | 0.000 | 0.000 | 0.000 | 0.000 | 0.001 | 0.001 | 0.003 | 0.006 | 0.010 | 0.010 | 0.022 |
| Zopiclone (30) | 0.000 | 0.000 | 0.000 | 0.004 | 0.016 | 0.025 | 0.036 | 0.052 | 0.055 | 0.060 | 0.086 |
| Zopiclone (90) | 0.000 | 0.000 | 0.000 | 0.002 | 0.011 | 0.018 | 0.028 | 0.040 | 0.044 | 0.049 | 0.071 |
| Zolpidem (30) | 0.000 | 0.000 | 0.000 | 0.003 | 0.009 | 0.016 | 0.027 | 0.042 | 0.048 | 0.053 | 0.071 |
| Zolpidem (90) | 0.000 | 0.000 | 0.000 | 0.001 | 0.005 | 0.010 | 0.020 | 0.032 | 0.038 | 0.041 | 0.058 |
| Antidepressant (30) | 0.000 | 0.000 | 0.000 | 0.002 | 0.006 | 0.009 | 0.012 | 0.014 | 0.014 | 0.015 | 0.030 |
| Antidepressant (90) | 0.000 | 0.000 | 0.000 | 0.001 | 0.003 | 0.005 | 0.008 | 0.009 | 0.010 | 0.011 | 0.024 |
| Antihistamines (30) | 0.000 | 0.000 | 0.000 | 0.001 | 0.001 | 0.001 | 0.002 | 0.002 | 0.001 | 0.001 | 0.001 |
| Antihistamines (90) | 0.000 | 0.000 | 0.000 | 0.000 | 0.001 | 0.001 | 0.001 | 0.001 | 0.001 | 0.001 | 0.001 |
| Antipsychotics (30) | 0.000 | 0.000 | 0.000 | 0.004 | 0.009 | 0.010 | 0.013 | 0.014 | 0.014 | 0.017 | 0.041 |
| Antipsychotics (90) | 0.000 | 0.000 | 0.000 | 0.002 | 0.006 | 0.007 | 0.010 | 0.011 | 0.011 | 0.014 | 0.034 |
| 2015 |  |  |  |  |  |  |  |  |  |  |  |
| Sedative (30) | 0.001 | 0.001 | 0.002 | 0.027 | 0.078 | 0.105 | 0.143 | 0.191 | 0.211 | 0.218 | 0.302 |
| Sedative (90) | 0.000 | 0.001 | 0.001 | 0.016 | 0.052 | 0.077 | 0.112 | 0.159 | 0.182 | 0.187 | 0.270 |
| Benzodiazepines and z-drugs (30) | 0.001 | 0.001 | 0.002 | 0.021 | 0.068 | 0.094 | 0.131 | 0.180 | 0.201 | 0.206 | 0.278 |
| Benzodiazepines and z-drugs after (90) | 0.000 | 0.001 | 0.001 | 0.011 | 0.044 | 0.067 | 0.101 | 0.148 | 0.171 | 0.175 | 0.245 |
| Long-acting benzodiazepines (30) | 0.001 | 0.001 | 0.001 | 0.009 | 0.033 | 0.044 | 0.053 | 0.062 | 0.060 | 0.055 | 0.058 |
| Long-acting benzodiazepines (90) | 0.000 | 0.001 | 0.001 | 0.005 | 0.021 | 0.031 | 0.038 | 0.048 | 0.049 | 0.044 | 0.047 |
| Clonazepam (30) | 0.000 | 0.000 | 0.000 | 0.002 | 0.004 | 0.005 | 0.007 | 0.007 | 0.007 | 0.006 | 0.006 |
| Clonazepam (90) | 0.000 | 0.000 | 0.000 | 0.001 | 0.003 | 0.004 | 0.006 | 0.006 | 0.006 | 0.005 | 0.005 |
| Diazepam (30) | 0.000 | 0.000 | 0.001 | 0.006 | 0.024 | 0.031 | 0.033 | 0.037 | 0.036 | 0.032 | 0.033 |
| Diazepam (90) | 0.000 | 0.000 | 0.000 | 0.003 | 0.015 | 0.021 | 0.023 | 0.027 | 0.028 | 0.024 | 0.026 |
| Chlordiazepoxide (30) | 0.000 | 0.000 | 0.000 | 0.000 | 0.002 | 0.004 | 0.005 | 0.005 | 0.003 | 0.002 | 0.002 |
| Chlordiazepoxide (90) | 0.000 | 0.000 | 0.000 | 0.000 | 0.001 | 0.001 | 0.002 | 0.002 | 0.002 | 0.001 | 0.002 |
| Clorazepate (30) | 0.000 | 0.000 | 0.000 | 0.000 | 0.000 | 0.000 | 0.000 | 0.000 | 0.000 | 0.000 | 0.000 |
| Clorazepate (90) | 0.000 | 0.000 | 0.000 | 0.000 | 0.000 | 0.000 | 0.000 | 0.000 | 0.000 | 0.000 | 0.000 |
| Lorazepam (30) | 0.000 | 0.000 | 0.000 | 0.001 | 0.002 | 0.002 | 0.002 | 0.004 | 0.005 | 0.005 | 0.008 |
| Lorazepam (90) | 0.000 | 0.000 | 0.000 | 0.000 | 0.001 | 0.001 | 0.002 | 0.003 | 0.004 | 0.004 | 0.006 |
| Bromazepam (30) | 0.000 | 0.000 | 0.000 | 0.000 | 0.001 | 0.002 | 0.004 | 0.008 | 0.012 | 0.012 | 0.013 |
| Bromazepam (90) | 0.000 | 0.000 | 0.000 | 0.000 | 0.001 | 0.001 | 0.003 | 0.007 | 0.011 | 0.010 | 0.012 |
| Clobazam (30) | 0.000 | 0.000 | 0.000 | 0.001 | 0.001 | 0.001 | 0.002 | 0.001 | 0.001 | 0.001 | 0.001 |
| Clobazam (90) | 0.000 | 0.000 | 0.000 | 0.001 | 0.001 | 0.001 | 0.001 | 0.001 | 0.001 | 0.001 | 0.001 |
| Prazepam (30) | 0.000 | 0.000 | 0.000 | 0.000 | 0.000 | 0.000 | 0.001 | 0.002 | 0.002 | 0.003 | 0.002 |
| Prazepam (90) | 0.000 | 0.000 | 0.000 | 0.000 | 0.000 | 0.000 | 0.001 | 0.002 | 0.002 | 0.002 | 0.002 |
| Alprazolam (30) | 0.000 | 0.000 | 0.000 | 0.005 | 0.016 | 0.020 | 0.027 | 0.036 | 0.038 | 0.039 | 0.051 |
| Alprazolam (90) | 0.000 | 0.000 | 0.000 | 0.002 | 0.009 | 0.013 | 0.019 | 0.027 | 0.030 | 0.030 | 0.041 |
| Flurazepam (30) | 0.000 | 0.000 | 0.000 | 0.001 | 0.004 | 0.007 | 0.010 | 0.014 | 0.013 | 0.011 | 0.010 |
| Flurazepam (90) | 0.000 | 0.000 | 0.000 | 0.000 | 0.003 | 0.006 | 0.009 | 0.012 | 0.012 | 0.010 | 0.009 |
| Nitrazepam (30) | 0.000 | 0.000 | 0.000 | 0.000 | 0.000 | 0.000 | 0.001 | 0.001 | 0.002 | 0.002 | 0.005 |
| Nitrazepam (90) | 0.000 | 0.000 | 0.000 | 0.000 | 0.000 | 0.000 | 0.001 | 0.001 | 0.002 | 0.002 | 0.004 |
| Triazolam (30) | 0.000 | 0.000 | 0.000 | 0.000 | 0.002 | 0.002 | 0.004 | 0.006 | 0.008 | 0.008 | 0.013 |
| Triazolam (90) | 0.000 | 0.000 | 0.000 | 0.000 | 0.002 | 0.002 | 0.003 | 0.005 | 0.007 | 0.007 | 0.012 |
| Lormetazepam (30) | 0.000 | 0.000 | 0.000 | 0.000 | 0.000 | 0.001 | 0.001 | 0.003 | 0.004 | 0.006 | 0.011 |
| Lormetazepam (90) | 0.000 | 0.000 | 0.000 | 0.000 | 0.000 | 0.000 | 0.001 | 0.002 | 0.004 | 0.005 | 0.010 |
| Temazepam (30) | 0.000 | 0.000 | 0.000 | 0.000 | 0.001 | 0.002 | 0.004 | 0.007 | 0.010 | 0.012 | 0.023 |
| Temazepam (90) | 0.000 | 0.000 | 0.000 | 0.000 | 0.001 | 0.001 | 0.003 | 0.006 | 0.009 | 0.010 | 0.021 |
| Zopiclone (30) | 0.000 | 0.000 | 0.000 | 0.004 | 0.015 | 0.025 | 0.036 | 0.051 | 0.055 | 0.057 | 0.083 |
| Zopiclone (90) | 0.000 | 0.000 | 0.000 | 0.002 | 0.011 | 0.019 | 0.029 | 0.042 | 0.046 | 0.048 | 0.072 |
| Zolpidem (30) | 0.000 | 0.000 | 0.000 | 0.002 | 0.009 | 0.016 | 0.027 | 0.041 | 0.047 | 0.050 | 0.068 |
| Zolpidem (90) | 0.000 | 0.000 | 0.000 | 0.001 | 0.005 | 0.011 | 0.020 | 0.033 | 0.039 | 0.041 | 0.059 |
| Antidepressant (30) | 0.000 | 0.000 | 0.000 | 0.003 | 0.007 | 0.010 | 0.013 | 0.015 | 0.015 | 0.017 | 0.033 |
| Antidepressant (90) | 0.000 | 0.000 | 0.000 | 0.002 | 0.004 | 0.006 | 0.009 | 0.011 | 0.012 | 0.014 | 0.027 |
| Antihistamines (30) | 0.000 | 0.000 | 0.000 | 0.001 | 0.002 | 0.002 | 0.003 | 0.002 | 0.002 | 0.001 | 0.002 |
| Antihistamines (90) | 0.000 | 0.000 | 0.000 | 0.000 | 0.001 | 0.001 | 0.002 | 0.002 | 0.001 | 0.001 | 0.001 |
| Antipsychotics (30) | 0.000 | 0.000 | 0.000 | 0.006 | 0.011 | 0.011 | 0.014 | 0.015 | 0.015 | 0.017 | 0.044 |
| Antipsychotics (90) | 0.000 | 0.000 | 0.000 | 0.003 | 0.007 | 0.009 | 0.011 | 0.012 | 0.013 | 0.015 | 0.038 |
| 2016 |  |  |  |  |  |  |  |  |  |  |  |
| Sedative (30) | 0.001 | 0.001 | 0.002 | 0.028 | 0.086 | 0.110 | 0.145 | 0.190 | 0.213 | 0.218 | 0.302 |
| Sedative (90) | 0.000 | 0.001 | 0.001 | 0.017 | 0.059 | 0.080 | 0.113 | 0.157 | 0.182 | 0.187 | 0.271 |
| Benzodiazepines and z-drugs (30) | 0.001 | 0.001 | 0.002 | 0.021 | 0.073 | 0.097 | 0.131 | 0.176 | 0.201 | 0.205 | 0.275 |
| Benzodiazepines and z-drugs after (90) | 0.000 | 0.001 | 0.001 | 0.011 | 0.047 | 0.069 | 0.100 | 0.144 | 0.170 | 0.174 | 0.242 |
| Long-acting benzodiazepines (30) | 0.001 | 0.001 | 0.001 | 0.009 | 0.035 | 0.046 | 0.053 | 0.060 | 0.060 | 0.054 | 0.055 |
| Long-acting benzodiazepines (90) | 0.000 | 0.001 | 0.001 | 0.005 | 0.023 | 0.032 | 0.038 | 0.046 | 0.048 | 0.043 | 0.046 |
| Clonazepam (30) | 0.000 | 0.000 | 0.000 | 0.002 | 0.005 | 0.005 | 0.007 | 0.008 | 0.007 | 0.006 | 0.006 |
| Clonazepam (90) | 0.000 | 0.000 | 0.000 | 0.001 | 0.004 | 0.005 | 0.006 | 0.006 | 0.006 | 0.005 | 0.006 |
| Diazepam (30) | 0.000 | 0.000 | 0.001 | 0.006 | 0.025 | 0.032 | 0.034 | 0.036 | 0.036 | 0.031 | 0.032 |
| Diazepam (90) | 0.000 | 0.000 | 0.000 | 0.003 | 0.016 | 0.022 | 0.023 | 0.026 | 0.027 | 0.023 | 0.025 |
| Chlordiazepoxide (30) | 0.000 | 0.000 | 0.000 | 0.000 | 0.002 | 0.004 | 0.005 | 0.005 | 0.003 | 0.002 | 0.002 |
| Chlordiazepoxide (90) | 0.000 | 0.000 | 0.000 | 0.000 | 0.001 | 0.002 | 0.002 | 0.002 | 0.001 | 0.001 | 0.001 |
| Clorazepate (30) | 0.000 | 0.000 | 0.000 | 0.000 | 0.000 | 0.000 | 0.000 | 0.000 | 0.000 | 0.000 | 0.000 |
| Clorazepate (90) | 0.000 | 0.000 | 0.000 | 0.000 | 0.000 | 0.000 | 0.000 | 0.000 | 0.000 | 0.000 | 0.000 |
| Lorazepam (30) | 0.000 | 0.000 | 0.000 | 0.001 | 0.002 | 0.002 | 0.002 | 0.004 | 0.004 | 0.005 | 0.007 |
| Lorazepam (90) | 0.000 | 0.000 | 0.000 | 0.000 | 0.001 | 0.001 | 0.002 | 0.003 | 0.004 | 0.004 | 0.006 |
| Bromazepam (30) | 0.000 | 0.000 | 0.000 | 0.000 | 0.001 | 0.002 | 0.004 | 0.008 | 0.012 | 0.012 | 0.013 |
| Bromazepam (90) | 0.000 | 0.000 | 0.000 | 0.000 | 0.001 | 0.001 | 0.003 | 0.006 | 0.010 | 0.010 | 0.012 |
| Clobazam (30) | 0.000 | 0.000 | 0.000 | 0.001 | 0.001 | 0.001 | 0.001 | 0.001 | 0.001 | 0.001 | 0.001 |
| Clobazam (90) | 0.000 | 0.000 | 0.000 | 0.001 | 0.001 | 0.001 | 0.001 | 0.001 | 0.001 | 0.001 | 0.001 |
| Prazepam (30) | 0.000 | 0.000 | 0.000 | 0.000 | 0.000 | 0.000 | 0.001 | 0.002 | 0.002 | 0.002 | 0.002 |
| Prazepam (90) | 0.000 | 0.000 | 0.000 | 0.000 | 0.000 | 0.000 | 0.001 | 0.001 | 0.002 | 0.002 | 0.002 |
| Alprazolam (30) | 0.000 | 0.000 | 0.000 | 0.004 | 0.018 | 0.021 | 0.027 | 0.035 | 0.039 | 0.038 | 0.052 |
| Alprazolam (90) | 0.000 | 0.000 | 0.000 | 0.002 | 0.010 | 0.013 | 0.019 | 0.027 | 0.030 | 0.030 | 0.041 |
| Flurazepam (30) | 0.000 | 0.000 | 0.000 | 0.001 | 0.004 | 0.007 | 0.010 | 0.013 | 0.013 | 0.011 | 0.010 |
| Flurazepam (90) | 0.000 | 0.000 | 0.000 | 0.000 | 0.003 | 0.006 | 0.009 | 0.011 | 0.011 | 0.010 | 0.008 |
| Nitrazepam (30) | 0.000 | 0.000 | 0.000 | 0.000 | 0.000 | 0.000 | 0.001 | 0.001 | 0.002 | 0.002 | 0.004 |
| Nitrazepam (90) | 0.000 | 0.000 | 0.000 | 0.000 | 0.000 | 0.000 | 0.001 | 0.001 | 0.002 | 0.002 | 0.004 |
| Triazolam (30) | 0.000 | 0.000 | 0.000 | 0.000 | 0.002 | 0.002 | 0.003 | 0.006 | 0.008 | 0.008 | 0.013 |
| Triazolam (90) | 0.000 | 0.000 | 0.000 | 0.000 | 0.002 | 0.002 | 0.003 | 0.005 | 0.007 | 0.007 | 0.012 |
| Lormetazepam (30) | 0.000 | 0.000 | 0.000 | 0.000 | 0.000 | 0.000 | 0.001 | 0.003 | 0.004 | 0.006 | 0.011 |
| Lormetazepam (90) | 0.000 | 0.000 | 0.000 | 0.000 | 0.000 | 0.000 | 0.001 | 0.002 | 0.003 | 0.005 | 0.010 |
| Temazepam (30) | 0.000 | 0.000 | 0.000 | 0.000 | 0.001 | 0.002 | 0.003 | 0.007 | 0.010 | 0.011 | 0.022 |
| Temazepam (90) | 0.000 | 0.000 | 0.000 | 0.000 | 0.001 | 0.001 | 0.003 | 0.005 | 0.009 | 0.009 | 0.019 |
| Zopiclone (30) | 0.000 | 0.000 | 0.000 | 0.005 | 0.017 | 0.026 | 0.036 | 0.050 | 0.055 | 0.057 | 0.083 |
| Zopiclone (90) | 0.000 | 0.000 | 0.000 | 0.002 | 0.012 | 0.020 | 0.029 | 0.042 | 0.046 | 0.048 | 0.072 |
| Zolpidem (30) | 0.000 | 0.000 | 0.000 | 0.003 | 0.010 | 0.016 | 0.027 | 0.041 | 0.048 | 0.051 | 0.069 |
| Zolpidem (90) | 0.000 | 0.000 | 0.000 | 0.001 | 0.006 | 0.011 | 0.020 | 0.032 | 0.040 | 0.042 | 0.059 |
| Antidepressant (30) | 0.000 | 0.000 | 0.000 | 0.003 | 0.009 | 0.011 | 0.014 | 0.016 | 0.016 | 0.019 | 0.036 |
| Antidepressant (90) | 0.000 | 0.000 | 0.000 | 0.002 | 0.006 | 0.007 | 0.010 | 0.012 | 0.013 | 0.015 | 0.030 |
| Antihistamines (30) | 0.000 | 0.000 | 0.000 | 0.001 | 0.003 | 0.003 | 0.003 | 0.003 | 0.002 | 0.002 | 0.002 |
| Antihistamines (90) | 0.000 | 0.000 | 0.000 | 0.001 | 0.002 | 0.002 | 0.002 | 0.002 | 0.001 | 0.001 | 0.001 |
| Antipsychotics (30) | 0.000 | 0.000 | 0.000 | 0.007 | 0.013 | 0.013 | 0.015 | 0.016 | 0.016 | 0.017 | 0.046 |
| Antipsychotics (90) | 0.000 | 0.000 | 0.000 | 0.004 | 0.009 | 0.010 | 0.012 | 0.013 | 0.014 | 0.015 | 0.040 |
| 2017 |  |  |  |  |  |  |  |  |  |  |  |
| Sedative (30) | 0.001 | 0.001 | 0.002 | 0.029 | 0.094 | 0.119 | 0.150 | 0.190 | 0.206 | 0.211 | 0.292 |
| Sedative (90) | 0.000 | 0.001 | 0.001 | 0.017 | 0.065 | 0.088 | 0.118 | 0.157 | 0.177 | 0.183 | 0.262 |
| Benzodiazepines and z-drugs (30) | 0.001 | 0.001 | 0.002 | 0.020 | 0.076 | 0.103 | 0.133 | 0.174 | 0.193 | 0.197 | 0.262 |
| Benzodiazepines and z-drugs after (90) | 0.000 | 0.001 | 0.001 | 0.011 | 0.051 | 0.074 | 0.103 | 0.142 | 0.164 | 0.169 | 0.231 |
| Long-acting benzodiazepines (30) | 0.001 | 0.001 | 0.001 | 0.009 | 0.037 | 0.049 | 0.054 | 0.059 | 0.057 | 0.052 | 0.052 |
| Long-acting benzodiazepines (90) | 0.000 | 0.001 | 0.001 | 0.005 | 0.025 | 0.034 | 0.040 | 0.045 | 0.046 | 0.042 | 0.043 |
| Clonazepam (30) | 0.000 | 0.000 | 0.000 | 0.002 | 0.006 | 0.006 | 0.007 | 0.008 | 0.007 | 0.007 | 0.007 |
| Clonazepam (90) | 0.000 | 0.000 | 0.000 | 0.001 | 0.004 | 0.005 | 0.006 | 0.007 | 0.006 | 0.006 | 0.006 |
| Diazepam (30) | 0.000 | 0.000 | 0.001 | 0.006 | 0.027 | 0.035 | 0.035 | 0.036 | 0.034 | 0.029 | 0.030 |
| Diazepam (90) | 0.000 | 0.000 | 0.000 | 0.003 | 0.017 | 0.024 | 0.025 | 0.026 | 0.027 | 0.023 | 0.023 |
| Chlordiazepoxide (30) | 0.000 | 0.000 | 0.000 | 0.000 | 0.003 | 0.004 | 0.005 | 0.004 | 0.003 | 0.002 | 0.002 |
| Chlordiazepoxide (90) | 0.000 | 0.000 | 0.000 | 0.000 | 0.001 | 0.002 | 0.002 | 0.002 | 0.001 | 0.001 | 0.001 |
| Clorazepate (30) | 0.000 | 0.000 | 0.000 | 0.000 | 0.000 | 0.000 | 0.000 | 0.000 | 0.000 | 0.000 | 0.000 |
| Clorazepate (90) | 0.000 | 0.000 | 0.000 | 0.000 | 0.000 | 0.000 | 0.000 | 0.000 | 0.000 | 0.000 | 0.000 |
| Lorazepam (30) | 0.000 | 0.000 | 0.000 | 0.001 | 0.002 | 0.002 | 0.003 | 0.004 | 0.004 | 0.004 | 0.007 |
| Lorazepam (90) | 0.000 | 0.000 | 0.000 | 0.000 | 0.001 | 0.001 | 0.002 | 0.003 | 0.003 | 0.004 | 0.005 |
| Bromazepam (30) | 0.000 | 0.000 | 0.000 | 0.000 | 0.001 | 0.002 | 0.004 | 0.007 | 0.011 | 0.012 | 0.012 |
| Bromazepam (90) | 0.000 | 0.000 | 0.000 | 0.000 | 0.001 | 0.001 | 0.003 | 0.006 | 0.009 | 0.010 | 0.011 |
| Clobazam (30) | 0.000 | 0.000 | 0.000 | 0.001 | 0.001 | 0.001 | 0.001 | 0.001 | 0.001 | 0.001 | 0.001 |
| Clobazam (90) | 0.000 | 0.000 | 0.000 | 0.001 | 0.001 | 0.001 | 0.001 | 0.001 | 0.001 | 0.001 | 0.001 |
| Prazepam (30) | 0.000 | 0.000 | 0.000 | 0.000 | 0.000 | 0.000 | 0.001 | 0.001 | 0.002 | 0.002 | 0.002 |
| Prazepam (90) | 0.000 | 0.000 | 0.000 | 0.000 | 0.000 | 0.000 | 0.000 | 0.001 | 0.002 | 0.002 | 0.002 |
| Alprazolam (30) | 0.000 | 0.000 | 0.000 | 0.004 | 0.018 | 0.022 | 0.027 | 0.035 | 0.037 | 0.037 | 0.048 |
| Alprazolam (90) | 0.000 | 0.000 | 0.000 | 0.002 | 0.011 | 0.014 | 0.019 | 0.026 | 0.029 | 0.029 | 0.038 |
| Flurazepam (30) | 0.000 | 0.000 | 0.000 | 0.001 | 0.003 | 0.007 | 0.010 | 0.012 | 0.012 | 0.011 | 0.009 |
| Flurazepam (90) | 0.000 | 0.000 | 0.000 | 0.000 | 0.002 | 0.006 | 0.009 | 0.011 | 0.011 | 0.009 | 0.008 |
| Nitrazepam (30) | 0.000 | 0.000 | 0.000 | 0.000 | 0.000 | 0.000 | 0.001 | 0.001 | 0.002 | 0.002 | 0.004 |
| Nitrazepam (90) | 0.000 | 0.000 | 0.000 | 0.000 | 0.000 | 0.000 | 0.001 | 0.001 | 0.001 | 0.002 | 0.003 |
| Triazolam (30) | 0.000 | 0.000 | 0.000 | 0.000 | 0.002 | 0.003 | 0.003 | 0.006 | 0.007 | 0.008 | 0.012 |
| Triazolam (90) | 0.000 | 0.000 | 0.000 | 0.000 | 0.002 | 0.002 | 0.003 | 0.005 | 0.007 | 0.007 | 0.011 |
| Lormetazepam (30) | 0.000 | 0.000 | 0.000 | 0.000 | 0.000 | 0.000 | 0.001 | 0.002 | 0.004 | 0.005 | 0.009 |
| Lormetazepam (90) | 0.000 | 0.000 | 0.000 | 0.000 | 0.000 | 0.000 | 0.001 | 0.002 | 0.003 | 0.004 | 0.009 |
| Temazepam (30) | 0.000 | 0.000 | 0.000 | 0.000 | 0.001 | 0.002 | 0.003 | 0.006 | 0.009 | 0.010 | 0.019 |
| Temazepam (90) | 0.000 | 0.000 | 0.000 | 0.000 | 0.001 | 0.001 | 0.002 | 0.005 | 0.008 | 0.009 | 0.017 |
| Zopiclone (30) | 0.000 | 0.000 | 0.000 | 0.004 | 0.018 | 0.027 | 0.038 | 0.050 | 0.054 | 0.055 | 0.080 |
| Zopiclone (90) | 0.000 | 0.000 | 0.000 | 0.002 | 0.013 | 0.021 | 0.030 | 0.042 | 0.046 | 0.047 | 0.070 |
| Zolpidem (30) | 0.000 | 0.000 | 0.000 | 0.002 | 0.010 | 0.016 | 0.027 | 0.040 | 0.047 | 0.050 | 0.067 |
| Zolpidem (90) | 0.000 | 0.000 | 0.000 | 0.001 | 0.006 | 0.012 | 0.021 | 0.032 | 0.040 | 0.042 | 0.058 |
| Antidepressant (30) | 0.000 | 0.000 | 0.000 | 0.003 | 0.011 | 0.013 | 0.015 | 0.017 | 0.017 | 0.019 | 0.038 |
| Antidepressant (90) | 0.000 | 0.000 | 0.000 | 0.002 | 0.006 | 0.008 | 0.011 | 0.013 | 0.013 | 0.016 | 0.032 |
| Antihistamines (30) | 0.000 | 0.000 | 0.000 | 0.002 | 0.004 | 0.004 | 0.004 | 0.004 | 0.003 | 0.002 | 0.002 |
| Antihistamines (90) | 0.000 | 0.000 | 0.000 | 0.001 | 0.002 | 0.003 | 0.003 | 0.003 | 0.002 | 0.002 | 0.002 |
| Antipsychotics (30) | 0.000 | 0.000 | 0.001 | 0.007 | 0.016 | 0.015 | 0.017 | 0.018 | 0.016 | 0.018 | 0.046 |
| Antipsychotics (90) | 0.000 | 0.000 | 0.000 | 0.005 | 0.011 | 0.012 | 0.014 | 0.015 | 0.014 | 0.016 | 0.041 |
| 2018 |  |  |  |  |  |  |  |  |  |  |  |
| Sedative (30) | 0.001 | 0.001 | 0.003 | 0.030 | 0.100 | 0.122 | 0.145 | 0.186 | 0.207 | 0.206 | 0.291 |
| Sedative (90) | 0.000 | 0.001 | 0.002 | 0.019 | 0.070 | 0.091 | 0.115 | 0.154 | 0.178 | 0.178 | 0.261 |
| Benzodiazepines and z-drugs (30) | 0.001 | 0.001 | 0.002 | 0.020 | 0.078 | 0.104 | 0.126 | 0.168 | 0.192 | 0.190 | 0.256 |
| Benzodiazepines and z-drugs after (90) | 0.000 | 0.001 | 0.001 | 0.011 | 0.053 | 0.075 | 0.098 | 0.138 | 0.164 | 0.163 | 0.226 |
| Long-acting benzodiazepines (30) | 0.001 | 0.001 | 0.001 | 0.009 | 0.038 | 0.050 | 0.052 | 0.057 | 0.057 | 0.049 | 0.051 |
| Long-acting benzodiazepines (90) | 0.000 | 0.000 | 0.001 | 0.005 | 0.026 | 0.035 | 0.038 | 0.044 | 0.045 | 0.040 | 0.042 |
| Clonazepam (30) | 0.000 | 0.000 | 0.000 | 0.002 | 0.006 | 0.006 | 0.007 | 0.008 | 0.007 | 0.007 | 0.007 |
| Clonazepam (90) | 0.000 | 0.000 | 0.000 | 0.001 | 0.005 | 0.005 | 0.006 | 0.007 | 0.006 | 0.006 | 0.006 |
| Diazepam (30) | 0.000 | 0.000 | 0.001 | 0.005 | 0.028 | 0.036 | 0.034 | 0.034 | 0.033 | 0.029 | 0.029 |
| Diazepam (90) | 0.000 | 0.000 | 0.000 | 0.003 | 0.018 | 0.025 | 0.024 | 0.025 | 0.026 | 0.022 | 0.023 |
| Chlordiazepoxide (30) | 0.000 | 0.000 | 0.000 | 0.000 | 0.003 | 0.004 | 0.004 | 0.004 | 0.003 | 0.002 | 0.002 |
| Chlordiazepoxide (90) | 0.000 | 0.000 | 0.000 | 0.000 | 0.001 | 0.002 | 0.002 | 0.002 | 0.002 | 0.001 | 0.001 |
| Clorazepate (30) | 0.000 | 0.000 | 0.000 | 0.000 | 0.000 | 0.000 | 0.000 | 0.000 | 0.000 | 0.000 | 0.000 |
| Clorazepate (90) | 0.000 | 0.000 | 0.000 | 0.000 | 0.000 | 0.000 | 0.000 | 0.000 | 0.000 | 0.000 | 0.000 |
| Lorazepam (30) | 0.000 | 0.000 | 0.000 | 0.001 | 0.002 | 0.002 | 0.003 | 0.004 | 0.004 | 0.004 | 0.006 |
| Lorazepam (90) | 0.000 | 0.000 | 0.000 | 0.001 | 0.002 | 0.001 | 0.002 | 0.003 | 0.004 | 0.003 | 0.005 |
| Bromazepam (30) | 0.000 | 0.000 | 0.000 | 0.000 | 0.001 | 0.002 | 0.003 | 0.007 | 0.010 | 0.011 | 0.012 |
| Bromazepam (90) | 0.000 | 0.000 | 0.000 | 0.000 | 0.001 | 0.001 | 0.003 | 0.005 | 0.009 | 0.010 | 0.011 |
| Clobazam (30) | 0.000 | 0.000 | 0.000 | 0.001 | 0.001 | 0.001 | 0.001 | 0.001 | 0.001 | 0.001 | 0.001 |
| Clobazam (90) | 0.000 | 0.000 | 0.000 | 0.001 | 0.001 | 0.001 | 0.001 | 0.001 | 0.001 | 0.001 | 0.001 |
| Prazepam (30) | 0.000 | 0.000 | 0.000 | 0.000 | 0.000 | 0.000 | 0.001 | 0.001 | 0.002 | 0.002 | 0.002 |
| Prazepam (90) | 0.000 | 0.000 | 0.000 | 0.000 | 0.000 | 0.000 | 0.000 | 0.001 | 0.002 | 0.002 | 0.002 |
| Alprazolam (30) | 0.000 | 0.000 | 0.000 | 0.005 | 0.019 | 0.023 | 0.026 | 0.033 | 0.036 | 0.035 | 0.047 |
| Alprazolam (90) | 0.000 | 0.000 | 0.000 | 0.002 | 0.012 | 0.015 | 0.019 | 0.026 | 0.028 | 0.027 | 0.037 |
| Flurazepam (30) | 0.000 | 0.000 | 0.000 | 0.001 | 0.003 | 0.006 | 0.009 | 0.011 | 0.012 | 0.010 | 0.008 |
| Flurazepam (90) | 0.000 | 0.000 | 0.000 | 0.000 | 0.002 | 0.005 | 0.008 | 0.010 | 0.011 | 0.009 | 0.008 |
| Nitrazepam (30) | 0.000 | 0.000 | 0.000 | 0.000 | 0.000 | 0.000 | 0.001 | 0.001 | 0.002 | 0.002 | 0.003 |
| Nitrazepam (90) | 0.000 | 0.000 | 0.000 | 0.000 | 0.000 | 0.000 | 0.000 | 0.001 | 0.001 | 0.001 | 0.003 |
| Triazolam (30) | 0.000 | 0.000 | 0.000 | 0.000 | 0.002 | 0.002 | 0.003 | 0.006 | 0.007 | 0.007 | 0.012 |
| Triazolam (90) | 0.000 | 0.000 | 0.000 | 0.000 | 0.002 | 0.002 | 0.003 | 0.005 | 0.006 | 0.006 | 0.011 |
| Lormetazepam (30) | 0.000 | 0.000 | 0.000 | 0.000 | 0.000 | 0.000 | 0.001 | 0.002 | 0.004 | 0.004 | 0.009 |
| Lormetazepam (90) | 0.000 | 0.000 | 0.000 | 0.000 | 0.000 | 0.000 | 0.001 | 0.002 | 0.003 | 0.004 | 0.008 |
| Temazepam (30) | 0.000 | 0.000 | 0.000 | 0.000 | 0.001 | 0.002 | 0.003 | 0.006 | 0.008 | 0.009 | 0.017 |
| Temazepam (90) | 0.000 | 0.000 | 0.000 | 0.000 | 0.001 | 0.001 | 0.002 | 0.005 | 0.007 | 0.008 | 0.015 |
| Zopiclone (30) | 0.000 | 0.000 | 0.000 | 0.004 | 0.018 | 0.027 | 0.036 | 0.049 | 0.054 | 0.054 | 0.079 |
| Zopiclone (90) | 0.000 | 0.000 | 0.000 | 0.002 | 0.014 | 0.021 | 0.029 | 0.041 | 0.046 | 0.046 | 0.069 |
| Zolpidem (30) | 0.000 | 0.000 | 0.000 | 0.002 | 0.010 | 0.016 | 0.025 | 0.039 | 0.048 | 0.049 | 0.066 |
| Zolpidem (90) | 0.000 | 0.000 | 0.000 | 0.001 | 0.006 | 0.012 | 0.020 | 0.031 | 0.041 | 0.041 | 0.058 |
| Antidepressant (30) | 0.000 | 0.000 | 0.000 | 0.004 | 0.013 | 0.014 | 0.016 | 0.019 | 0.019 | 0.020 | 0.040 |
| Antidepressant (90) | 0.000 | 0.000 | 0.000 | 0.002 | 0.008 | 0.010 | 0.012 | 0.014 | 0.015 | 0.016 | 0.034 |
| Antihistamines (30) | 0.000 | 0.000 | 0.000 | 0.002 | 0.005 | 0.005 | 0.005 | 0.005 | 0.003 | 0.003 | 0.003 |
| Antihistamines (90) | 0.000 | 0.000 | 0.000 | 0.001 | 0.003 | 0.003 | 0.004 | 0.004 | 0.002 | 0.002 | 0.002 |
| Antipsychotics (30) | 0.000 | 0.000 | 0.001 | 0.009 | 0.020 | 0.017 | 0.019 | 0.019 | 0.018 | 0.019 | 0.048 |
| Antipsychotics (90) | 0.000 | 0.000 | 0.001 | 0.006 | 0.014 | 0.013 | 0.015 | 0.016 | 0.015 | 0.017 | 0.042 |
| 2019 |  |  |  |  |  |  |  |  |  |  |  |
| Sedative (30) | 0.001 | 0.001 | 0.002 | 0.032 | 0.103 | 0.123 | 0.146 | 0.184 | 0.206 | 0.205 | 0.285 |
| Sedative (90) | 0.000 | 0.001 | 0.001 | 0.020 | 0.073 | 0.092 | 0.115 | 0.154 | 0.178 | 0.177 | 0.257 |
| Benzodiazepines and z-drugs (30) | 0.000 | 0.001 | 0.002 | 0.019 | 0.078 | 0.102 | 0.124 | 0.164 | 0.189 | 0.187 | 0.247 |
| Benzodiazepines and z-drugs after (90) | 0.000 | 0.001 | 0.001 | 0.011 | 0.053 | 0.074 | 0.096 | 0.136 | 0.162 | 0.160 | 0.219 |
| Long-acting benzodiazepines (30) | 0.000 | 0.001 | 0.001 | 0.008 | 0.038 | 0.050 | 0.052 | 0.056 | 0.056 | 0.048 | 0.050 |
| Long-acting benzodiazepines (90) | 0.000 | 0.001 | 0.001 | 0.005 | 0.026 | 0.035 | 0.038 | 0.043 | 0.045 | 0.039 | 0.041 |
| Clonazepam (30) | 0.000 | 0.000 | 0.000 | 0.002 | 0.006 | 0.006 | 0.007 | 0.008 | 0.007 | 0.007 | 0.007 |
| Clonazepam (90) | 0.000 | 0.000 | 0.000 | 0.001 | 0.005 | 0.005 | 0.006 | 0.007 | 0.006 | 0.006 | 0.006 |
| Diazepam (30) | 0.000 | 0.000 | 0.001 | 0.005 | 0.028 | 0.036 | 0.034 | 0.034 | 0.034 | 0.028 | 0.028 |
| Diazepam (90) | 0.000 | 0.000 | 0.000 | 0.003 | 0.018 | 0.025 | 0.025 | 0.025 | 0.025 | 0.021 | 0.022 |
| Chlordiazepoxide (30) | 0.000 | 0.000 | 0.000 | 0.000 | 0.003 | 0.005 | 0.005 | 0.004 | 0.003 | 0.002 | 0.001 |
| Chlordiazepoxide (90) | 0.000 | 0.000 | 0.000 | 0.000 | 0.001 | 0.002 | 0.002 | 0.002 | 0.001 | 0.001 | 0.001 |
| Clorazepate (30) | 0.000 | 0.000 | 0.000 | 0.000 | 0.000 | 0.000 | 0.000 | 0.000 | 0.000 | 0.000 | 0.000 |
| Clorazepate (90) | 0.000 | 0.000 | 0.000 | 0.000 | 0.000 | 0.000 | 0.000 | 0.000 | 0.000 | 0.000 | 0.000 |
| Lorazepam (30) | 0.000 | 0.000 | 0.000 | 0.001 | 0.002 | 0.003 | 0.003 | 0.004 | 0.005 | 0.004 | 0.006 |
| Lorazepam (90) | 0.000 | 0.000 | 0.000 | 0.001 | 0.002 | 0.002 | 0.002 | 0.003 | 0.004 | 0.003 | 0.005 |
| Bromazepam (30) | 0.000 | 0.000 | 0.000 | 0.000 | 0.001 | 0.002 | 0.003 | 0.006 | 0.010 | 0.011 | 0.012 |
| Bromazepam (90) | 0.000 | 0.000 | 0.000 | 0.000 | 0.001 | 0.001 | 0.003 | 0.005 | 0.009 | 0.010 | 0.010 |
| Clobazam (30) | 0.000 | 0.000 | 0.000 | 0.001 | 0.001 | 0.001 | 0.001 | 0.001 | 0.001 | 0.001 | 0.001 |
| Clobazam (90) | 0.000 | 0.000 | 0.000 | 0.001 | 0.001 | 0.001 | 0.001 | 0.001 | 0.001 | 0.001 | 0.001 |
| Prazepam (30) | 0.000 | 0.000 | 0.000 | 0.000 | 0.000 | 0.000 | 0.000 | 0.001 | 0.002 | 0.002 | 0.002 |
| Prazepam (90) | 0.000 | 0.000 | 0.000 | 0.000 | 0.000 | 0.000 | 0.000 | 0.001 | 0.002 | 0.002 | 0.002 |
| Alprazolam (30) | 0.000 | 0.000 | 0.000 | 0.005 | 0.020 | 0.022 | 0.027 | 0.032 | 0.036 | 0.035 | 0.045 |
| Alprazolam (90) | 0.000 | 0.000 | 0.000 | 0.002 | 0.012 | 0.015 | 0.019 | 0.025 | 0.029 | 0.027 | 0.036 |
| Flurazepam (30) | 0.000 | 0.000 | 0.000 | 0.000 | 0.003 | 0.006 | 0.008 | 0.011 | 0.011 | 0.009 | 0.008 |
| Flurazepam (90) | 0.000 | 0.000 | 0.000 | 0.000 | 0.002 | 0.005 | 0.007 | 0.010 | 0.010 | 0.008 | 0.007 |
| Nitrazepam (30) | 0.000 | 0.000 | 0.000 | 0.000 | 0.000 | 0.000 | 0.001 | 0.001 | 0.001 | 0.001 | 0.003 |
| Nitrazepam (90) | 0.000 | 0.000 | 0.000 | 0.000 | 0.000 | 0.000 | 0.000 | 0.001 | 0.001 | 0.001 | 0.003 |
| Triazolam (30) | 0.000 | 0.000 | 0.000 | 0.000 | 0.002 | 0.002 | 0.003 | 0.006 | 0.007 | 0.007 | 0.011 |
| Triazolam (90) | 0.000 | 0.000 | 0.000 | 0.000 | 0.001 | 0.002 | 0.003 | 0.005 | 0.006 | 0.006 | 0.010 |
| Lormetazepam (30) | 0.000 | 0.000 | 0.000 | 0.000 | 0.000 | 0.000 | 0.001 | 0.002 | 0.003 | 0.003 | 0.007 |
| Lormetazepam (90) | 0.000 | 0.000 | 0.000 | 0.000 | 0.000 | 0.000 | 0.001 | 0.001 | 0.002 | 0.003 | 0.006 |
| Temazepam (30) | 0.000 | 0.000 | 0.000 | 0.000 | 0.001 | 0.001 | 0.002 | 0.005 | 0.007 | 0.008 | 0.016 |
| Temazepam (90) | 0.000 | 0.000 | 0.000 | 0.000 | 0.001 | 0.001 | 0.002 | 0.004 | 0.006 | 0.007 | 0.014 |
| Zopiclone (30) | 0.000 | 0.000 | 0.000 | 0.004 | 0.018 | 0.026 | 0.036 | 0.049 | 0.055 | 0.054 | 0.078 |
| Zopiclone (90) | 0.000 | 0.000 | 0.000 | 0.002 | 0.013 | 0.021 | 0.029 | 0.041 | 0.047 | 0.046 | 0.069 |
| Zolpidem (30) | 0.000 | 0.000 | 0.000 | 0.002 | 0.010 | 0.016 | 0.025 | 0.039 | 0.048 | 0.050 | 0.067 |
| Zolpidem (90) | 0.000 | 0.000 | 0.000 | 0.001 | 0.006 | 0.011 | 0.019 | 0.032 | 0.040 | 0.042 | 0.058 |
| Antidepressant (30) | 0.000 | 0.000 | 0.000 | 0.004 | 0.014 | 0.015 | 0.017 | 0.020 | 0.021 | 0.022 | 0.042 |
| Antidepressant (90) | 0.000 | 0.000 | 0.000 | 0.002 | 0.009 | 0.010 | 0.013 | 0.015 | 0.017 | 0.017 | 0.036 |
| Antihistamines (30) | 0.000 | 0.000 | 0.000 | 0.003 | 0.007 | 0.007 | 0.007 | 0.006 | 0.004 | 0.003 | 0.003 |
| Antihistamines (90) | 0.000 | 0.000 | 0.000 | 0.002 | 0.004 | 0.004 | 0.005 | 0.005 | 0.003 | 0.002 | 0.002 |
| Antipsychotics (30) | 0.000 | 0.000 | 0.001 | 0.010 | 0.023 | 0.020 | 0.020 | 0.021 | 0.019 | 0.020 | 0.049 |
| Antipsychotics (90) | 0.000 | 0.000 | 0.001 | 0.007 | 0.016 | 0.015 | 0.016 | 0.018 | 0.017 | 0.017 | 0.043 |
| 2020 |  |  |  |  |  |  |  |  |  |  |  |
| Sedative (30) | 0.001 | 0.001 | 0.002 | 0.028 | 0.099 | 0.119 | 0.144 | 0.180 | 0.201 | 0.200 | 0.280 |
| Sedative (90) | 0.000 | 0.001 | 0.001 | 0.019 | 0.072 | 0.091 | 0.115 | 0.152 | 0.173 | 0.175 | 0.254 |
| Benzodiazepines and z-drugs (30) | 0.001 | 0.001 | 0.001 | 0.016 | 0.072 | 0.096 | 0.120 | 0.157 | 0.181 | 0.181 | 0.240 |
| Benzodiazepines and z-drugs after (90) | 0.000 | 0.001 | 0.001 | 0.009 | 0.049 | 0.070 | 0.094 | 0.131 | 0.155 | 0.157 | 0.212 |
| Long-acting benzodiazepines (30) | 0.001 | 0.001 | 0.001 | 0.007 | 0.035 | 0.047 | 0.051 | 0.053 | 0.053 | 0.047 | 0.049 |
| Long-acting benzodiazepines (90) | 0.000 | 0.001 | 0.001 | 0.004 | 0.024 | 0.034 | 0.038 | 0.042 | 0.042 | 0.038 | 0.040 |
| Clonazepam (30) | 0.000 | 0.000 | 0.000 | 0.002 | 0.005 | 0.006 | 0.007 | 0.008 | 0.008 | 0.007 | 0.008 |
| Clonazepam (90) | 0.000 | 0.000 | 0.000 | 0.001 | 0.004 | 0.005 | 0.006 | 0.007 | 0.007 | 0.006 | 0.007 |
| Diazepam (30) | 0.000 | 0.000 | 0.000 | 0.004 | 0.026 | 0.035 | 0.034 | 0.033 | 0.032 | 0.028 | 0.028 |
| Diazepam (90) | 0.000 | 0.000 | 0.000 | 0.002 | 0.017 | 0.024 | 0.025 | 0.025 | 0.024 | 0.021 | 0.022 |
| Chlordiazepoxide (30) | 0.000 | 0.000 | 0.000 | 0.000 | 0.003 | 0.004 | 0.005 | 0.004 | 0.002 | 0.002 | 0.001 |
| Chlordiazepoxide (90) | 0.000 | 0.000 | 0.000 | 0.000 | 0.001 | 0.002 | 0.002 | 0.002 | 0.001 | 0.001 | 0.001 |
| Clorazepate (30) | 0.000 | 0.000 | 0.000 | 0.000 | 0.000 | 0.000 | 0.000 | 0.000 | 0.000 | 0.000 | 0.000 |
| Clorazepate (90) | 0.000 | 0.000 | 0.000 | 0.000 | 0.000 | 0.000 | 0.000 | 0.000 | 0.000 | 0.000 | 0.000 |
| Lorazepam (30) | 0.000 | 0.000 | 0.000 | 0.001 | 0.002 | 0.002 | 0.003 | 0.004 | 0.004 | 0.004 | 0.006 |
| Lorazepam (90) | 0.000 | 0.000 | 0.000 | 0.001 | 0.002 | 0.002 | 0.002 | 0.003 | 0.003 | 0.003 | 0.005 |
| Bromazepam (30) | 0.000 | 0.000 | 0.000 | 0.000 | 0.001 | 0.002 | 0.003 | 0.006 | 0.009 | 0.010 | 0.011 |
| Bromazepam (90) | 0.000 | 0.000 | 0.000 | 0.000 | 0.001 | 0.001 | 0.002 | 0.005 | 0.008 | 0.009 | 0.010 |
| Clobazam (30) | 0.000 | 0.000 | 0.000 | 0.001 | 0.001 | 0.001 | 0.001 | 0.001 | 0.001 | 0.001 | 0.001 |
| Clobazam (90) | 0.000 | 0.000 | 0.000 | 0.001 | 0.001 | 0.001 | 0.001 | 0.001 | 0.001 | 0.001 | 0.001 |
| Prazepam (30) | 0.000 | 0.000 | 0.000 | 0.000 | 0.000 | 0.000 | 0.001 | 0.001 | 0.002 | 0.002 | 0.002 |
| Prazepam (90) | 0.000 | 0.000 | 0.000 | 0.000 | 0.000 | 0.000 | 0.000 | 0.001 | 0.002 | 0.002 | 0.002 |
| Alprazolam (30) | 0.000 | 0.000 | 0.000 | 0.004 | 0.018 | 0.022 | 0.025 | 0.031 | 0.036 | 0.035 | 0.044 |
| Alprazolam (90) | 0.000 | 0.000 | 0.000 | 0.002 | 0.012 | 0.015 | 0.019 | 0.024 | 0.028 | 0.027 | 0.035 |
| Flurazepam (30) | 0.000 | 0.000 | 0.000 | 0.000 | 0.002 | 0.005 | 0.008 | 0.010 | 0.010 | 0.008 | 0.008 |
| Flurazepam (90) | 0.000 | 0.000 | 0.000 | 0.000 | 0.002 | 0.004 | 0.007 | 0.009 | 0.009 | 0.008 | 0.007 |
| Nitrazepam (30) | 0.000 | 0.000 | 0.000 | 0.000 | 0.000 | 0.000 | 0.000 | 0.001 | 0.001 | 0.001 | 0.003 |
| Nitrazepam (90) | 0.000 | 0.000 | 0.000 | 0.000 | 0.000 | 0.000 | 0.000 | 0.001 | 0.001 | 0.001 | 0.003 |
| Triazolam (30) | 0.000 | 0.000 | 0.000 | 0.000 | 0.001 | 0.002 | 0.003 | 0.005 | 0.006 | 0.007 | 0.011 |
| Triazolam (90) | 0.000 | 0.000 | 0.000 | 0.000 | 0.001 | 0.002 | 0.003 | 0.005 | 0.005 | 0.006 | 0.010 |
| Lormetazepam (30) | 0.000 | 0.000 | 0.000 | 0.000 | 0.000 | 0.000 | 0.000 | 0.000 | 0.000 | 0.000 | 0.001 |
| Lormetazepam (90) | 0.000 | 0.000 | 0.000 | 0.000 | 0.000 | 0.000 | 0.000 | 0.000 | 0.000 | 0.000 | 0.001 |
| Temazepam (30) | 0.000 | 0.000 | 0.000 | 0.000 | 0.001 | 0.001 | 0.002 | 0.005 | 0.006 | 0.008 | 0.015 |
| Temazepam (90) | 0.000 | 0.000 | 0.000 | 0.000 | 0.001 | 0.001 | 0.002 | 0.004 | 0.005 | 0.007 | 0.013 |
| Zopiclone (30) | 0.000 | 0.000 | 0.000 | 0.003 | 0.017 | 0.025 | 0.035 | 0.047 | 0.054 | 0.053 | 0.077 |
| Zopiclone (90) | 0.000 | 0.000 | 0.000 | 0.002 | 0.012 | 0.020 | 0.029 | 0.040 | 0.046 | 0.045 | 0.068 |
| Zolpidem (30) | 0.000 | 0.000 | 0.000 | 0.002 | 0.009 | 0.015 | 0.024 | 0.038 | 0.047 | 0.049 | 0.067 |
| Zolpidem (90) | 0.000 | 0.000 | 0.000 | 0.001 | 0.006 | 0.011 | 0.019 | 0.032 | 0.040 | 0.042 | 0.058 |
| Antidepressant (30) | 0.000 | 0.000 | 0.000 | 0.004 | 0.015 | 0.016 | 0.018 | 0.021 | 0.022 | 0.023 | 0.045 |
| Antidepressant (90) | 0.000 | 0.000 | 0.000 | 0.002 | 0.010 | 0.011 | 0.014 | 0.017 | 0.018 | 0.019 | 0.038 |
| Antihistamines (30) | 0.000 | 0.000 | 0.000 | 0.003 | 0.008 | 0.009 | 0.009 | 0.008 | 0.006 | 0.004 | 0.004 |
| Antihistamines (90) | 0.000 | 0.000 | 0.000 | 0.002 | 0.005 | 0.006 | 0.006 | 0.006 | 0.004 | 0.003 | 0.003 |
| Antipsychotics (30) | 0.000 | 0.000 | 0.001 | 0.010 | 0.024 | 0.022 | 0.022 | 0.022 | 0.020 | 0.020 | 0.051 |
| Antipsychotics (90) | 0.000 | 0.000 | 0.001 | 0.007 | 0.018 | 0.017 | 0.018 | 0.019 | 0.017 | 0.018 | 0.045 |
| 2021 |  |  |  |  |  |  |  |  |  |  |  |
| Sedative (30) | 0.001 | 0.001 | 0.002 | 0.031 | 0.106 | 0.125 | 0.148 | 0.180 | 0.202 | 0.198 | 0.276 |
| Sedative (90) | 0.000 | 0.001 | 0.001 | 0.021 | 0.078 | 0.096 | 0.119 | 0.153 | 0.175 | 0.173 | 0.251 |
| Benzodiazepines and z-drugs (30) | 0.001 | 0.001 | 0.001 | 0.017 | 0.073 | 0.097 | 0.120 | 0.154 | 0.180 | 0.177 | 0.232 |
| Benzodiazepines and z-drugs after (90) | 0.000 | 0.001 | 0.001 | 0.010 | 0.050 | 0.071 | 0.094 | 0.129 | 0.154 | 0.153 | 0.207 |
| Long-acting benzodiazepines (30) | 0.001 | 0.001 | 0.001 | 0.008 | 0.037 | 0.048 | 0.052 | 0.053 | 0.053 | 0.045 | 0.047 |
| Long-acting benzodiazepines (90) | 0.000 | 0.001 | 0.001 | 0.005 | 0.025 | 0.034 | 0.039 | 0.041 | 0.042 | 0.037 | 0.039 |
| Clonazepam (30) | 0.000 | 0.000 | 0.000 | 0.002 | 0.006 | 0.006 | 0.007 | 0.008 | 0.008 | 0.007 | 0.008 |
| Clonazepam (90) | 0.000 | 0.000 | 0.000 | 0.001 | 0.004 | 0.005 | 0.006 | 0.007 | 0.007 | 0.006 | 0.007 |
| Diazepam (30) | 0.000 | 0.000 | 0.000 | 0.005 | 0.027 | 0.035 | 0.035 | 0.034 | 0.032 | 0.028 | 0.027 |
| Diazepam (90) | 0.000 | 0.000 | 0.000 | 0.003 | 0.018 | 0.025 | 0.026 | 0.025 | 0.024 | 0.021 | 0.021 |
| Chlordiazepoxide (30) | 0.000 | 0.000 | 0.000 | 0.000 | 0.003 | 0.004 | 0.005 | 0.004 | 0.002 | 0.002 | 0.001 |
| Chlordiazepoxide (90) | 0.000 | 0.000 | 0.000 | 0.000 | 0.001 | 0.002 | 0.002 | 0.001 | 0.001 | 0.001 | 0.001 |
| Clorazepate (30) | 0.000 | 0.000 | 0.000 | 0.000 | 0.000 | 0.000 | 0.000 | 0.000 | 0.000 | 0.000 | 0.000 |
| Clorazepate (90) | 0.000 | 0.000 | 0.000 | 0.000 | 0.000 | 0.000 | 0.000 | 0.000 | 0.000 | 0.000 | 0.000 |
| Lorazepam (30) | 0.000 | 0.000 | 0.000 | 0.001 | 0.003 | 0.002 | 0.003 | 0.004 | 0.004 | 0.004 | 0.006 |
| Lorazepam (90) | 0.000 | 0.000 | 0.000 | 0.001 | 0.002 | 0.002 | 0.002 | 0.003 | 0.003 | 0.003 | 0.005 |
| Bromazepam (30) | 0.000 | 0.000 | 0.000 | 0.000 | 0.001 | 0.002 | 0.003 | 0.005 | 0.008 | 0.010 | 0.011 |
| Bromazepam (90) | 0.000 | 0.000 | 0.000 | 0.000 | 0.001 | 0.001 | 0.002 | 0.004 | 0.007 | 0.008 | 0.010 |
| Clobazam (30) | 0.000 | 0.000 | 0.000 | 0.001 | 0.001 | 0.001 | 0.001 | 0.001 | 0.001 | 0.001 | 0.001 |
| Clobazam (90) | 0.000 | 0.000 | 0.000 | 0.000 | 0.001 | 0.001 | 0.001 | 0.001 | 0.001 | 0.001 | 0.001 |
| Prazepam (30) | 0.000 | 0.000 | 0.000 | 0.000 | 0.000 | 0.000 | 0.000 | 0.001 | 0.001 | 0.002 | 0.002 |
| Prazepam (90) | 0.000 | 0.000 | 0.000 | 0.000 | 0.000 | 0.000 | 0.000 | 0.001 | 0.001 | 0.002 | 0.001 |
| Alprazolam (30) | 0.000 | 0.000 | 0.000 | 0.004 | 0.018 | 0.022 | 0.025 | 0.031 | 0.035 | 0.033 | 0.042 |
| Alprazolam (90) | 0.000 | 0.000 | 0.000 | 0.002 | 0.011 | 0.015 | 0.019 | 0.024 | 0.028 | 0.027 | 0.034 |
| Flurazepam (30) | 0.000 | 0.000 | 0.000 | 0.001 | 0.002 | 0.004 | 0.007 | 0.009 | 0.010 | 0.008 | 0.007 |
| Flurazepam (90) | 0.000 | 0.000 | 0.000 | 0.000 | 0.002 | 0.004 | 0.006 | 0.008 | 0.009 | 0.007 | 0.007 |
| Nitrazepam (30) | 0.000 | 0.000 | 0.000 | 0.000 | 0.000 | 0.000 | 0.000 | 0.001 | 0.001 | 0.001 | 0.002 |
| Nitrazepam (90) | 0.000 | 0.000 | 0.000 | 0.000 | 0.000 | 0.000 | 0.000 | 0.001 | 0.001 | 0.001 | 0.002 |
| Triazolam (30) | 0.000 | 0.000 | 0.000 | 0.000 | 0.001 | 0.002 | 0.003 | 0.005 | 0.006 | 0.007 | 0.010 |
| Triazolam (90) | 0.000 | 0.000 | 0.000 | 0.000 | 0.001 | 0.002 | 0.003 | 0.005 | 0.005 | 0.006 | 0.009 |
| Lormetazepam (30) | 0.000 | 0.000 | 0.000 | 0.000 | 0.000 | 0.000 | 0.000 | 0.000 | 0.000 | 0.000 | 0.000 |
| Lormetazepam (90) | 0.000 | 0.000 | 0.000 | 0.000 | 0.000 | 0.000 | 0.000 | 0.000 | 0.000 | 0.000 | 0.000 |
| Temazepam (30) | 0.000 | 0.000 | 0.000 | 0.000 | 0.001 | 0.001 | 0.002 | 0.004 | 0.006 | 0.007 | 0.013 |
| Temazepam (90) | 0.000 | 0.000 | 0.000 | 0.000 | 0.001 | 0.001 | 0.002 | 0.004 | 0.005 | 0.006 | 0.012 |
| Zopiclone (30) | 0.000 | 0.000 | 0.000 | 0.003 | 0.016 | 0.025 | 0.034 | 0.047 | 0.053 | 0.052 | 0.076 |
| Zopiclone (90) | 0.000 | 0.000 | 0.000 | 0.002 | 0.012 | 0.020 | 0.028 | 0.040 | 0.046 | 0.045 | 0.067 |
| Zolpidem (30) | 0.000 | 0.000 | 0.000 | 0.002 | 0.009 | 0.015 | 0.024 | 0.037 | 0.047 | 0.049 | 0.065 |
| Zolpidem (90) | 0.000 | 0.000 | 0.000 | 0.001 | 0.006 | 0.011 | 0.019 | 0.031 | 0.041 | 0.042 | 0.058 |
| Antidepressant (30) | 0.000 | 0.000 | 0.000 | 0.004 | 0.016 | 0.017 | 0.020 | 0.023 | 0.024 | 0.025 | 0.048 |
| Antidepressant (90) | 0.000 | 0.000 | 0.000 | 0.003 | 0.011 | 0.012 | 0.015 | 0.018 | 0.020 | 0.021 | 0.041 |
| Antihistamines (30) | 0.000 | 0.000 | 0.000 | 0.004 | 0.011 | 0.011 | 0.011 | 0.010 | 0.007 | 0.005 | 0.005 |
| Antihistamines (90) | 0.000 | 0.000 | 0.000 | 0.003 | 0.007 | 0.008 | 0.008 | 0.007 | 0.006 | 0.004 | 0.004 |
| Antipsychotics (30) | 0.000 | 0.000 | 0.001 | 0.011 | 0.027 | 0.024 | 0.024 | 0.024 | 0.022 | 0.021 | 0.052 |
| Antipsychotics (90) | 0.000 | 0.000 | 0.001 | 0.008 | 0.021 | 0.019 | 0.020 | 0.021 | 0.019 | 0.018 | 0.046 |
| 2022 |  |  |  |  |  |  |  |  |  |  |  |
| Sedative (30) | 0.001 | 0.001 | 0.002 | 0.026 | 0.095 | 0.120 | 0.142 | 0.173 | 0.191 | 0.189 | 0.268 |
| Sedative (90) | 0.000 | 0.001 | 0.001 | 0.018 | 0.070 | 0.092 | 0.115 | 0.146 | 0.166 | 0.165 | 0.242 |
| Benzodiazepines and z-drugs (30) | 0.001 | 0.001 | 0.001 | 0.015 | 0.064 | 0.091 | 0.114 | 0.145 | 0.168 | 0.168 | 0.222 |
| Benzodiazepines and z-drugs after (90) | 0.000 | 0.000 | 0.001 | 0.009 | 0.044 | 0.067 | 0.090 | 0.121 | 0.144 | 0.145 | 0.198 |
| Long-acting benzodiazepines (30) | 0.001 | 0.001 | 0.001 | 0.007 | 0.032 | 0.045 | 0.049 | 0.050 | 0.049 | 0.043 | 0.043 |
| Long-acting benzodiazepines (90) | 0.000 | 0.000 | 0.001 | 0.004 | 0.022 | 0.032 | 0.037 | 0.039 | 0.039 | 0.035 | 0.035 |
| Clonazepam (30) | 0.000 | 0.000 | 0.000 | 0.001 | 0.005 | 0.006 | 0.007 | 0.008 | 0.008 | 0.007 | 0.008 |
| Clonazepam (90) | 0.000 | 0.000 | 0.000 | 0.001 | 0.004 | 0.005 | 0.006 | 0.007 | 0.007 | 0.006 | 0.007 |
| Diazepam (30) | 0.000 | 0.000 | 0.000 | 0.005 | 0.025 | 0.035 | 0.035 | 0.034 | 0.032 | 0.028 | 0.027 |
| Diazepam (90) | 0.000 | 0.000 | 0.000 | 0.003 | 0.016 | 0.024 | 0.026 | 0.025 | 0.024 | 0.021 | 0.021 |
| Chlordiazepoxide (30) | 0.000 | 0.000 | 0.000 | 0.000 | 0.002 | 0.004 | 0.004 | 0.003 | 0.002 | 0.002 | 0.001 |
| Chlordiazepoxide (90) | 0.000 | 0.000 | 0.000 | 0.000 | 0.001 | 0.001 | 0.002 | 0.001 | 0.001 | 0.001 | 0.001 |
| Clorazepate (30) | 0.000 | 0.000 | 0.000 | 0.000 | 0.000 | 0.000 | 0.000 | 0.000 | 0.000 | 0.000 | 0.000 |
| Clorazepate (90) | 0.000 | 0.000 | 0.000 | 0.000 | 0.000 | 0.000 | 0.000 | 0.000 | 0.000 | 0.000 | 0.000 |
| Lorazepam (30) | 0.000 | 0.000 | 0.000 | 0.001 | 0.003 | 0.003 | 0.003 | 0.004 | 0.004 | 0.004 | 0.006 |
| Lorazepam (90) | 0.000 | 0.000 | 0.000 | 0.001 | 0.002 | 0.002 | 0.002 | 0.003 | 0.003 | 0.003 | 0.005 |
| Bromazepam (30) | 0.000 | 0.000 | 0.000 | 0.000 | 0.001 | 0.001 | 0.003 | 0.005 | 0.008 | 0.009 | 0.011 |
| Bromazepam (90) | 0.000 | 0.000 | 0.000 | 0.000 | 0.001 | 0.001 | 0.002 | 0.004 | 0.007 | 0.008 | 0.009 |
| Clobazam (30) | 0.000 | 0.000 | 0.000 | 0.001 | 0.001 | 0.001 | 0.001 | 0.001 | 0.001 | 0.001 | 0.001 |
| Clobazam (90) | 0.000 | 0.000 | 0.000 | 0.001 | 0.001 | 0.001 | 0.001 | 0.001 | 0.001 | 0.001 | 0.001 |
| Prazepam (30) | 0.000 | 0.000 | 0.000 | 0.000 | 0.000 | 0.000 | 0.000 | 0.001 | 0.001 | 0.001 | 0.001 |
| Prazepam (90) | 0.000 | 0.000 | 0.000 | 0.000 | 0.000 | 0.000 | 0.000 | 0.000 | 0.001 | 0.001 | 0.001 |
| Alprazolam (30) | 0.000 | 0.000 | 0.000 | 0.003 | 0.016 | 0.021 | 0.025 | 0.030 | 0.033 | 0.031 | 0.040 |
| Alprazolam (90) | 0.000 | 0.000 | 0.000 | 0.001 | 0.010 | 0.015 | 0.018 | 0.023 | 0.026 | 0.025 | 0.032 |
| Flurazepam (30) | 0.000 | 0.000 | 0.000 | 0.000 | 0.002 | 0.004 | 0.006 | 0.008 | 0.009 | 0.008 | 0.007 |
| Flurazepam (90) | 0.000 | 0.000 | 0.000 | 0.000 | 0.001 | 0.003 | 0.006 | 0.008 | 0.008 | 0.007 | 0.006 |
| Nitrazepam (30) | 0.000 | 0.000 | 0.000 | 0.000 | 0.000 | 0.000 | 0.000 | 0.000 | 0.000 | 0.000 | 0.000 |
| Nitrazepam (90) | 0.000 | 0.000 | 0.000 | 0.000 | 0.000 | 0.000 | 0.000 | 0.000 | 0.000 | 0.000 | 0.000 |
| Triazolam (30) | 0.000 | 0.000 | 0.000 | 0.000 | 0.001 | 0.002 | 0.003 | 0.005 | 0.005 | 0.006 | 0.010 |
| Triazolam (90) | 0.000 | 0.000 | 0.000 | 0.000 | 0.001 | 0.002 | 0.003 | 0.004 | 0.005 | 0.006 | 0.009 |
| Lormetazepam (30) | 0.000 | 0.000 | 0.000 | 0.000 | 0.000 | 0.000 | 0.000 | 0.000 | 0.000 | 0.000 | 0.000 |
| Lormetazepam (90) | 0.000 | 0.000 | 0.000 | 0.000 | 0.000 | 0.000 | 0.000 | 0.000 | 0.000 | 0.000 | 0.000 |
| Temazepam (30) | 0.000 | 0.000 | 0.000 | 0.000 | 0.001 | 0.001 | 0.002 | 0.004 | 0.005 | 0.006 | 0.012 |
| Temazepam (90) | 0.000 | 0.000 | 0.000 | 0.000 | 0.000 | 0.001 | 0.001 | 0.003 | 0.005 | 0.006 | 0.011 |
| Zopiclone (30) | 0.000 | 0.000 | 0.000 | 0.003 | 0.014 | 0.024 | 0.032 | 0.044 | 0.051 | 0.050 | 0.072 |
| Zopiclone (90) | 0.000 | 0.000 | 0.000 | 0.002 | 0.011 | 0.019 | 0.027 | 0.038 | 0.044 | 0.044 | 0.065 |
| Zolpidem (30) | 0.000 | 0.000 | 0.000 | 0.002 | 0.007 | 0.013 | 0.022 | 0.035 | 0.044 | 0.046 | 0.063 |
| Zolpidem (90) | 0.000 | 0.000 | 0.000 | 0.001 | 0.005 | 0.010 | 0.018 | 0.030 | 0.038 | 0.040 | 0.056 |
| Antidepressant (30) | 0.000 | 0.000 | 0.000 | 0.003 | 0.015 | 0.017 | 0.020 | 0.023 | 0.024 | 0.025 | 0.049 |
| Antidepressant (90) | 0.000 | 0.000 | 0.000 | 0.002 | 0.010 | 0.012 | 0.015 | 0.019 | 0.020 | 0.021 | 0.043 |
| Antihistamines (30) | 0.000 | 0.000 | 0.000 | 0.003 | 0.011 | 0.011 | 0.012 | 0.011 | 0.008 | 0.006 | 0.006 |
| Antihistamines (90) | 0.000 | 0.000 | 0.000 | 0.002 | 0.007 | 0.008 | 0.009 | 0.009 | 0.006 | 0.005 | 0.005 |
| Antipsychotics (30) | 0.000 | 0.000 | 0.000 | 0.009 | 0.026 | 0.025 | 0.025 | 0.025 | 0.022 | 0.021 | 0.050 |
| Antipsychotics (90) | 0.000 | 0.000 | 0.000 | 0.007 | 0.020 | 0.020 | 0.020 | 0.021 | 0.019 | 0.018 | 0.045 |

**Prevalence of chronic use >30/>90 days by sex**

|  | F | M |
| --- | --- | --- |
| 2014 |  |  |
| Sedative (30) | 0.147 | 0.094 |
| Sedative (90) | 0.118 | 0.073 |
| Benzodiazepines and z-drugs (30) | 0.138 | 0.087 |
| Benzodiazepines and z-drugs after (90) | 0.109 | 0.066 |
| Long-acting benzodiazepines (30) | 0.040 | 0.032 |
| Long-acting benzodiazepines (90) | 0.029 | 0.023 |
| Clonazepam (30) | 0.004 | 0.004 |
| Clonazepam (90) | 0.003 | 0.003 |
| Diazepam (30) | 0.025 | 0.019 |
| Diazepam (90) | 0.017 | 0.013 |
| Chlordiazepoxide (30) | 0.002 | 0.003 |
| Chlordiazepoxide (90) | 0.001 | 0.001 |
| Clorazepate (30) | 0.000 | 0.000 |
| Clorazepate (90) | 0.000 | 0.000 |
| Lorazepam (30) | 0.003 | 0.002 |
| Lorazepam (90) | 0.002 | 0.002 |
| Bromazepam (30) | 0.007 | 0.003 |
| Bromazepam (90) | 0.006 | 0.002 |
| Clobazam (30) | 0.001 | 0.001 |
| Clobazam (90) | 0.001 | 0.001 |
| Prazepam (30) | 0.001 | 0.001 |
| Prazepam (90) | 0.001 | 0.000 |
| Alprazolam (30) | 0.028 | 0.015 |
| Alprazolam (90) | 0.019 | 0.010 |
| Flurazepam (30) | 0.007 | 0.006 |
| Flurazepam (90) | 0.006 | 0.005 |
| Nitrazepam (30) | 0.002 | 0.001 |
| Nitrazepam (90) | 0.001 | 0.001 |
| Triazolam (30) | 0.005 | 0.003 |
| Triazolam (90) | 0.004 | 0.003 |
| Lormetazepam (30) | 0.004 | 0.002 |
| Lormetazepam (90) | 0.003 | 0.001 |
| Temazepam (30) | 0.008 | 0.004 |
| Temazepam (90) | 0.007 | 0.003 |
| Zopiclone (30) | 0.037 | 0.024 |
| Zopiclone (90) | 0.029 | 0.019 |
| Zolpidem (30) | 0.032 | 0.017 |
| Zolpidem (90) | 0.025 | 0.012 |
| Antidepressant (30) | 0.012 | 0.008 |
| Antidepressant (90) | 0.008 | 0.005 |
| Antihistamines (30) | 0.001 | 0.001 |
| Antihistamines (90) | 0.001 | 0.001 |
| Antipsychotics (30) | 0.014 | 0.010 |
| Antipsychotics (90) | 0.011 | 0.008 |
| 2015 |  |  |
| Sedative (30) | 0.146 | 0.094 |
| Sedative (90) | 0.121 | 0.076 |
| Benzodiazepines and z-drugs (30) | 0.135 | 0.086 |
| Benzodiazepines and z-drugs after (90) | 0.111 | 0.068 |
| Long-acting benzodiazepines (30) | 0.039 | 0.031 |
| Long-acting benzodiazepines (90) | 0.030 | 0.023 |
| Clonazepam (30) | 0.004 | 0.004 |
| Clonazepam (90) | 0.004 | 0.003 |
| Diazepam (30) | 0.025 | 0.019 |
| Diazepam (90) | 0.018 | 0.014 |
| Chlordiazepoxide (30) | 0.002 | 0.003 |
| Chlordiazepoxide (90) | 0.001 | 0.001 |
| Clorazepate (30) | 0.000 | 0.000 |
| Clorazepate (90) | 0.000 | 0.000 |
| Lorazepam (30) | 0.003 | 0.002 |
| Lorazepam (90) | 0.002 | 0.002 |
| Bromazepam (30) | 0.007 | 0.003 |
| Bromazepam (90) | 0.006 | 0.002 |
| Clobazam (30) | 0.001 | 0.001 |
| Clobazam (90) | 0.001 | 0.001 |
| Prazepam (30) | 0.001 | 0.001 |
| Prazepam (90) | 0.001 | 0.000 |
| Alprazolam (30) | 0.028 | 0.015 |
| Alprazolam (90) | 0.021 | 0.011 |
| Flurazepam (30) | 0.007 | 0.006 |
| Flurazepam (90) | 0.006 | 0.005 |
| Nitrazepam (30) | 0.001 | 0.001 |
| Nitrazepam (90) | 0.001 | 0.001 |
| Triazolam (30) | 0.005 | 0.003 |
| Triazolam (90) | 0.005 | 0.003 |
| Lormetazepam (30) | 0.004 | 0.001 |
| Lormetazepam (90) | 0.003 | 0.001 |
| Temazepam (30) | 0.007 | 0.004 |
| Temazepam (90) | 0.006 | 0.003 |
| Zopiclone (30) | 0.037 | 0.025 |
| Zopiclone (90) | 0.031 | 0.020 |
| Zolpidem (30) | 0.032 | 0.016 |
| Zolpidem (90) | 0.026 | 0.013 |
| Antidepressant (30) | 0.013 | 0.009 |
| Antidepressant (90) | 0.010 | 0.006 |
| Antihistamines (30) | 0.002 | 0.001 |
| Antihistamines (90) | 0.001 | 0.001 |
| Antipsychotics (30) | 0.015 | 0.012 |
| Antipsychotics (90) | 0.013 | 0.009 |
| 2016 |  |  |
| Sedative (30) | 0.151 | 0.098 |
| Sedative (90) | 0.126 | 0.080 |
| Benzodiazepines and z-drugs (30) | 0.138 | 0.088 |
| Benzodiazepines and z-drugs after (90) | 0.114 | 0.070 |
| Long-acting benzodiazepines (30) | 0.040 | 0.032 |
| Long-acting benzodiazepines (90) | 0.030 | 0.024 |
| Clonazepam (30) | 0.005 | 0.004 |
| Clonazepam (90) | 0.004 | 0.004 |
| Diazepam (30) | 0.026 | 0.019 |
| Diazepam (90) | 0.018 | 0.014 |
| Chlordiazepoxide (30) | 0.002 | 0.003 |
| Chlordiazepoxide (90) | 0.001 | 0.001 |
| Clorazepate (30) | 0.000 | 0.000 |
| Clorazepate (90) | 0.000 | 0.000 |
| Lorazepam (30) | 0.003 | 0.002 |
| Lorazepam (90) | 0.002 | 0.002 |
| Bromazepam (30) | 0.007 | 0.003 |
| Bromazepam (90) | 0.006 | 0.002 |
| Clobazam (30) | 0.001 | 0.001 |
| Clobazam (90) | 0.001 | 0.001 |
| Prazepam (30) | 0.001 | 0.001 |
| Prazepam (90) | 0.001 | 0.000 |
| Alprazolam (30) | 0.029 | 0.015 |
| Alprazolam (90) | 0.021 | 0.011 |
| Flurazepam (30) | 0.007 | 0.006 |
| Flurazepam (90) | 0.006 | 0.005 |
| Nitrazepam (30) | 0.001 | 0.001 |
| Nitrazepam (90) | 0.001 | 0.001 |
| Triazolam (30) | 0.005 | 0.003 |
| Triazolam (90) | 0.005 | 0.003 |
| Lormetazepam (30) | 0.003 | 0.001 |
| Lormetazepam (90) | 0.003 | 0.001 |
| Temazepam (30) | 0.007 | 0.004 |
| Temazepam (90) | 0.006 | 0.003 |
| Zopiclone (30) | 0.038 | 0.026 |
| Zopiclone (90) | 0.032 | 0.021 |
| Zolpidem (30) | 0.033 | 0.017 |
| Zolpidem (90) | 0.027 | 0.013 |
| Antidepressant (30) | 0.015 | 0.010 |
| Antidepressant (90) | 0.012 | 0.007 |
| Antihistamines (30) | 0.002 | 0.002 |
| Antihistamines (90) | 0.001 | 0.001 |
| Antipsychotics (30) | 0.017 | 0.013 |
| Antipsychotics (90) | 0.014 | 0.010 |
| 2017 |  |  |
| Sedative (30) | 0.155 | 0.102 |
| Sedative (90) | 0.129 | 0.083 |
| Benzodiazepines and z-drugs (30) | 0.140 | 0.090 |
| Benzodiazepines and z-drugs after (90) | 0.115 | 0.072 |
| Long-acting benzodiazepines (30) | 0.040 | 0.032 |
| Long-acting benzodiazepines (90) | 0.030 | 0.024 |
| Clonazepam (30) | 0.005 | 0.005 |
| Clonazepam (90) | 0.004 | 0.004 |
| Diazepam (30) | 0.026 | 0.019 |
| Diazepam (90) | 0.018 | 0.014 |
| Chlordiazepoxide (30) | 0.002 | 0.003 |
| Chlordiazepoxide (90) | 0.001 | 0.001 |
| Clorazepate (30) | 0.000 | 0.000 |
| Clorazepate (90) | 0.000 | 0.000 |
| Lorazepam (30) | 0.003 | 0.002 |
| Lorazepam (90) | 0.002 | 0.002 |
| Bromazepam (30) | 0.007 | 0.003 |
| Bromazepam (90) | 0.006 | 0.002 |
| Clobazam (30) | 0.001 | 0.001 |
| Clobazam (90) | 0.001 | 0.001 |
| Prazepam (30) | 0.001 | 0.001 |
| Prazepam (90) | 0.001 | 0.000 |
| Alprazolam (30) | 0.029 | 0.016 |
| Alprazolam (90) | 0.022 | 0.012 |
| Flurazepam (30) | 0.006 | 0.006 |
| Flurazepam (90) | 0.006 | 0.005 |
| Nitrazepam (30) | 0.001 | 0.001 |
| Nitrazepam (90) | 0.001 | 0.001 |
| Triazolam (30) | 0.005 | 0.003 |
| Triazolam (90) | 0.005 | 0.003 |
| Lormetazepam (30) | 0.003 | 0.001 |
| Lormetazepam (90) | 0.003 | 0.001 |
| Temazepam (30) | 0.006 | 0.004 |
| Temazepam (90) | 0.006 | 0.003 |
| Zopiclone (30) | 0.039 | 0.027 |
| Zopiclone (90) | 0.033 | 0.022 |
| Zolpidem (30) | 0.034 | 0.017 |
| Zolpidem (90) | 0.028 | 0.014 |
| Antidepressant (30) | 0.017 | 0.011 |
| Antidepressant (90) | 0.013 | 0.008 |
| Antihistamines (30) | 0.003 | 0.002 |
| Antihistamines (90) | 0.002 | 0.001 |
| Antipsychotics (30) | 0.018 | 0.014 |
| Antipsychotics (90) | 0.015 | 0.012 |
| 2018 |  |  |
| Sedative (30) | 0.158 | 0.104 |
| Sedative (90) | 0.132 | 0.086 |
| Benzodiazepines and z-drugs (30) | 0.140 | 0.090 |
| Benzodiazepines and z-drugs after (90) | 0.116 | 0.073 |
| Long-acting benzodiazepines (30) | 0.040 | 0.032 |
| Long-acting benzodiazepines (90) | 0.030 | 0.025 |
| Clonazepam (30) | 0.005 | 0.005 |
| Clonazepam (90) | 0.004 | 0.004 |
| Diazepam (30) | 0.026 | 0.019 |
| Diazepam (90) | 0.018 | 0.015 |
| Chlordiazepoxide (30) | 0.002 | 0.003 |
| Chlordiazepoxide (90) | 0.001 | 0.001 |
| Clorazepate (30) | 0.000 | 0.000 |
| Clorazepate (90) | 0.000 | 0.000 |
| Lorazepam (30) | 0.003 | 0.003 |
| Lorazepam (90) | 0.002 | 0.002 |
| Bromazepam (30) | 0.007 | 0.003 |
| Bromazepam (90) | 0.006 | 0.002 |
| Clobazam (30) | 0.001 | 0.001 |
| Clobazam (90) | 0.001 | 0.001 |
| Prazepam (30) | 0.001 | 0.001 |
| Prazepam (90) | 0.001 | 0.000 |
| Alprazolam (30) | 0.029 | 0.016 |
| Alprazolam (90) | 0.022 | 0.012 |
| Flurazepam (30) | 0.006 | 0.006 |
| Flurazepam (90) | 0.006 | 0.005 |
| Nitrazepam (30) | 0.001 | 0.001 |
| Nitrazepam (90) | 0.001 | 0.001 |
| Triazolam (30) | 0.005 | 0.003 |
| Triazolam (90) | 0.005 | 0.003 |
| Lormetazepam (30) | 0.003 | 0.001 |
| Lormetazepam (90) | 0.003 | 0.001 |
| Temazepam (30) | 0.006 | 0.003 |
| Temazepam (90) | 0.005 | 0.003 |
| Zopiclone (30) | 0.039 | 0.027 |
| Zopiclone (90) | 0.033 | 0.022 |
| Zolpidem (30) | 0.034 | 0.018 |
| Zolpidem (90) | 0.028 | 0.014 |
| Antidepressant (30) | 0.018 | 0.012 |
| Antidepressant (90) | 0.014 | 0.009 |
| Antihistamines (30) | 0.004 | 0.003 |
| Antihistamines (90) | 0.002 | 0.002 |
| Antipsychotics (30) | 0.020 | 0.016 |
| Antipsychotics (90) | 0.017 | 0.013 |
| 2019 |  |  |
| Sedative (30) | 0.159 | 0.105 |
| Sedative (90) | 0.134 | 0.087 |
| Benzodiazepines and z-drugs (30) | 0.139 | 0.089 |
| Benzodiazepines and z-drugs after (90) | 0.115 | 0.073 |
| Long-acting benzodiazepines (30) | 0.039 | 0.032 |
| Long-acting benzodiazepines (90) | 0.030 | 0.025 |
| Clonazepam (30) | 0.005 | 0.005 |
| Clonazepam (90) | 0.004 | 0.004 |
| Diazepam (30) | 0.026 | 0.020 |
| Diazepam (90) | 0.018 | 0.015 |
| Chlordiazepoxide (30) | 0.002 | 0.003 |
| Chlordiazepoxide (90) | 0.001 | 0.001 |
| Clorazepate (30) | 0.000 | 0.000 |
| Clorazepate (90) | 0.000 | 0.000 |
| Lorazepam (30) | 0.003 | 0.003 |
| Lorazepam (90) | 0.002 | 0.002 |
| Bromazepam (30) | 0.006 | 0.002 |
| Bromazepam (90) | 0.006 | 0.002 |
| Clobazam (30) | 0.001 | 0.001 |
| Clobazam (90) | 0.001 | 0.001 |
| Prazepam (30) | 0.001 | 0.001 |
| Prazepam (90) | 0.001 | 0.000 |
| Alprazolam (30) | 0.029 | 0.015 |
| Alprazolam (90) | 0.022 | 0.012 |
| Flurazepam (30) | 0.006 | 0.005 |
| Flurazepam (90) | 0.005 | 0.005 |
| Nitrazepam (30) | 0.001 | 0.001 |
| Nitrazepam (90) | 0.001 | 0.000 |
| Triazolam (30) | 0.005 | 0.003 |
| Triazolam (90) | 0.005 | 0.003 |
| Lormetazepam (30) | 0.003 | 0.001 |
| Lormetazepam (90) | 0.002 | 0.001 |
| Temazepam (30) | 0.006 | 0.003 |
| Temazepam (90) | 0.005 | 0.003 |
| Zopiclone (30) | 0.039 | 0.027 |
| Zopiclone (90) | 0.034 | 0.023 |
| Zolpidem (30) | 0.035 | 0.018 |
| Zolpidem (90) | 0.029 | 0.014 |
| Antidepressant (30) | 0.020 | 0.013 |
| Antidepressant (90) | 0.016 | 0.010 |
| Antihistamines (30) | 0.005 | 0.003 |
| Antihistamines (90) | 0.003 | 0.002 |
| Antipsychotics (30) | 0.022 | 0.017 |
| Antipsychotics (90) | 0.018 | 0.014 |
| 2020 |  |  |
| Sedative (30) | 0.155 | 0.102 |
| Sedative (90) | 0.132 | 0.086 |
| Benzodiazepines and z-drugs (30) | 0.133 | 0.085 |
| Benzodiazepines and z-drugs after (90) | 0.111 | 0.070 |
| Long-acting benzodiazepines (30) | 0.038 | 0.031 |
| Long-acting benzodiazepines (90) | 0.029 | 0.024 |
| Clonazepam (30) | 0.005 | 0.005 |
| Clonazepam (90) | 0.005 | 0.004 |
| Diazepam (30) | 0.025 | 0.019 |
| Diazepam (90) | 0.018 | 0.014 |
| Chlordiazepoxide (30) | 0.001 | 0.003 |
| Chlordiazepoxide (90) | 0.001 | 0.001 |
| Clorazepate (30) | 0.000 | 0.000 |
| Clorazepate (90) | 0.000 | 0.000 |
| Lorazepam (30) | 0.003 | 0.003 |
| Lorazepam (90) | 0.002 | 0.002 |
| Bromazepam (30) | 0.006 | 0.002 |
| Bromazepam (90) | 0.005 | 0.002 |
| Clobazam (30) | 0.001 | 0.001 |
| Clobazam (90) | 0.001 | 0.001 |
| Prazepam (30) | 0.001 | 0.000 |
| Prazepam (90) | 0.001 | 0.000 |
| Alprazolam (30) | 0.029 | 0.015 |
| Alprazolam (90) | 0.021 | 0.011 |
| Flurazepam (30) | 0.006 | 0.005 |
| Flurazepam (90) | 0.005 | 0.004 |
| Nitrazepam (30) | 0.001 | 0.001 |
| Nitrazepam (90) | 0.001 | 0.000 |
| Triazolam (30) | 0.005 | 0.003 |
| Triazolam (90) | 0.004 | 0.003 |
| Lormetazepam (30) | 0.000 | 0.000 |
| Lormetazepam (90) | 0.000 | 0.000 |
| Temazepam (30) | 0.005 | 0.003 |
| Temazepam (90) | 0.005 | 0.002 |
| Zopiclone (30) | 0.038 | 0.026 |
| Zopiclone (90) | 0.033 | 0.022 |
| Zolpidem (30) | 0.034 | 0.017 |
| Zolpidem (90) | 0.029 | 0.014 |
| Antidepressant (30) | 0.021 | 0.014 |
| Antidepressant (90) | 0.017 | 0.011 |
| Antihistamines (30) | 0.006 | 0.004 |
| Antihistamines (90) | 0.004 | 0.003 |
| Antipsychotics (30) | 0.023 | 0.018 |
| Antipsychotics (90) | 0.020 | 0.015 |
| 2021 |  |  |
| Sedative (30) | 0.160 | 0.105 |
| Sedative (90) | 0.136 | 0.089 |
| Benzodiazepines and z-drugs (30) | 0.134 | 0.086 |
| Benzodiazepines and z-drugs after (90) | 0.112 | 0.071 |
| Long-acting benzodiazepines (30) | 0.038 | 0.031 |
| Long-acting benzodiazepines (90) | 0.030 | 0.024 |
| Clonazepam (30) | 0.006 | 0.005 |
| Clonazepam (90) | 0.005 | 0.004 |
| Diazepam (30) | 0.026 | 0.019 |
| Diazepam (90) | 0.019 | 0.015 |
| Chlordiazepoxide (30) | 0.001 | 0.003 |
| Chlordiazepoxide (90) | 0.001 | 0.001 |
| Clorazepate (30) | 0.000 | 0.000 |
| Clorazepate (90) | 0.000 | 0.000 |
| Lorazepam (30) | 0.003 | 0.003 |
| Lorazepam (90) | 0.002 | 0.002 |
| Bromazepam (30) | 0.006 | 0.002 |
| Bromazepam (90) | 0.005 | 0.002 |
| Clobazam (30) | 0.001 | 0.001 |
| Clobazam (90) | 0.001 | 0.001 |
| Prazepam (30) | 0.001 | 0.000 |
| Prazepam (90) | 0.001 | 0.000 |
| Alprazolam (30) | 0.029 | 0.015 |
| Alprazolam (90) | 0.022 | 0.011 |
| Flurazepam (30) | 0.005 | 0.005 |
| Flurazepam (90) | 0.005 | 0.004 |
| Nitrazepam (30) | 0.001 | 0.000 |
| Nitrazepam (90) | 0.001 | 0.000 |
| Triazolam (30) | 0.005 | 0.003 |
| Triazolam (90) | 0.004 | 0.003 |
| Lormetazepam (30) | 0.000 | 0.000 |
| Lormetazepam (90) | 0.000 | 0.000 |
| Temazepam (30) | 0.005 | 0.003 |
| Temazepam (90) | 0.005 | 0.002 |
| Zopiclone (30) | 0.039 | 0.026 |
| Zopiclone (90) | 0.033 | 0.022 |
| Zolpidem (30) | 0.034 | 0.018 |
| Zolpidem (90) | 0.030 | 0.015 |
| Antidepressant (30) | 0.023 | 0.015 |
| Antidepressant (90) | 0.019 | 0.012 |
| Antihistamines (30) | 0.008 | 0.005 |
| Antihistamines (90) | 0.005 | 0.004 |
| Antipsychotics (30) | 0.025 | 0.019 |
| Antipsychotics (90) | 0.022 | 0.016 |
| 2022 |  |  |
| Sedative (30) | 0.153 | 0.102 |
| Sedative (90) | 0.131 | 0.086 |
| Benzodiazepines and z-drugs (30) | 0.127 | 0.082 |
| Benzodiazepines and z-drugs after (90) | 0.106 | 0.068 |
| Long-acting benzodiazepines (30) | 0.036 | 0.030 |
| Long-acting benzodiazepines (90) | 0.027 | 0.023 |
| Clonazepam (30) | 0.005 | 0.005 |
| Clonazepam (90) | 0.005 | 0.004 |
| Diazepam (30) | 0.025 | 0.019 |
| Diazepam (90) | 0.018 | 0.015 |
| Chlordiazepoxide (30) | 0.001 | 0.003 |
| Chlordiazepoxide (90) | 0.001 | 0.001 |
| Clorazepate (30) | 0.000 | 0.000 |
| Clorazepate (90) | 0.000 | 0.000 |
| Lorazepam (30) | 0.003 | 0.003 |
| Lorazepam (90) | 0.002 | 0.002 |
| Bromazepam (30) | 0.006 | 0.002 |
| Bromazepam (90) | 0.005 | 0.002 |
| Clobazam (30) | 0.001 | 0.001 |
| Clobazam (90) | 0.001 | 0.001 |
| Prazepam (30) | 0.001 | 0.000 |
| Prazepam (90) | 0.001 | 0.000 |
| Alprazolam (30) | 0.027 | 0.014 |
| Alprazolam (90) | 0.020 | 0.011 |
| Flurazepam (30) | 0.005 | 0.004 |
| Flurazepam (90) | 0.004 | 0.004 |
| Nitrazepam (30) | 0.000 | 0.000 |
| Nitrazepam (90) | 0.000 | 0.000 |
| Triazolam (30) | 0.004 | 0.003 |
| Triazolam (90) | 0.004 | 0.002 |
| Lormetazepam (30) | 0.000 | 0.000 |
| Lormetazepam (90) | 0.000 | 0.000 |
| Temazepam (30) | 0.005 | 0.003 |
| Temazepam (90) | 0.004 | 0.002 |
| Zopiclone (30) | 0.037 | 0.025 |
| Zopiclone (90) | 0.032 | 0.022 |
| Zolpidem (30) | 0.033 | 0.017 |
| Zolpidem (90) | 0.028 | 0.014 |
| Antidepressant (30) | 0.024 | 0.015 |
| Antidepressant (90) | 0.019 | 0.012 |
| Antihistamines (30) | 0.008 | 0.006 |
| Antihistamines (90) | 0.006 | 0.004 |
| Antipsychotics (30) | 0.025 | 0.019 |
| Antipsychotics (90) | 0.022 | 0.016 |

**Benzodiazepines or z-drugs dispensed at a dose equivalent to >40 mg diazepam per day by age group**

|  | <5 | 5-11 | 12-15 | 16-24 | 25-34 | 35-44 | 45-54 | 55-64 | 65-69 | 70-74 | 75+ |
| --- | --- | --- | --- | --- | --- | --- | --- | --- | --- | --- | --- |
| 2014 |  |  |  |  |  |  |  |  |  |  |  |
| Benzodiazepines | 0.000 | 0.000 | 0.000 | 0.002 | 0.011 | 0.015 | 0.018 | 0.020 | 0.019 | 0.014 | 0.012 |
| Long-acting benzodiazepines | 0.000 | 0.000 | 0.000 | 0.002 | 0.008 | 0.012 | 0.013 | 0.013 | 0.010 | 0.007 | 0.004 |
| 2015 |  |  |  |  |  |  |  |  |  |  |  |
| Benzodiazepines | 0.000 | 0.000 | 0.000 | 0.002 | 0.011 | 0.015 | 0.017 | 0.019 | 0.017 | 0.014 | 0.011 |
| Long-acting benzodiazepines | 0.000 | 0.000 | 0.000 | 0.002 | 0.008 | 0.011 | 0.012 | 0.012 | 0.010 | 0.007 | 0.004 |
| 2016 |  |  |  |  |  |  |  |  |  |  |  |
| Benzodiazepines | 0.000 | 0.000 | 0.000 | 0.002 | 0.012 | 0.015 | 0.017 | 0.018 | 0.017 | 0.013 | 0.011 |
| Long-acting benzodiazepines | 0.000 | 0.000 | 0.000 | 0.002 | 0.009 | 0.012 | 0.012 | 0.012 | 0.009 | 0.007 | 0.004 |
| 2017 |  |  |  |  |  |  |  |  |  |  |  |
| Benzodiazepines | 0.000 | 0.000 | 0.000 | 0.002 | 0.012 | 0.017 | 0.018 | 0.018 | 0.015 | 0.013 | 0.010 |
| Long-acting benzodiazepines | 0.000 | 0.000 | 0.000 | 0.002 | 0.009 | 0.013 | 0.013 | 0.012 | 0.009 | 0.006 | 0.004 |
| 2018 |  |  |  |  |  |  |  |  |  |  |  |
| Benzodiazepines | 0.000 | 0.000 | 0.000 | 0.002 | 0.013 | 0.017 | 0.017 | 0.018 | 0.015 | 0.012 | 0.009 |
| Long-acting benzodiazepines | 0.000 | 0.000 | 0.000 | 0.002 | 0.010 | 0.013 | 0.012 | 0.011 | 0.009 | 0.006 | 0.004 |
| 2019 |  |  |  |  |  |  |  |  |  |  |  |
| Benzodiazepines | 0.000 | 0.000 | 0.000 | 0.002 | 0.013 | 0.017 | 0.016 | 0.017 | 0.014 | 0.012 | 0.009 |
| Long-acting benzodiazepines | 0.000 | 0.000 | 0.000 | 0.001 | 0.009 | 0.013 | 0.012 | 0.011 | 0.008 | 0.006 | 0.004 |
| 2020 |  |  |  |  |  |  |  |  |  |  |  |
| Benzodiazepines | 0.000 | 0.000 | 0.000 | 0.002 | 0.011 | 0.015 | 0.016 | 0.016 | 0.014 | 0.011 | 0.009 |
| Long-acting benzodiazepines | 0.000 | 0.000 | 0.000 | 0.001 | 0.008 | 0.011 | 0.011 | 0.011 | 0.008 | 0.006 | 0.004 |
| 2021 |  |  |  |  |  |  |  |  |  |  |  |
| Benzodiazepines | 0.000 | 0.000 | 0.000 | 0.002 | 0.011 | 0.015 | 0.016 | 0.016 | 0.014 | 0.010 | 0.008 |
| Long-acting benzodiazepines | 0.000 | 0.000 | 0.000 | 0.001 | 0.008 | 0.011 | 0.011 | 0.010 | 0.008 | 0.005 | 0.004 |
| 2022 |  |  |  |  |  |  |  |  |  |  |  |
| Benzodiazepines | 0.000 | 0.000 | 0.000 | 0.002 | 0.010 | 0.014 | 0.015 | 0.015 | 0.013 | 0.010 | 0.008 |
| Long-acting benzodiazepines | 0.000 | 0.000 | 0.000 | 0.001 | 0.007 | 0.010 | 0.011 | 0.010 | 0.008 | 0.005 | 0.004 |

**Benzodiazepines or z-drugs dispensed at a dose equivalent to >40 mg diazepam per day by sex**

|  | F | M |
| --- | --- | --- |
| 2014 |  |  |
| Benzodiazepines | 0.011 | 0.010 |
| Long-acting benzodiazepines | 0.006 | 0.007 |
| 2015 |  |  |
| Benzodiazepines | 0.011 | 0.009 |
| Long-acting benzodiazepines | 0.006 | 0.007 |
| 2016 |  |  |
| Benzodiazepines | 0.011 | 0.010 |
| Long-acting benzodiazepines | 0.006 | 0.007 |
| 2017 |  |  |
| Benzodiazepines | 0.011 | 0.010 |
| Long-acting benzodiazepines | 0.006 | 0.007 |
| 2018 |  |  |
| Benzodiazepines | 0.011 | 0.010 |
| Long-acting benzodiazepines | 0.006 | 0.007 |
| 2019 |  |  |
| Benzodiazepines | 0.010 | 0.010 |
| Long-acting benzodiazepines | 0.006 | 0.007 |
| 2020 |  |  |
| Benzodiazepines | 0.010 | 0.009 |
| Long-acting benzodiazepines | 0.006 | 0.006 |
| 2021 |  |  |
| Benzodiazepines | 0.009 | 0.009 |
| Long-acting benzodiazepines | 0.006 | 0.006 |
| 2022 |  |  |
| Benzodiazepines | 0.009 | 0.009 |
| Long-acting benzodiazepines | 0.005 | 0.006 |
